# On the lag between deaths and infections in the first phase of the Covid-19 pandemic

**DOI:** 10.1101/2021.01.01.21249115

**Authors:** Piotr T. Chruściel, Sebastian J. Szybka

**Author notes:** Corresponding author;, URL homepage.univie.ac.at/piotr.chrusciel.

## Abstract

One of the key issues in fighting the current pandemic, or the ones to come, is to obtain objective quantitative indicators of the effectiveness of the measures taken to contain the epidemic. The aim of this work is to point out that the lag between the daily number of infections and casualties provides one such indicator. For this we determined the lag during the first phase of the Covid-19 pandemic for a series of countries using the data available at the server of the John Hopkins University using three different methods. Somewhat surprisingly, we find a lag varying substantially between countries, taking negative values (thus the maximum daily number of casulties preceding the maximum daily namber of new infections) in countries where no steps to contain the epidemic have been taken at the outset, with an average lag of 7 ± 0.3 days. Our results can be useful to health authorities in a search for the best strategy to fight the epidemic.

**Key Messages:**

- The lags between the maximum daily infections and casualties during the first phase of the Covid-19 pandemic differ widely between countries.
- These lags are clear for some countries, but impossible to determine confidently for most.
- In some countries the day at which the maximal number of daily deaths is attained precedes the day of the maximal number of casualties, indicating a failure to protect the most vulnerable part of the population.
- The lags can serve as an objective quantitative measure of the effectiveness of the measures taken to contain the epidemic.

---

In several countries one observes a first phase of the Covid-19 pandemic, where the daily infections drop significantly from a clear maximum to a low rampant level, see Figures 1 and 2. The curves of daily Covid-19 related casualties for these countries follow a similar pattern. In *some* of the associated time series one can observe a *clear* lag between the maximum number of reported daily infections of Covid-19 and the maximum number of resulting daily deaths.

**Figure 1:**
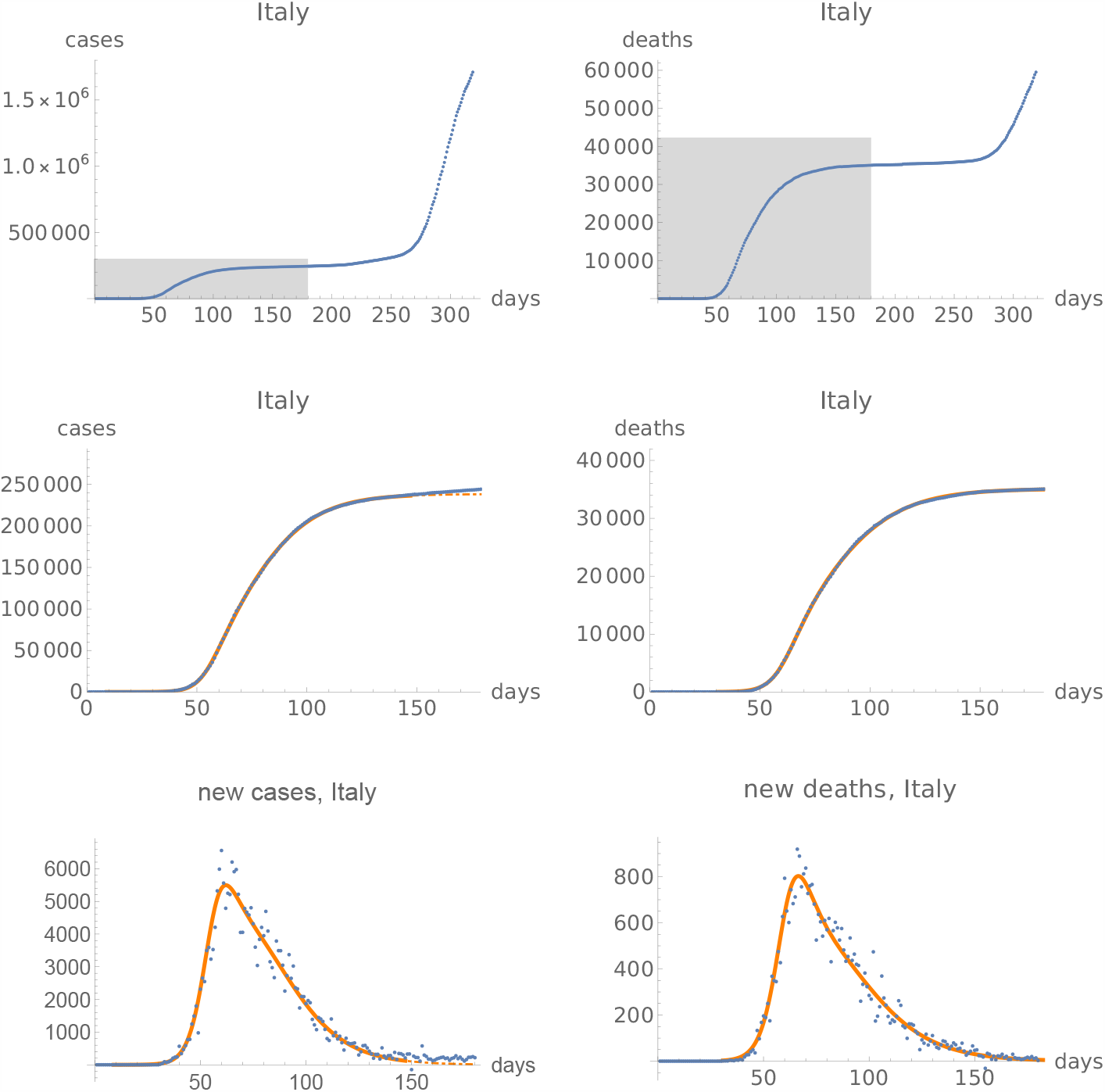
Fits of the function *I* to the first phase of the Covid-19 pandemic in Italy. First line: the cumulative cases time-series and death time-series up to December 6, 2020. The second line is a zoom to the shaded windows in the first line, where moreover the fits of the function *I*, are shown. The continuous orange curve showing the fit to the curve of accumulated cases becomes dashed on day 148 (June 16, 2020), on which the fit starts deviating from the data; we interpret this as the end of the first phase of the epidemic. Third line: Daily infections and deaths, together with the derivative of the function *I*. Note the different scales on all plots.

**Figure 2:**
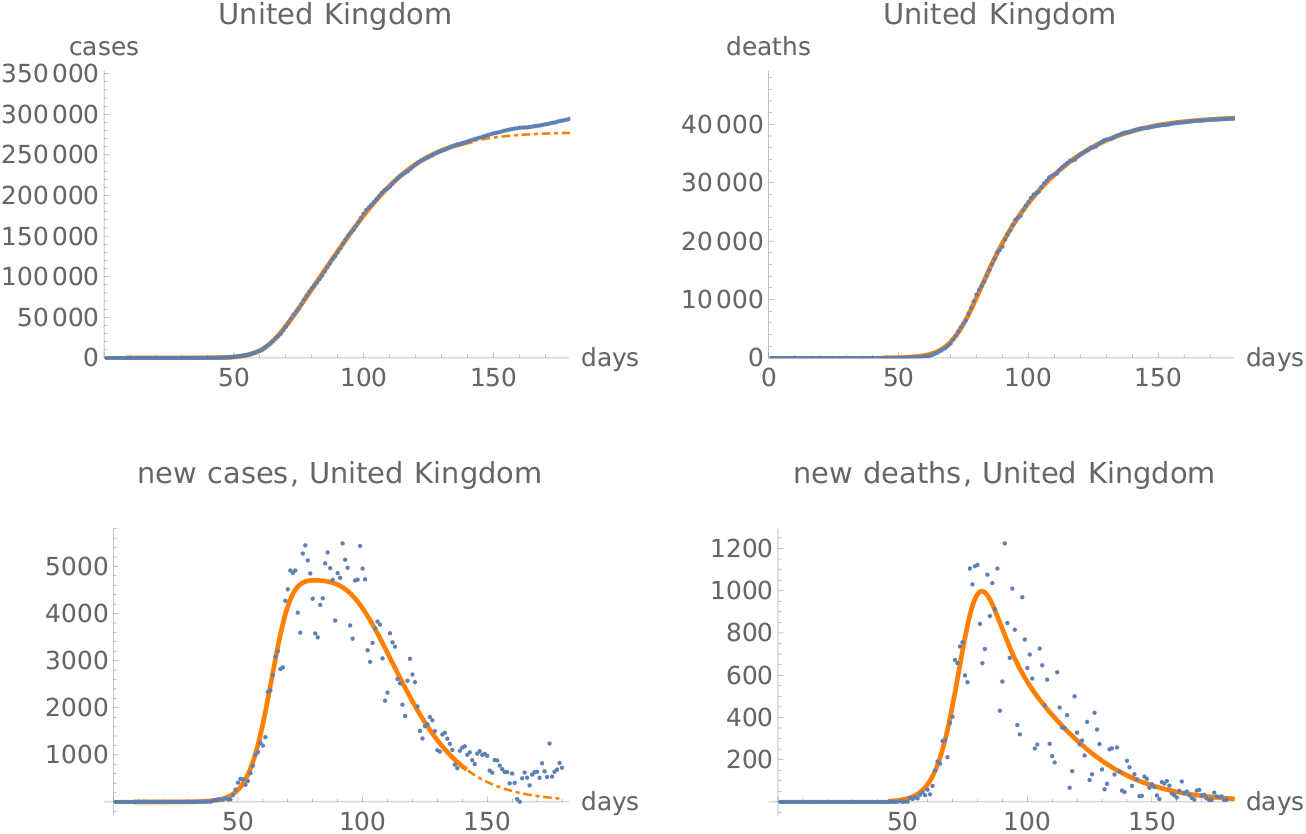
Same as the last two lines of Figure 1 for the United Kingdom.

One might expect that this lag is closely related to the number of days between infection and death of the patients. The lag should therefore be longer in countries with better health care. Since most deaths occur in older patients, this lag will be affected by the measures taken to protect the elderly population. Hence we expect that, at equal quality of health care, the lag will provide a qualitative measure of the efficacity of the measures taken to protect the older population.

The question then arises, how to determine this lag in a systematic way across countries, to be able to observe its variation from country to country and draw conclusions concerning the protection measures chosen.

Indeed, inspection of time series for various countries reveals that it is often not clear how to determine this lag in an algorithmic way from the data, without manual adjustements based on visual inspection of individual time series, or based on other considerations. The difficulty of pinpointing a precise lag are well illustrated by Figure 3, where we show the time series for daily deaths and daily new cases for Poland, Romania and Sweden in the first 180 days of the epidemic.

**Figure 3:**
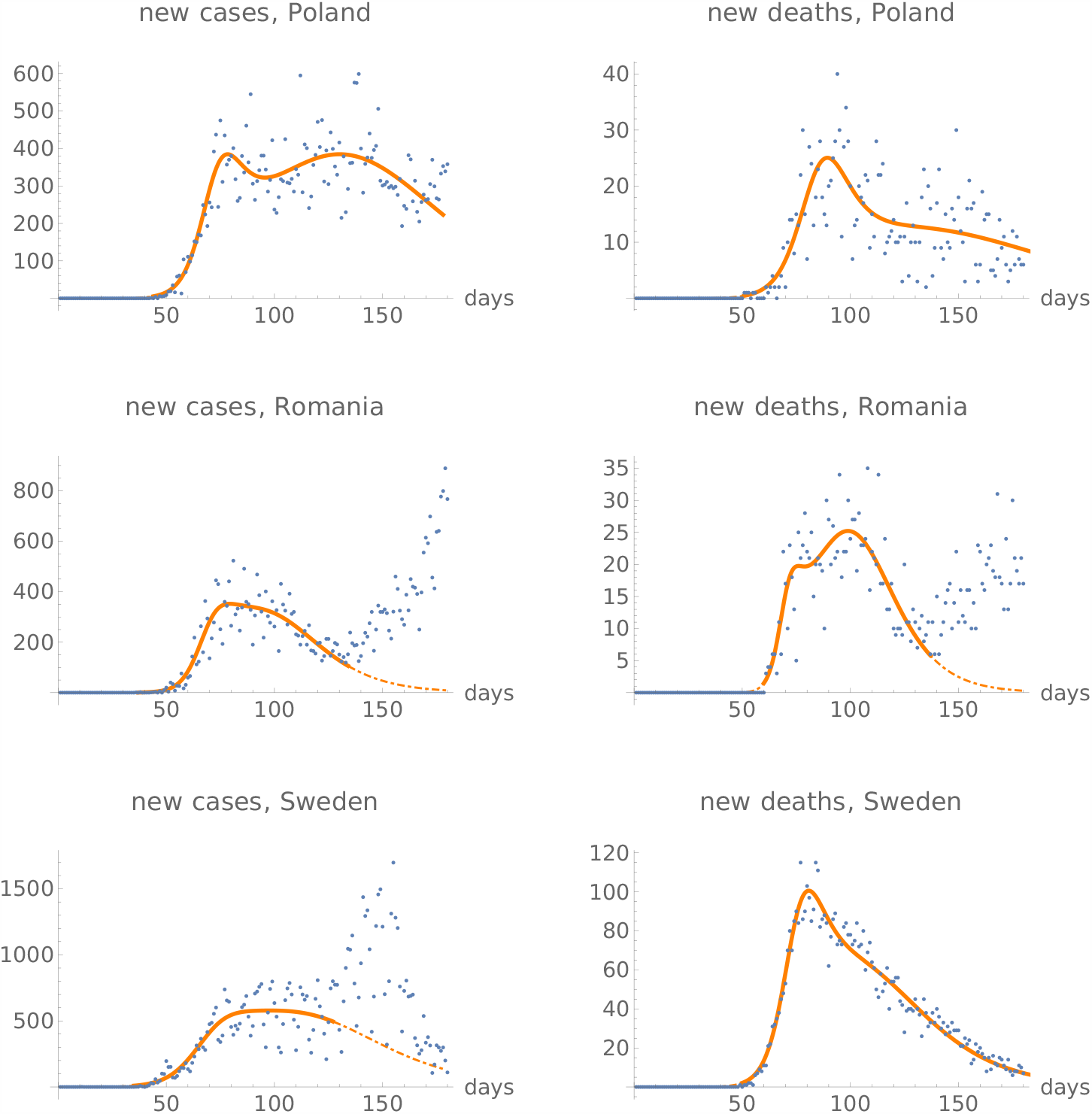
Time series for Poland, Romania and Sweden, with the derivative of the best fit of the function *I* indicated in orange. The orange lines become dashed on the day at which the fit of the function *I* to the data is optimal, when compared to fits with different lengths, indicating the start of a new phase of the epidemic. The lack of a clear peak, as well as the width of the region where the numbers of daily new infections are high before starting to decline, explain the difference seen in Table 1 between the lags as determined from the function *I* and from the integral method.

A possible tool to study this question has been provided in [1]. In that work we observed that the function

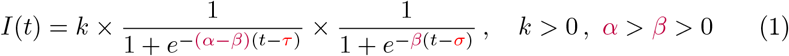

parameterised by five parameters *α, β, τ, σ* and *k*, describes surprisingly well the global features of the confirmed-cases time-series and death time-series for the *first phase* of the Covid-19 epidemic, whenever a fit to the data is available. We can therefore attempt to use the function given in Equation (1) to determine this lag. Indeed, after fitting the function *I* to the time-series of the total number of infected, the first derivative of *I* provides a fit to the daily number of new cases. The day at which the maximum number of cases has been achieved is obtained by studying the zeros of the second derivative. Similarly for the time series of the total number of deaths.

**Table 1:** Lags between the day of maximum number of deaths and the maximum number of cases for the first wave of Covid-19, ordered decreasingly. The table has been split in two, with the second part listing the countries where the lags determined by different methods differ in a significant way. The first data column is the lag determined by fitting the function (1) to the data, the third column is obtained from the same function fitted to the data averaged over a week. The second and fourth data columns are obtained from determining the maximum max *f* of the integral overlap function (2), with the interval indicating days during which *f* (*r*) ≥ .99 max *f*.

| Country | Lag from $I$ | Lag from<br>integral overlap | Lag from $I$,<br>after averaging | Lag from<br>integral overlap<br>after averaging |
| --- | --- | --- | --- | --- |
| Romania | 18 | 16 (16 – 16) | 20 | 13 (4 – 19) |
| Germany | 15 | 13 (13 – 13) | 17 | 13 (11 – 16) |
| Austria | 14 | 13 (12 – 13) | 14 | 12 (11 – 14) |
| Poland | 11 | 10 (5 – 11) | 11 | 9 (5 – 14) |
| US | 7 | 6 (-1 – 7) | 6 | 4 (0 – 7) |
| Belgium | 5 | 6 (6 – 7) | 3 | 6 (3 – 9) |
| Italy | 4 | 6 (1 – 7) | 4 | 4 (1 – 6) |
| Hungary | 4 | 7 (0 – 7) | 4 | 8 (5 – 10) |
| United Kingdom | 1 | 1 (1 – 1) | -2 | 3 (0 – 6) |
| Spain | 0 | 0 (0 – 0) | 0 | 3 (1 – 5) |
| Denmark | -1 | 0 (0 – 1) | -1 | 1 (-1 – 3) |
| Netherlands | -3 | 5 (5 – 6) | -3 | 4 (2 – 7) |
| Portugal | 10 | 5 (3 – 7) | 11 | 5 (3 – 9) |
| France | poor fit | 3 (3 – 3) | 7 | 3 (2 – 4) |
| Sweden | -17 | -6 (-9 – -2) | -18 | -5 (-11 – 0) |

In order to determine the lag we attempted to find fits of the function (1) to all cases time-series and all death time-series as available on the John Hopkins University (JHU) server on December 6, 2020. In order to guarantee statistical significance of the data we restricted the analysis to these time series where the total number of deaths was larger than 500 on the 180th day of the epidemic. Here “180th day of the epidemic” is July 19, corresponding to the 180th day of the time series available on the JHU server, While most of the fits were optically satisfactory, many of them had large uncertainties in the values of the parameters. This made them useless for any significant analysis. To address this we imposed a threshold of 30 for the sum of our “fit quality” parameters (cf. [2] for the definition) of cases time-series and death time-series. We were left with the list of countries given in Table 1, see also Figure 4. The countries in the table are therefore those for which there were at least 500 casualties due to Covid-19 reported on July 19, 2020, and for which parameters exist so that the function *I* gives meaningful fits for both the total cases and total death time series. All these countries had a clear first wave ending before the 180th day of the epidemic. The first data-column of the table is the lag determined in this way, in days.

**Figure 4:**
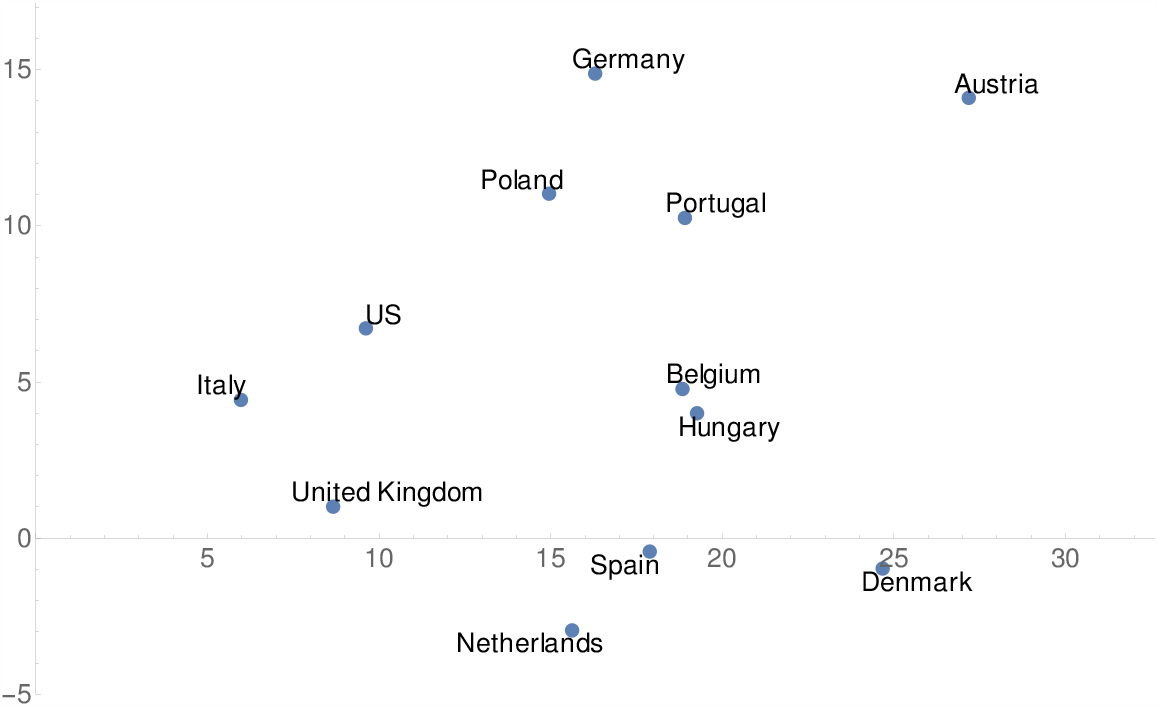
Lag between days of the maximum number of casualties and the maximum number of daily infections for the first wave of Covid-19 determined from the function *I*, ordered with decreasing quality of the fits to the function *I*. Romania and Sweden have not been included in the plot, because of the width of the error interval when calculated by the integral overlap method.

As an example, we show the fits for Romania (largest lag) and Sweden (largest negative lag) in Figure 5. The fits satisfy our quality criterion, but Figure 3 makes it clear that no clear-cut lag can be determined from the data for these countries.

**Figure 5:**
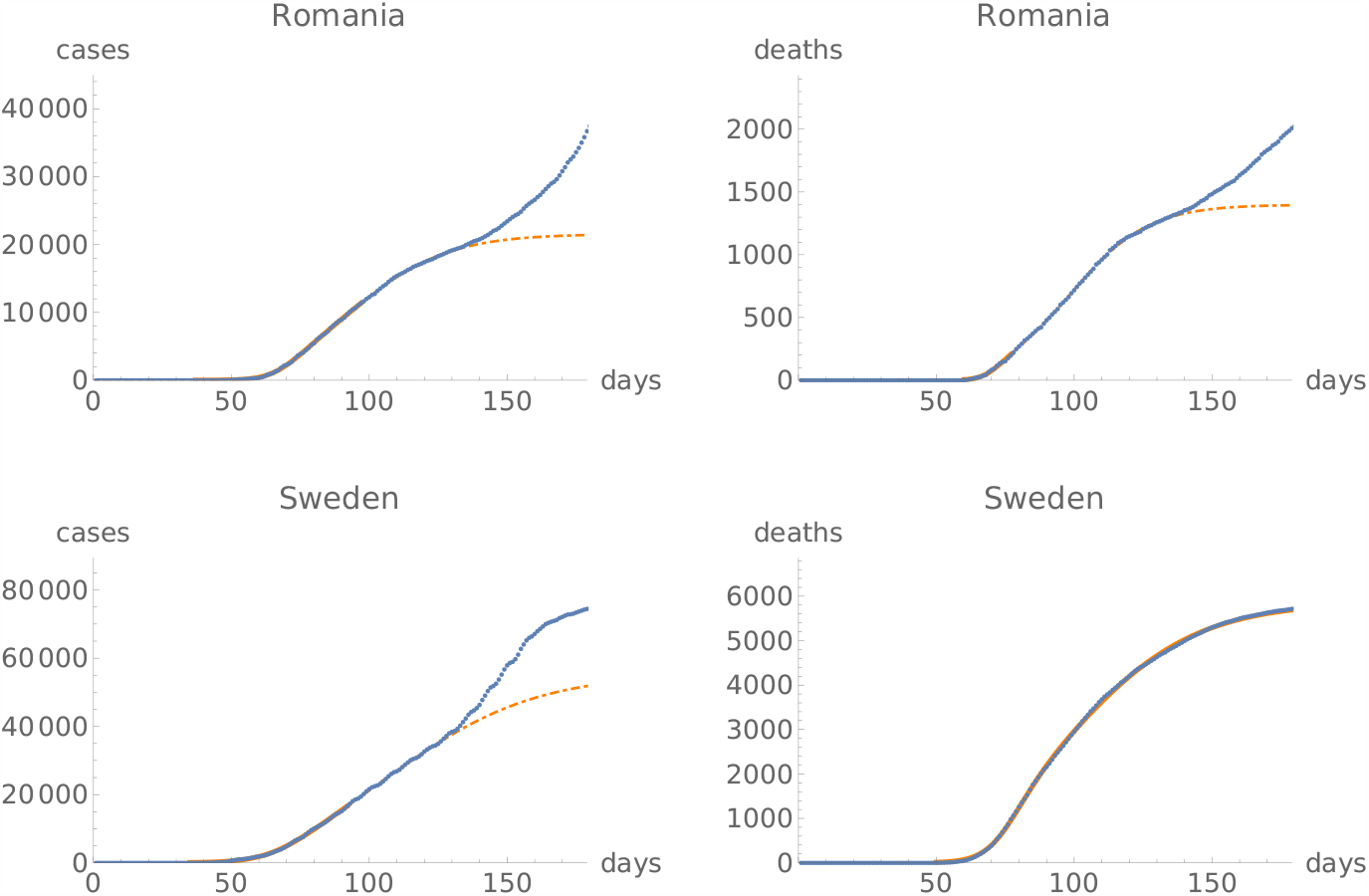
Fits of the function *I* to the first phase of the Covid-19 pandemic for Romania and Sweden. A clear transition from one phase of the epidemic to another for Romania can be seen in Figure 3, which is perhaps less clear on the plots here. Similarly a clear transition to a second phase for the death time-series of Sweden can be seen in Figure 3, but less so in the cases time-series of Sweden, or in the plots here. We draw attention to different scales on different plots.

In Figure 4 the data points are ordered by decreasing quality of the fit, as measured by the “fit quality parameter” described in detail in [2]. Thus the smaller the abscissa, the better is the approximation of the data by the function *I*. The fits to the data for the time series with the best fits, namely Italy and United Kingdom, have already been seen in Figures 1 and 2.

As we did not expect negative lags, i.e. time series where the maximum number of daily deaths precedes the day of maximum number of daily infections, it became important to devise an alternative systematic method, other than the above and the visual inspection, to determine the lag. For this we used an “integral overlap method”, illustrated in Figure 6, which proceeds as follows. Let *c*(*t*) denote the number of new infections on day *t*, and let *d*(*t*) denote the number of deaths on the same day. Consider the sum

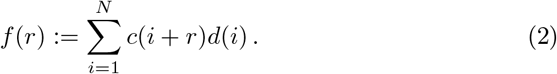

**Figure 6:**
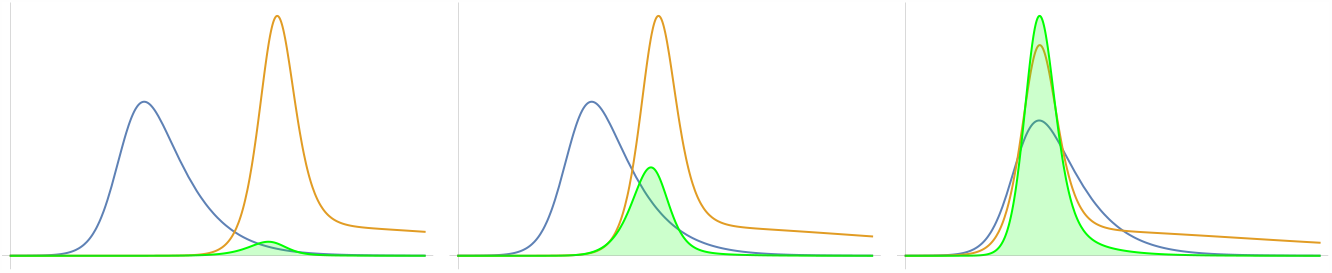
Illustration of the integral overlap method: Consider the blue and orange functions in the left plot. The green function is their product, and the shaded area under the green curve is the integral equivalent of the value *f* (0) of the function *f* of (2). In the middle plot the orange function has been shifted to the left by some value *r*, leading to an increased area, equal to *f* (*r*), under the new product curve. The area under the graph of the product function is maximised by shifting the orange function as shown in the right plot.

(Obviosly, *N* and the largest value of *r* need to be chosen so that *N* + *r* does not exceed the length of the time series; in our analysis of the first 180 days of the epidemic we used *N* = 152 and *r* ∈ [−28, 28], with *c*(*t*) extended by zero for negative values of *t*.) Keeping in mind that both *c* and *d* should be positive, one expects *f* (*r*) to be maximised when the shift *r* between the functions *c* and *d* is such that the maximum of the function *c* overlaps with the maximum of the function *d*. So finding the value of *r* for which the function *f* attains its maximum determines the lag.

The results of this analysis, applied to the first 180 days of the Covid-19 epidemic, are presented in the third column of Table 1. There the intervals for which *f* (*r*) was larger than .99 of its maximum value are also indicated. The length of such intervals provides a joint measure of the widths of the peaks of maximum numbers of new infections and new deaths.

One of the problems arising in the integral-overlap method is that outlying values of the time series sometimes unduly influence the location of the maximum of the function *f*. In order to avoid this we repeated the analysis using time series averaged over seven consecutive days. Here seven days have been chosen to take into account effects arising from different reporting habits during weekends. The results are shown in the last column of Table 1. We will refer to this method as the average-data-integral (ADI) method.

We split Table 1 into a first part, where the lags are consistent with the lags determined from the function *I*, and a second one where the values differ. By inspection of the data (see Figure 7), the difference between the lag determined from the function *I* and the integral average for Netherlands and Portugal is due to some extreme outliers in the time series. One can most likely get a better estimation of the lags by removing outliers country by country, but we did not attempt this.

**Figure 7:**
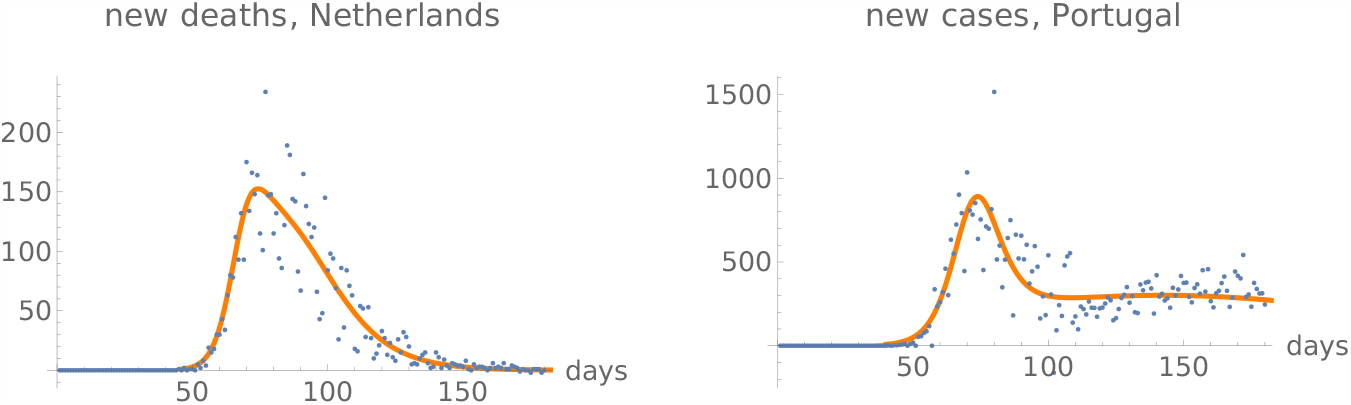
Daily death time-series for Netherlands and daily infections time-series for Portugal, with the derivative of the best fit of the function *I* indicated in orange.

The ADI method can be applied to any time series, without the need to have a significant fit of the function *I*. We used the method to determine the lags for all time series from the JHU server for which there was a clear first phase of the Covid-19 pandemic which lasted less than 180 days from February 22 (the beginning of the data on the JHU server), thus ending before July 19, 2020.

A histogram of the lags can be found in Figure 8. The histogram hints at two peaks, centered around day five and day ten. These arise from a peak around ten that is present in the histogram for the US counties, and a peak around five when the US counties are removed from the analysis. However, a somewhat different story is told when a weighted average is determined, where the weights are inversely proportional to the width of the peak, determined as the region where the integral-overlap function *f* of Equation (2) is larger than 99 % of its maximum. One then finds a weighted mean of 7.5± 0.5 for US counties, essentially consistent with a weighted mean of 6.7± 0.4 days for all remaining time series, with an overall weighted mean of

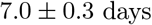

for all time series which had a clear end of the first phase of the epidemic before July 19, 2020, with at least 500 Covid-19 deaths on that date.

**Figure 8:**
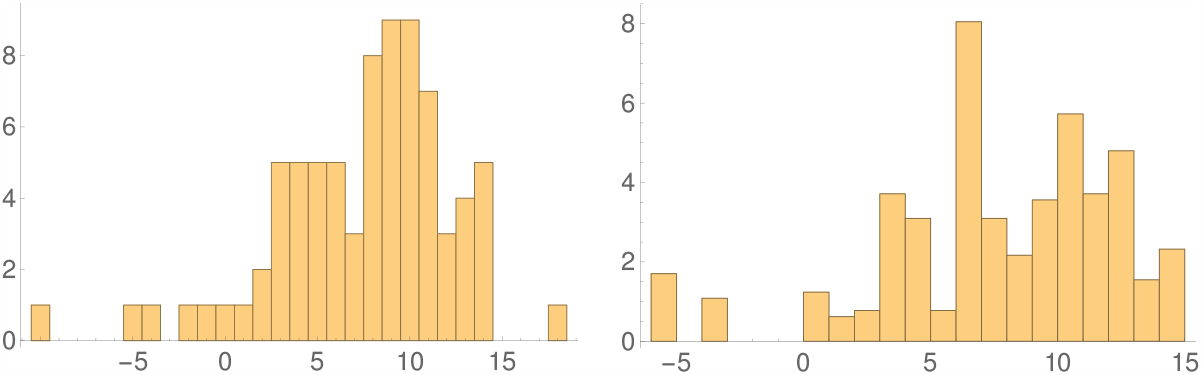
The histograms of the lags between days of the maximum number of casualties and the maximum number of daily infections for the first wave of Covid-19, as determined by the “average-data-integral” method. The first histogram is direct, the second is obtained by weighting the data inversely proportionally to the width of the peak.

The leftermost outlier on the histogram, with a negative lag of −10 days arises from the data of the county Hennepin in Minessota, while the rightermost one with a lag of 18 days is determined from the time series for Ecuador. The associated time series are seen in Figure 9. These time series suggest strongly that no clear lag can be determined for them in any case.

**Figure 9:**
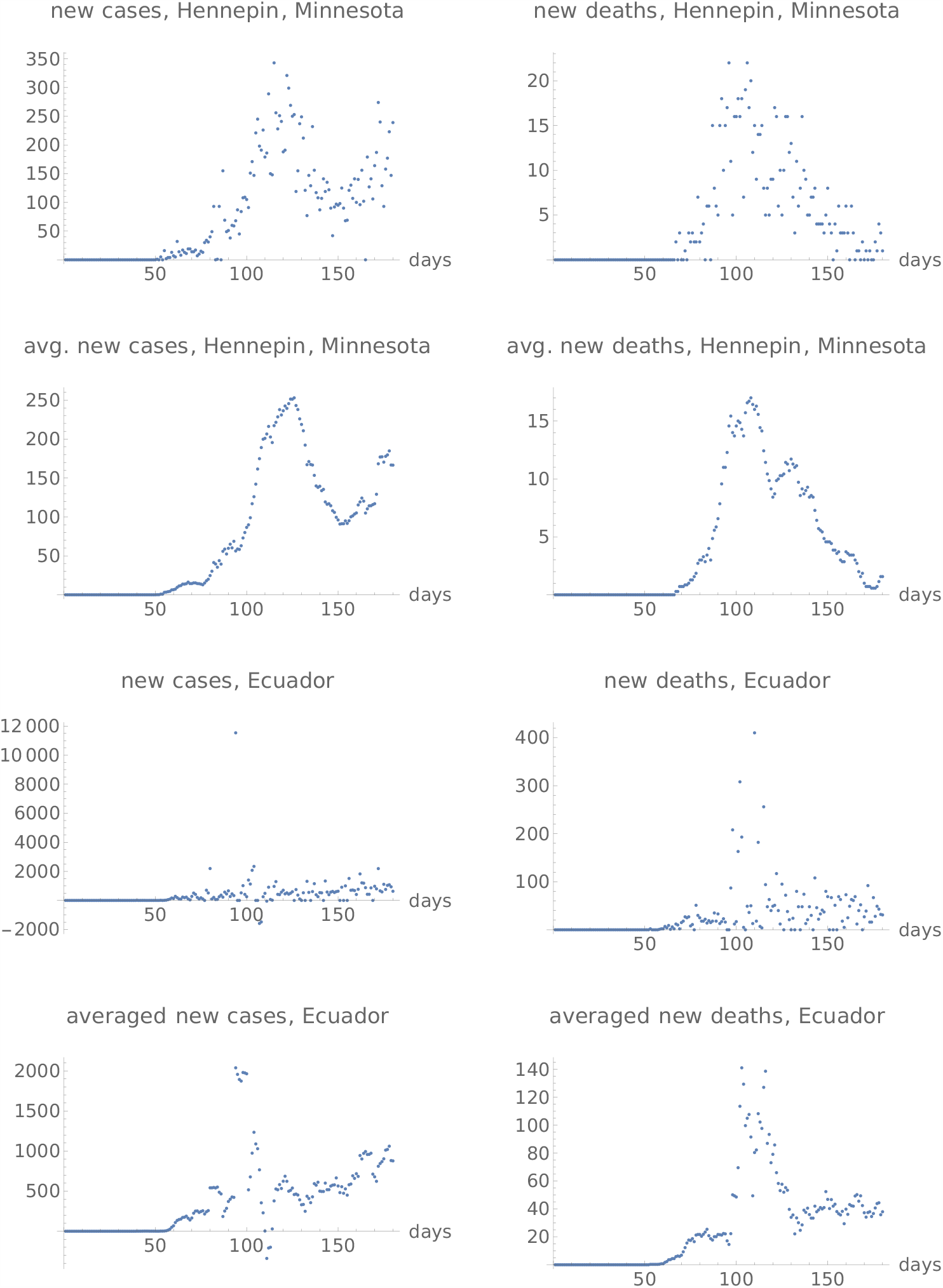
Time series for the outliers in the histogram of Figure 8, namely Hennepin, Minnesota and Ecuador.

Summarising, we have analysed the cases-and-deaths lags in the first phase of the Covid-19 pandemic. We have found that the peaks of deaths preceded the peaks of infections in some cases, witnessing inadequateness of measures taken to protect the most vulnerable population. This can, however be biased by the age pyramid of the population, and by local usages concerning when new cases are reported and when new deaths are reported.

While the precise lags are not evident for some countries, they provide an objective quantitative indicator of the effectiveness of preventive measures taken whenever they are clearly determined.

Our results are consistent with the folklore knowledge, that countries such as Sweden did not manage to protect the most fragile part of their population. They confirm that measures taken in Germany were effective in that respect. The large uncertainty in the lag for US is consistent with the lack of global preventive measures taken.

There exists another obvious indicator of the effectiveness of measures taken to protect a population, namely the number of deaths relative to the size of the population. We note that we did not find any obvious correlation between this indicator and the lags determined by our methods.

All fits and integral averages used for the analysis here can be found in the Supplementary Material.

## Data Availability

Data publicly available on the server of the John Hopkins University

