## Supplementary Materials for "On the lag between deaths and infections in the first phase of the Covid-19 pandemic"

**supplementary material**

### Introduction

We present here the time series and fits used for the analysis in the main accompanying paper. The analysis is concerned with the first 180 days of the epidemic, using data available on the John Hopkins University server on December 6, 2020. The best fits, and the data of the end of the first phase of the epidemic, are obtained as described in https://www.medrxiv.org/content/early/2020/08/30/2020.08.24.20181214.

### List of countries and states used in our analysis

##### a) global

Out[ ]= {{, Austria}, {, Belgium}, {, Denmark}, {, France}, {, Germany}, {, Hungary}, {, Ireland}, {, Italy}, {, Netherlands}, {, Poland}, {, Portugal}, {, Romania}, {, Spain}, {, Sweden}, {, United Kingdom}, {, US}, {, Afghanistan}, {, Algeria}, {, Argentina}, {, Armenia}, {, Bangladesh}, {, Bolivia}, {, Brazil}, {, Chile}, {, Colombia}, {, Dominican Republic}, {, Ecuador}, {, Egypt}, {, Guatemala}, {, Honduras}, {, India}, {, Indonesia}, {, Iran}, {, Iraq}, {, Japan}, {, Mexico}, {, Moldova}, {, Nigeria}, {, Pakistan}, {, Panama}, {, Peru}, {, Philippines}, {, Russia}, {, Saudi Arabia}, {, South Africa}, {, Sudan}, {, Switzerland}, {, Turkey}, {, Ukraine}, {Hubei, China}, {Ontario, Canada}, {Quebec, Canada}}

### b) US

Out[ ]= {{Maricopa, Arizona}, {Los Angeles, California}, {Riverside, California}, {Fairfield, Connecticut}, {Hartford, Connecticut}, {New Haven, Connecticut}, {District of Columbia, District of Columbia}, {Miami-Dade, Florida}, {Palm Beach, Florida}, {Cook, Illinois}, {Marion, Indiana}, {Orleans, Louisiana}, {Baltimore, Maryland}, {Montgomery, Maryland}, {Prince George's, Maryland}, {Bristol, Massachusetts}, {Essex, Massachusetts}, {Hampden, Massachusetts}, {Middlesex, Massachusetts}, {Norfolk, Massachusetts}, {Plymouth, Massachusetts}, {Suffolk, Massachusetts}, {Worcester, Massachusetts}, {Macomb, Michigan}, {Oakland, Michigan}, {Wayne, Michigan}, {Hennepin, Minnesota}, {St. Louis, Missouri}, {Clark, Nevada}, {Bergen, New Jersey}, {Camden, New Jersey}, {Essex, New Jersey}, {Hudson, New Jersey}, {Mercer, New Jersey}, {Middlesex, New Jersey}, {Monmouth, New Jersey}, {Morris, New Jersey}, {Ocean, New Jersey}, {Passaic, New Jersey}, {Somerset, New Jersey}, {Union, New Jersey}, {Bronx, New York}, {Erie, New York}, {Kings, New York}, {Nassau, New York}, {New York, New York}, {Queens, New York}, {Richmond, New York}, {Rockland, New York}, {Suffolk, New York}, {Westchester, New York}, {Bucks, Pennsylvania}, {Delaware, Pennsylvania}, {Montgomery, Pennsylvania}, {Philadelphia, Pennsylvania}, {Providence, Rhode Island}, {Harris, Texas}, {Fairfax, Virginia}, {King, Washington}}

### Plots, fits and integral averages, not including US counties

#### Austria

lag from integral overlap between 12 and 13 days, maximum overlap on day 13

lag from integral overlap, using data averaged over 7 days, between 11 and 14 days, maximum overlap on day 12

lag from fits to the function I: 14.1083 days

lag from fits to the function I (averaged data): 14.1546 days

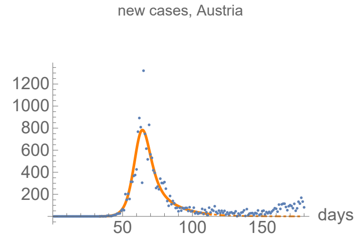


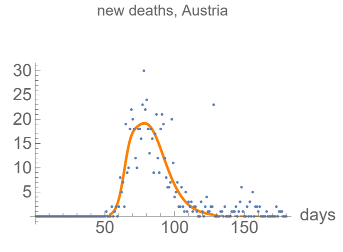


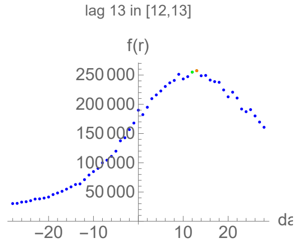

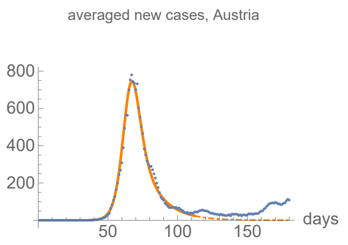


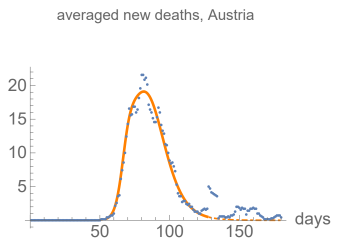


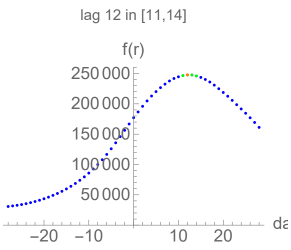


#### Parameters of fits to cases data.

The best fit ends on Wed 13 May 2020.

| α | β | k | τ | σ |
|---|---|---|---|---|
| 0.322±0.0071 | 0.0705±0.00793 | 16200.±75. | 28.7±0.234 | 22.3±1.18 |

| badness | | α | β | k | τ | σ | quality |
|---|---|---|---|---|---|---|---|
| 2.1% | error | 2.2% | 11.% | 0.46% | 0.81% | 5.3% | 12.6% |

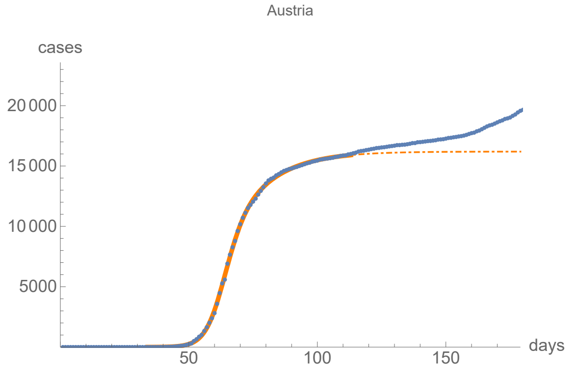


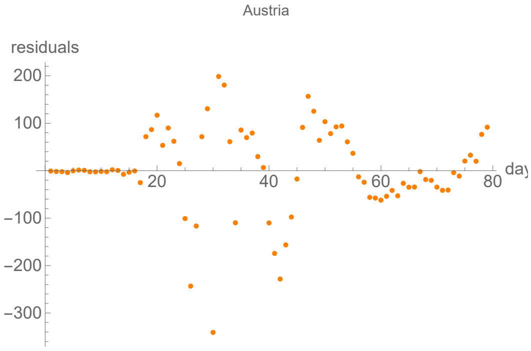


#### Parameters of fits to averaged cases data.

The best fit ends on Tue 12 May 2020.

| α | β | k | τ | σ |
|---|---|---|---|---|
| 0.31±0.00351 | 0.0736±0.00376 | 16100.±41.7 | 28.6±0.142 | 22.7±0.633 |

| badness | | α | β | k | τ | σ | quality |
|---|---|---|---|---|---|---|---|
| 0.65% | error | 1.1% | 5.1% | 0.26% | 0.5% | 2.8% | 6.09% |

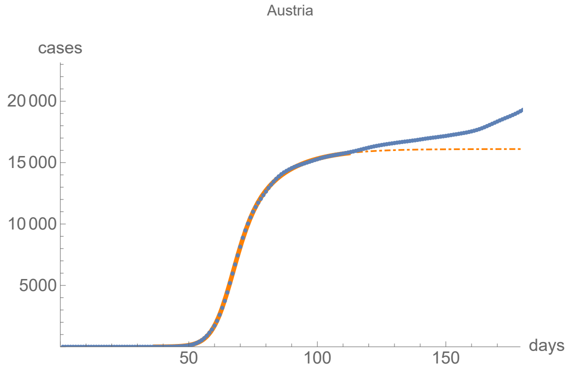


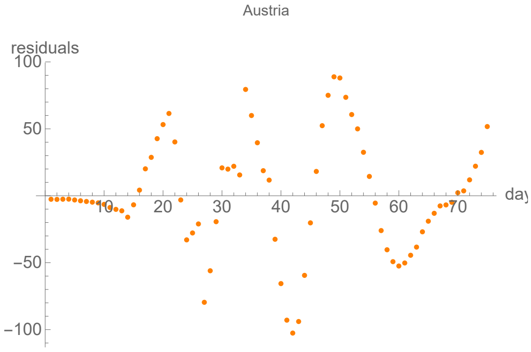


#### Parameters of fits to death data.

The best fit ends on Wed 27 May 2020.

| α | β | k | τ | σ |
|---|---|---|---|---|
| 0.409±0.0283 | 0.113±0.00206 | 642.±1.54 | 26.6±0.136 | 11.1±0.365 |

| badness | | α | β | k | τ | σ | quality |
|---|---|---|---|---|---|---|---|
| 1.8% | error | 6.9% | 1.8% | 0.24% | 0.51% | 3.3% | 14.6% |

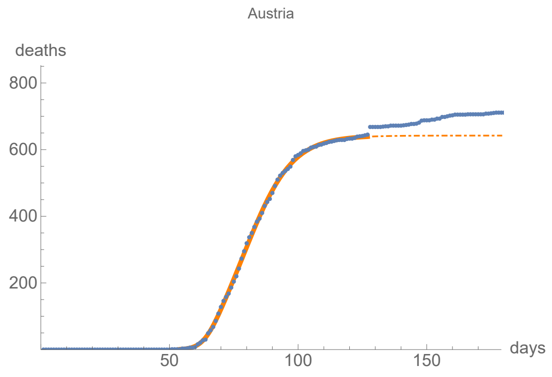


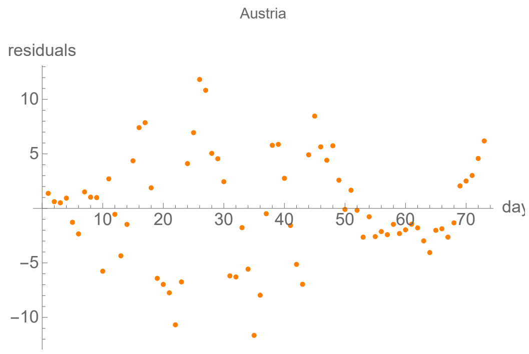


#### Parameters of fits to averaged death data.

The best fit ends on Wed 27 May 2020.

| α | β | k | τ | σ |
|---|---|---|---|---|
| 0.39±0.0139 | 0.113±0.00112 | 641.±0.878 | 28.5±0.0742 | 12.7±0.206 |

| badness | | α | β | k | τ | σ | quality |
|---|---|---|---|---|---|---|---|
| 1.% | error | 3.6% | 1.% | 0.14% | 0.26% | 1.6% | 7.62% |

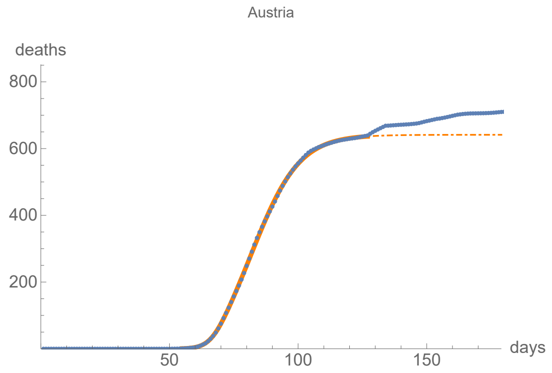


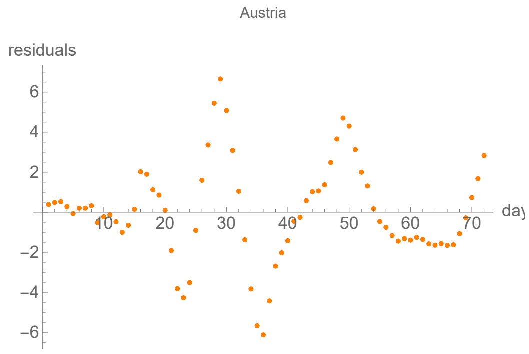


### Belgium

lag from integral overlap between 6 and 7 days, maximum overlap on day 6

lag from integral overlap, using data averaged over 7 days, between 3 and 9 days, maximum overlap on day 6

lag from fits to the function I: 4.76359 days

lag from fits to the function I (averaged data): 3.14838 days

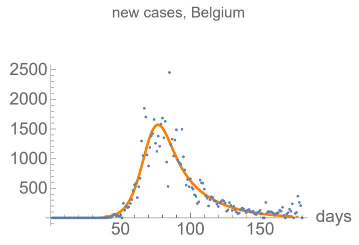


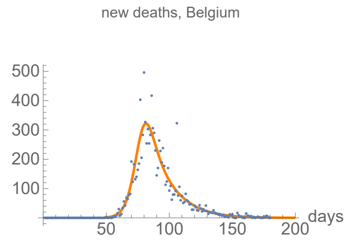


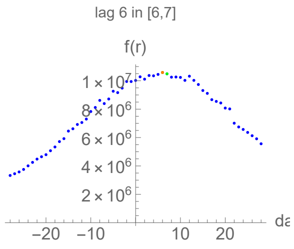


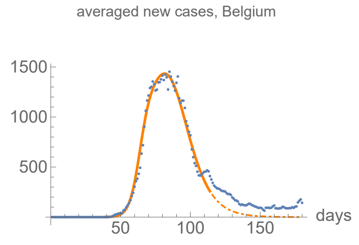


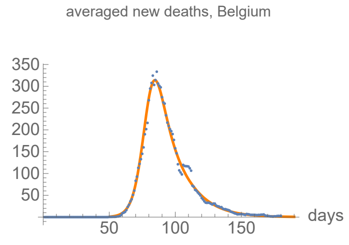


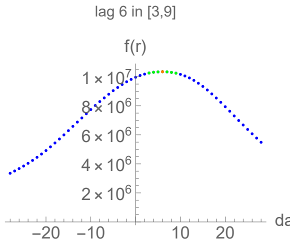

#### Parameters of fits to cases data.

The best fit ends on Sat 11 Jul 2020.

| α | β | k | τ | σ |
|---|---|---|---|---|
| 0.188±0.00417 | 0.0473±0.00503 | 62200.±165. | 33.6±0.461 | 33.6±1.14 |

| badness | | α | β | k | τ | σ | quality |
|---|---|---|---|---|---|---|---|
| 2.6% | error | 2.2% | 11.% | 0.27% | 1.4% | 3.4% | 12.% |

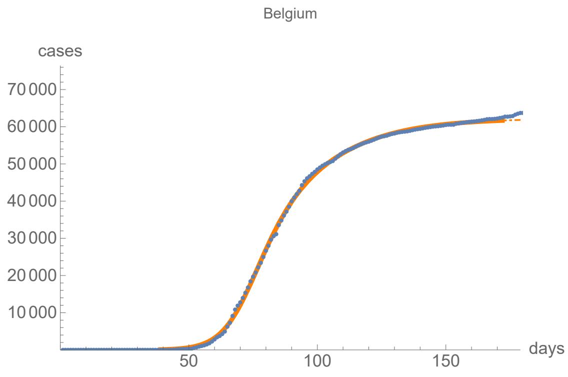


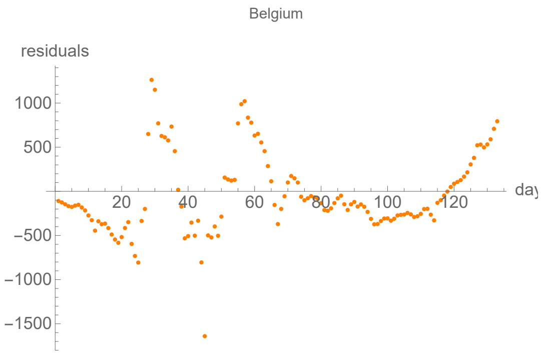


#### Parameters of fits to averaged cases data.

The best fit ends on Thu 14 May 2020.

| α | β | k | τ | σ |
|---|---|---|---|---|
| 0.309±0.0109 | 0.0965±0.00143 | 55800.±187. | 44.6±0.0882 | 25.5±0.317 |

| badness | | α | β | k | τ | σ | quality |
|---|---|---|---|---|---|---|---|
| 0.65% | error | 3.5% | 1.5% | 0.34% | 0.2% | 1.2% | 7.44% |

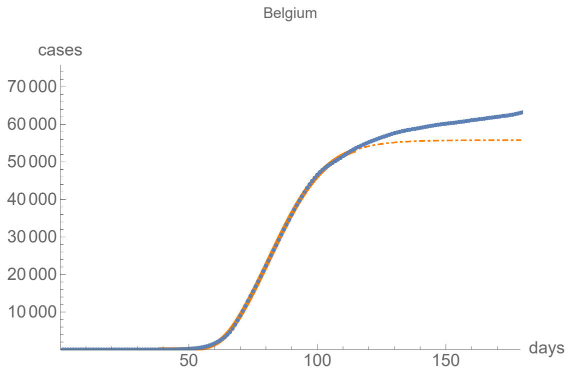


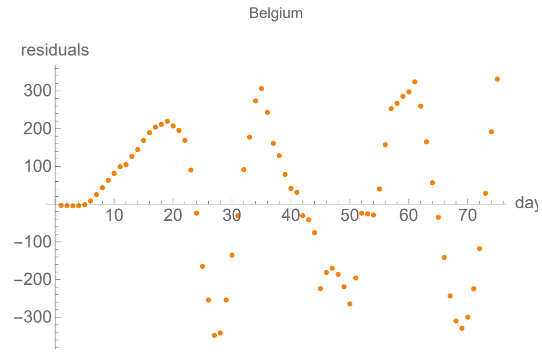


#### Parameters of fits to death data.

The best fit ends on Fri 7 Aug 2020.

| α | β | k | τ | σ |
|---|---|---|---|---|
| 0.25±0.00353 | 0.0648±0.00106 | 9820.±6.9 | 30.7±0.451 | 29.±0.205 |

| badness | | α | β | k | τ | σ | quality |
|---|---|---|---|---|---|---|---|
| 1.6% | error | 1.4% | 1.6% | 0.07% | 1.5% | 0.71% | 6.86% |

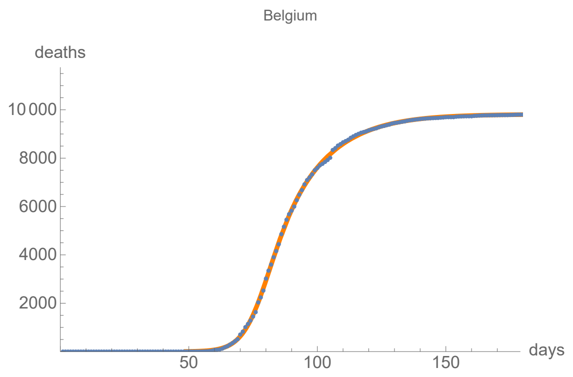


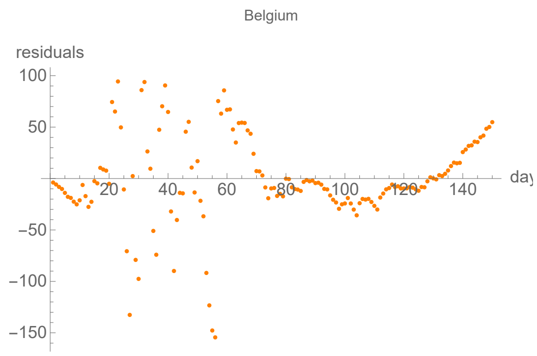


#### Parameters of fits to averaged death data.

The best fit ends on Wed 29 Jul 2020.

| α | β | k | τ | σ |
|---|---|---|---|---|
| 0.246±0.00195 | 0.0659±0.00245 | 9800.±4.29 | 29.7±0.125 | 32.±0.255 |

| badness | | α | β | k | τ | σ | quality |
|---|---|---|---|---|---|---|---|
| 0.82% | error | 0.79% | 3.7% | 0.044% | 0.42% | 0.8% | 3.71% |

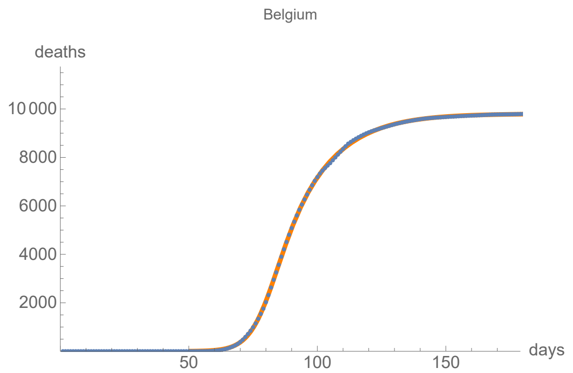


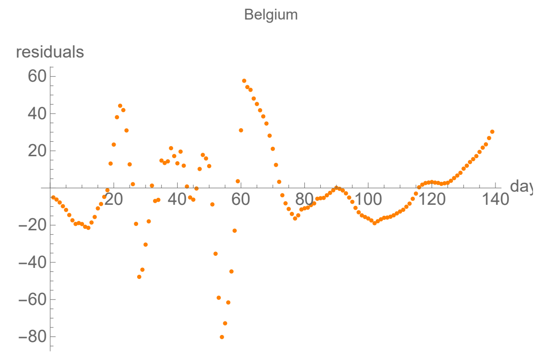


### Denmark

lag from integral overlap between 0 and 1 days, maximum overlap on day 0

lag from integral overlap, using data averaged over 7 days, between −1 and 3 days, maximum overlap on day 1

lag from fits to the function I: −0.98422 days

lag from fits to the function I (averaged data): −0.883999 days

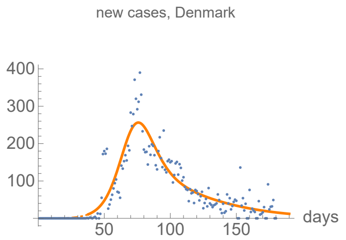


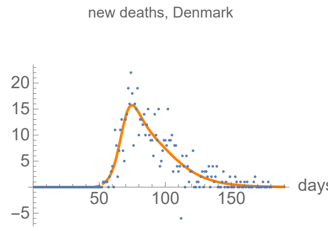


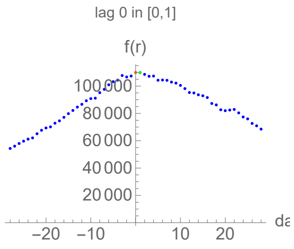


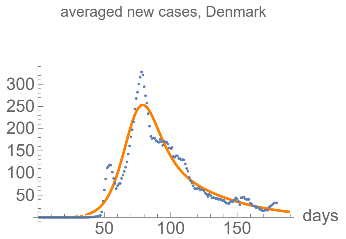


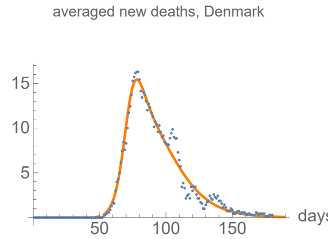


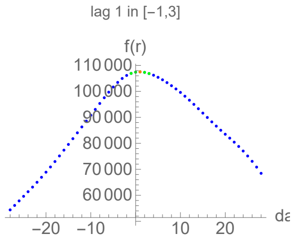

#### Parameters of fits to cases data.

The best fit ends on Wed 29 Jul 2020.

| α | β | k | τ | σ |
|---|---|---|---|---|
| 0.142±0.00374 | 0.029±0.00457 | 13800.±85.8 | 33.6±0.559 | 33.6±1.54 |

| badness | | α | β | k | τ | σ | quality |
|---|---|---|---|---|---|---|---|
| 2.4% | error | 2.6% | 16.% | 0.62% | 1.7% | 4.6% | 14.3% |

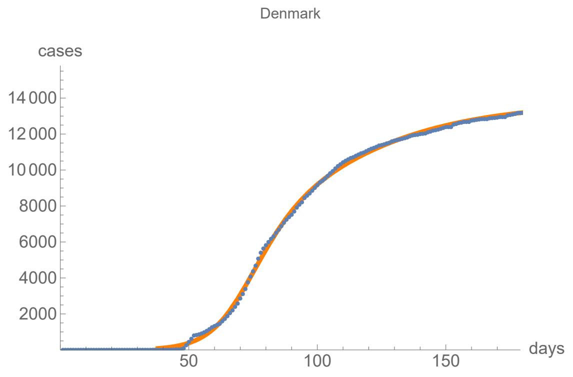


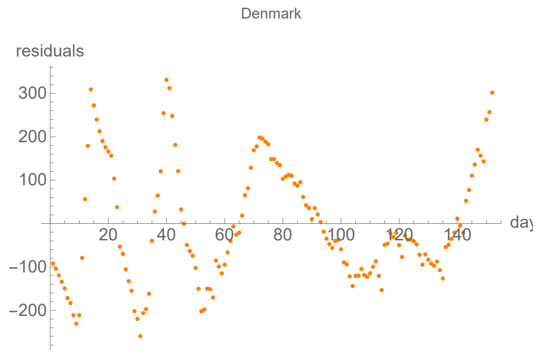


#### Parameters of fits to averaged cases data.

The best fit ends on Tue 28 Jul 2020.

| α | β | k | τ | σ |
|---|---|---|---|---|
| 0.141±0.00323 | 0.0299±0.00389 | 13700.±75.3 | 35.4±0.519 | 35.4±1.39 |

| badness | | α | β | k | τ | σ | quality |
|---|---|---|---|---|---|---|---|
| 1.7% | error | 2.3% | 13.% | 0.55% | 1.5% | 3.9% | 12.% |

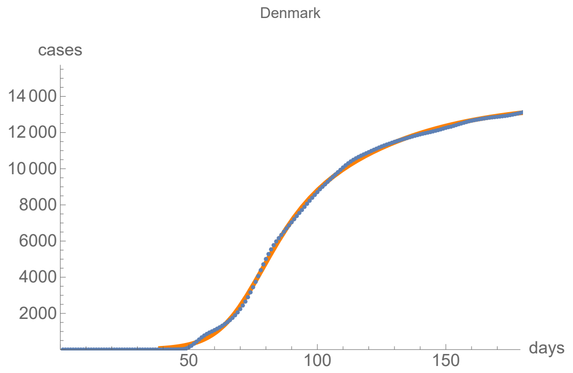


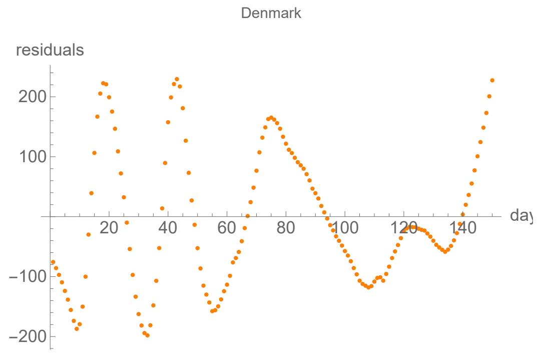


#### Parameters of fits to death data.

The best fit ends on Wed 29 Jul 2020.

| α | β | k | τ | σ |
|---|---|---|---|---|
| 0.261±0.0082 | 0.0628±0.00109 | 610.±0.821 | 29.5±0.327 | 17.±0.298 |

| badness | | α | β | k | τ | σ | quality |
|---|---|---|---|---|---|---|---|
| 2.5% | error | 3.1% | 1.7% | 0.13% | 1.1% | 1.8% | 10.3% |

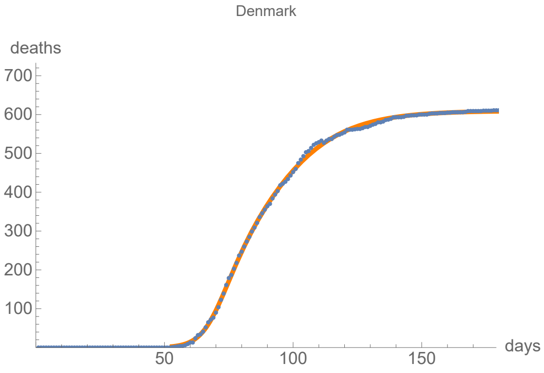


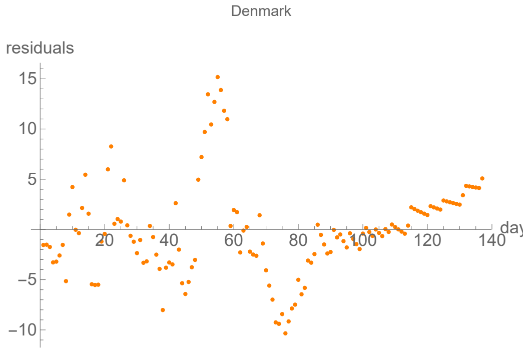


#### Parameters of fits to averaged death data.

The best fit ends on Tue 28 Jul 2020.

| α | β | k | τ | σ |
|---|---|---|---|---|
| 0.254±0.00672 | 0.0628±0.00095 | 609.±0.726 | 30.5±0.289 | 17.8±0.269 |

| badness | | α | β | k | τ | σ | quality |
|---|---|---|---|---|---|---|---|
| 2.% | error | 2.7% | 1.5% | 0.12% | 0.95% | 1.5% | 8.77% |

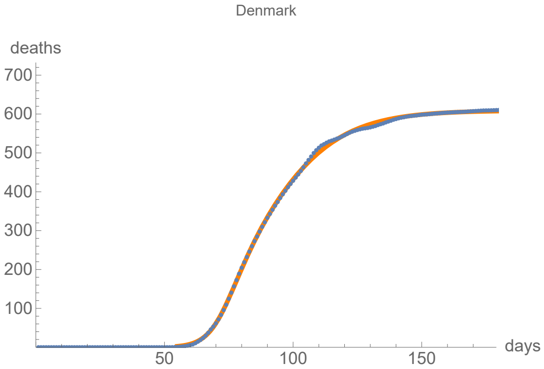


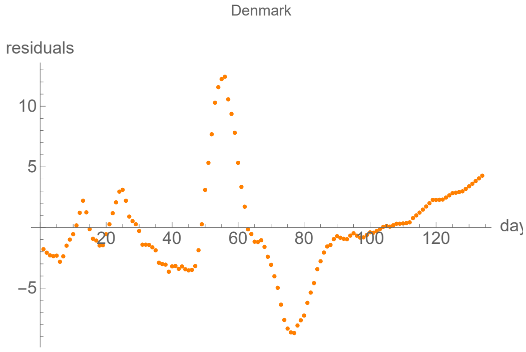


### France

lag from integral overlap between 3 and 3 days, maximum overlap on day 3

lag from integral overlap, using data averaged over 7 days, between 2 and 4 days, maximum overlap on day 3

lag from fits to the function I: 4.01184 days

lag from fits to the function I (averaged data): 7.40324 days

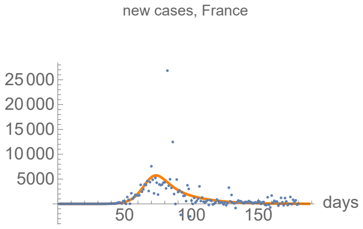


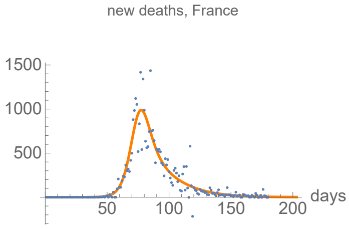


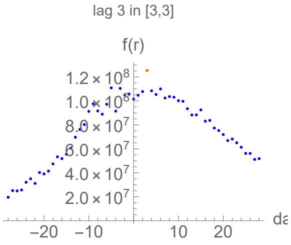


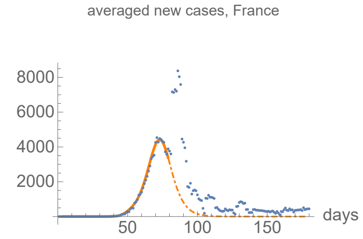


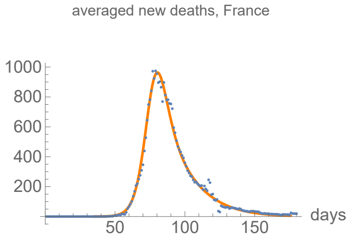


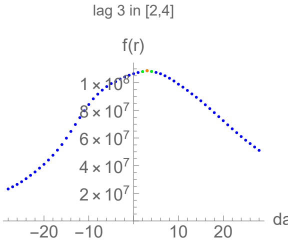

#### Parameters of fits to cases data.

The best fit ends on Mon 27 Jul 2020.

| α | β | k | τ | σ |
|---|---|---|---|---|
| 0.217±0.0162 | 0.0546±0.0203 | 196000.±987. | 67.8±1.26 | 67.9±3.18 |

| badness | | α | β | k | τ | σ | quality |
|---|---|---|---|---|---|---|---|
| 7.4% | error | 7.5% | 37.% | 0.5% | 1.9% | 4.7% | 29.6% |

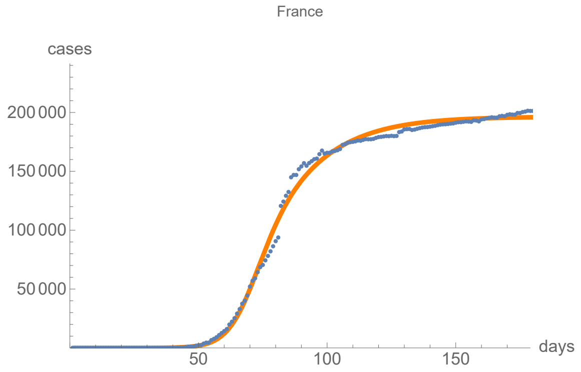


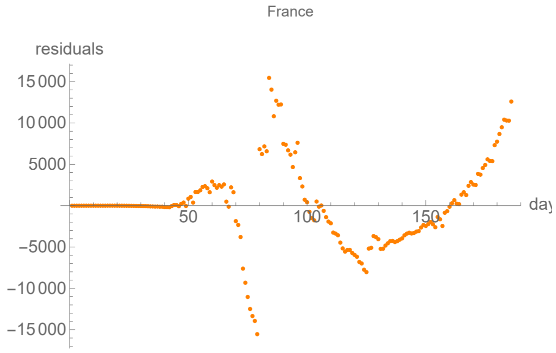


#### Parameters of fits to averaged cases data.

The best fit ends on Thu 9 Apr 2020.

| α | β | k | τ | σ |
|---|---|---|---|---|
| 0.482±0.0313 | 0.175±0.000988 | 101000.±493. | 69.±0.0597 | 43.9±0.372 |

| badness | | α | β | k | τ | σ | quality |
|---|---|---|---|---|---|---|---|
| 0.56% | error | 6.5% | 0.56% | 0.49% | 0.087% | 0.85% | 9.04% |

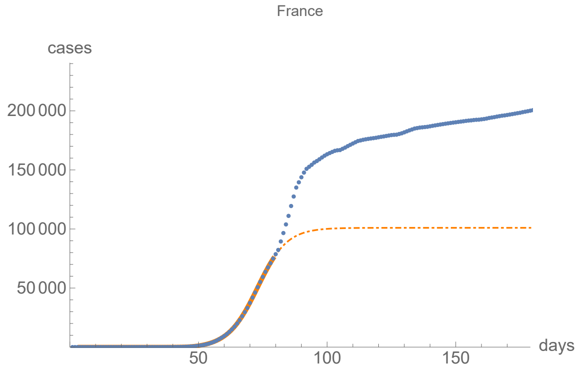


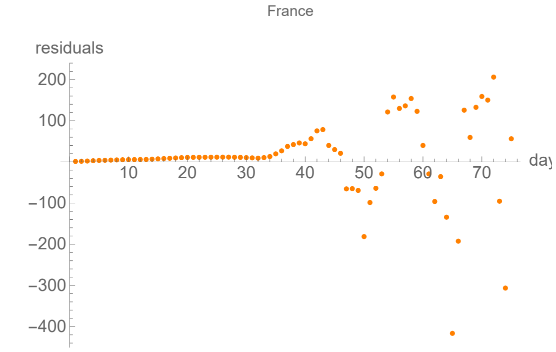


#### Parameters of fits to death data.

The best fit ends on Tue 11 Aug 2020.

| α | β | k | τ | σ |
|---|---|---|---|---|
| 0.242±0.00333 | 0.0553±0.00426 | 30100.±24.5 | 39.8±0.177 | 38.5±0.527 |

| badness | | α | β | k | τ | σ | quality |
|---|---|---|---|---|---|---|---|
| 2.2% | error | 1.4% | 7.7% | 0.082% | 0.44% | 1.4% | 6.9% |

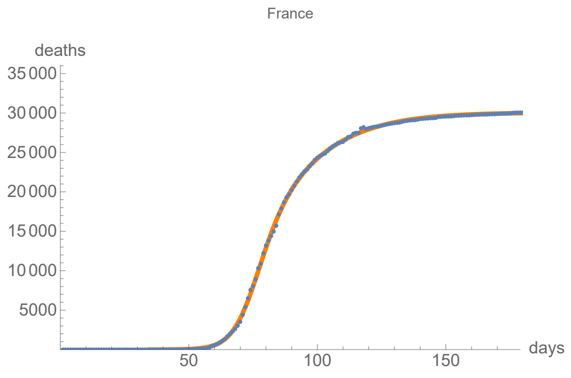


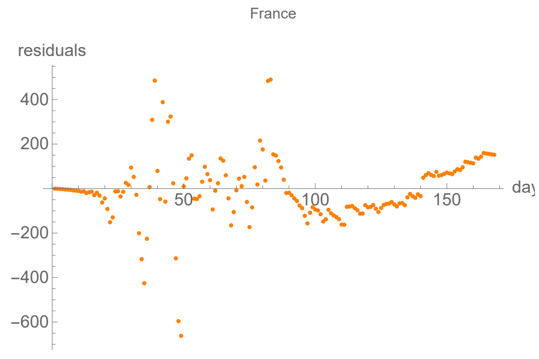


#### Parameters of fits to averaged death data.

The best fit ends on Tue 14 Jul 2020.

| α | β | k | τ | σ |
|---|---|---|---|---|
| 0.243±0.00199 | 0.0591±0.0025 | 29900.±19.5 | 42.2±0.116 | 42.8±0.288 |

| badness | | α | β | k | τ | σ | quality |
|---|---|---|---|---|---|---|---|
| 0.65% | error | 0.82% | 4.2% | 0.065% | 0.28% | 0.67% | 3.31% |

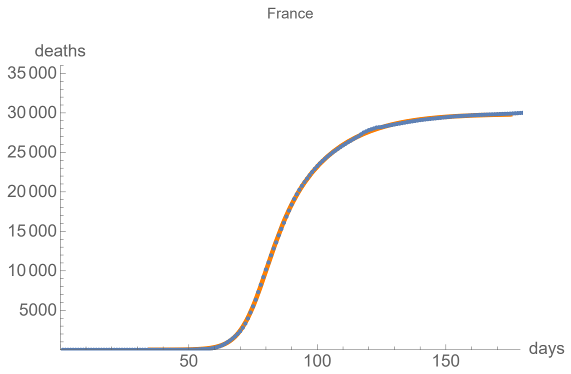


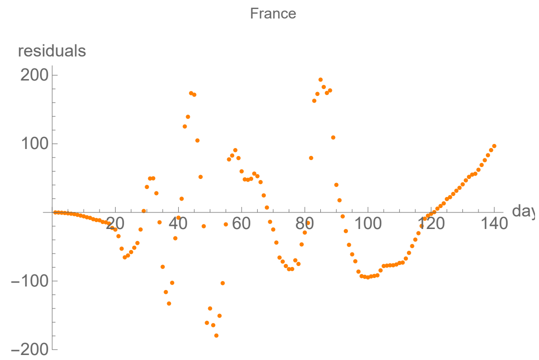


### Germany

lag from integral overlap between 13 and 13 days, maximum overlap on day 13

lag from integral overlap, using data averaged over 7 days, between 11 and 16 days, maximum overlap on day 13

lag from fits to the function I: 14.8609 days

lag from fits to the function I (averaged data): 16.5692 days

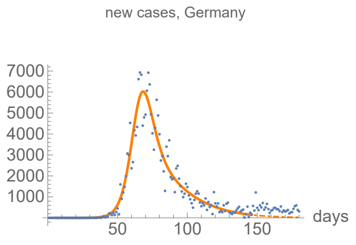


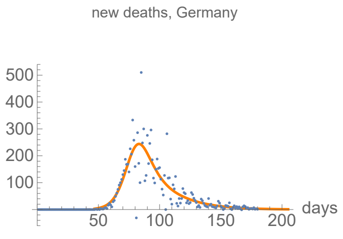


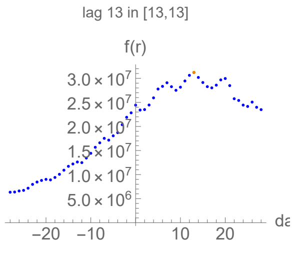


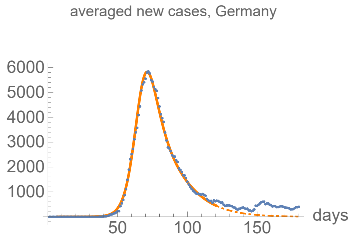


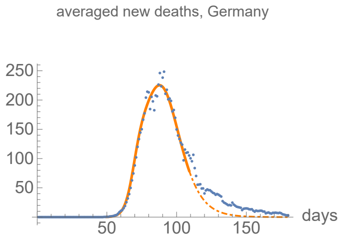


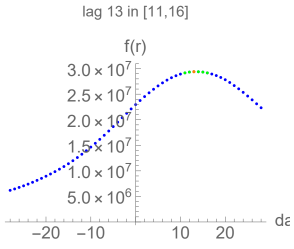

#### Parameters of fits to cases data.

The best fit ends on Mon 15 Jun 2020.

| α | β | k | τ | σ |
|---|---|---|---|---|
| 0.241±0.003 | 0.0521±0.00373 | 190000.±355. | 59.6±0.161 | 59.6±0.445 |

| badness | | α | β | k | τ | σ | quality |
|---|---|---|---|---|---|---|---|
| 1.2% | error | 1.2% | 7.2% | 0.19% | 0.27% | 0.75% | 4.87% |

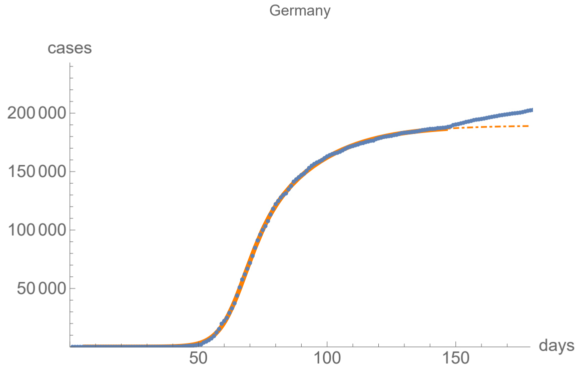


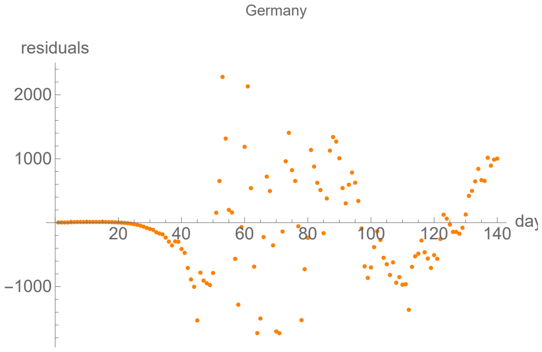


#### Parameters of fits to averaged cases data.

The best fit ends on Wed 20 May 2020.

| α | β | k | τ | σ |
|---|---|---|---|---|
| 0.247±0.00185 | 0.0625±0.00211 | 184000.±381. | 60.5±0.129 | 63.1±0.233 |

| badness | | α | β | k | τ | σ | quality |
|---|---|---|---|---|---|---|---|
| 0.46% | error | 0.75% | 3.4% | 0.21% | 0.21% | 0.37% | 2.61% |

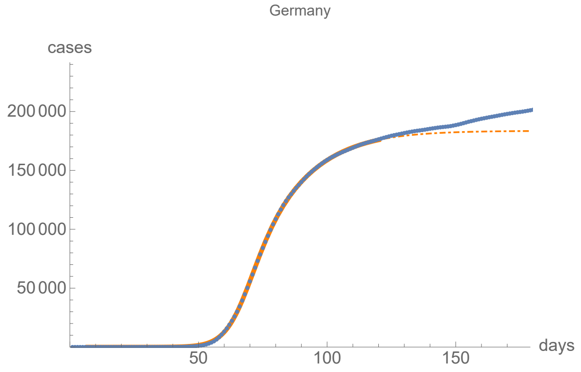


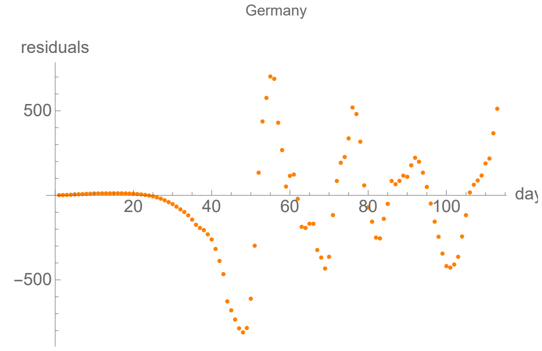


#### Parameters of fits to death data.

The best fit ends on Thu 13 Aug 2020.

| α | β | k | τ | σ |
|---|---|---|---|---|
| 0.191±0.00351 | 0.0481±0.00428 | 9160.±12.8 | 32.4±0.371 | 29.5±1.08 |

| badness | | α | β | k | τ | σ | quality |
|---|---|---|---|---|---|---|---|
| 2.9% | error | 1.8% | 8.9% | 0.14% | 1.1% | 3.7% | 11.4% |

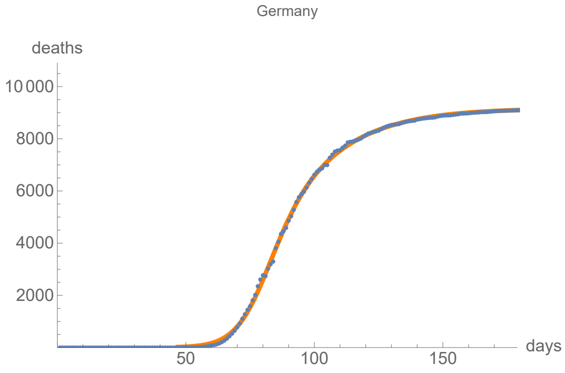


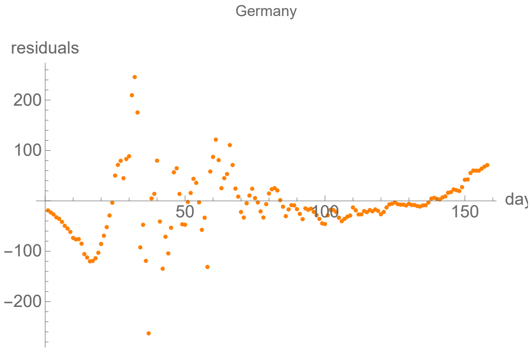


#### Parameters of fits to averaged death data.

The best fit ends on Sat 9 May 2020.

| α | β | k | τ | σ |
|---|---|---|---|---|
| 0.348±0.0111 | 0.108±0.00147 | 8000.±36.6 | 38.9±0.0722 | 19.9±0.249 |

| badness | | α | β | k | τ | σ | quality |
|---|---|---|---|---|---|---|---|
| 0.71% | error | 3.2% | 1.4% | 0.46% | 0.19% | 1.3% | 7.15% |

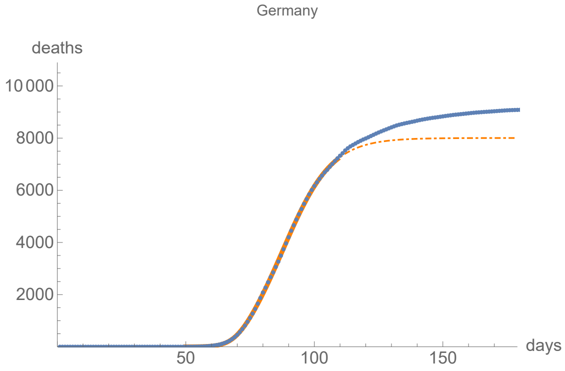


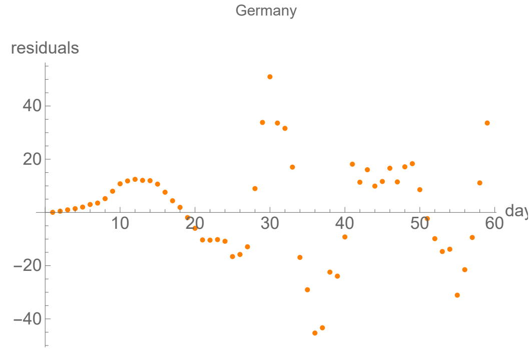


### Hungary

lag from integral overlap between 0 and 7 days, maximum overlap on day 7

lag from integral overlap, using data averaged over 7 days, between 5 and 10 days, maximum overlap on day 8

lag from fits to the function I: 4.00293 days

lag from fits to the function I (averaged data): 4.20852 days

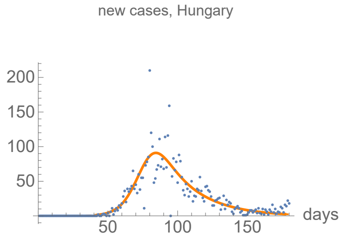


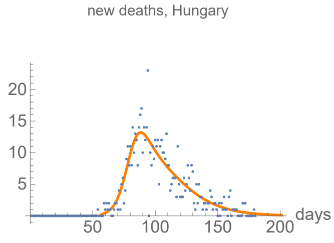


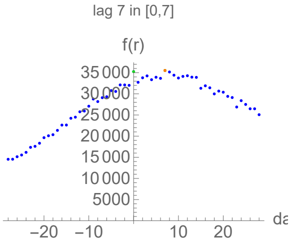


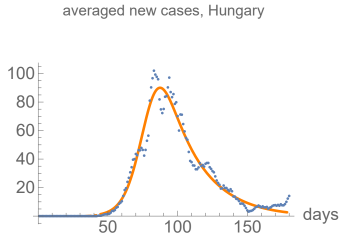


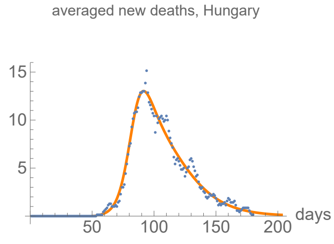


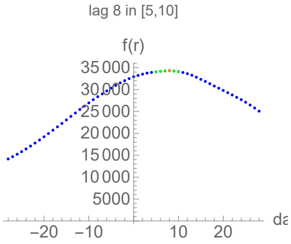

#### Parameters of fits to cases data.

The best fit ends on Thu 16 Jul 2020.

| α | β | k | τ | σ |
|---|---|---|---|---|
| 0.162±0.00386 | 0.0449±0.00452 | 4290.±13.8 | 36.2±0.685 | 39.5±1.29 |

| badness | | α | β | k | τ | σ | quality |
|---|---|---|---|---|---|---|---|
| 2.1% | error | 2.4% | 10.% | 0.32% | 1.9% | 3.3% | 12.1% |

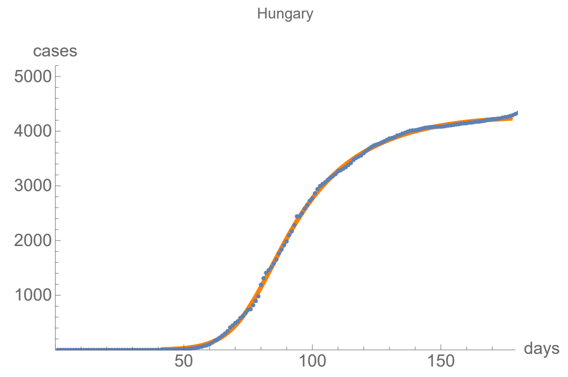


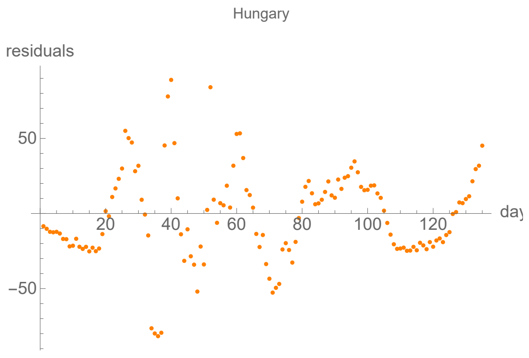


#### Parameters of fits to averaged cases data.

The best fit ends on Fri 17 Jul 2020.

| α | β | k | τ | σ |
|---|---|---|---|---|
| 0.161±0.00279 | 0.0457±0.00326 | 4280.±9.96 | 35.9±0.514 | 39.9±0.909 |

| badness | | α | β | k | τ | σ | quality |
|---|---|---|---|---|---|---|---|
| 1.1% | error | 1.7% | 7.1% | 0.23% | 1.4% | 2.3% | 8.34% |

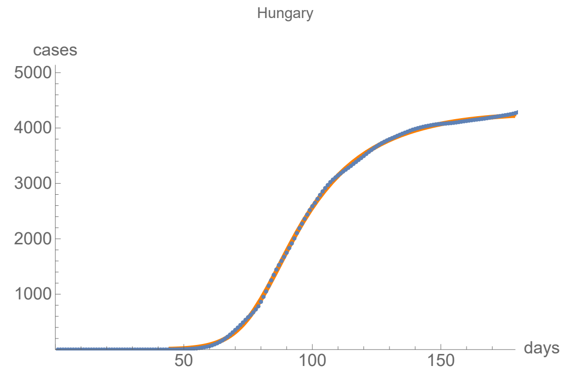


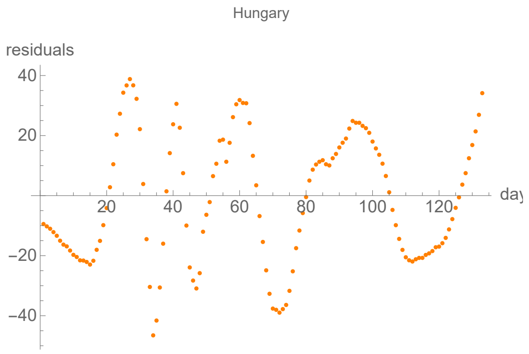


#### Parameters of fits to death data.

The best fit ends on Sun 9 Aug 2020.

| α | β | k | τ | σ |
|---|---|---|---|---|
| 0.209±0.00383 | 0.0531±0.000672 | 602.±0.647 | 37.6±0.283 | 24.2±0.249 |

| badness | | α | β | k | τ | σ | quality |
|---|---|---|---|---|---|---|---|
| 2.2% | error | 1.8% | 1.3% | 0.11% | 0.75% | 1.% | 7.14% |

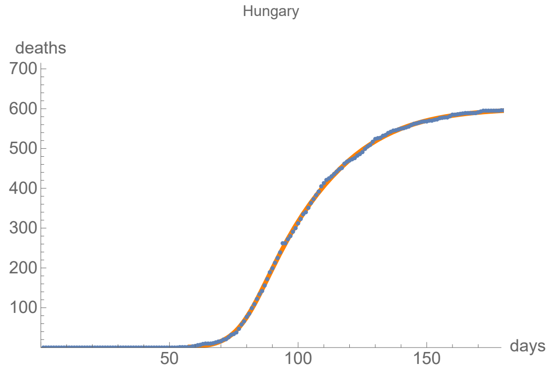


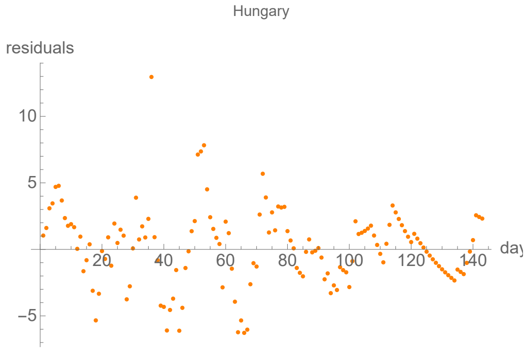


#### Parameters of fits to averaged death data.

The best fit ends on Tue 11 Aug 2020.

| α | β | k | τ | σ |
|---|---|---|---|---|
| 0.205±0.00262 | 0.0529±0.000482 | 602.±0.464 | 40.5±0.208 | 27.1±0.184 |

| badness | | α | β | k | τ | σ | quality |
|---|---|---|---|---|---|---|---|
| 0.77% | error | 1.3% | 0.91% | 0.077% | 0.51% | 0.68% | 4.23% |

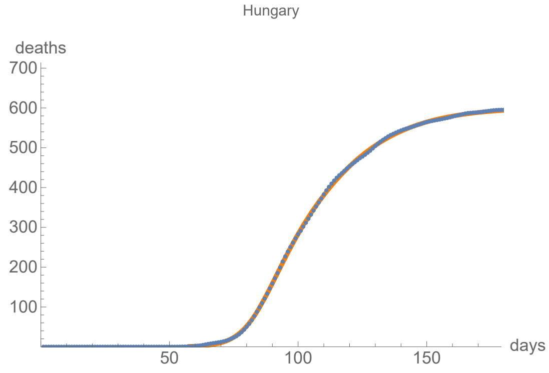


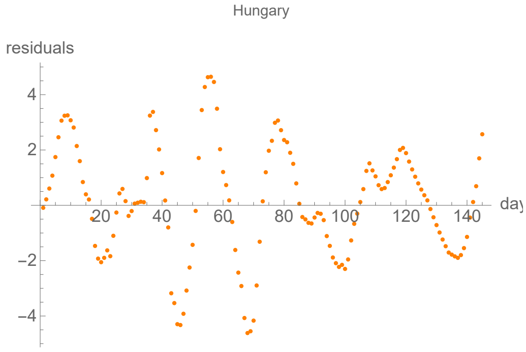


### Ireland

lag from integral overlap between 14 and 14 days, maximum overlap on day 14

lag from integral overlap, using data averaged over 7 days, between 7 and 10 days, maximum overlap on day 9

lag from fits to the function I: 5.58788 days

lag from fits to the function I (averaged data): 5.4144 days

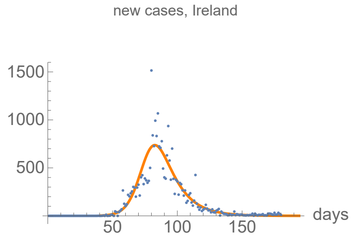


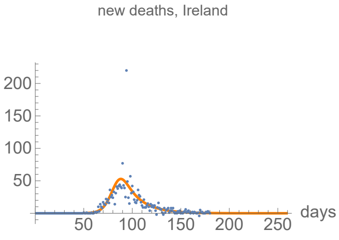


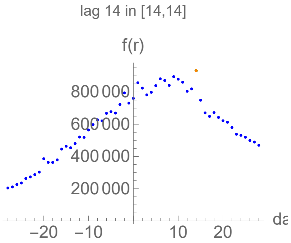


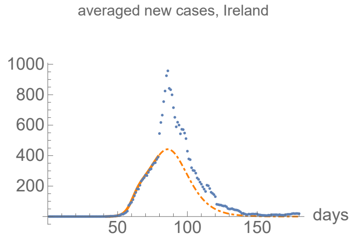


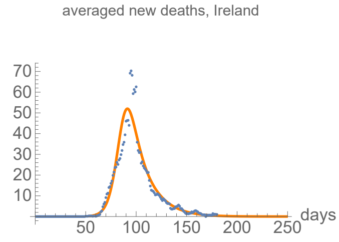


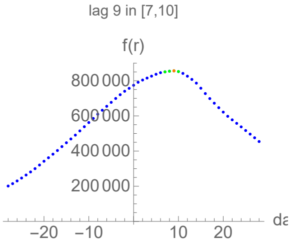

#### Parameters of fits to cases data.

The best fit ends on Sun 2 Aug 2020.

| α | β | k | τ | σ |
|---|---|---|---|---|
| 0.202±0.00738 | 0.0709±0.00822 | 25600.±41.7 | 35.9±1.44 | 35.9±2.51 |

| badness | | α | β | k | τ | σ | quality |
|---|---|---|---|---|---|---|---|
| 4.7% | error | 3.6% | 12.% | 0.16% | 4.% | 7.% | 22.7% |

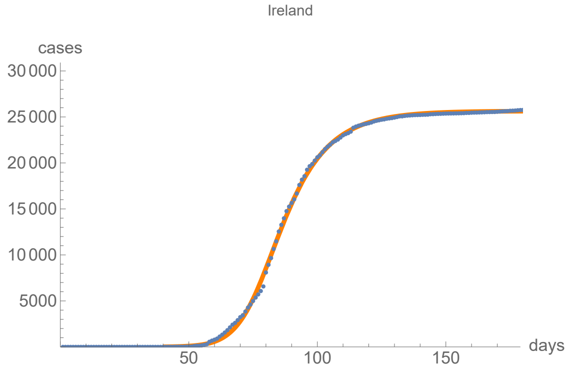


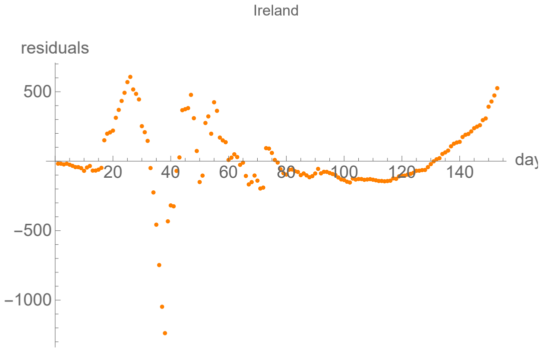


#### Parameters of fits to averaged cases data.

The best fit ends on Thu 9 Apr 2020.

| α | β | k | τ | σ |
|---|---|---|---|---|
| 0.368±0.00693 | 0.104±0.00761 | 16900.±1470. | 19.8±0.245 | 44.2±1.46 |

| badness | | α | β | k | τ | σ | quality |
|---|---|---|---|---|---|---|---|
| 0.34% | error | 1.9% | 7.3% | 8.7% | 1.2% | 3.3% | 16.9% |

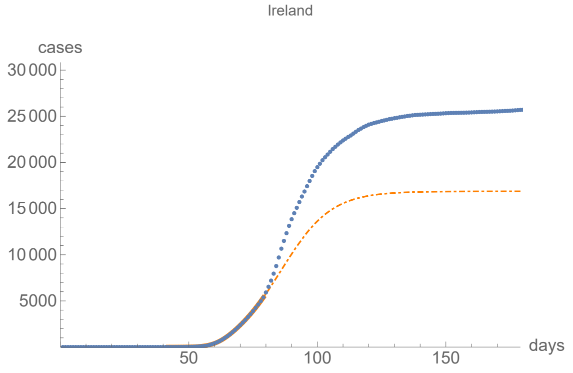


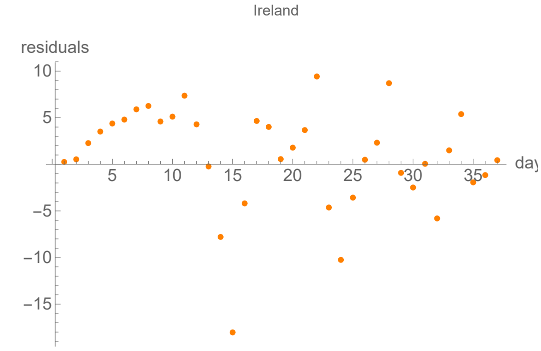


#### Parameters of fits to death data.

The best fit ends on Tue 6 Oct 2020.

| α | β | k | τ | σ |
|---|---|---|---|---|
| 0.22±0.00913 | 0.0596±0.00313 | 1770.±2.75 | 32.3±1.8 | 32.2±0.766 |

| badness | | α | β | k | τ | σ | quality |
|---|---|---|---|---|---|---|---|
| 5.4% | error | 4.2% | 5.2% | 0.16% | 5.6% | 2.4% | 22.9% |

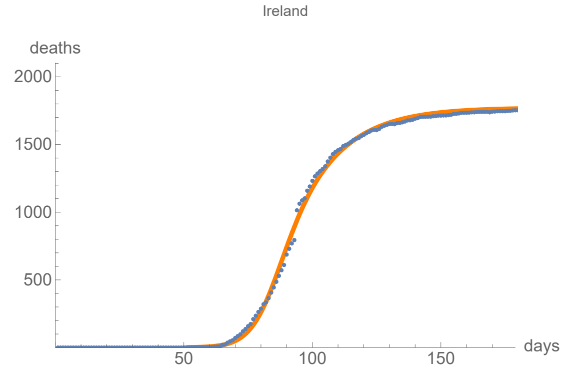


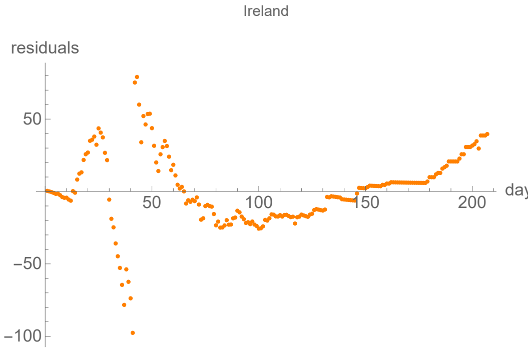


#### Parameters of fits to averaged death data.

The best fit ends on Sat 26 Sep 2020.

| α | β | k | τ | σ |
|---|---|---|---|---|
| 0.216±0.00741 | 0.0603±0.00913 | 1770.±2.4 | 33.1±0.689 | 33.1±1.57 |

| badness | | α | β | k | τ | σ | quality |
|---|---|---|---|---|---|---|---|
| 3.9% | error | 3.4% | 15.% | 0.14% | 2.1% | 4.7% | 17.8% |

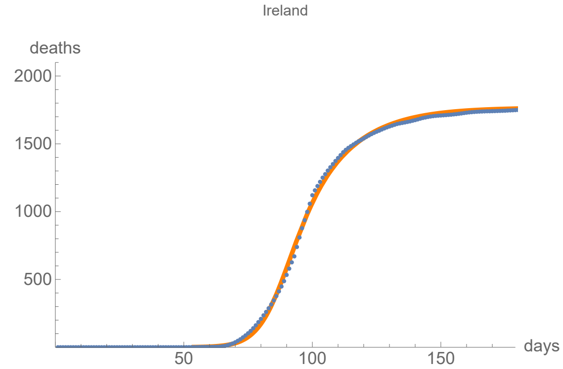


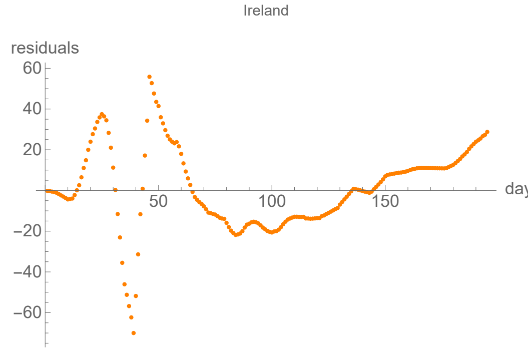


### Italy

lag from integral overlap between 1 and 7 days, maximum overlap on day 6

lag from integral overlap, using data averaged over 7 days, between 1 and 6 days, maximum overlap on day 4

lag from fits to the function I: 4.43773 days

lag from fits to the function I (averaged data): 3.81535 days

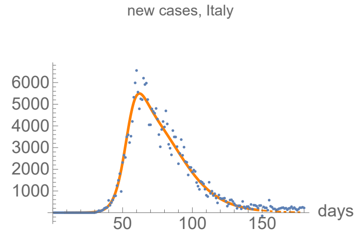


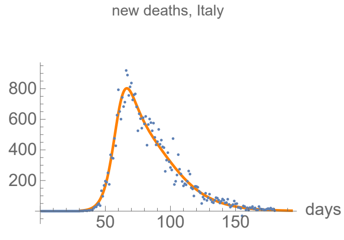


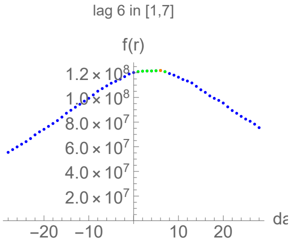


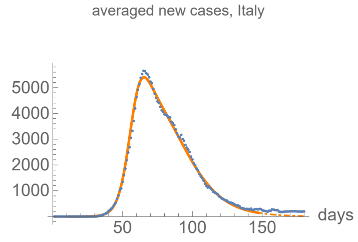


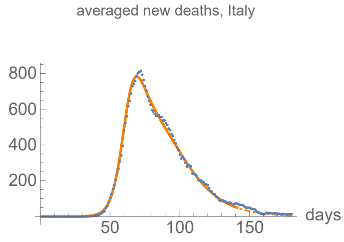


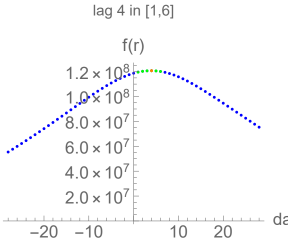

#### Parameters of fits to cases data.

The best fit ends on Tue 16 Jun 2020.

| α | β | k | τ | σ |
|---|---|---|---|---|
| 0.252±0.00293 | 0.0636±0.000406 | 238000.±178. | 62.6±0.0932 | 46.8±0.118 |

| badness | | α | β | k | τ | σ | quality |
|---|---|---|---|---|---|---|---|
| 0.77% | error | 1.2% | 0.64% | 0.075% | 0.15% | 0.25% | 3.05% |

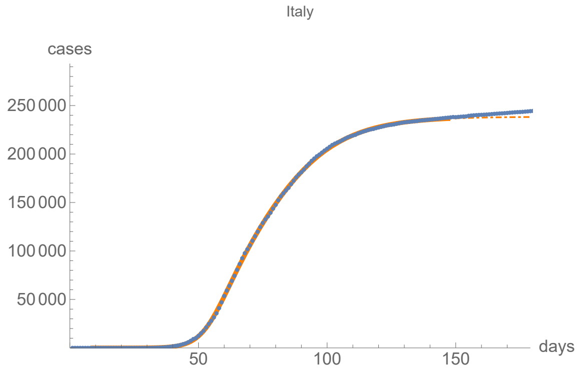


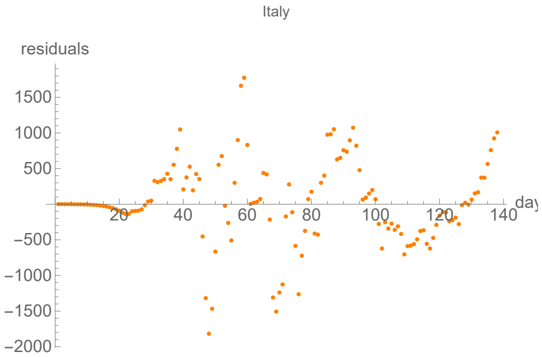


#### Parameters of fits to averaged cases data.

The best fit ends on Tue 16 Jun 2020.

| α | β | k | τ | σ |
|---|---|---|---|---|
| 0.246±0.00209 | 0.0636±0.000308 | 238000.±138. | 62.6±0.0699 | 46.6±0.0916 |

| badness | | α | β | k | τ | σ | quality |
|---|---|---|---|---|---|---|---|
| 0.35% | error | 0.85% | 0.48% | 0.058% | 0.11% | 0.2% | 2.05% |

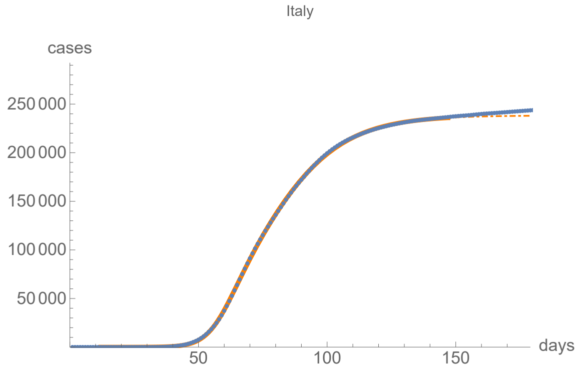


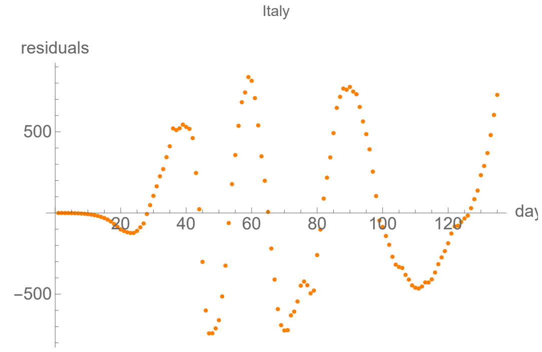


#### Parameters of fits to death data.

The best fit ends on Sat 1 Aug 2020.

| α | β | k | τ | σ |
|---|---|---|---|---|
| 0.233±0.00232 | 0.0555±0.000302 | 35100.±15.7 | 44.5±0.113 | 30.1±0.105 |

| badness | | α | β | k | τ | σ | quality |
|---|---|---|---|---|---|---|---|
| 0.76% | error | 1.% | 0.54% | 0.045% | 0.25% | 0.35% | 2.94% |

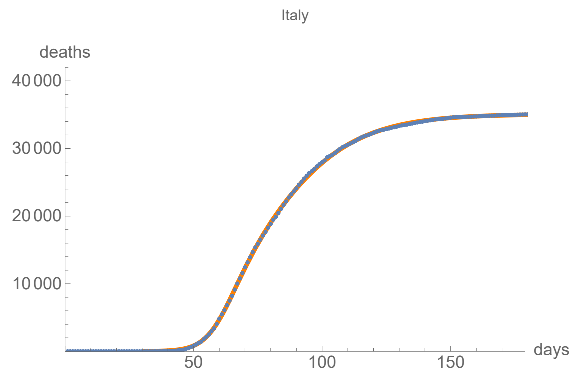


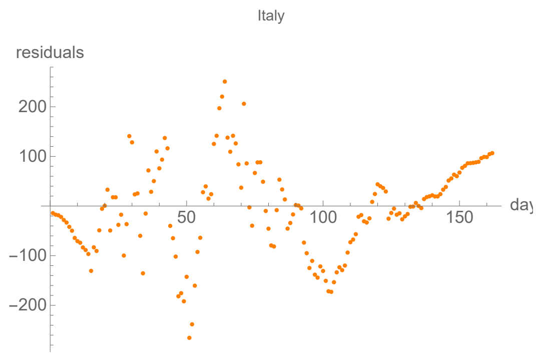


#### Parameters of fits to averaged death data.

The best fit ends on Wed 10 Jun 2020.

| α | β | k | τ | σ |
|---|---|---|---|---|
| 0.241±0.00176 | 0.0591±0.00031 | 34700.±28.4 | 45.9±0.0632 | 30.2±0.0832 |

| badness | | α | β | k | τ | σ | quality |
|---|---|---|---|---|---|---|---|
| 0.36% | error | 0.73% | 0.52% | 0.082% | 0.14% | 0.28% | 2.11% |

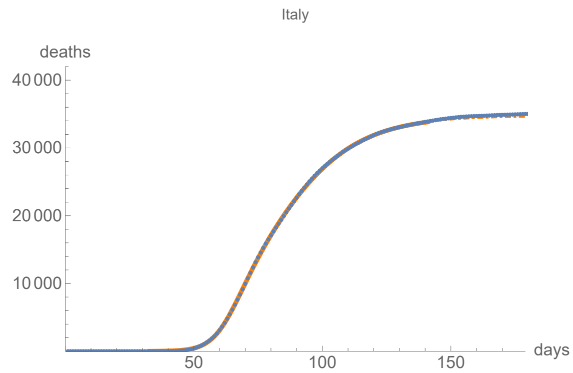


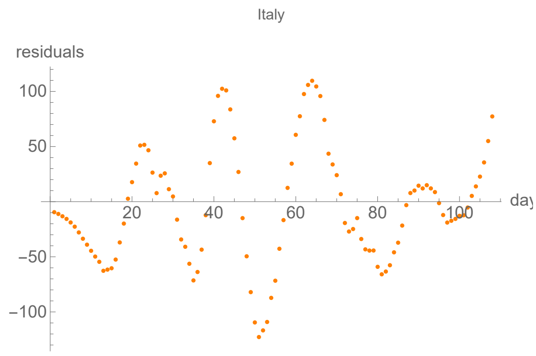


### Netherlands

lag from integral overlap between 5 and 6 days, maximum overlap on day 5

lag from integral overlap, using data averaged over 7 days, between 2 and 7 days, maximum overlap on day 4

lag from fits to the function I: -2.95931 days

lag from fits to the function I (averaged data): -3.4936 days

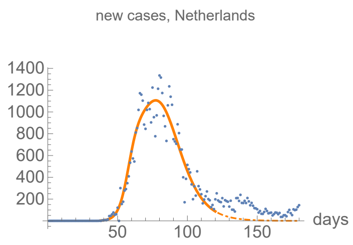


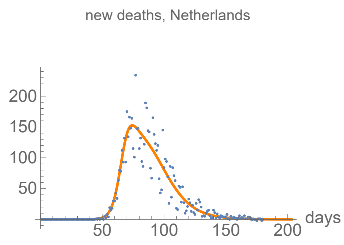


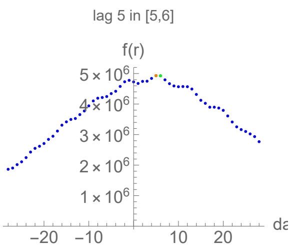


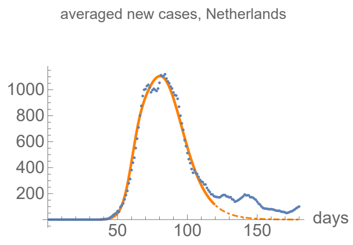


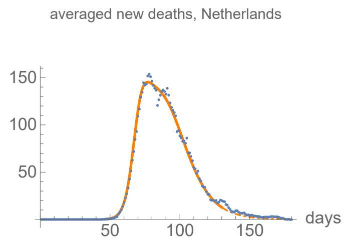


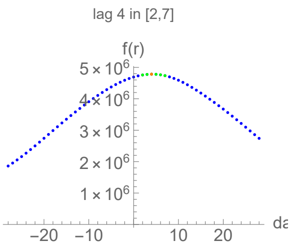

#### Parameters of fits to cases data.

The best fit ends on Wed 20 May 2020.

| α | β | k | τ | σ |
|---|---|---|---|---|
| 0.314±0.0148 | 0.0938±0.00133 | 45100.±115. | 41.1±0.104 | 20.7±0.35 |

| badness | | α | β | k | τ | σ | quality |
|---|---|---|---|---|---|---|---|
| 1.2% | error | 4.7% | 1.4% | 0.26% | 0.25% | 1.7% | 9.52% |

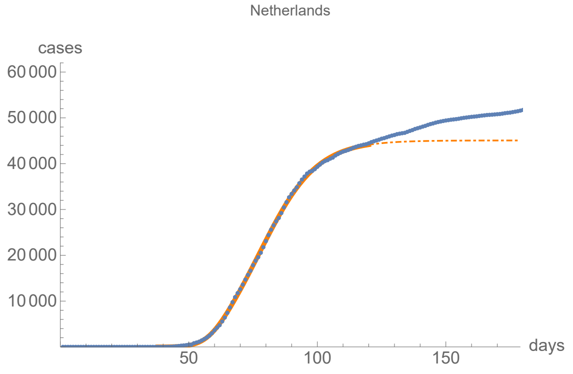


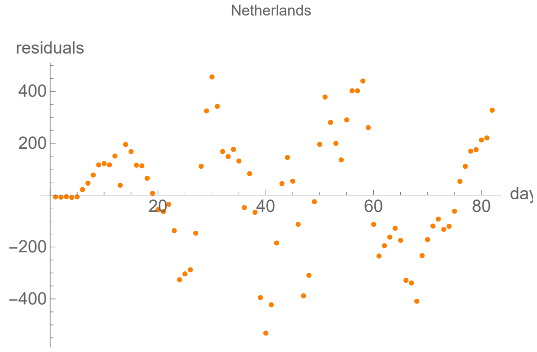


#### Parameters of fits to averaged cases data.

The best fit ends on Tue 19 May 2020.

| α | β | k | τ | σ |
|---|---|---|---|---|
| 0.309±0.0103 | 0.0944±0.000979 | 44900.±93.3 | 44.±0.0708 | 23.2±0.258 |

| badness | | α | β | k | τ | σ | quality |
|---|---|---|---|---|---|---|---|
| 0.63% | error | 3.3% | 1.% | 0.21% | 0.16% | 1.1% | 6.48% |

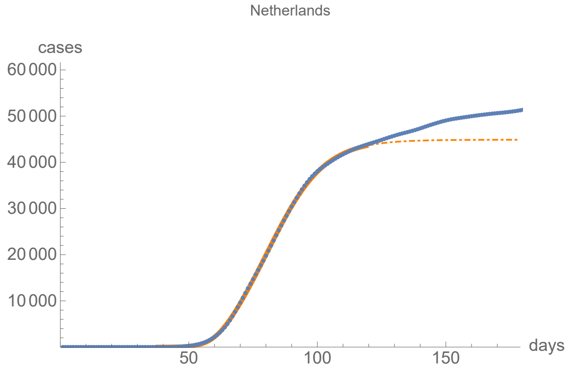


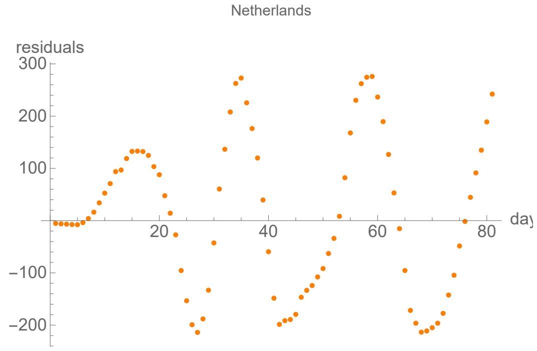


#### Parameters of fits to death data.

The best fit ends on Tue 11 Aug 2020.

| α | β | k | τ | σ |
|---|---|---|---|---|
| 0.291±0.00649 | 0.0762±0.000622 | 6130.±3.44 | 37.3±0.129 | 21.6±0.179 |

| badness | | α | β | k | τ | σ | quality |
|---|---|---|---|---|---|---|---|
| 1.8% | error | 2.2% | 0.82% | 0.056% | 0.35% | 0.83% | 6.12% |

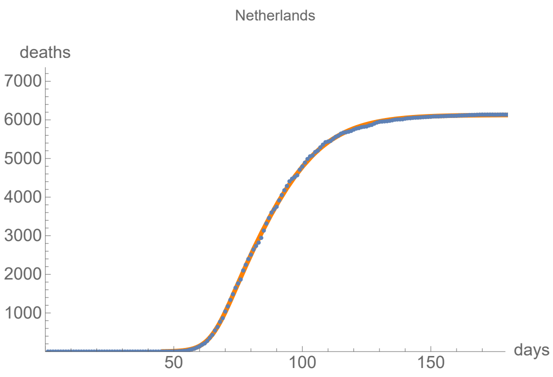


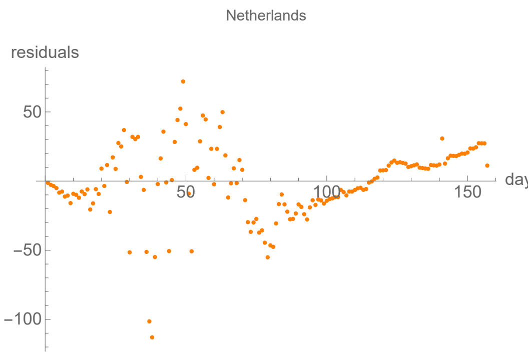


#### Parameters of fits to averaged death data.

The best fit ends on Sat 30 May 2020.

| α | β | k | τ | σ |
|---|---|---|---|---|
| 0.317±0.00344 | 0.0829±0.000395 | 6030.±5.35 | 39.5±0.0359 | 22.2±0.0817 |

| badness | | α | β | k | τ | σ | quality |
|---|---|---|---|---|---|---|---|
| 0.4% | error | 1.1% | 0.48% | 0.089% | 0.091% | 0.37% | 2.51% |

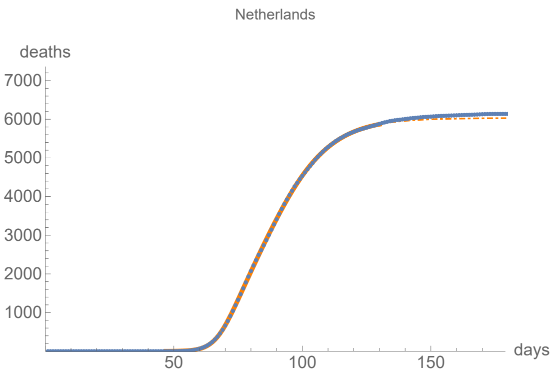


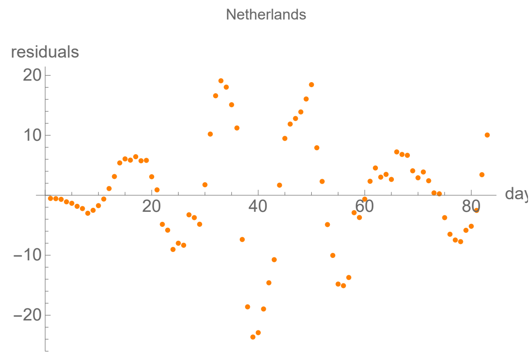


### Poland

lag from integral overlap between 5 and 11 days, maximum overlap on day 10

lag from integral overlap, using data averaged over 7 days, between 5 and 14 days, maximum overlap on day 9

lag from fits to the function I: 11.0418 days

lag from fits to the function I (averaged data): 10.6938 days

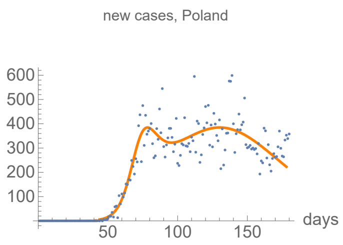


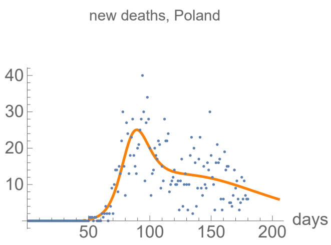


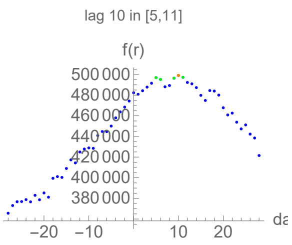


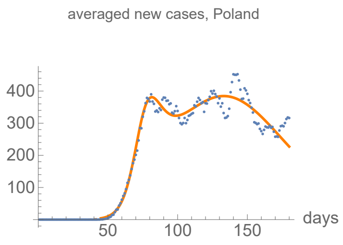


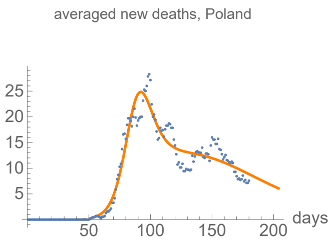


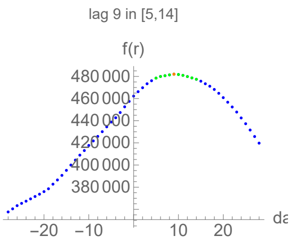

#### Parameters of fits to cases data.

The best fit ends on Fri 17 Jul 2020.

| α | β | k | τ | σ |
|---|---|---|---|---|
| 0.191±0.00406 | 0.0324±0.000236 | 47400.±225. | 86.4±0.329 | 28.6±0.188 |

| badness | | α | β | k | τ | σ | quality |
|---|---|---|---|---|---|---|---|
| 1.2% | error | 2.1% | 0.73% | 0.47% | 0.38% | 0.66% | 5.54% |

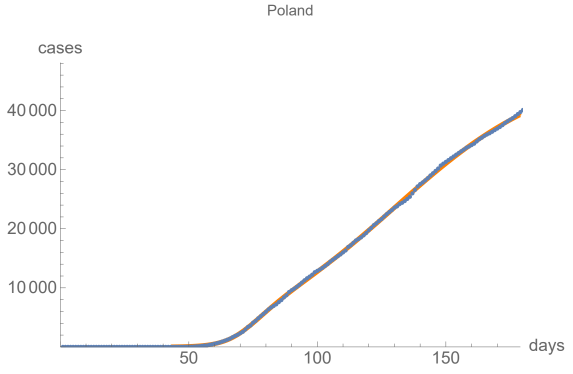


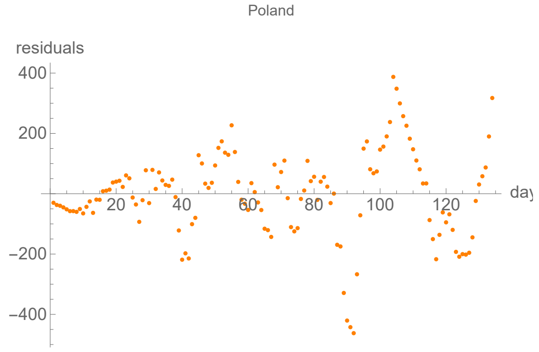


#### Parameters of fits to averaged cases data.

The best fit ends on Sun 19 Jul 2020.

| α | β | k | τ | σ |
|---|---|---|---|---|
| 0.188±0.0032 | 0.0324±0.000195 | 47400.±188. | 88.3±0.274 | 30.5±0.154 |

| badness | | α | β | k | τ | σ | quality |
|---|---|---|---|---|---|---|---|
| 0.84% | error | 1.7% | 0.6% | 0.4% | 0.31% | 0.5% | 4.35% |

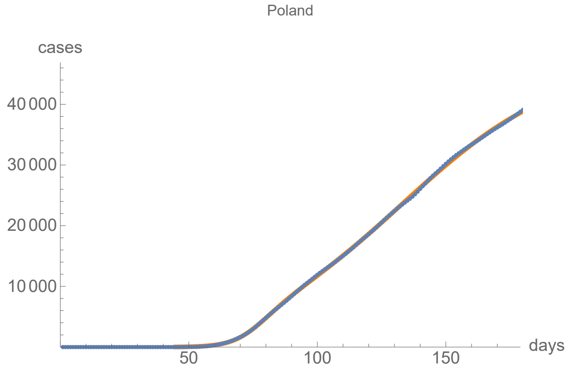


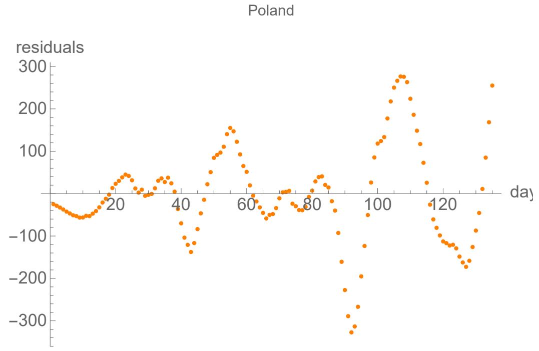


#### Parameters of fits to death data.

The best fit ends on Thu 13 Aug 2020.

| α | β | k | τ | σ |
|---|---|---|---|---|
| 0.157±0.00414 | 0.0238±0.000519 | 2110.±20.5 | 77.±0.787 | 34.1±0.312 |

| badness | | α | β | k | τ | σ | quality |
|---|---|---|---|---|---|---|---|
| 1.7% | error | 2.6% | 2.2% | 0.97% | 1.% | 0.91% | 9.41% |

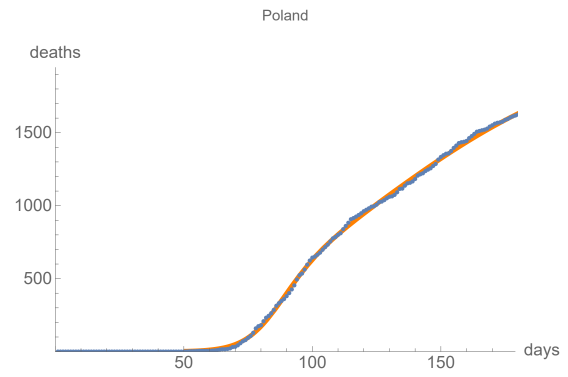


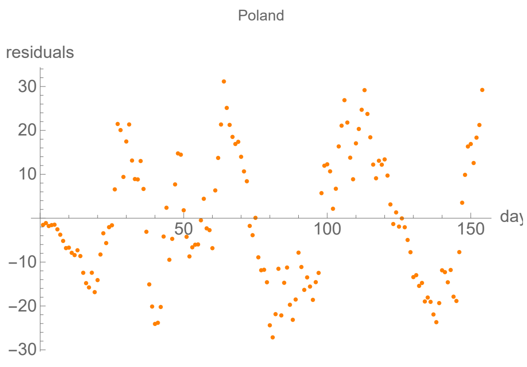


#### Parameters of fits to averaged death data.

The best fit ends on Wed 12 Aug 2020.

| α | β | k | τ | σ |
|---|---|---|---|---|
| 0.158±0.00365 | 0.0245±0.000477 | 2070.±17.9 | 76.6±0.674 | 34.8±0.276 |

| badness | | α | β | k | τ | σ | quality |
|---|---|---|---|---|---|---|---|
| 1.3% | error | 2.3% | 1.9% | 0.87% | 0.88% | 0.79% | 8.06% |

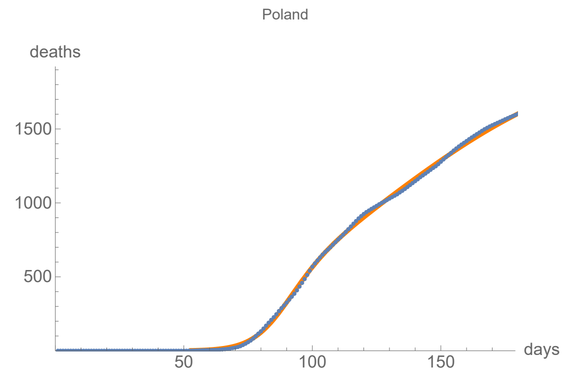


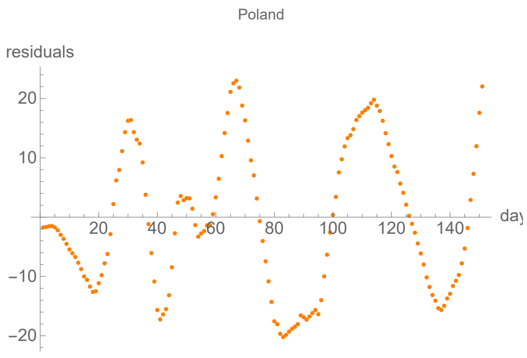


### Portugal

lag from integral overlap between 3 and 7 days, maximum overlap on day 5

lag from integral overlap, using data averaged over 7 days, between 3 and 9 days, maximum overlap on day 5

lag from fits to the function I: 10.2502 days

lag from fits to the function I (averaged data): 10.7209 days

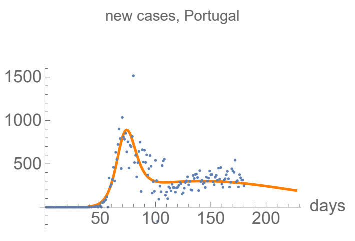


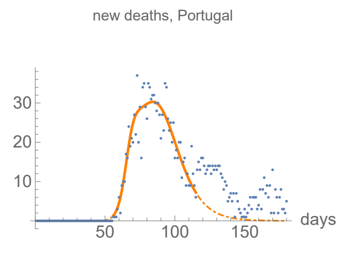


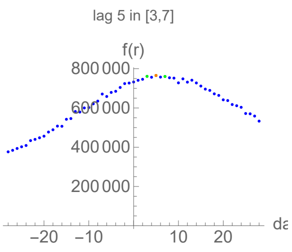


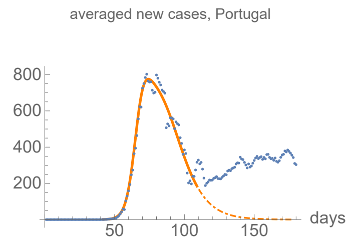


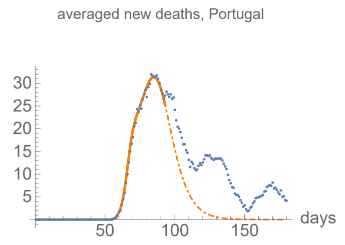


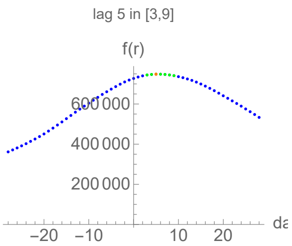

#### Parameters of fits to cases data.

The best fit ends on Fri 4 Sep 2020.

| α | β | k | τ | σ |
|---|---|---|---|---|
| 0.191±0.0055 | 0.0164±0.000275 | 73300.±813. | 103.±1.47 | 32.2±0.217 |

| badness | | α | β | k | τ | σ | quality |
|---|---|---|---|---|---|---|---|
| 1.8% | error | 2.9% | 1.7% | 1.1% | 1.4% | 0.67% | 9.61% |

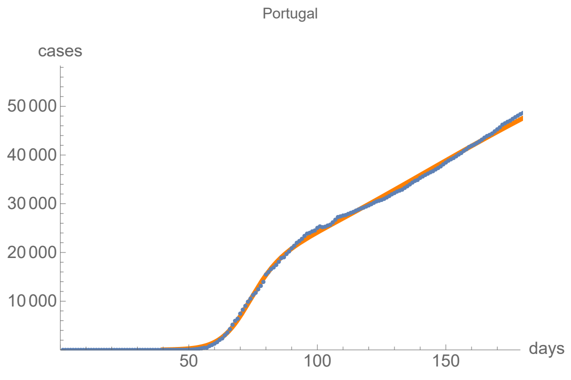


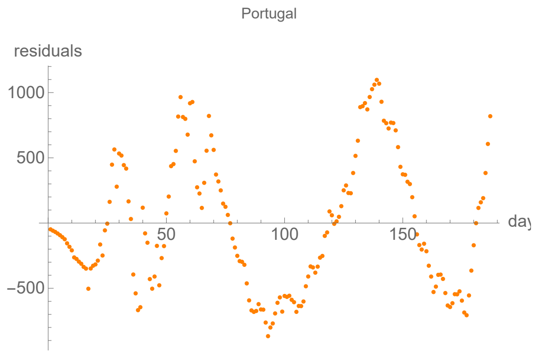


#### Parameters of fits to averaged cases data.

The best fit ends on Sat 9 May 2020.

| α | β | k | τ | σ |
|---|---|---|---|---|
| 0.323±0.00697 | 0.0893±0.00145 | 28500.±121. | 39.5±0.0824 | 24.7±0.211 |

| badness | | α | β | k | τ | σ | quality |
|---|---|---|---|---|---|---|---|
| 0.69% | error | 2.2% | 1.6% | 0.42% | 0.21% | 0.85% | 5.96% |

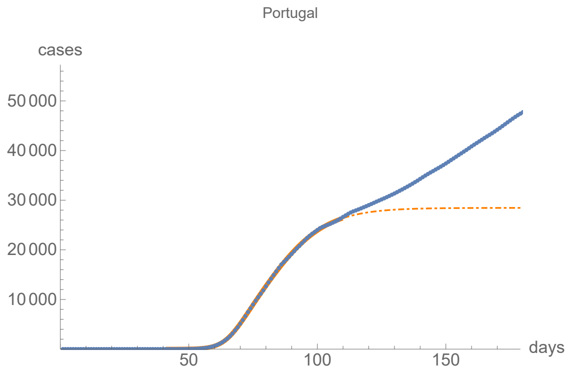


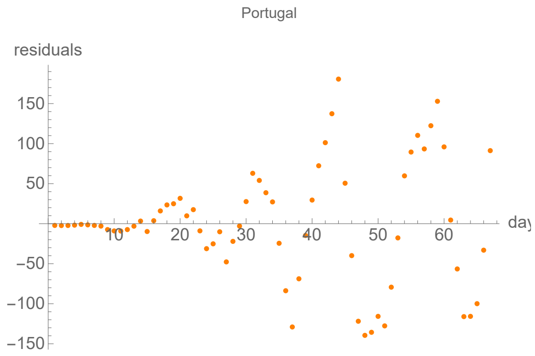


#### Parameters of fits to death data.

The best fit ends on Fri 15 May 2020.

| α | β | k | τ | σ |
|---|---|---|---|---|
| 0.351±0.0133 | 0.0921±0.00135 | 1270.±5.48 | 30.3±0.0916 | 10.9±0.24 |

| badness | | α | β | k | τ | σ | quality |
|---|---|---|---|---|---|---|---|
| 1.1% | error | 3.8% | 1.5% | 0.43% | 0.3% | 2.2% | 9.29% |

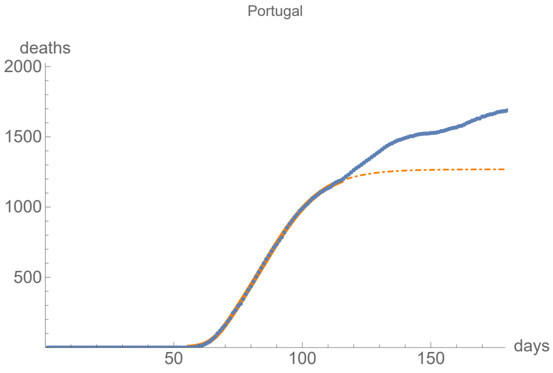


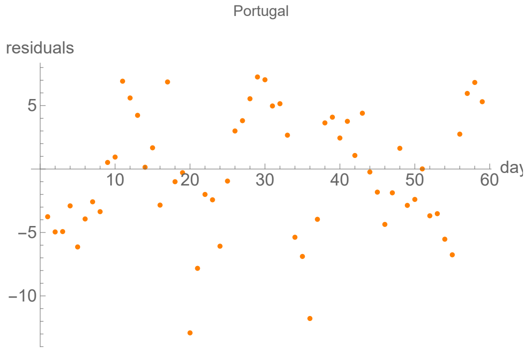


#### Parameters of fits to averaged death data.

The best fit ends on Thu 23 Apr 2020.

| α | β | k | τ | σ |
|---|---|---|---|---|
| 0.451±0.00599 | 0.121±0.000926 | 1030.±6.08 | 27.3±0.101 | 9.15±0.0747 |

| badness | | α | β | k | τ | σ | quality |
|---|---|---|---|---|---|---|---|
| 0.22% | error | 1.3% | 0.77% | 0.59% | 0.37% | 0.82% | 4.09% |

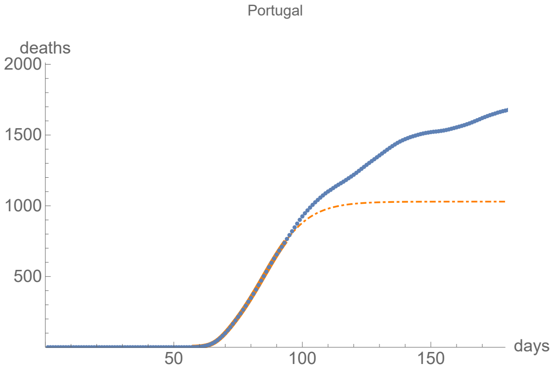


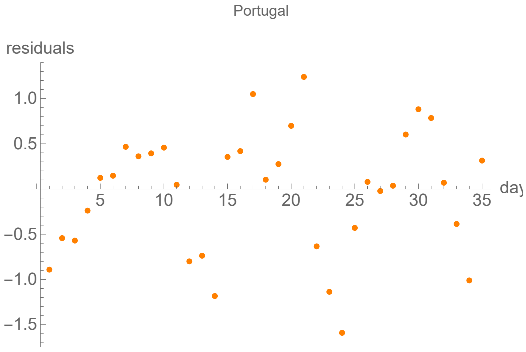


### Romania

lag from integral overlap between 16 and 16 days, maximum overlap on day 16

lag from integral overlap, using data averaged over 7 days, between 4 and 19 days, maximum overlap on day 13

lag from fits to the function I: -3.24463 days

lag from fits to the function I (averaged data): 19.8699 days

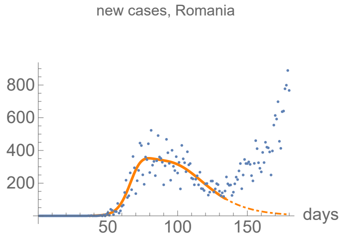


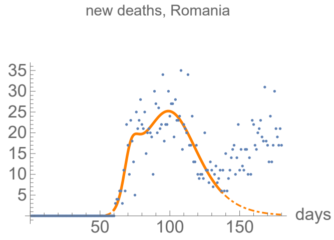


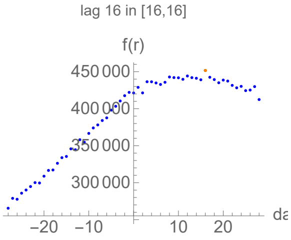


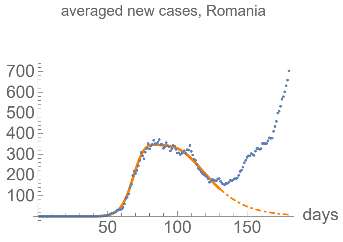


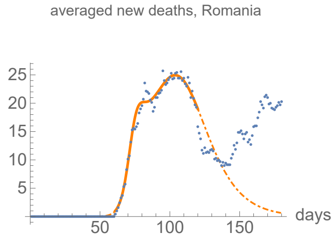


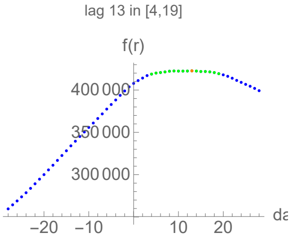

#### Parameters of fits to cases data.

The best fit ends on Wed 3 Jun 2020.

| α | β | k | τ | σ |
|---|---|---|---|---|
| 0.231±0.00568 | 0.0581±0.000842 | 21600.±103. | 57.9±0.141 | 32.±0.266 |

| badness | | α | β | k | τ | σ | quality |
|---|---|---|---|---|---|---|---|
| 0.98% | error | 2.5% | 1.5% | 0.48% | 0.24% | 0.83% | 6.44% |

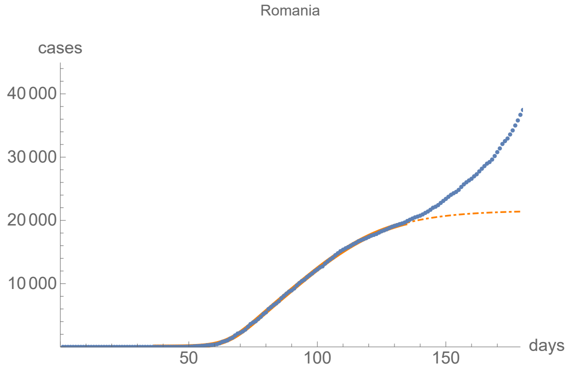


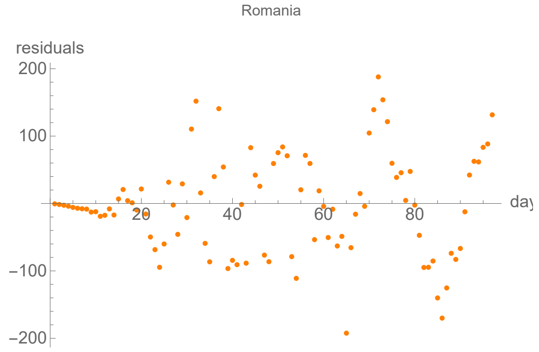


#### Parameters of fits to averaged cases data.

The best fit ends on Sat 30 May 2020.

| α | β | k | τ | σ |
|---|---|---|---|---|
| 0.237±0.00338 | 0.0608±0.000543 | 21100.±71.5 | 59.2±0.0966 | 33.2±0.157 |

| badness | | α | β | k | τ | σ | quality |
|---|---|---|---|---|---|---|---|
| 0.49% | error | 1.4% | 0.89% | 0.34% | 0.16% | 0.47% | 3.78% |

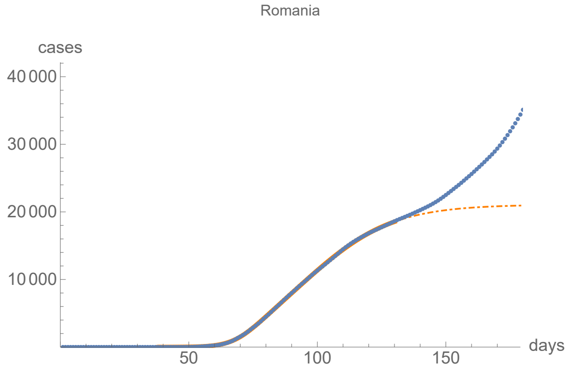


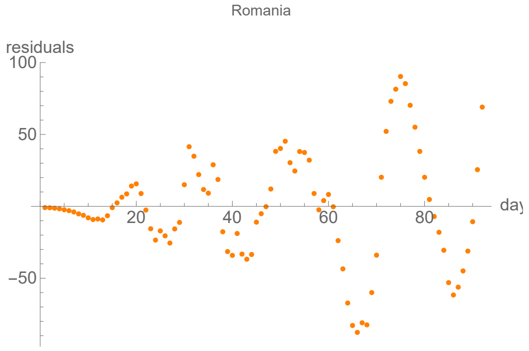


#### Parameters of fits to death data.

The best fit ends on Sat 6 Jun 2020.

| α | β | k | τ | σ |
|---|---|---|---|---|
| 0.348±0.0166 | 0.072±0.000663 | 1400.±4.77 | 39.1±0.116 | 10.2±0.24 |

| badness | | α | β | k | τ | σ | quality |
|---|---|---|---|---|---|---|---|
| 1.1% | error | 4.8% | 0.92% | 0.34% | 0.3% | 2.4% | 9.85% |

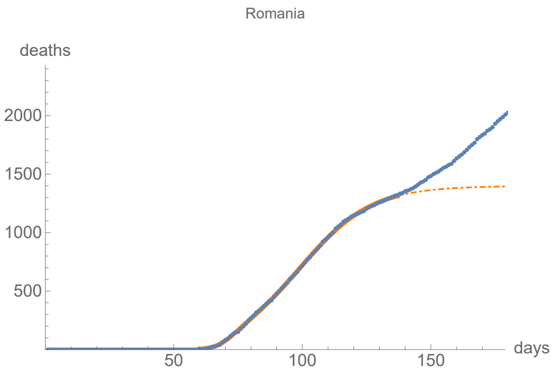


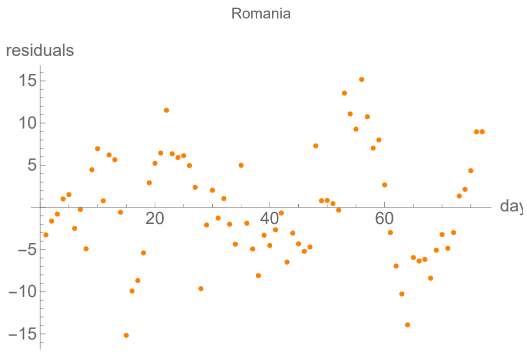


#### Parameters of fits to averaged death data.

The best fit ends on Wed 20 May 2020.

| α | β | k | τ | σ |
|---|---|---|---|---|
| 0.31±0.00551 | 0.0666±0.000587 | 1490.±10.4 | 43.1±0.224 | 12.6±0.114 |

| badness | | α | β | k | τ | σ | quality |
|---|---|---|---|---|---|---|---|
| 0.42% | error | 1.8% | 0.88% | 0.7% | 0.52% | 0.9% | 5.2% |

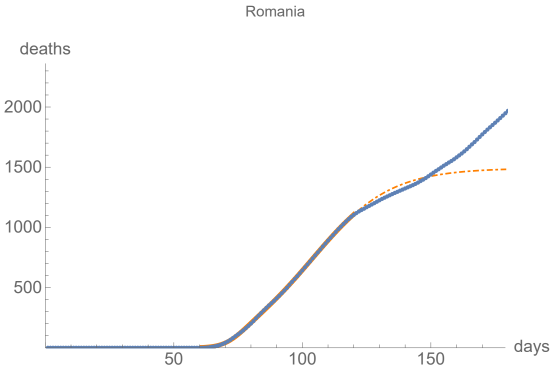


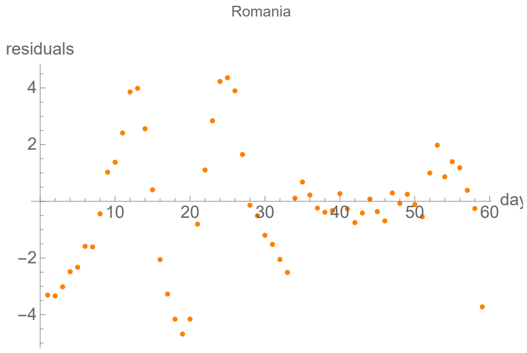


### Spain

lag from integral overlap between 0 and 0 days, maximum overlap on day 0

lag from integral overlap, using data averaged over 7 days, between 1 and 5 days, maximum overlap on day 3

lag from fits to the function I: −0.415911 days

lag from fits to the function I (averaged data): −0.309878 days

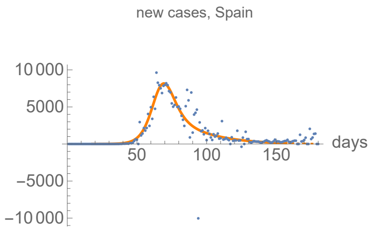


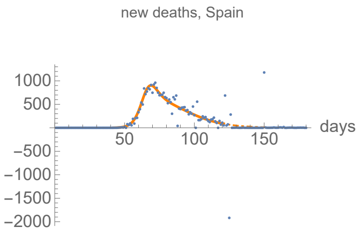


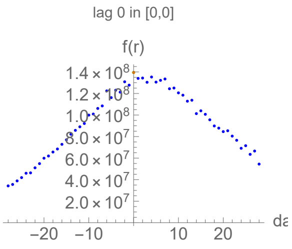


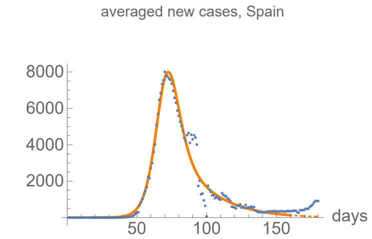


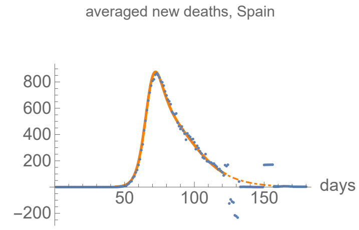


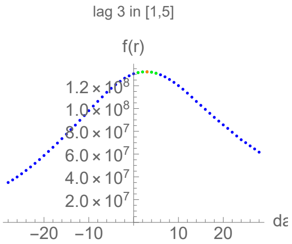

#### Parameters of fits to cases data.

The best fit ends on Fri 26 Jun 2020.

| α | β | k | τ | σ |
|---|---|---|---|---|
| 0.228±0.00532 | 0.0471±0.00646 | 248000.±855. | 49.2±0.312 | 42.6±1.36 |

| badness | | α | β | k | τ | σ | quality |
|---|---|---|---|---|---|---|---|
| 4.5% | error | 2.3% | 14.% | 0.34% | 0.63% | 3.2% | 13.1% |

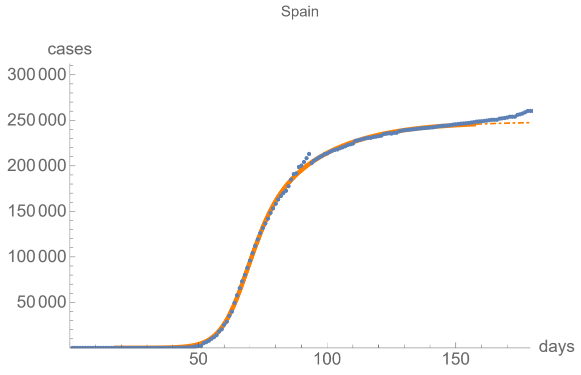


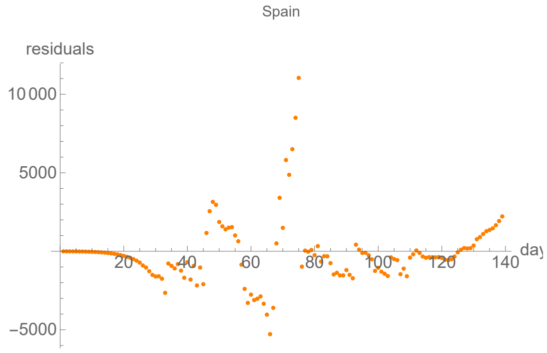


#### Parameters of fits to averaged cases data.

The best fit ends on Mon 29 Jun 2020.

| α | β | k | τ | σ |
|---|---|---|---|---|
| 0.221±0.00392 | 0.0459±0.00471 | 249000.±689. | 52.3±0.25 | 44.9±1.13 |

| badness | | α | β | k | τ | σ | quality |
|---|---|---|---|---|---|---|---|
| 2.4% | error | 1.8% | 10.% | 0.28% | 0.48% | 2.5% | 8.99% |

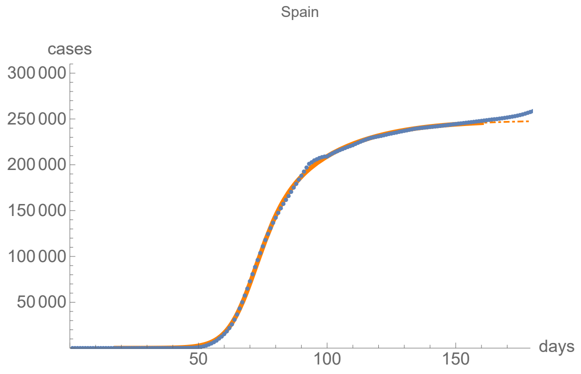


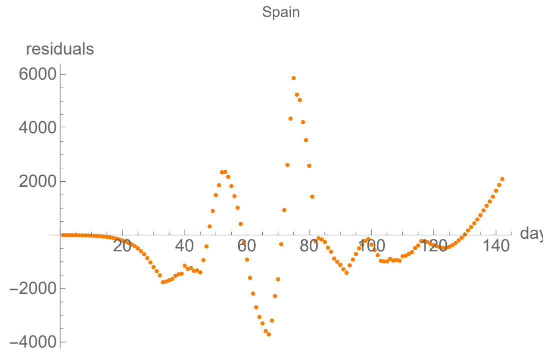


#### Parameters of fits to death data.

The best fit ends on Thu 21 May 2020.

| α | β | k | τ | σ |
|---|---|---|---|---|
| 0.315±0.00418 | 0.0683±0.000853 | 29300.±71.8 | 34.4±0.0959 | 23.6±0.111 |

| badness | | α | β | k | τ | σ | quality |
|---|---|---|---|---|---|---|---|
| 1.2% | error | 1.3% | 1.2% | 0.24% | 0.28% | 0.47% | 4.8% |

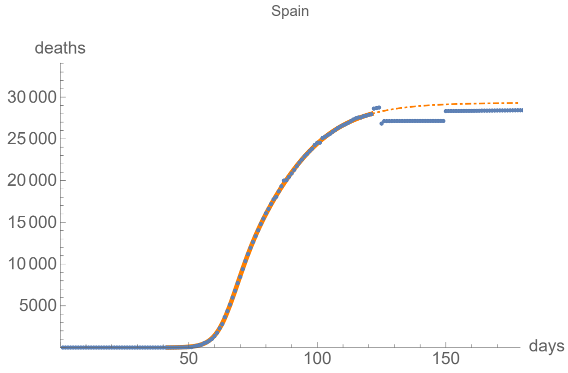


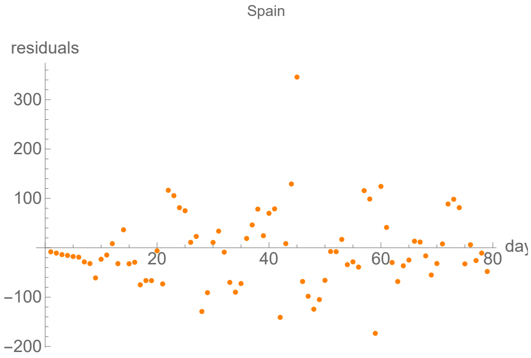


#### Parameters of fits to averaged death data.

The best fit ends on Fri 22 May 2020.

| α | β | k | τ | σ |
|---|---|---|---|---|
| 0.3±0.00168 | 0.0671±0.000397 | 29400.±35.7 | 35.3±0.0437 | 24.6±0.0529 |

| badness | | α | β | k | τ | σ | quality |
|---|---|---|---|---|---|---|---|
| 0.26% | error | 0.56% | 0.59% | 0.12% | 0.12% | 0.22% | 1.88% |

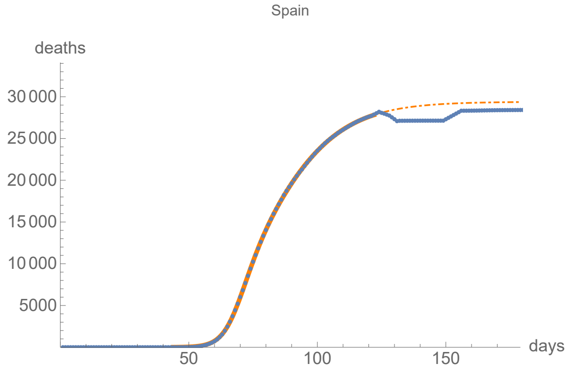


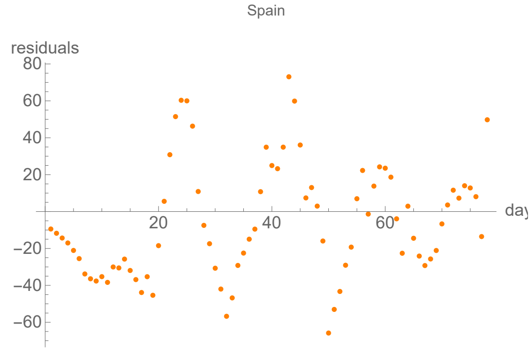


### Sweden

```
lag from integral overlap between -9 and -2 days, maximum overlap on day -6

lag from integral overlap, using data averaged over 7 days, between -11 and 0 days, maximum overlap on day -5

lag from fits to the function I: -16.8232 days

lag from fits to the function I (averaged data): -17.9024 days
```

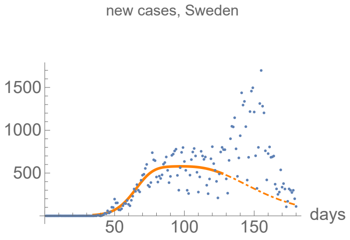


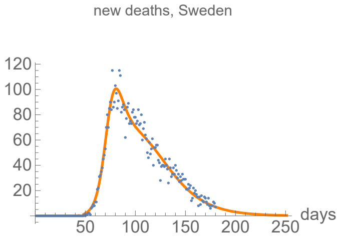


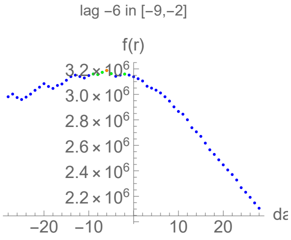


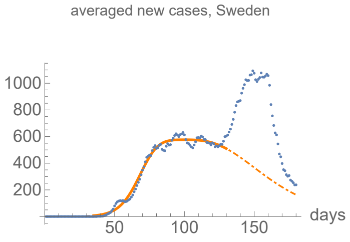


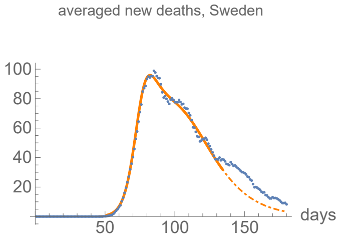


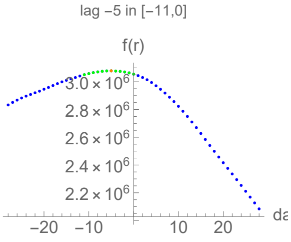

#### Parameters of fits to cases data.

The best fit ends on Wed 27 May 2020.

| α | β | k | τ | σ |
|---|---|---|---|---|
| 0.147±0.00528 | 0.0389±0.00623 | 55500.±2450. | 34.2±0.988 | 75.8±2.22 |

| badness | | α | β | k | τ | σ | quality |
|---|---|---|---|---|---|---|---|
| 1.% | error | 3.6% | 16.% | 4.4% | 2.9% | 2.9% | 18.1% |

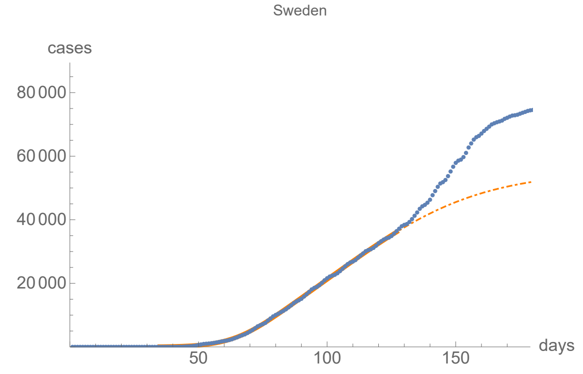


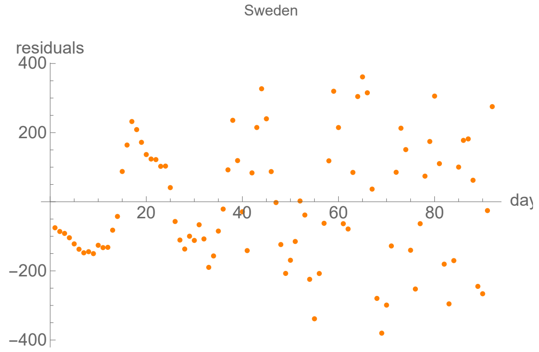


#### Parameters of fits to averaged cases data.

The best fit ends on Sat 30 May 2020.

| α | β | k | τ | σ |
|---|---|---|---|---|
| 0.144±0.00273 | 0.0375±0.00318 | 57600.±1560. | 37.6±0.539 | 80.7±1.42 |

| badness | | α | β | k | τ | σ | quality |
|---|---|---|---|---|---|---|---|
| 0.46% | error | 1.9% | 8.5% | 2.7% | 1.4% | 1.8% | 9.91% |

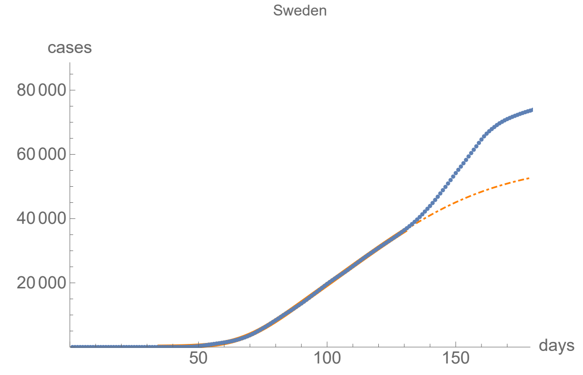


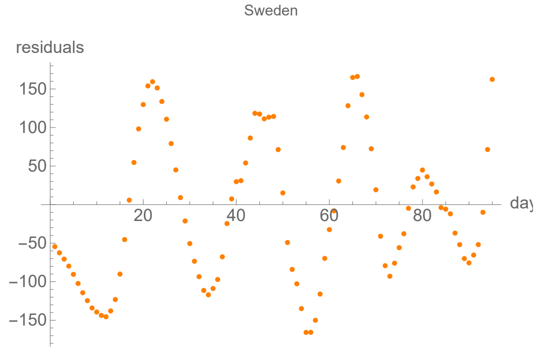


#### Parameters of fits to death data.

The best fit ends on Mon 28 Sep 2020.

| α | β | k | τ | σ |
|---|---|---|---|---|
| 0.206±0.0027 | 0.0432±0.000212 | 5870.±2.92 | 48.7±0.107 | 25.2±0.126 |

| badness | | α | β | k | τ | σ | quality |
|---|---|---|---|---|---|---|---|
| 0.92% | error | 1.3% | 0.49% | 0.05% | 0.22% | 0.5% | 3.49% |

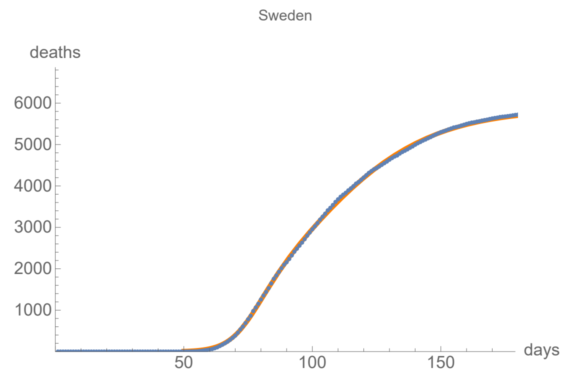


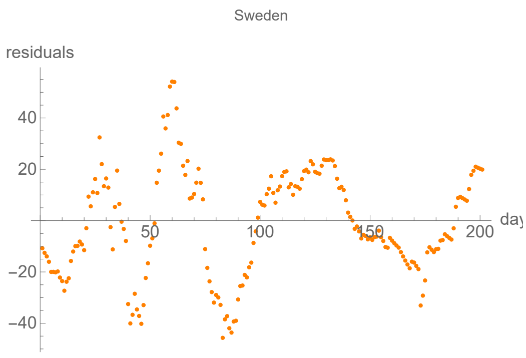


#### Parameters of fits to averaged death data.

The best fit ends on Wed 3 Jun 2020.

| α | β | k | τ | σ |
|---|---|---|---|---|
| 0.238±0.00234 | 0.0548±0.000455 | 5320.±16.6 | 46.7±0.0943 | 23.6±0.109 |

| badness | | α | β | k | τ | σ | quality |
|---|---|---|---|---|---|---|---|
| 0.36% | error | 0.98% | 0.83% | 0.31% | 0.2% | 0.46% | 3.15% |

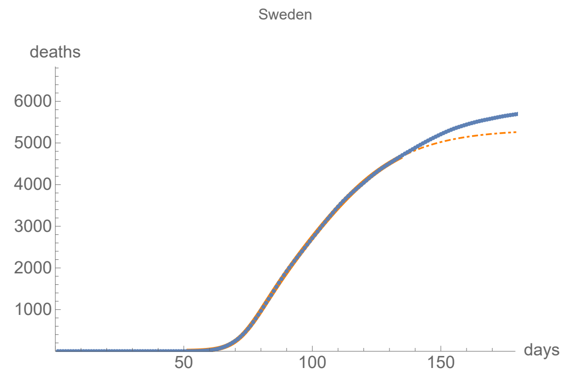


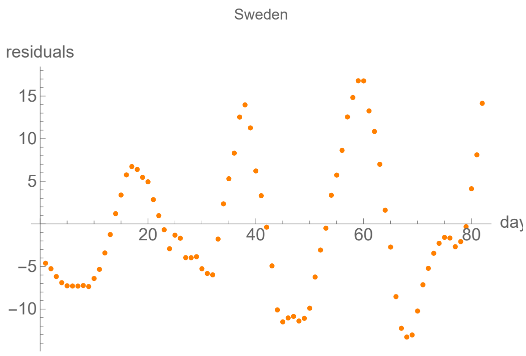


### United Kingdom

lag from integral overlap between 1 and 1 days, maximum overlap on day 1

lag from integral overlap, using data averaged over 7 days, between 0 and 6 days, maximum overlap on day 3

lag from fits to the function I: 0.996832 days

lag from fits to the function I (averaged data): -1.64298 days

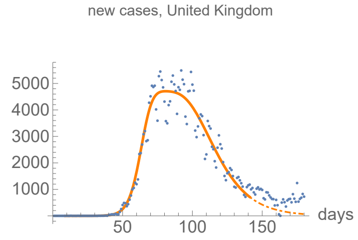


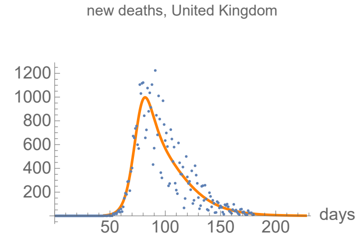


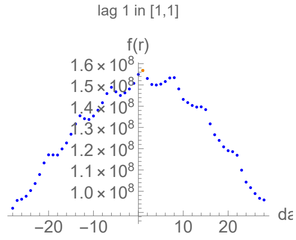


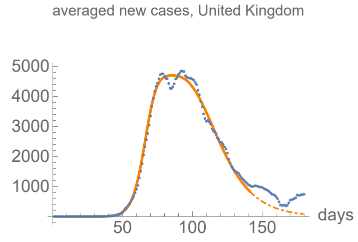


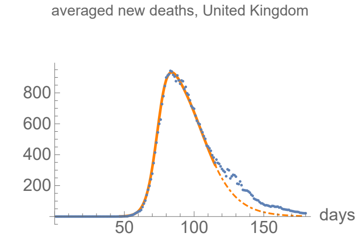


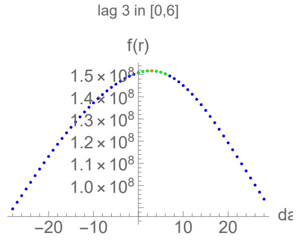

#### Parameters of fits to cases data.

The best fit ends on Wed 10 Jun 2020.

| α | β | k | τ | σ |
|---|---|---|---|---|
| 0.236±0.00435 | 0.0625±0.00049 | 278000.±508. | 82.3±0.0682 | 56.9±0.184 |

| badness | | α | β | k | τ | σ | quality |
|---|---|---|---|---|---|---|---|
| 1.% | error | 1.8% | 0.78% | 0.18% | 0.083% | 0.32% | 4.26% |

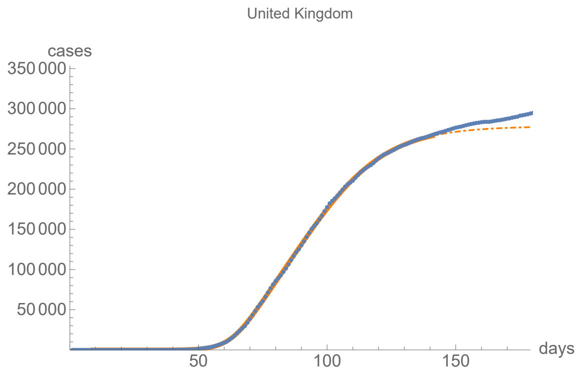


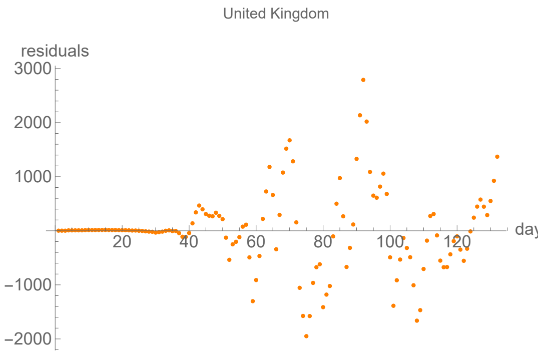


#### Parameters of fits to averaged cases data.

The best fit ends on Wed 10 Jun 2020.

| α | β | k | τ | σ |
|---|---|---|---|---|
| 0.232±0.00304 | 0.0625±0.000371 | 278000.±405. | 82.3±0.0499 | 56.6±0.138 |

| badness | | α | β | k | τ | σ | quality |
|---|---|---|---|---|---|---|---|
| 0.54% | error | 1.3% | 0.59% | 0.15% | 0.061% | 0.24% | 2.89% |

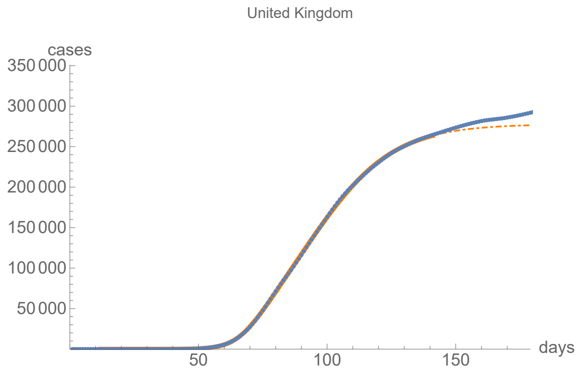


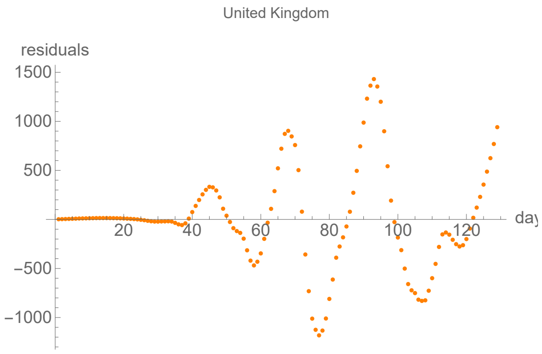


#### Parameters of fits to death data.

The best fit ends on Fri 4 Sep 2020.

| α | β | k | τ | σ |
|---|---|---|---|---|
| 0.222±0.00263 | 0.0522±0.00041 | 41400.±21.5 | 42.9±0.198 | 32.1±0.138 |

| badness | | α | β | k | τ | σ | quality |
|---|---|---|---|---|---|---|---|
| 1.5% | error | 1.2% | 0.79% | 0.052% | 0.46% | 0.43% | 4.41% |

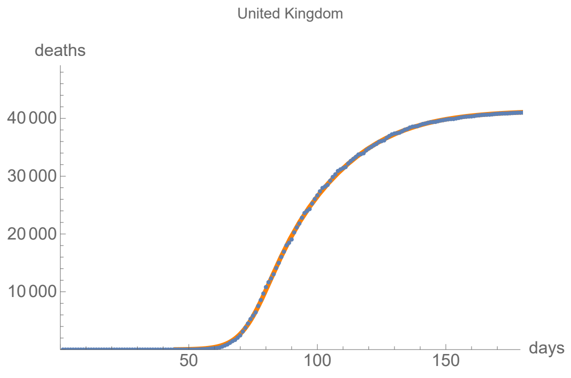


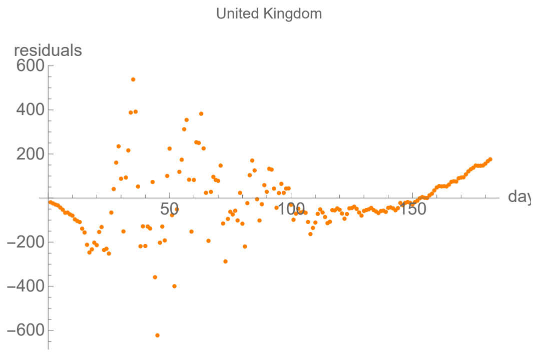


#### Parameters of fits to averaged death data.

The best fit ends on Fri 15 May 2020.

| α | β | k | τ | σ |
|---|---|---|---|---|
| 0.274±0.00152 | 0.0779±0.00185 | 37100.±78.6 | 28.8±0.0828 | 43.9±0.0354 |

| badness | | α | β | k | τ | σ | quality |
|---|---|---|---|---|---|---|---|
| 0.18% | error | 0.55% | 2.4% | 0.21% | 0.29% | 0.081% | 1.87% |

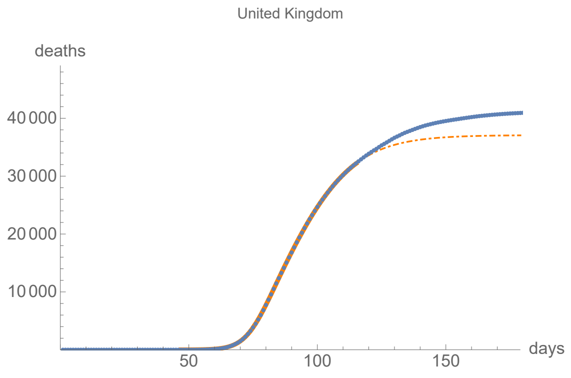


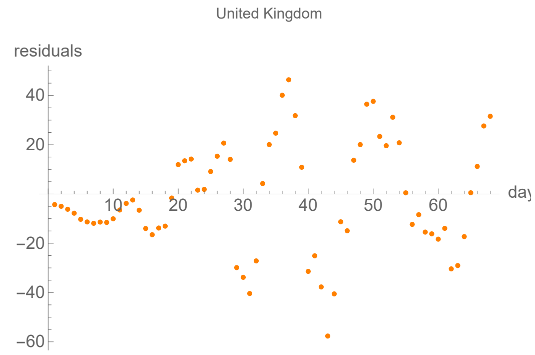


# US

lag from integral overlap between -1 and 7 days, maximum overlap on day 6

lag from integral overlap, using data averaged over 7 days, between 0 and 7 days, maximum overlap on day 4

lag from fits to the function I: 6.72005 days

lag from fits to the function I (averaged data): 6.4673 days

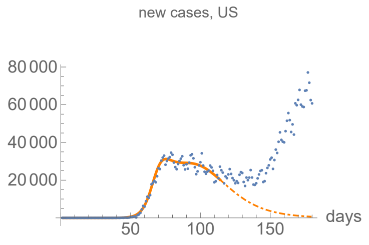


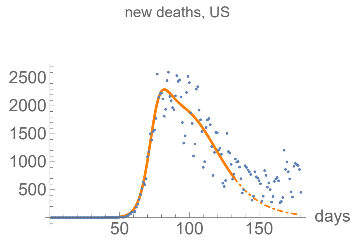


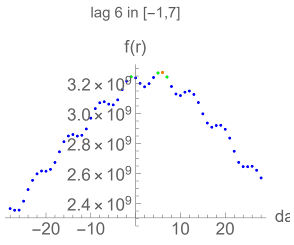


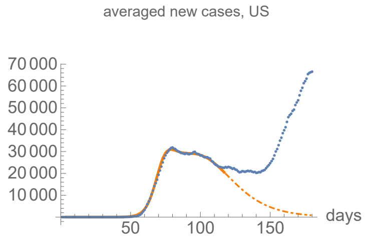


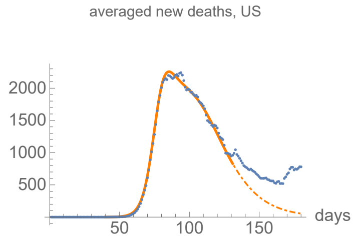


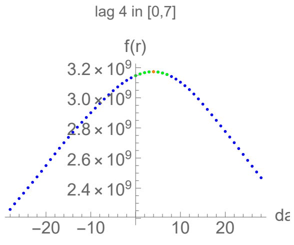

#### Parameters of fits to cases data.

The best fit ends on Thu 14 May 2020.

| α | β | k | τ | σ |
|---|---|---|---|---|
| 0.271±0.0032 | 0.0606±0.000644 | 1860000.±12400. | 92.6±0.221 | 66.7±0.112 |

| badness | | α | β | k | τ | σ | quality |
|---|---|---|---|---|---|---|---|
| 0.46% | error | 1.2% | 1.1% | 0.67% | 0.24% | 0.17% | 3.78% |

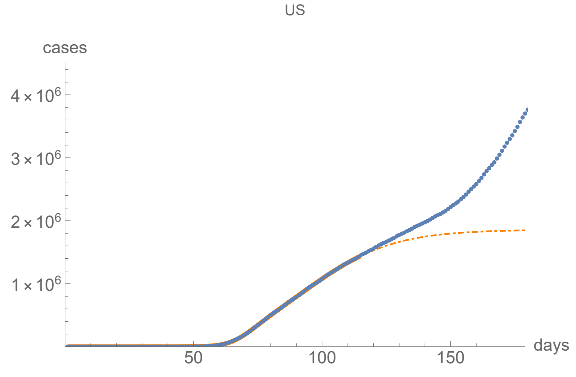


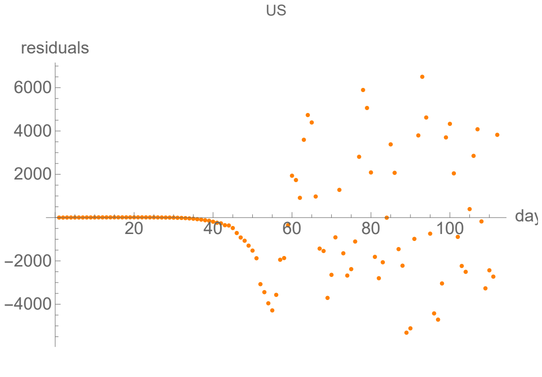


#### Parameters of fits to averaged cases data.

The best fit ends on Sun 17 May 2020.

| α | β | k | τ | σ |
|---|---|---|---|---|
| 0.262±0.00158 | 0.0596±0.000342 | 1880000.±6810. | 93.9±0.122 | 67.7±0.0621 |

| badness | | α | β | k | τ | σ | quality |
|---|---|---|---|---|---|---|---|
| 0.24% | error | 0.6% | 0.57% | 0.36% | 0.13% | 0.092% | 2.% |

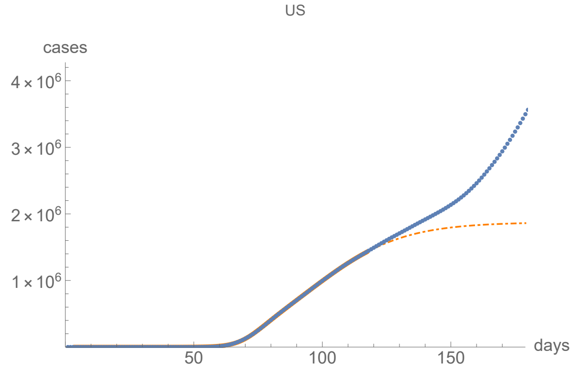


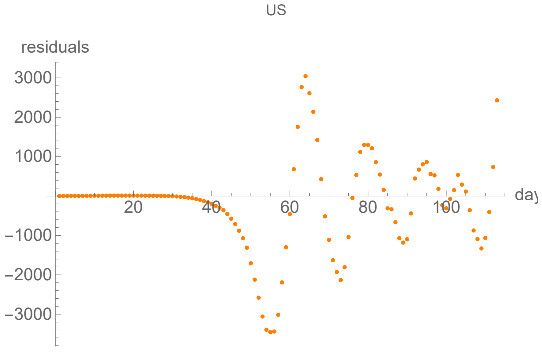


#### Parameters of fits to death data.

The best fit ends on Mon 1 Jun 2020.

| α | β | k | τ | σ |
|---|---|---|---|---|
| 0.251±0.00516 | 0.0597±0.000935 | 122000.±669. | 57.2±0.151 | 35.2±0.214 |

| badness | | α | β | k | τ | σ | quality |
|---|---|---|---|---|---|---|---|
| 0.8% | error | 2.1% | 1.6% | 0.55% | 0.26% | 0.61% | 5.85% |

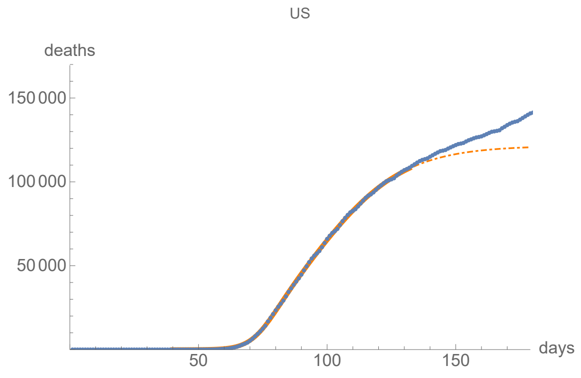


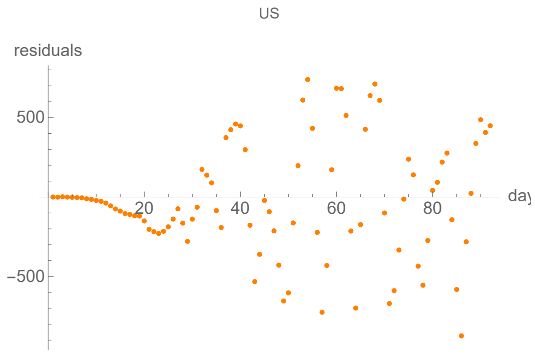


#### Parameters of fits to averaged death data.

The best fit ends on Sun 31 May 2020.

| α | β | k | τ | σ |
|---|---|---|---|---|
| 0.25±0.002 | 0.0609±0.000406 | 120000.±303. | 59.8±0.0677 | 37.8±0.0897 |

| badness | | α | β | k | τ | σ | quality |
|---|---|---|---|---|---|---|---|
| 0.29% | error | 0.8% | 0.67% | 0.25% | 0.11% | 0.24% | 2.36% |

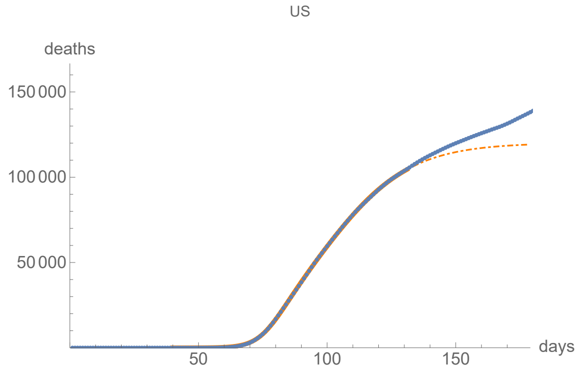


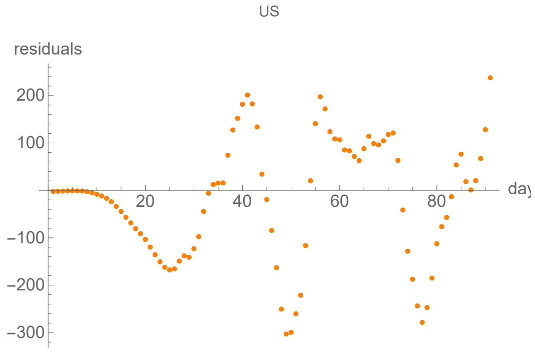


### Integral averages, not including US counties

#### Afghanistan

no clear lag in the first six months of the pandemic from integral overlap

no clear lag in the first six months of the pandemic from integral overlap using data averaged over 7 days

no meaningful fits to the function I available

no meaningful fits to the function I available (averaged data)

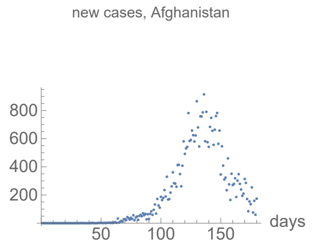


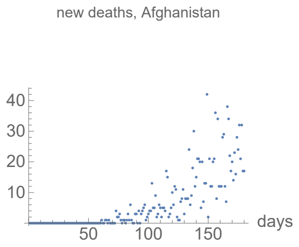


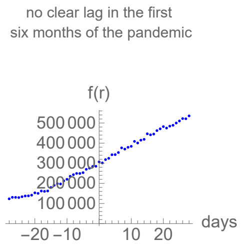

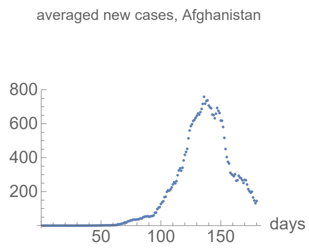


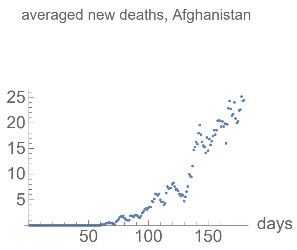


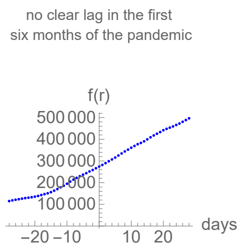


### Algeria

lag from integral overlap between 1 and 7 days, maximum overlap on day 3

lag from integral overlap, using data averaged over 7 days, between 3 and 8 days, maximum overlap on day 5

no meaningful fits to the function I available

no meaningful fits to the function I available (averaged data)

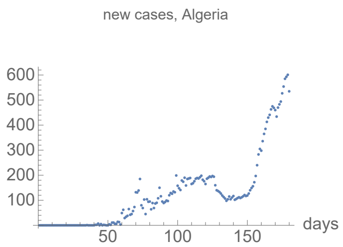


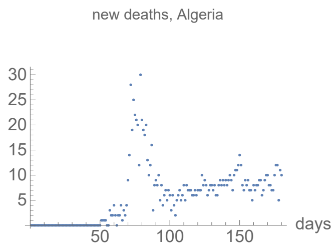


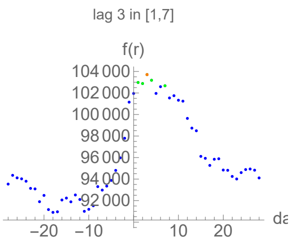


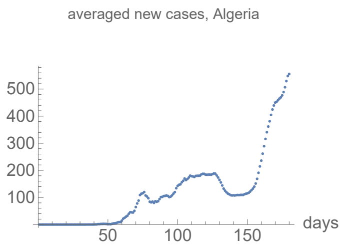


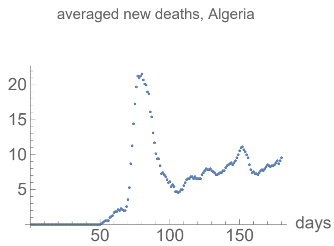


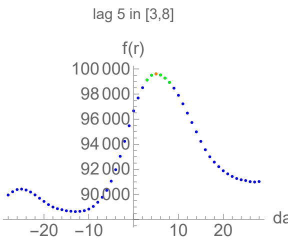


### Argentina

no clear lag in the first six months of the pandemic from integral overlap

no clear lag in the first six months of the pandemic from integral overlap using data averaged over 7 days

no meaningful fits to the function I available

no meaningful fits to the function I available (averaged data)

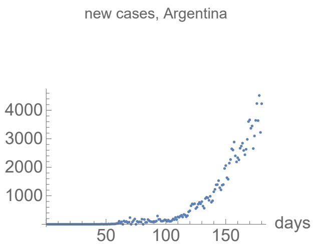


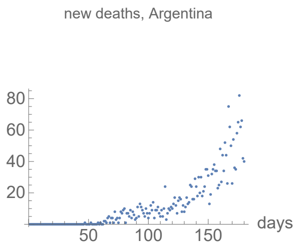


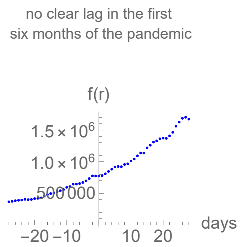


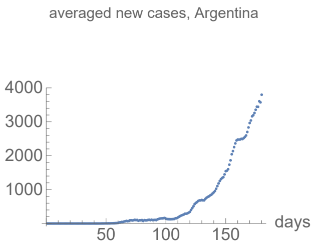


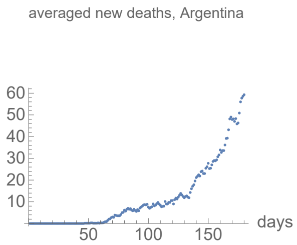


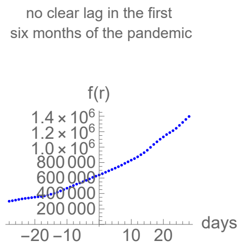


### Armenia

no clear lag in the first six months of the pandemic from integral overlap

no clear lag in the first six months of the pandemic from integral overlap using data averaged over 7 days

no meaningful fits to the function I available

no meaningful fits to the function I available (averaged data)

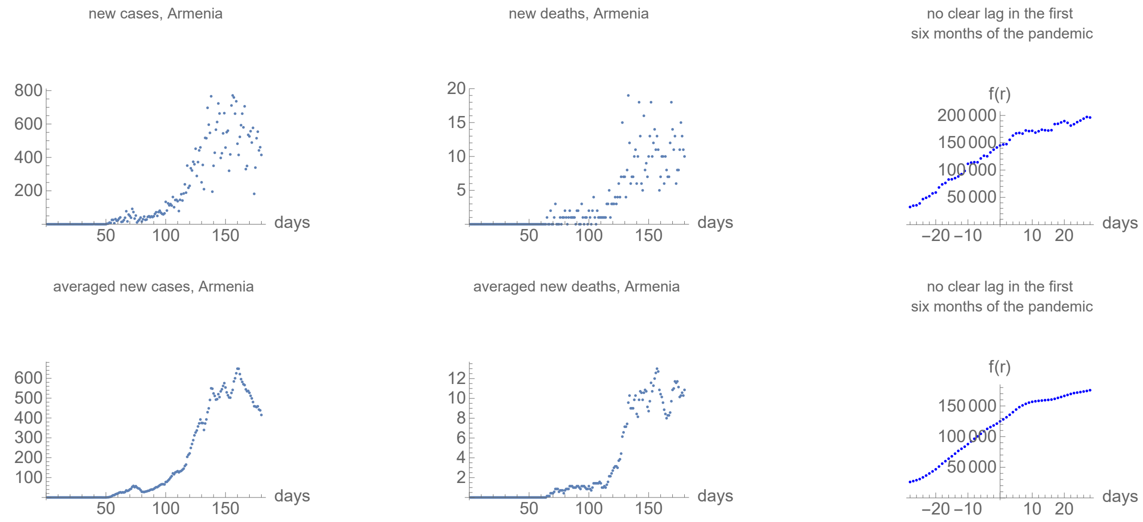


### Bangladesh

no clear lag in the first six months of the pandemic from integral overlap

no clear lag in the first six months of the pandemic from integral overlap using data averaged over 7 days

no meaningful fits to the function I available

no meaningful fits to the function I available (averaged data)

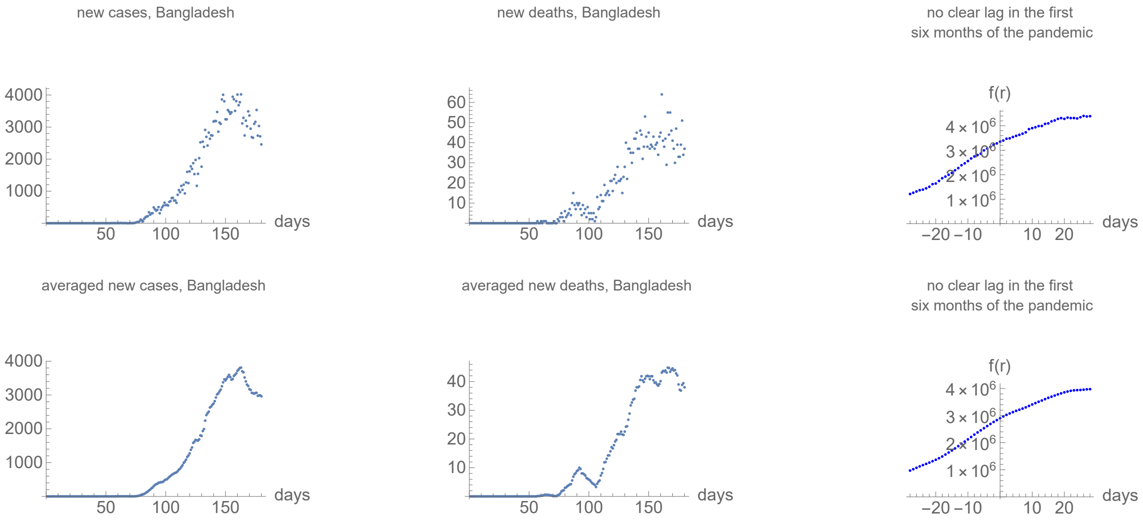


### Bolivia

no clear lag in the first six months of the pandemic from integral overlap

no clear lag in the first six months of the pandemic from integral overlap using data averaged over 7 days

no meaningful fits to the function I available

no meaningful fits to the function I available (averaged data)

new cases, Bolivia

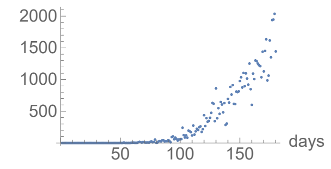


new deaths, Bolivia

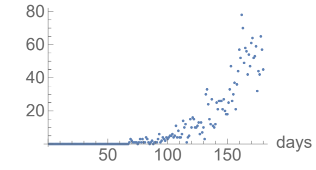


no clear lag in the first
six months of the pandemic

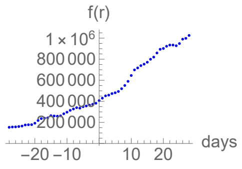


averaged new cases, Bolivia

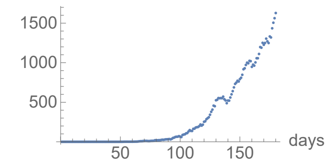


averaged new deaths, Bolivia

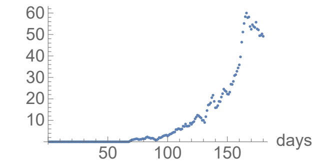


no clear lag in the first
six months of the pandemic

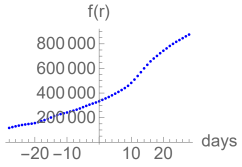


### Brazil

```
no clear lag in the first six months of the pandemic from integral overlap

no clear lag in the first six months of the pandemic from integral overlap using data averaged over 7 days

no meaningful fits to the function I available

no meaningful fits to the function I available (averaged data)
```

new cases, Brazil

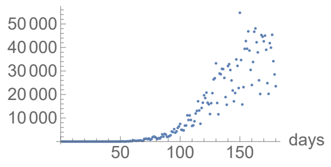


new deaths, Brazil

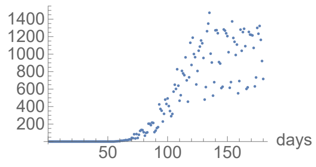


no clear lag in the first
six months of the pandemic

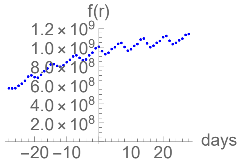


averaged new cases, Brazil

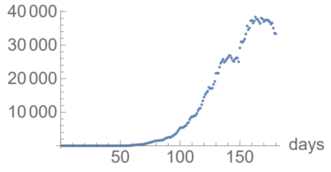


averaged new deaths, Brazil

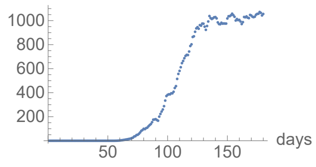


no clear lag in the first
six months of the pandemic

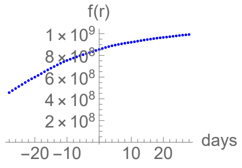


### Chile

```
no clear lag in the first six months of the pandemic from integral overlap

no clear lag in the first six months of the pandemic from integral overlap using data averaged over 7 days

no meaningful fits to the function I available

no meaningful fits to the function I available (averaged data)
```

new cases, Chile

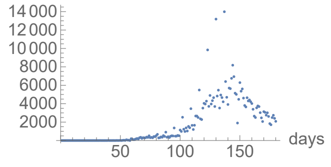


new deaths, Chile

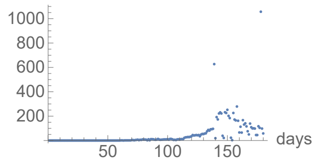


no clear lag in the first
six months of the pandemic

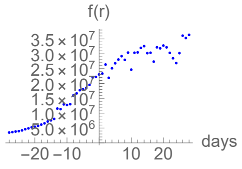

averaged new cases, Chile

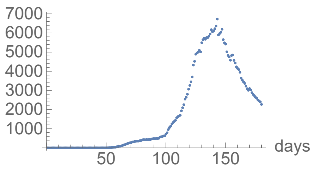


averaged new deaths, Chile

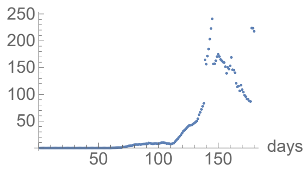


no clear lag in the first
six months of the pandemic

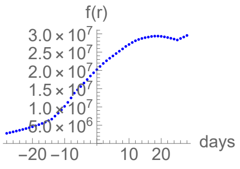


### Colombia

no clear lag in the first six months of the pandemic from integral overlap

no clear lag in the first six months of the pandemic from integral overlap using data averaged over 7 days

no meaningful fits to the function I available

no meaningful fits to the function I available (averaged data)

new cases, Colombia

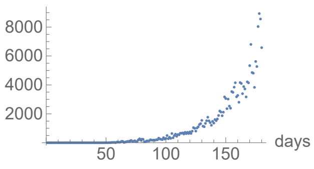


new deaths, Colombia

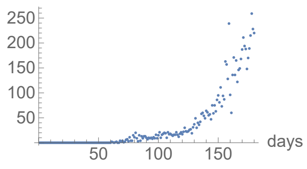


no clear lag in the first
six months of the pandemic

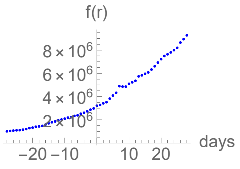


averaged new cases, Colombia

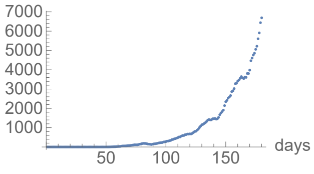


averaged new deaths, Colombia

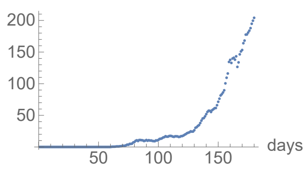


no clear lag in the first
six months of the pandemic

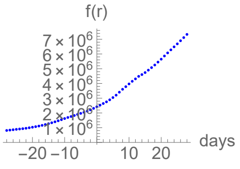


### Dominican Republic

no clear lag in the first six months of the pandemic from integral overlap

no clear lag in the first six months of the pandemic from integral overlap using data averaged over 7 days

no meaningful fits to the function I available

no meaningful fits to the function I available (averaged data)

new cases, Dominican Republic

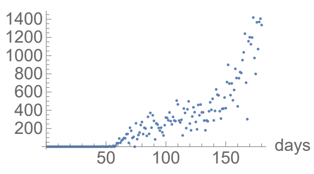


new deaths, Dominican Republic

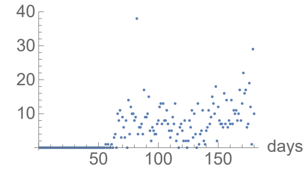


no clear lag in the first
six months of the pandemic

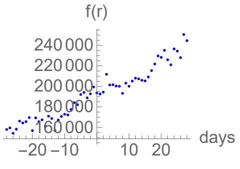


averaged new cases, Dominican Republic

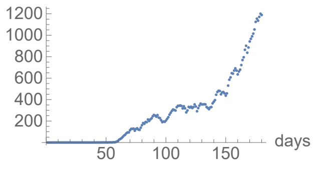


averaged new deaths, Dominican Republic

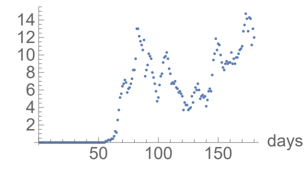


no clear lag in the first
six months of the pandemic

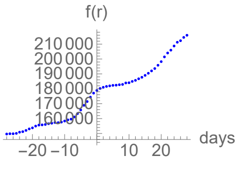


### Ecuador

```
lag from integral overlap between 16 and 16 days, maximum overlap on day 16

lag from integral overlap, using data averaged over 7 days, between 18 and 18 days, maximum overlap on day 18

no meaningful fits to the function I available

no meaningful fits to the function I available (averaged data)
```

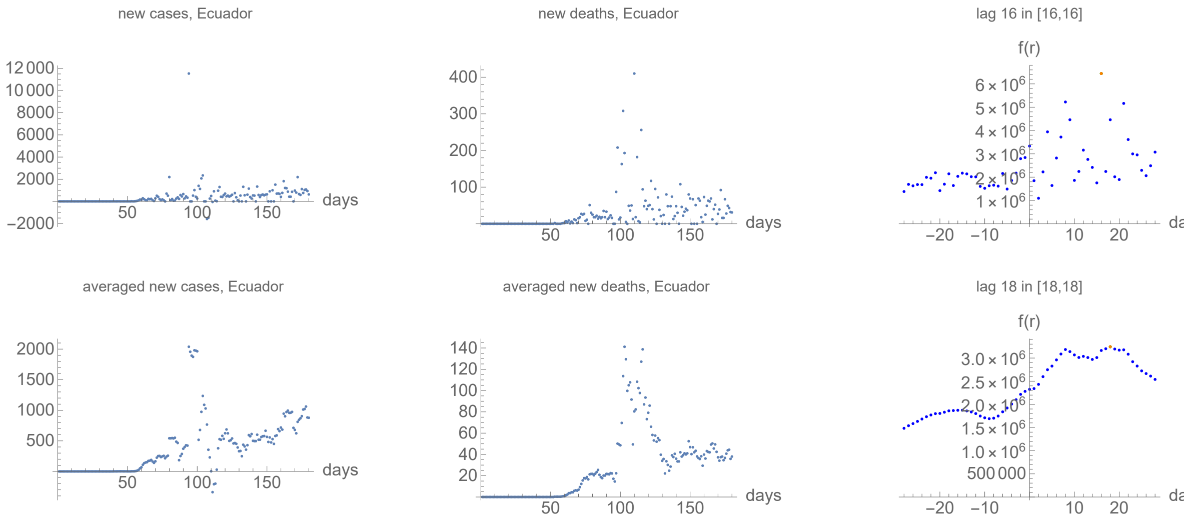


### Egypt

```
no clear lag in the first six months of the pandemic from integral overlap

no clear lag in the first six months of the pandemic from integral overlap using data averaged over 7 days

no meaningful fits to the function I available

no meaningful fits to the function I available (averaged data)
```

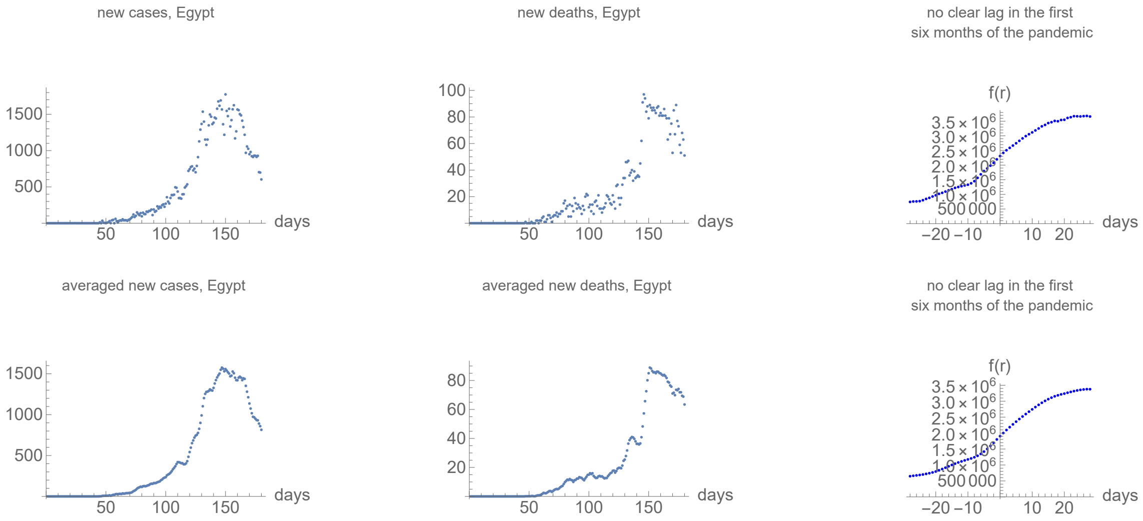


### Guatemala

```
no clear lag in the first six months of the pandemic from integral overlap

no clear lag in the first six months of the pandemic from integral overlap using data averaged over 7 days

no meaningful fits to the function I available

no meaningful fits to the function I available (averaged data)
```

new cases, Guatemala

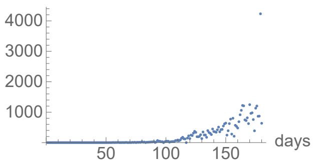


new deaths, Guatemala

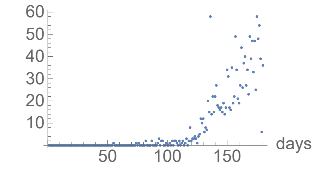


no clear lag in the first
six months of the pandemic

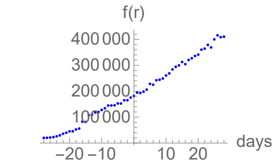


averaged new cases, Guatemala

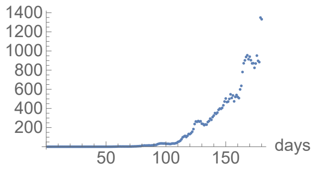


averaged new deaths, Guatemala

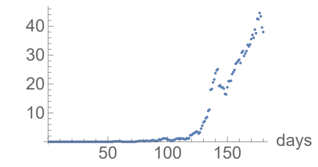


no clear lag in the first
six months of the pandemic

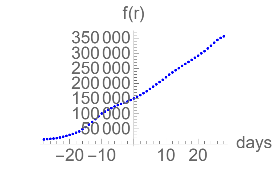


### Honduras

lag from integral overlap between 20 and 20 days, maximum overlap on day 20

no clear lag in the first six months of the pandemic from integral overlap using data averaged over 7 days

no meaningful fits to the function I available

no meaningful fits to the function I available (averaged data)

new cases, Honduras

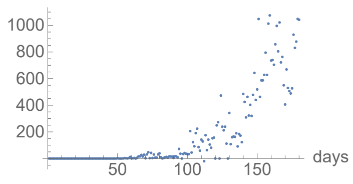


new deaths, Honduras

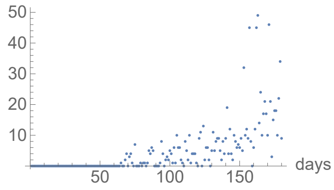


lag 20 in [20,20]

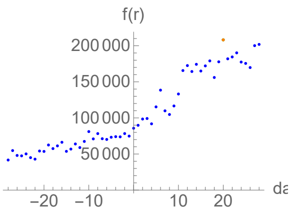


averaged new cases, Honduras

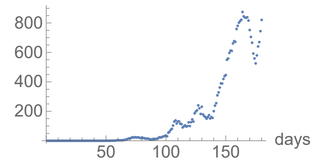


averaged new deaths, Honduras

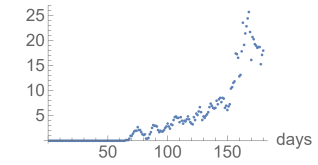


no clear lag in the first
six months of the pandemic

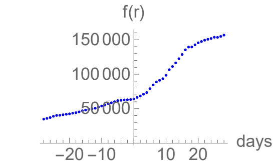


### India

no clear lag in the first six months of the pandemic from integral overlap

no clear lag in the first six months of the pandemic from integral overlap using data averaged over 7 days

no meaningful fits to the function I available

no meaningful fits to the function I available (averaged data)

new cases, India

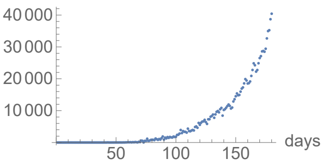


new deaths, India

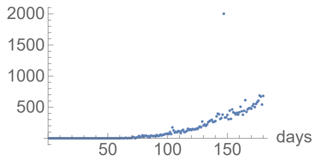


no clear lag in the first
six months of the pandemic

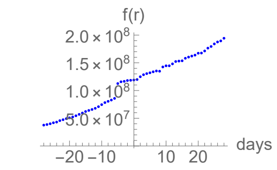

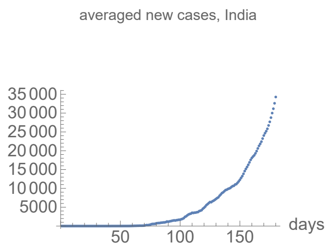


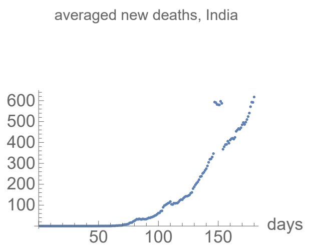


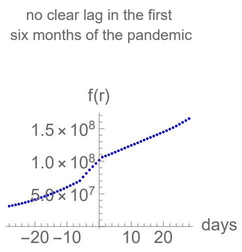


### Indonesia

```
no clear lag in the first six months of the pandemic from integral overlap

no clear lag in the first six months of the pandemic from integral overlap using data averaged over 7 days

no meaningful fits to the function I available

no meaningful fits to the function I available (averaged data)
```

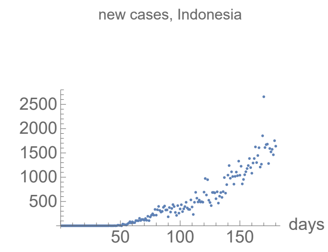


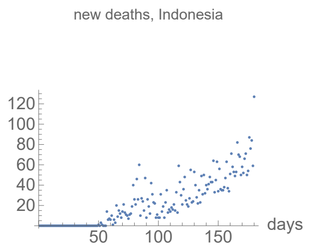


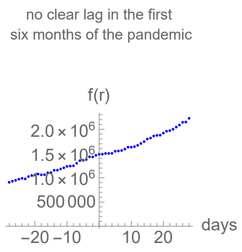


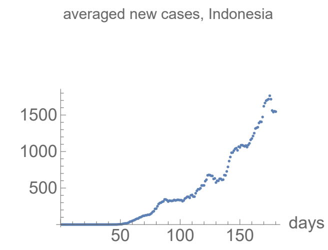


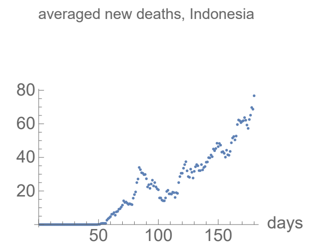


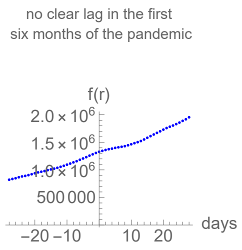


### Iran

```
no clear lag in the first six months of the pandemic from integral overlap

no clear lag in the first six months of the pandemic from integral overlap using data averaged over 7 days

no meaningful fits to the function I available

no meaningful fits to the function I available (averaged data)
```

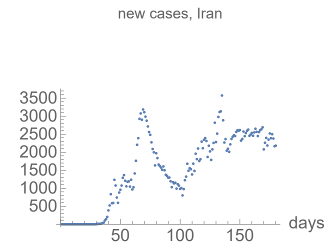


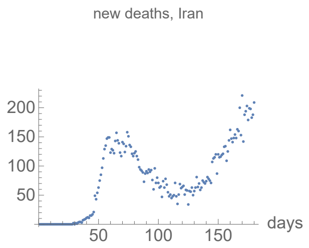


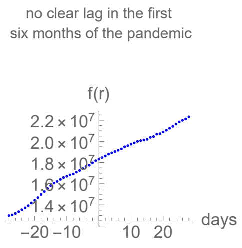


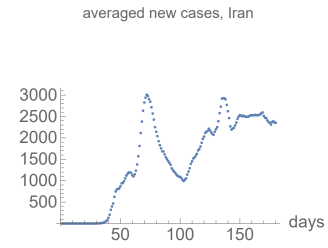


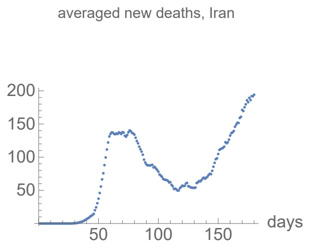


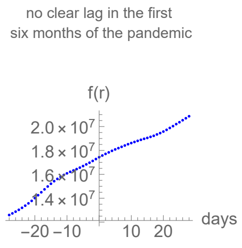


### Iraq

no clear lag in the first six months of the pandemic from integral overlap

no clear lag in the first six months of the pandemic from integral overlap using data averaged over 7 days

no meaningful fits to the function I available

no meaningful fits to the function I available (averaged data)

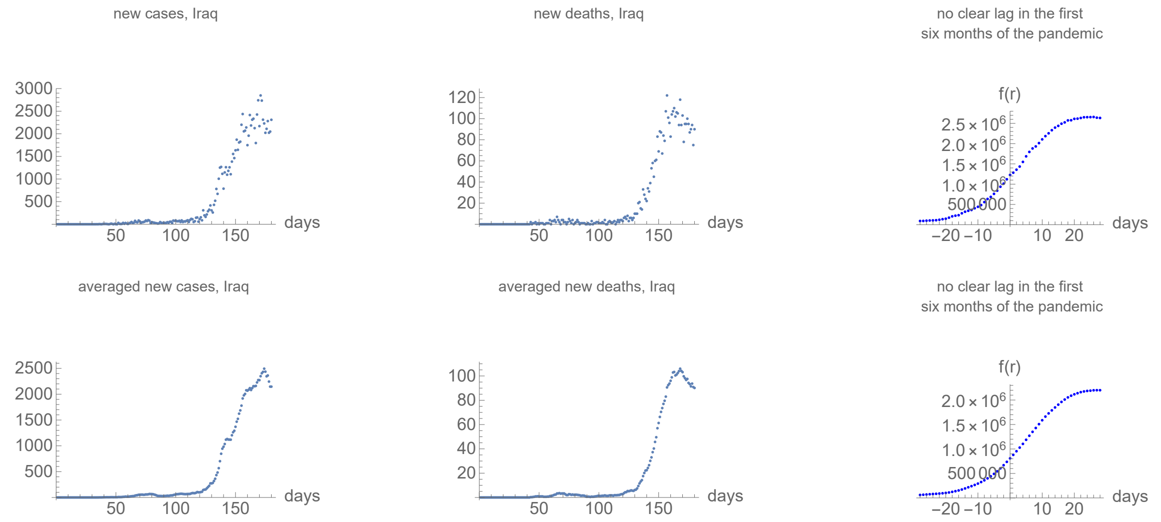


### Japan

lag from integral overlap between 11 and 16 days, maximum overlap on day 14

lag from integral overlap, using data averaged over 7 days, between 12 and 17 days, maximum overlap on day 14

no meaningful fits to the function I available

no meaningful fits to the function I available (averaged data)

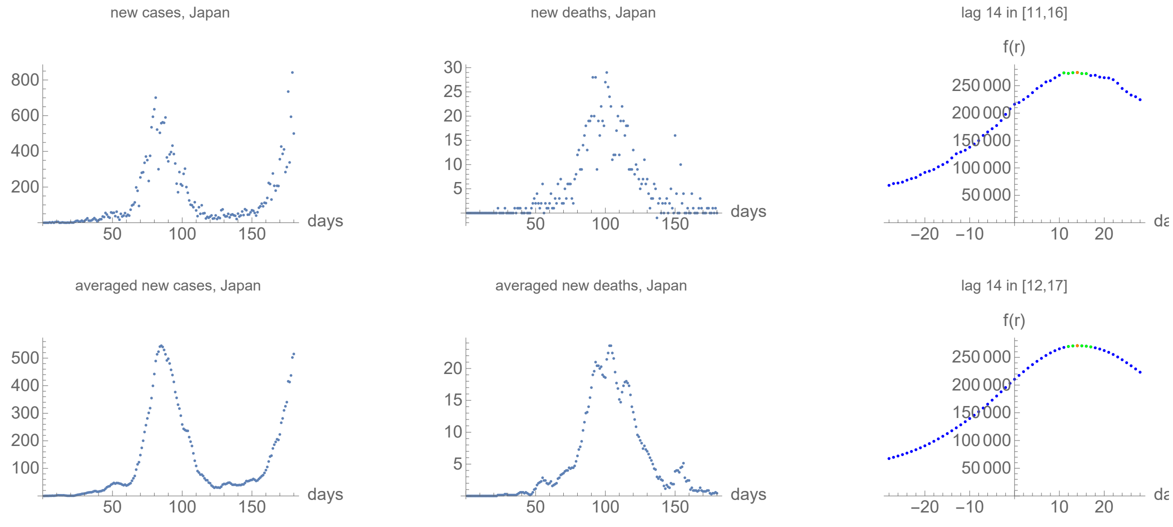


### Mexico

no clear lag in the first six months of the pandemic from integral overlap

no clear lag in the first six months of the pandemic from integral overlap using data averaged over 7 days

no meaningful fits to the function I available

no meaningful fits to the function I available (averaged data)

new cases, Mexico

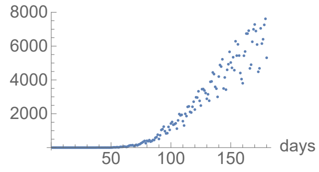


new deaths, Mexico

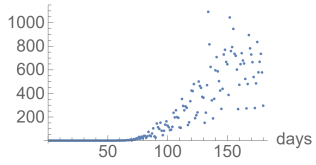


no clear lag in the first
six months of the pandemic

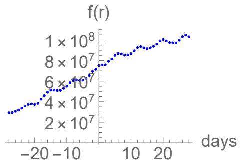


averaged new cases, Mexico

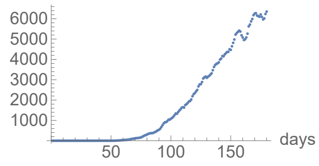


averaged new deaths, Mexico

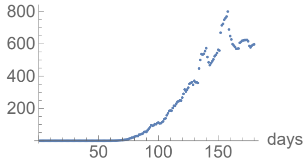


no clear lag in the first
six months of the pandemic

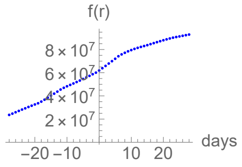


### Moldova

`lag from integral overlap between 20 and 20 days, maximum overlap on day 20`

`lag from integral overlap, using data averaged over 7 days, between 21 and 26 days, maximum overlap on day 23`

`no meaningful fits to the function I available`

`no meaningful fits to the function I available (averaged data)`

new cases, Moldova

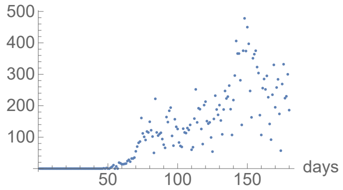


new deaths, Moldova

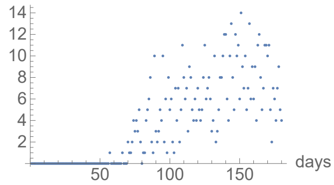


lag 20 in [20,20]

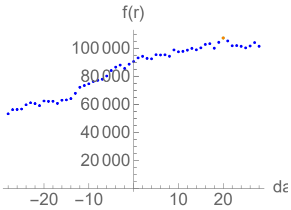


averaged new cases, Moldova

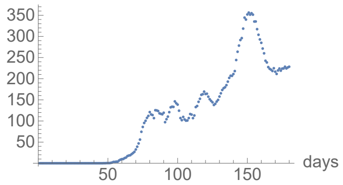


averaged new deaths, Moldova

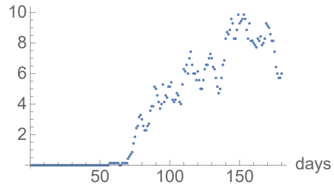


lag 23 in [21,26]

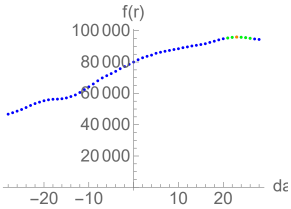


### Nigeria

`lag from integral overlap between 21 and 21 days, maximum overlap on day 21`

`no clear lag in the first six months of the pandemic from integral overlap using data averaged over 7 days`

`no meaningful fits to the function I available`

`no meaningful fits to the function I available (averaged data)`

new cases, Nigeria

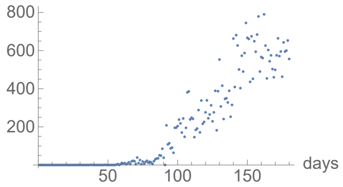


new deaths, Nigeria

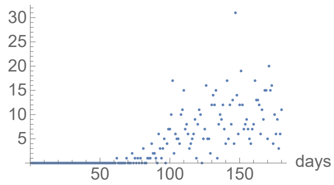


lag 21 in [21,21]

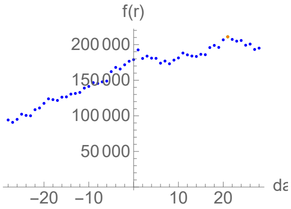

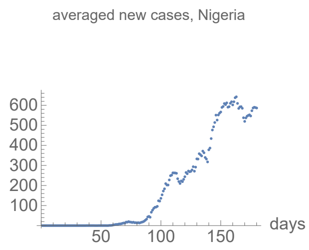


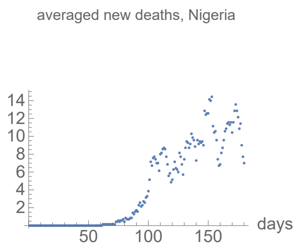


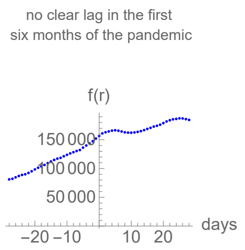


### Pakistan

```
lag from integral overlap between 10 and 21 days, maximum overlap on day 10

lag from integral overlap, using data averaged over 7 days, between 9 and 16 days, maximum overlap on day 11

no meaningful fits to the function I available

no meaningful fits to the function I available (averaged data)
```

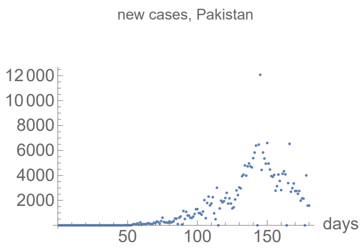


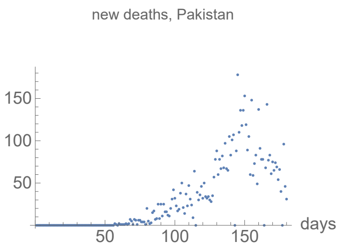


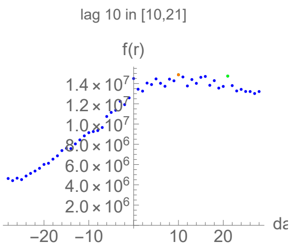


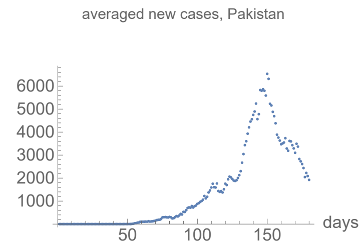


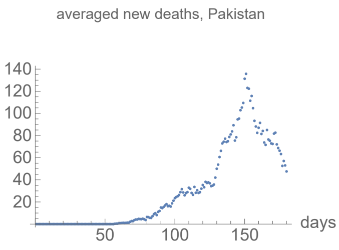


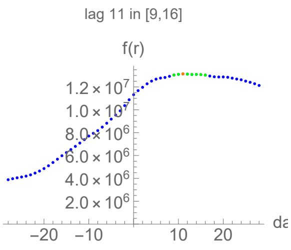


### Panama

```
no clear lag in the first six months of the pandemic from integral overlap

no clear lag in the first six months of the pandemic from integral overlap using data averaged over 7 days

no meaningful fits to the function I available

no meaningful fits to the function I available (averaged data)
```

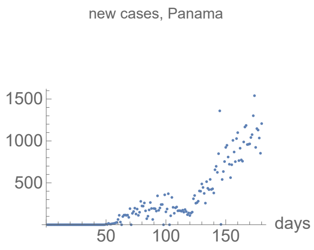


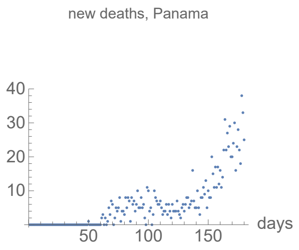


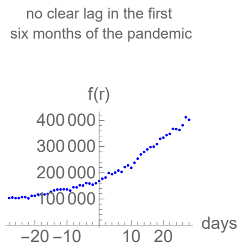


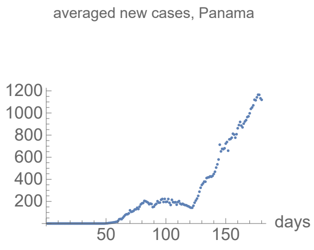


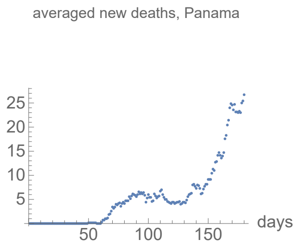


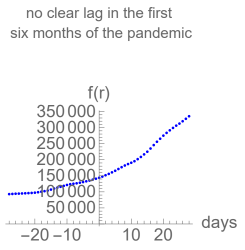


### Peru

no clear lag in the first six months of the pandemic from integral overlap

no clear lag in the first six months of the pandemic from integral overlap using data averaged over 7 days

no meaningful fits to the function I available

no meaningful fits to the function I available (averaged data)

new cases, Peru

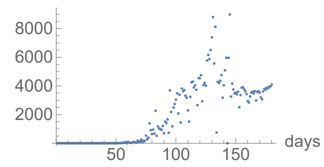


new deaths, Peru

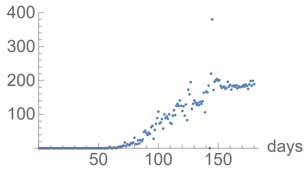


no clear lag in the first six months of the pandemic

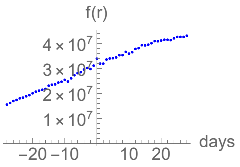


averaged new cases, Peru

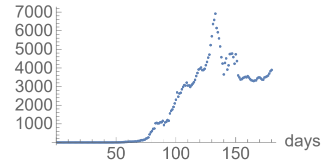


averaged new deaths, Peru

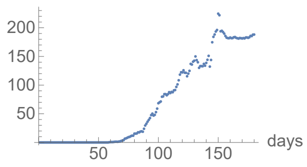


no clear lag in the first six months of the pandemic

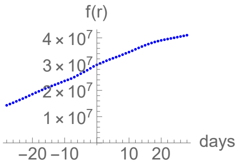


### Philippines

no clear lag in the first six months of the pandemic from integral overlap

no clear lag in the first six months of the pandemic from integral overlap using data averaged over 7 days

no meaningful fits to the function I available

no meaningful fits to the function I available (averaged data)

new cases, Philippines

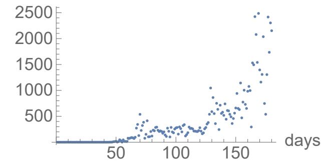


new deaths, Philippines

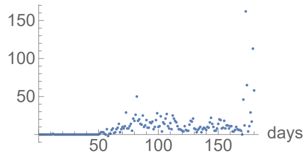


no clear lag in the first six months of the pandemic

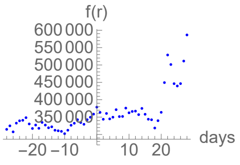


averaged new cases, Philippines

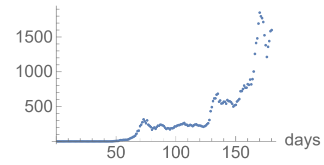


averaged new deaths, Philippines

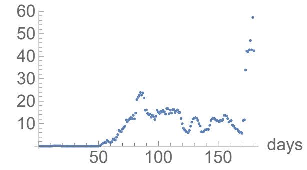


no clear lag in the first six months of the pandemic

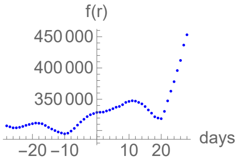


### Russia

no clear lag in the first six months of the pandemic from integral overlap

no clear lag in the first six months of the pandemic from integral overlap using data averaged over 7 days

no meaningful fits to the function I available

no meaningful fits to the function I available (averaged data)

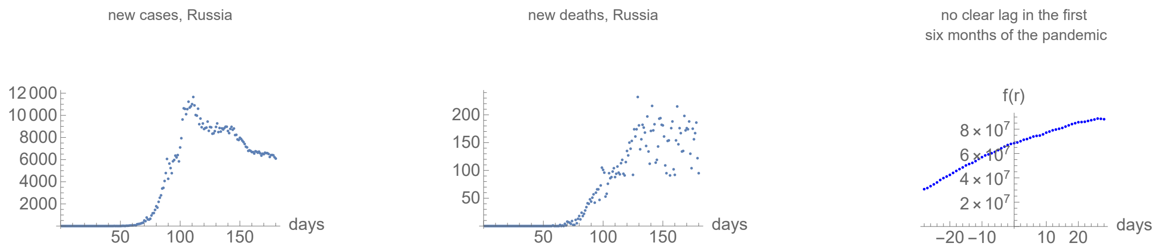


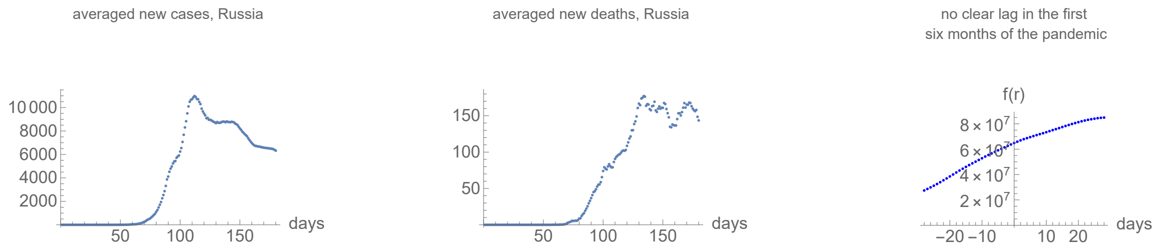


### Saudi Arabia

no clear lag in the first six months of the pandemic from integral overlap

no clear lag in the first six months of the pandemic from integral overlap using data averaged over 7 days

no meaningful fits to the function I available

no meaningful fits to the function I available (averaged data)

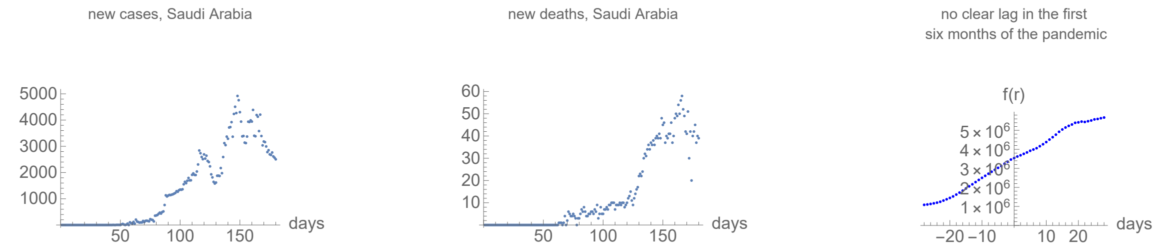


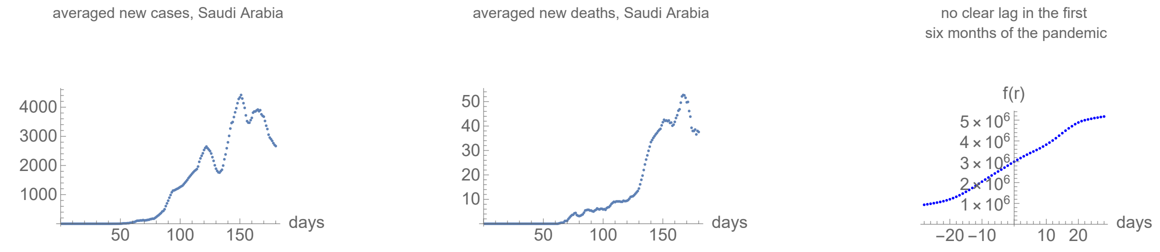


### South Africa

no clear lag in the first six months of the pandemic from integral overlap

no clear lag in the first six months of the pandemic from integral overlap using data averaged over 7 days

no meaningful fits to the function I available

no meaningful fits to the function I available (averaged data)

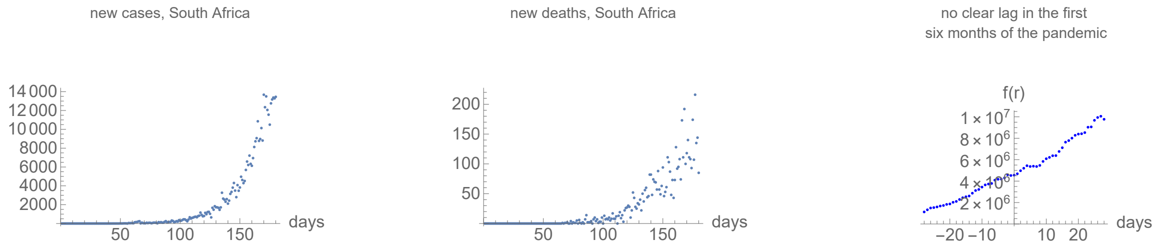

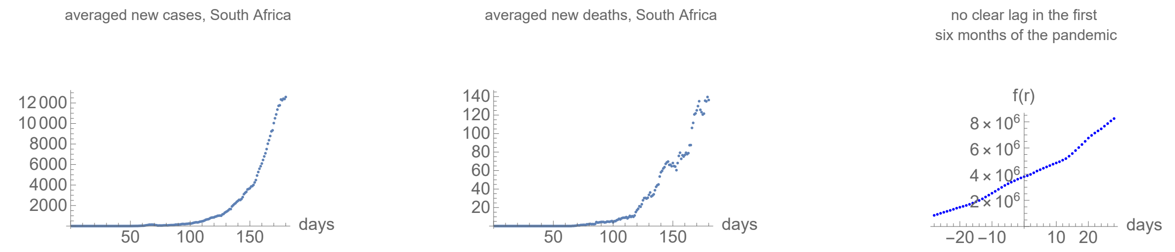


### Sudan

```
lag from integral overlap between 0 and 0 days, maximum overlap on day 0

lag from integral overlap, using data averaged over 7 days, between 7 and 11 days, maximum overlap on day 8

no meaningful fits to the function I available

no meaningful fits to the function I available (averaged data)
```

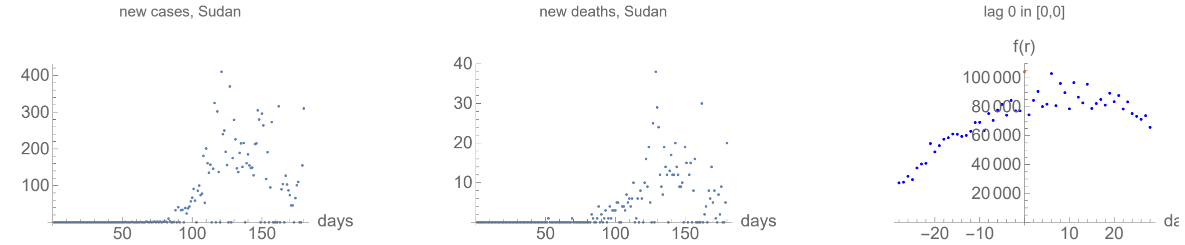


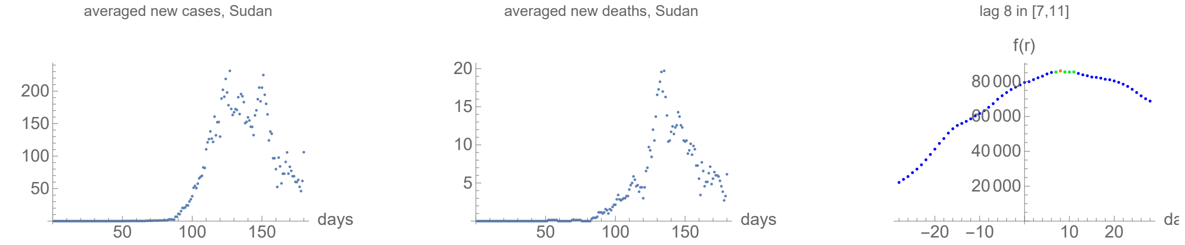


### Switzerland

```
lag from integral overlap between 11 and 11 days, maximum overlap on day 11

lag from integral overlap, using data averaged over 7 days, between 9 and 12 days, maximum overlap on day 10

no meaningful fits to the function I available

no meaningful fits to the function I available (averaged data)
```

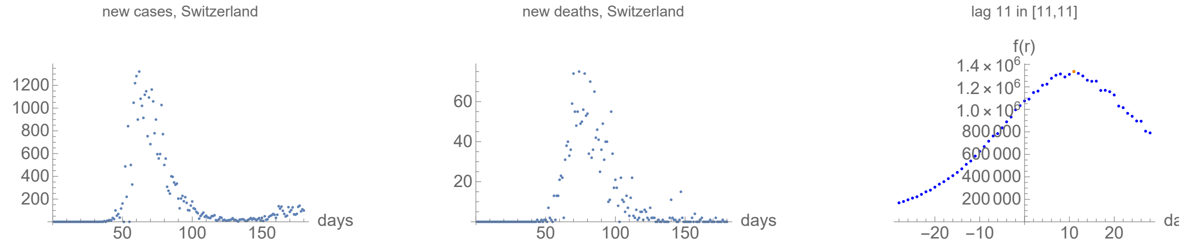


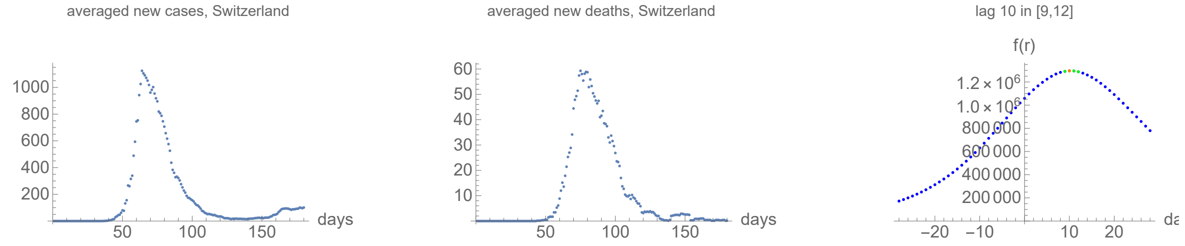


### Turkey

lag from integral overlap between 2 and 6 days, maximum overlap on day 5

lag from integral overlap, using data averaged over 7 days, between 2 and 7 days, maximum overlap on day 4

no meaningful fits to the function I available

no meaningful fits to the function I available (averaged data)

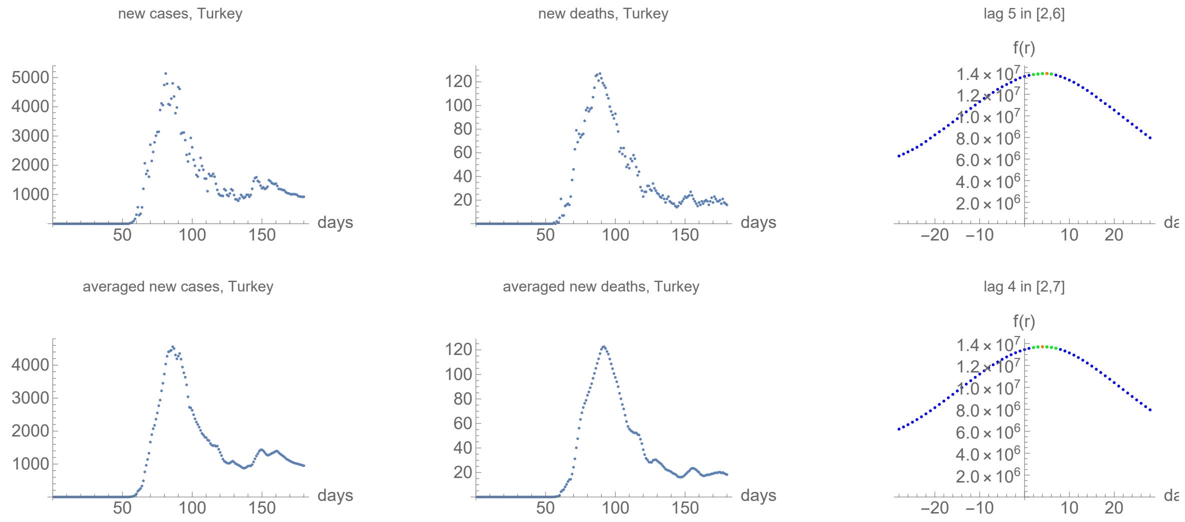


### Ukraine

no clear lag in the first six months of the pandemic from integral overlap

no clear lag in the first six months of the pandemic from integral overlap using data averaged over 7 days

no meaningful fits to the function I available

no meaningful fits to the function I available (averaged data)

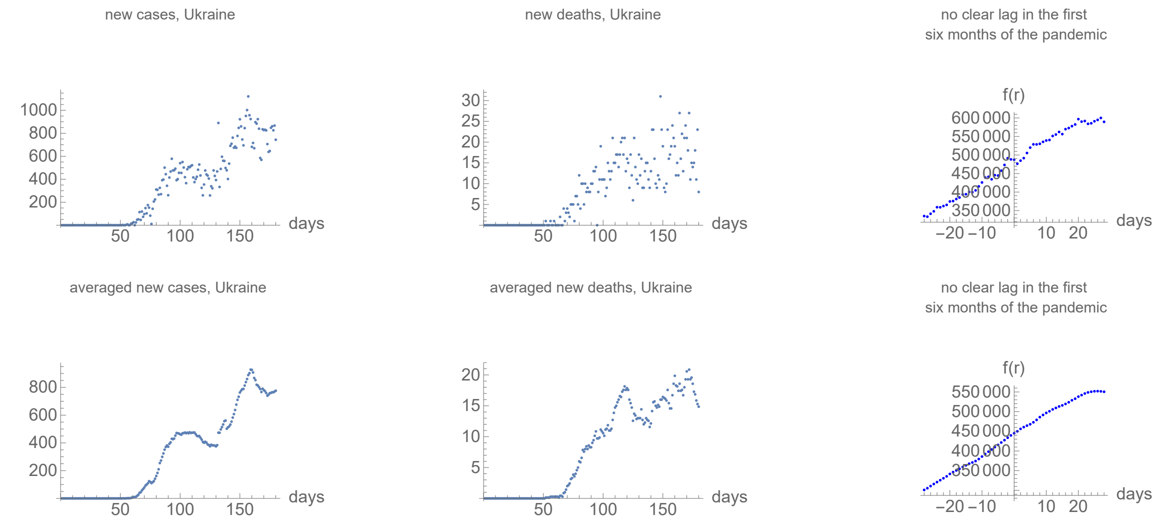


### Hubei, China

lag from integral overlap between 0 and 0 days, maximum overlap on day 0

lag from integral overlap, using data averaged over 7 days, between 3 and 6 days, maximum overlap on day 5

no meaningful fits to the function I available

no meaningful fits to the function I available (averaged data)

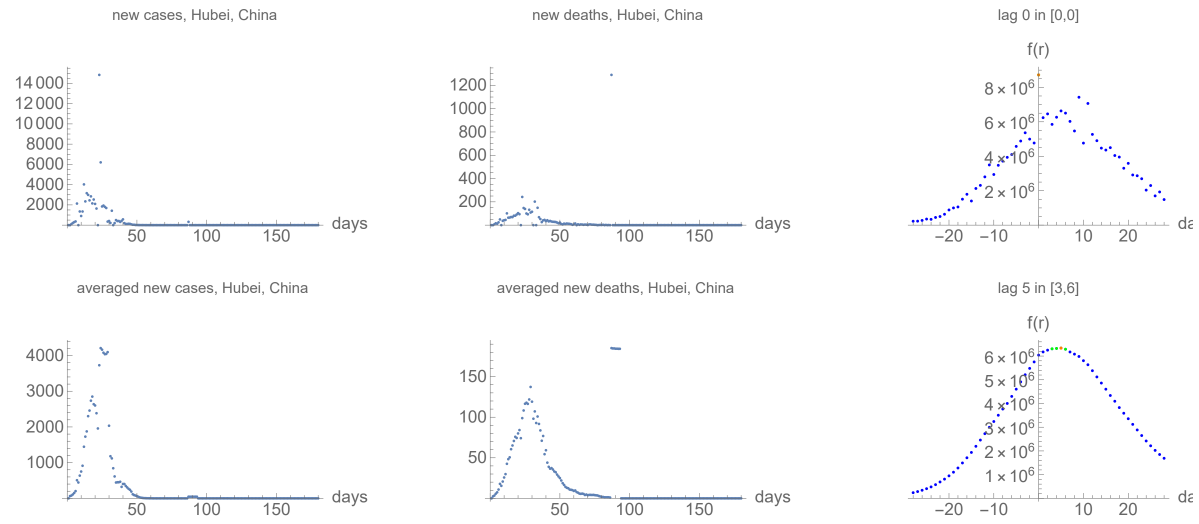


### Ontario, Canada

```
lag from integral overlap between 0 and 0 days, maximum overlap on day 0

lag from integral overlap, using data averaged over 7 days, between 3 and 11 days, maximum overlap on day 7

no meaningful fits to the function I available

no meaningful fits to the function I available (averaged data)
```

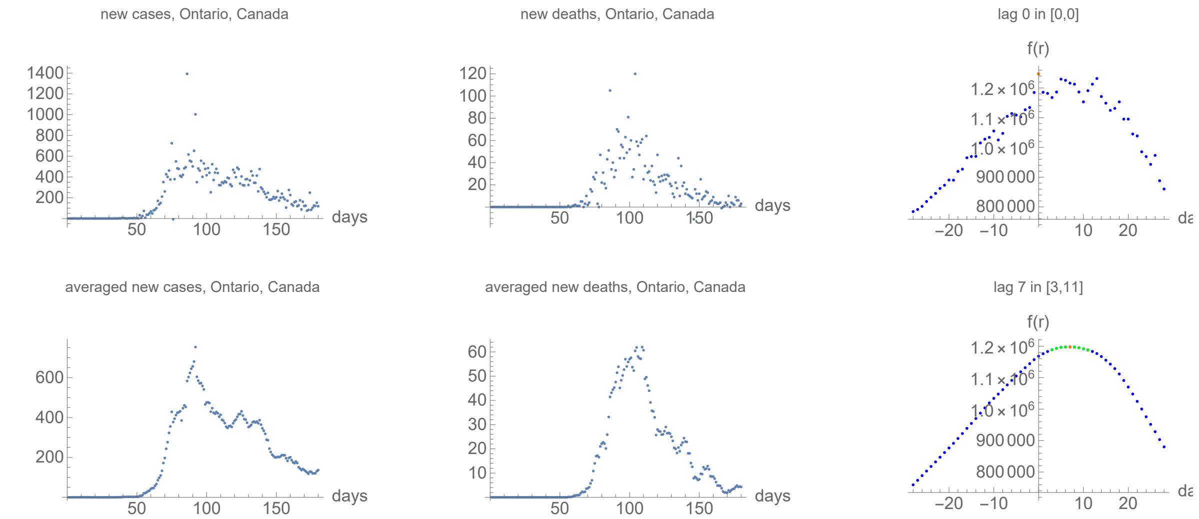


### Quebec, Canada

```
lag from integral overlap between 11 and 14 days, maximum overlap on day 11

lag from integral overlap, using data averaged over 7 days, between 7 and 14 days, maximum overlap on day 11

no meaningful fits to the function I available

no meaningful fits to the function I available (averaged data)
```

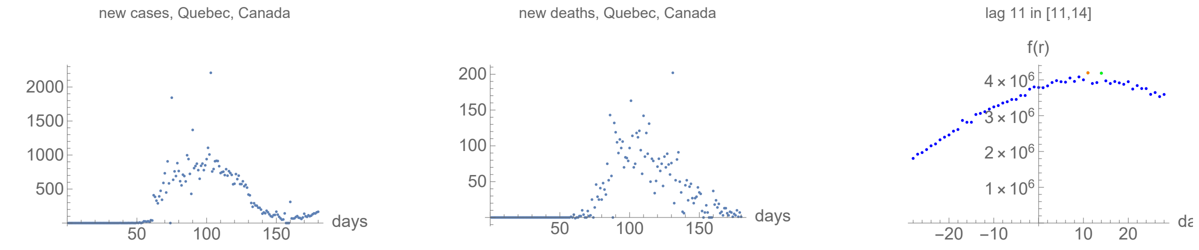

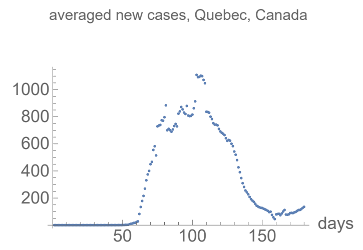


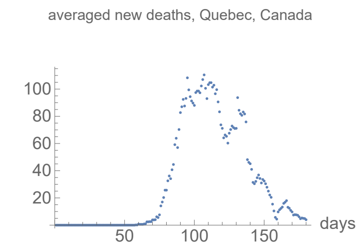


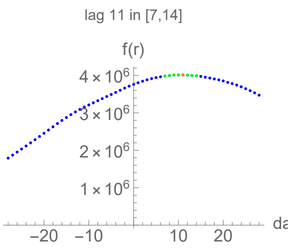


### Plots, fits and integral averages for US counties (ordered as on the JHU server)

#### Maricopa, Arizona

no clear lag in the first six months of the pandemic from integral overlap using data averaged over 7 days

lag from fits to the function I (averaged data): -1.31252 days

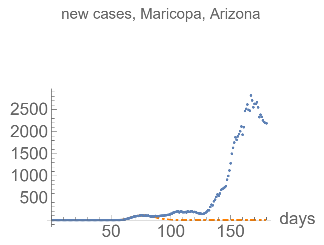


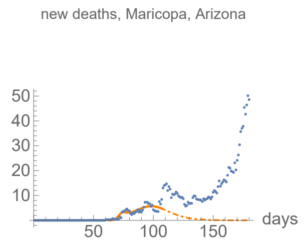


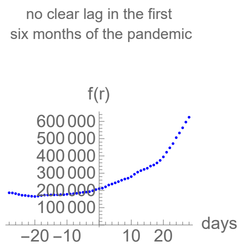


##### Parameters of fits to averaged cases data.

The best fit ends on Wed 15 Apr 2020.

| α | β | k | τ | σ |
|---|---|---|---|---|
| 0.485±0.0115 | 0.157±0.00312 | 2530.±26.3 | 33.6±0.112 | 21.3±0.2 |

| badness | | α | β | k | τ | σ | quality |
|---|---|---|---|---|---|---|---|
| 0.35% | error | 2.4% | 2.% | 1.% | 0.33% | 0.94% | 7.02% |

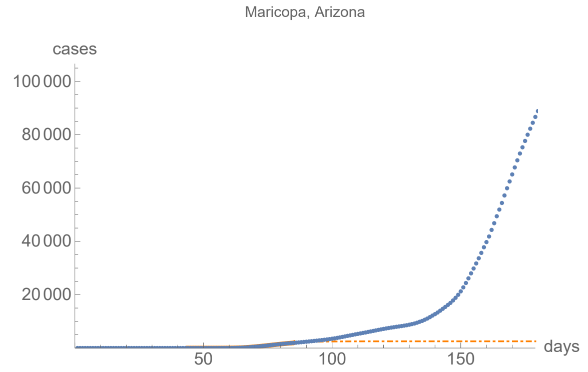


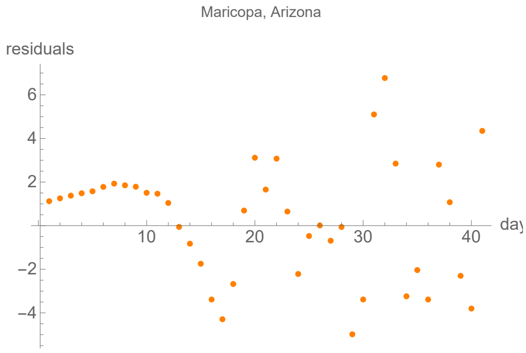


##### Parameters of fits to averaged death data.

The best fit ends on Thu 7 May 2020.

| α | β | k | τ | σ |
|---|---|---|---|---|
| 0.586±0.123 | 0.0991±0.00472 | 230.±12. | 35.3±1.14 | 9.94±0.587 |

| badness | | α | β | k | τ | σ | quality |
|---|---|---|---|---|---|---|---|
| 2.8% | error | 21.% | 4.8% | 5.2% | 3.2% | 5.9% | 42.8% |

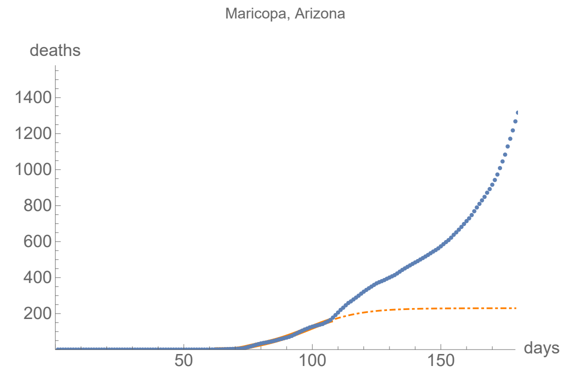


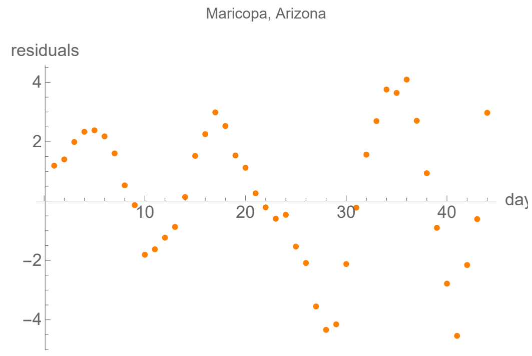


### Los Angeles, California

```
lag from integral overlap, using data averaged over 7 days, between -3 and 28 days, maximum overlap on day 4

lag from fits to the function I (averaged data): 21.5503 days
```

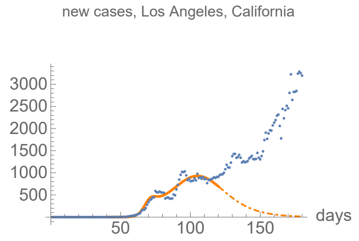


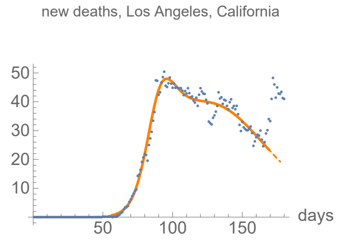


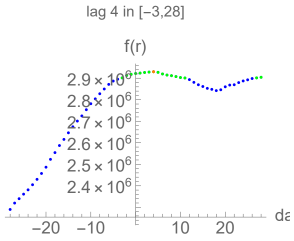


#### Parameters of fits to averaged cases data.

The best fit ends on Wed 20 May 2020.

| $\alpha$ | $\beta$ | k | $\tau$ | $\sigma$ |
|---|---|---|---|---|
| 0.396±0.0349 | 0.0744±0.0012 | 50000.±772. | 63.1±0.465 | 27.5±0.379 |

| badness | | $\alpha$ | $\beta$ | k | $\tau$ | $\sigma$ | quality |
|---|---|---|---|---|---|---|---|
| 1.7% | error | 8.8% | 1.6% | 1.5% | 0.74% | 1.4% | 15.8% |

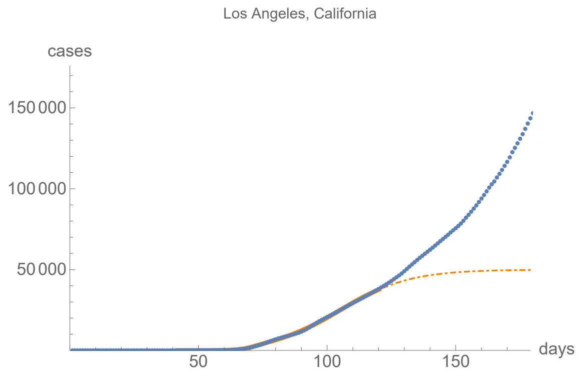


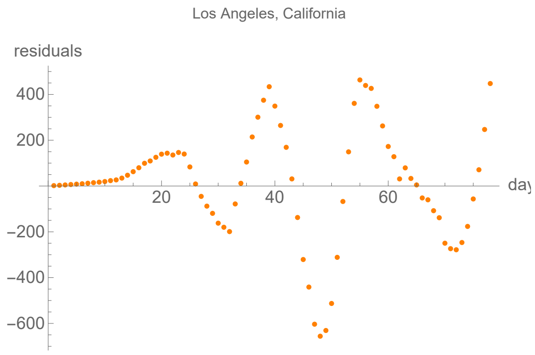


#### Parameters of fits to averaged death data.

The best fit ends on Wed 8 Jul 2020.

| $\alpha$ | $\beta$ | k | $\tau$ | $\sigma$ |
|---|---|---|---|---|
| 0.175±0.00184 | 0.036±0.000298 | 4300.±19.3 | 71.±0.248 | 30.9±0.136 |

| badness | | $\alpha$ | $\beta$ | k | $\tau$ | $\sigma$ | quality |
|---|---|---|---|---|---|---|---|
| 0.55% | error | 1.% | 0.83% | 0.45% | 0.35% | 0.44% | 3.67% |

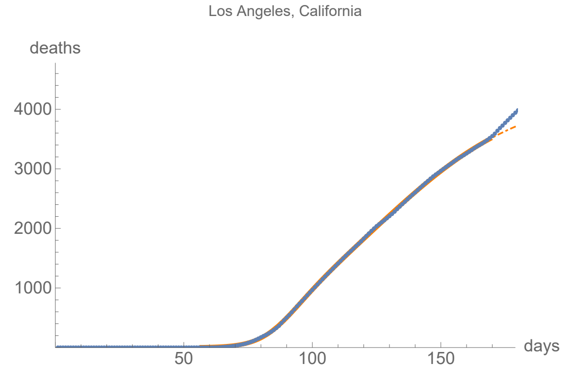


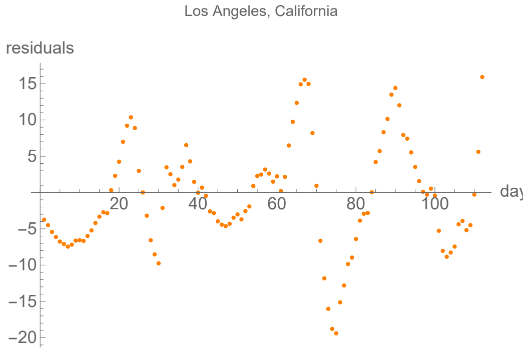


### Riverside, California

no clear lag in the first six months of the pandemic from integral overlap using data averaged over 7 days

lag from fits to the function I (averaged data): 11.8012 days

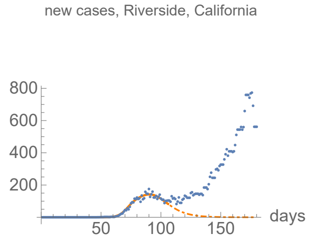


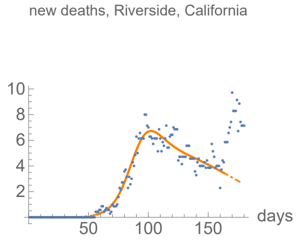


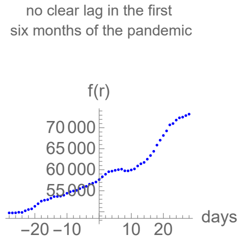


#### Parameters of fits to averaged cases data.

The best fit ends on Wed 6 May 2020.

| α | β | k | τ | σ |
|---|---|---|---|---|
| 0.322±0.0132 | 0.104±0.00271 | 5220.±62.5 | 42.8±0.191 | 23.2±0.444 |

| badness | | α | β | k | τ | σ | quality |
|---|---|---|---|---|---|---|---|
| 0.96% | error | 4.1% | 2.6% | 1.2% | 0.45% | 1.9% | 11.2% |

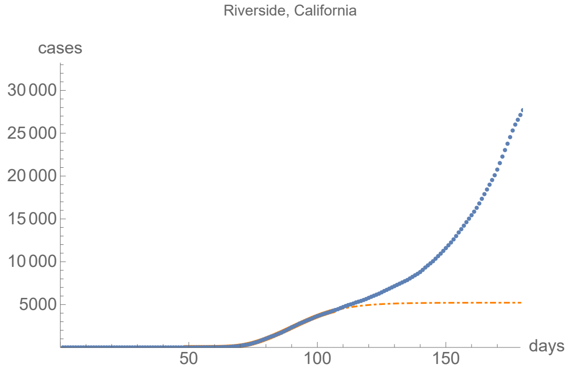


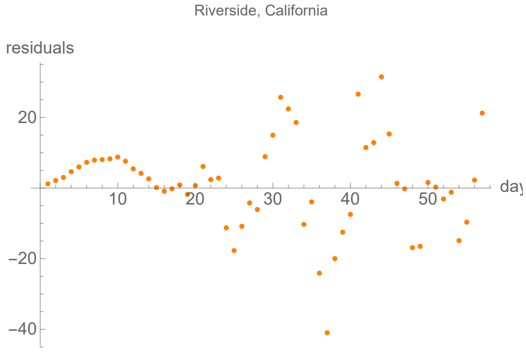


#### Parameters of fits to averaged death data.

The best fit ends on Thu 2 Jul 2020.

| α | β | k | τ | σ |
|---|---|---|---|---|
| 0.132±0.00275 | 0.0296±0.00318 | 603.±14.7 | 35.3±0.568 | 70.5±1.5 |

| badness | | α | β | k | τ | σ | quality |
|---|---|---|---|---|---|---|---|
| 0.96% | error | 2.1% | 11.% | 2.4% | 1.6% | 2.1% | 10.9% |

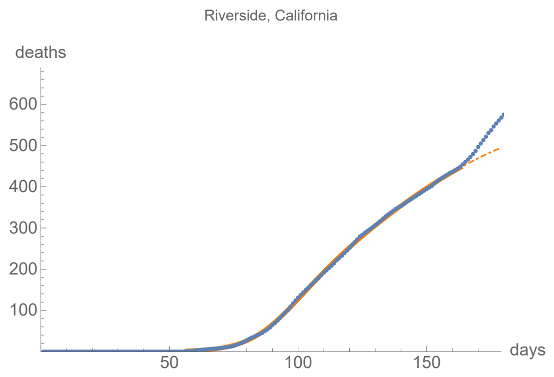


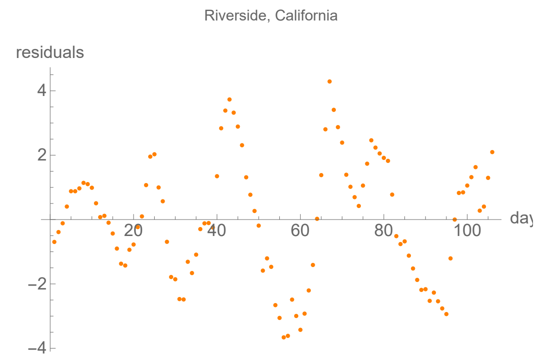


### Fairfield, Connecticut

lag from integral overlap, using data averaged over 7 days, between 4 and 12 days, maximum overlap on day 9

lag from fits to the function I (averaged data): 6.90919 days

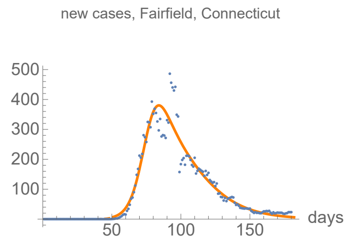


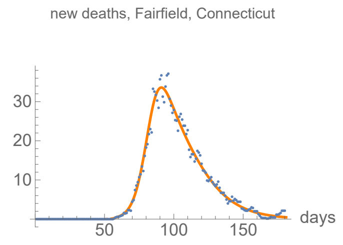


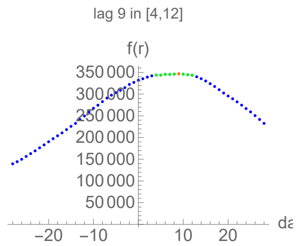


#### Parameters of fits to averaged cases data.

The best fit ends on Tue 21 Jul 2020.

| α | β | k | τ | σ |
|---|---|---|---|---|
| 0.202±0.00548 | 0.0536±0.00126 | 17000.±35.1 | 38.2±0.537 | 26.6±0.447 |

| badness | | α | β | k | τ | σ | quality |
|---|---|---|---|---|---|---|---|
| 2.5% | error | 2.7% | 2.3% | 0.21% | 1.4% | 1.7% | 10.8% |

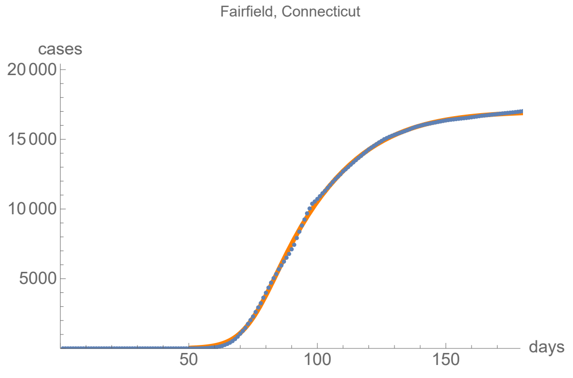


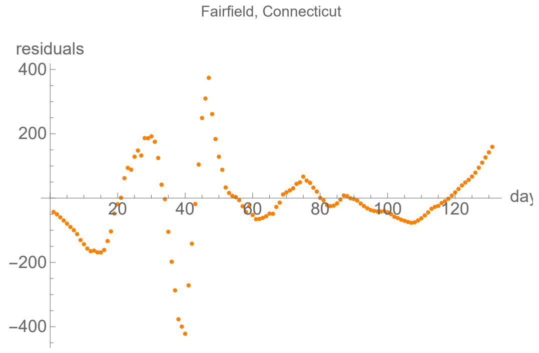


#### Parameters of fits to averaged death data.

The best fit ends on Mon 20 Jul 2020.

| α | β | k | τ | σ |
|---|---|---|---|---|
| 0.219±0.00177 | 0.0609±0.000403 | 1400.±0.759 | 37.6±0.141 | 26.3±0.128 |

| badness | | α | β | k | τ | σ | quality |
|---|---|---|---|---|---|---|---|
| 0.46% | error | 0.81% | 0.66% | 0.054% | 0.37% | 0.49% | 2.84% |

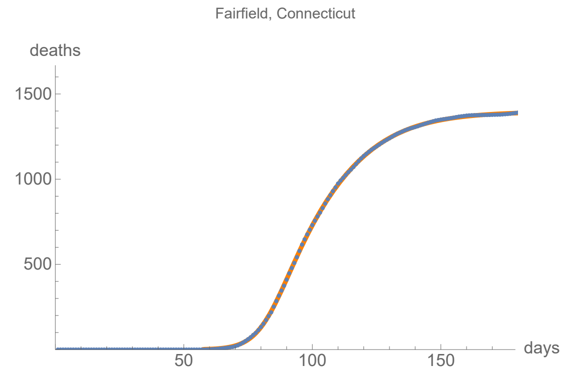


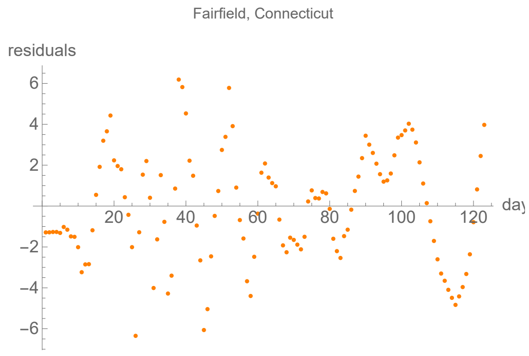


### Hartford, Connecticut

lag from integral overlap, using data averaged over 7 days, between 0 and 5 days, maximum overlap on day 3

lag from fits to the function I (averaged data): 4.14423 days

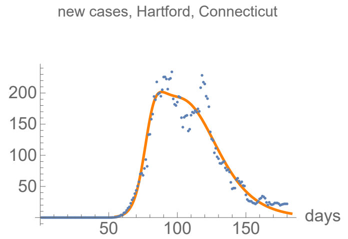


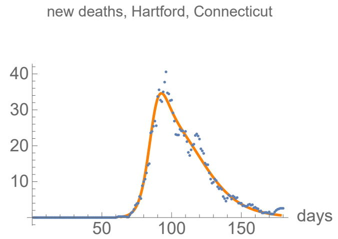


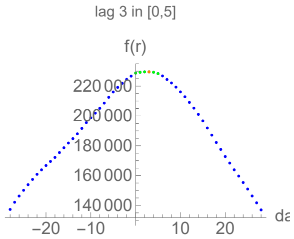


#### Parameters of fits to averaged cases data.

The best fit ends on Wed 22 Jul 2020.

| α | β | k | τ | σ |
|---|---|---|---|---|
| 0.25±0.0107 | 0.0605±0.000763 | 12000.±23.6 | 50.±0.152 | 24.6±0.348 |

| badness | | α | β | k | τ | σ | quality |
|---|---|---|---|---|---|---|---|
| 1.7% | error | 4.3% | 1.3% | 0.2% | 0.3% | 1.4% | 9.13% |

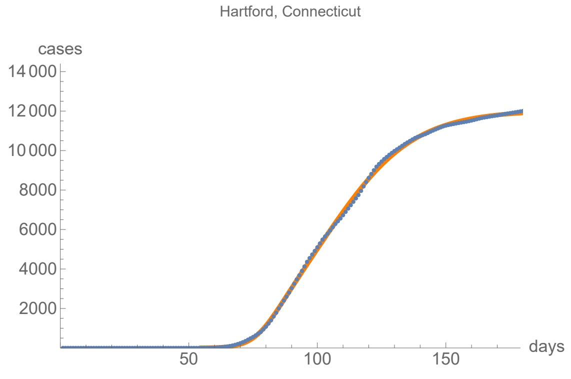


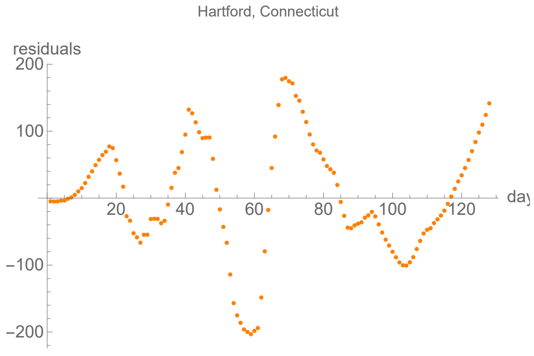


#### Parameters of fits to averaged death data.

The best fit ends on Fri 17 Jul 2020.

| α | β | k | τ | σ |
|---|---|---|---|---|
| 0.274±0.00561 | 0.0652±0.000705 | 1400.±1.77 | 38.4±0.149 | 23.6±0.178 |

| badness | | α | β | k | τ | σ | quality |
|---|---|---|---|---|---|---|---|
| 1.1% | error | 2.% | 1.1% | 0.13% | 0.39% | 0.75% | 5.51% |

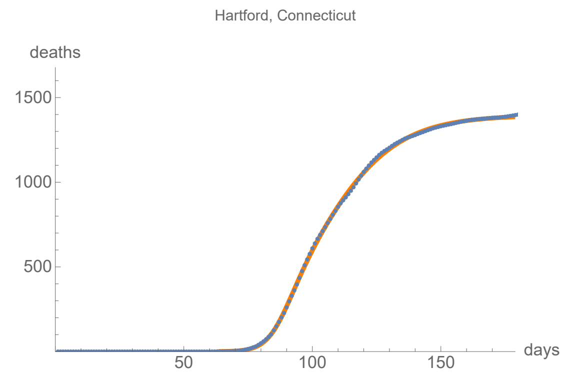


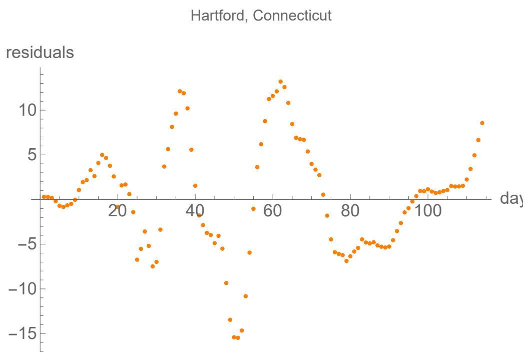


### New Haven, Connecticut

lag from integral overlap, using data averaged over 7 days, between 4 and 9 days, maximum overlap on day 6

lag from fits to the function I (averaged data): 5.79625 days

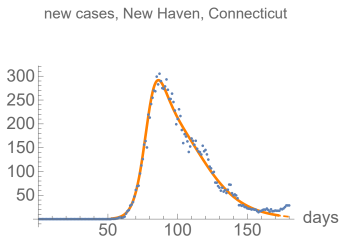


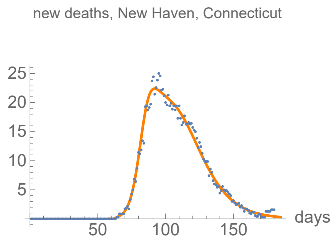


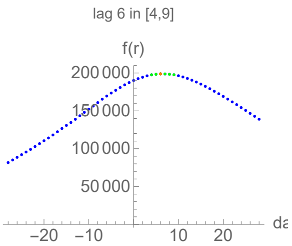


#### Parameters of fits to averaged cases data.

The best fit ends on Sat 11 Jul 2020.

| α | β | k | τ | σ |
|---|---|---|---|---|
| 0.247±0.00281 | 0.0593±0.000413 | 12600.±10.8 | 42.8±0.103 | 27.7±0.117 |

| badness | | α | β | k | τ | σ | quality |
|---|---|---|---|---|---|---|---|
| 0.63% | error | 1.1% | 0.7% | 0.086% | 0.24% | 0.42% | 3.21% |

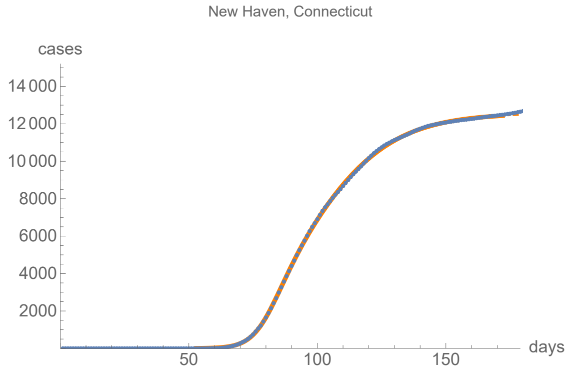


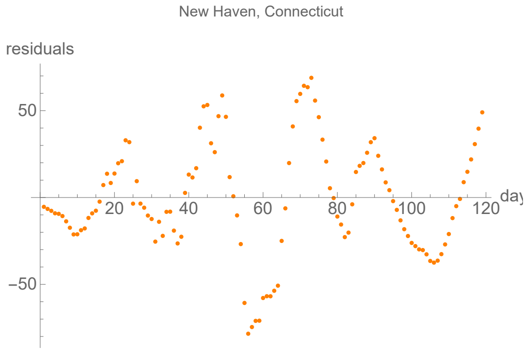


#### Parameters of fits to averaged death data.

The best fit ends on Fri 24 Jul 2020.

| α | β | k | τ | σ |
|---|---|---|---|---|
| 0.269±0.00375 | 0.0683±0.000323 | 1090.±0.617 | 39.2±0.0612 | 19.±0.113 |

| badness | | α | β | k | τ | σ | quality |
|---|---|---|---|---|---|---|---|
| 0.55% | error | 1.4% | 0.47% | 0.056% | 0.16% | 0.59% | 3.22% |

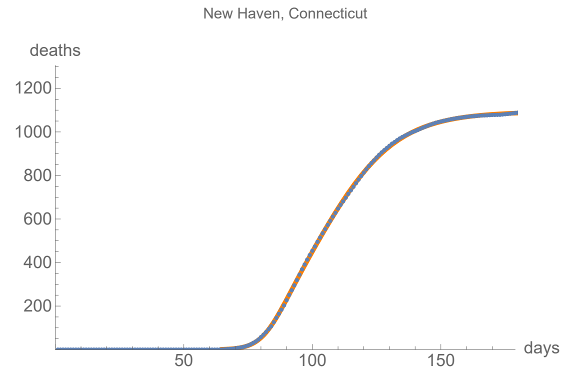


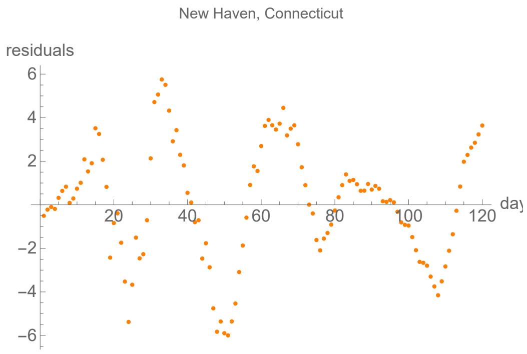


### District of Columbia, District of Columbia

lag from integral overlap, using data averaged over 7 days, between 1 and 8 days, maximum overlap on day 4

lag from fits to the function I (averaged data): -4.96504 days

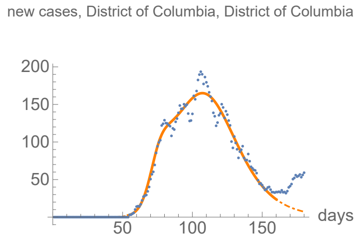


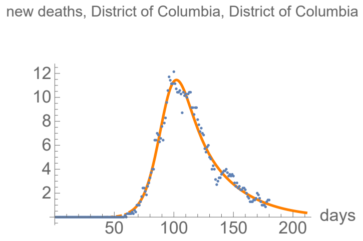


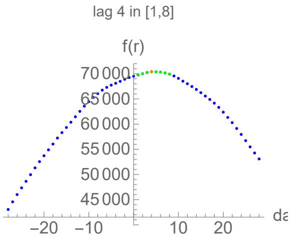


#### Parameters of fits to averaged cases data.

The best fit ends on Mon 29 Jun 2020.

| α | β | k | τ | σ |
|---|---|---|---|---|
| 0.25±0.00712 | 0.0622±0.000359 | 10500.±17.1 | 53.8±0.0657 | 19.3±0.22 |

| badness | | α | β | k | τ | σ | quality |
|---|---|---|---|---|---|---|---|
| 0.88% | error | 2.8% | 0.58% | 0.16% | 0.12% | 1.1% | 5.73% |

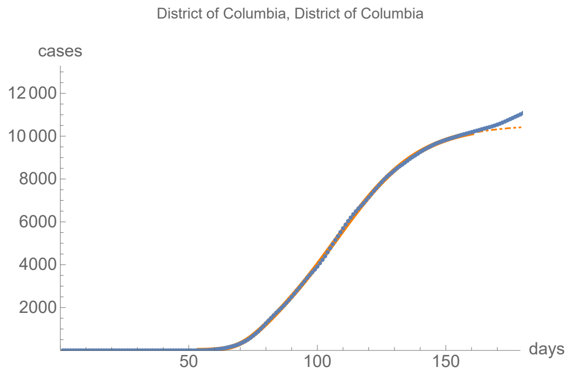


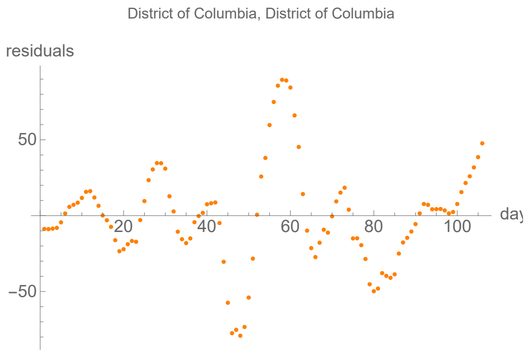


#### Parameters of fits to averaged death data.

The best fit ends on Wed 19 Aug 2020.

| α | β | k | τ | σ |
|---|---|---|---|---|
| 0.146±0.00156 | 0.0366±0.00187 | 604.±0.981 | 33.3±0.28 | 36.5±0.593 |

| badness | | α | β | k | τ | σ | quality |
|---|---|---|---|---|---|---|---|
| 0.78% | error | 1.1% | 5.1% | 0.16% | 0.84% | 1.6% | 5.47% |

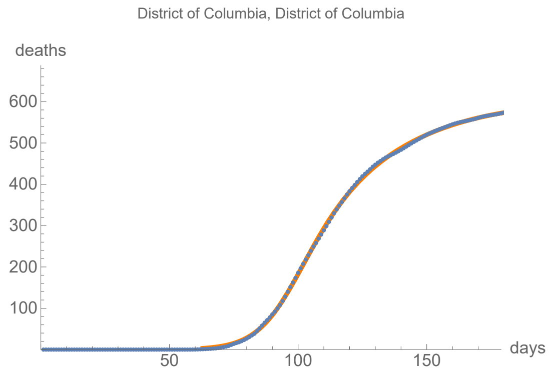


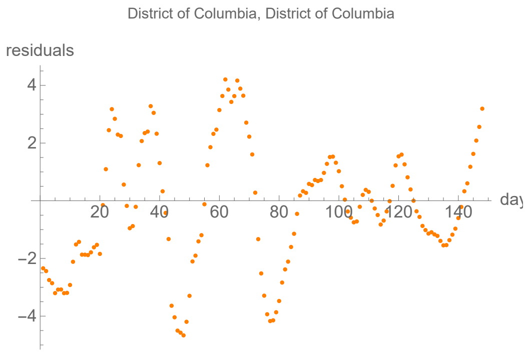


### Miami–Dade, Florida

no clear lag in the first six months of the pandemic from integral overlap using data averaged over 7 days

lag from fits to the function I (averaged data): 12.7839 days

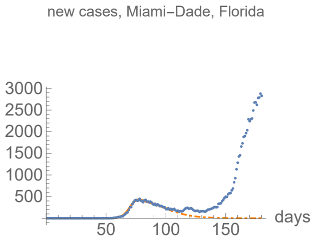


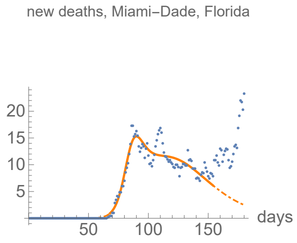


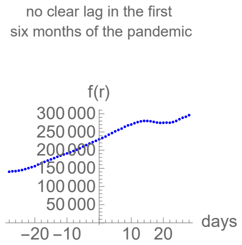


#### Parameters of fits to averaged cases data.

The best fit ends on Tue 5 May 2020.

| α | β | k | τ | σ |
|---|---|---|---|---|
| 0.344±0.00441 | 0.0861±0.00551 | 15000.±91.4 | 20.1±0.126 | 33.9±0.109 |

| badness | | α | β | k | τ | σ | quality |
|---|---|---|---|---|---|---|---|
| 0.44% | error | 1.3% | 6.4% | 0.61% | 0.63% | 0.32% | 4.59% |

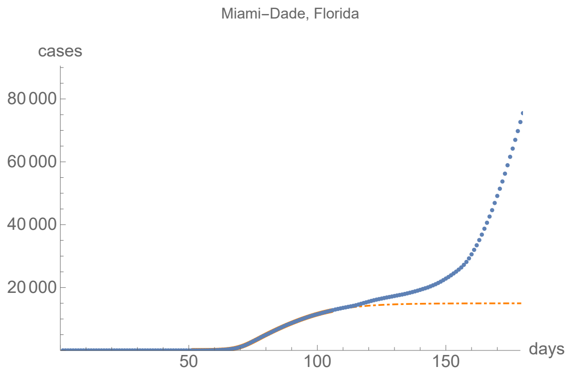


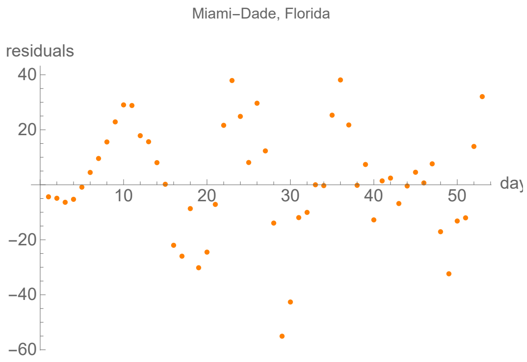


#### Parameters of fits to averaged death data.

The best fit ends on Tue 23 Jun 2020.

| α | β | k | τ | σ |
|---|---|---|---|---|
| 0.231±0.00848 | 0.0438±0.00113 | 1030.±13.4 | 48.5±0.594 | 17.1±0.325 |

| badness | | α | β | k | τ | σ | quality |
|---|---|---|---|---|---|---|---|
| 1.5% | error | 3.7% | 2.6% | 1.3% | 1.2% | 1.9% | 12.2% |

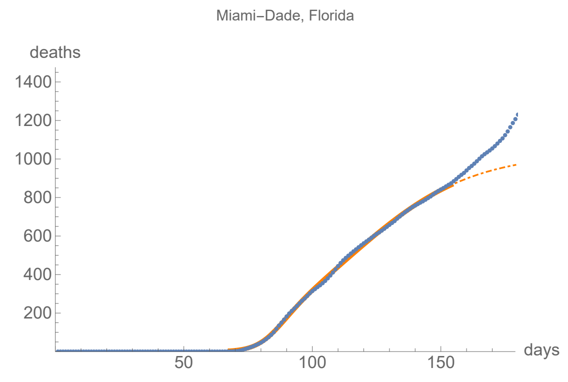


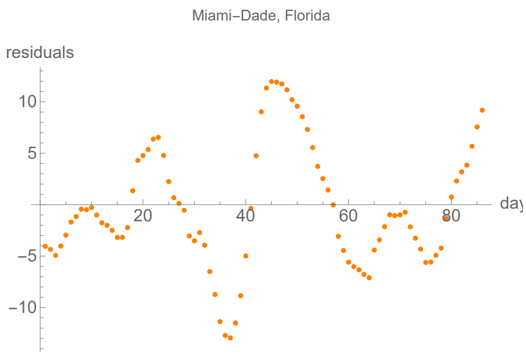


### Palm Beach, Florida

no clear lag in the first six months of the pandemic from integral overlap using data averaged over 7 days

lag from fits to the function I (averaged data): 2.79552 days

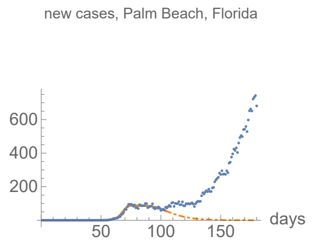


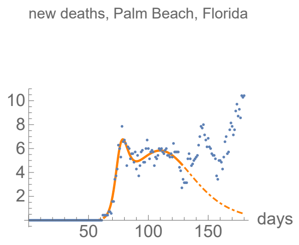


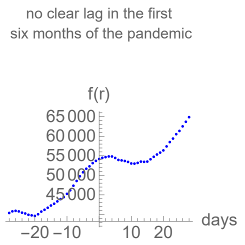


#### Parameters of fits to averaged cases data.

The best fit ends on Mon 4 May 2020.

| α | β | k | τ | σ |
|---|---|---|---|---|
| 0.372±0.0117 | 0.0865±0.00214 | 3850.±55. | 35.9±0.328 | 17.1±0.216 |

| badness | | α | β | k | τ | σ | quality |
|---|---|---|---|---|---|---|---|
| 0.71% | error | 3.1% | 2.5% | 1.4% | 0.91% | 1.3% | 9.95% |

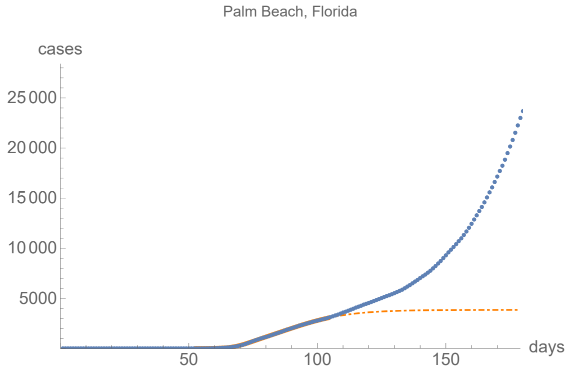


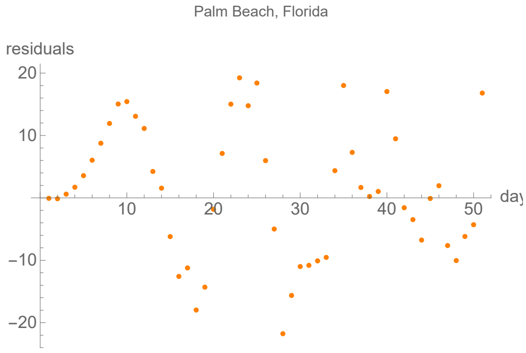


#### Parameters of fits to averaged death data.

The best fit ends on Thu 28 May 2020.

| α | β | k | τ | σ |
|---|---|---|---|---|
| 0.352±0.00711 | 0.053±0.000551 | 440.±4.13 | 46.9±0.382 | 13.2±0.0959 |

| badness | | α | β | k | τ | σ | quality |
|---|---|---|---|---|---|---|---|
| 0.6% | error | 2.% | 1.% | 0.94% | 0.82% | 0.72% | 6.14% |

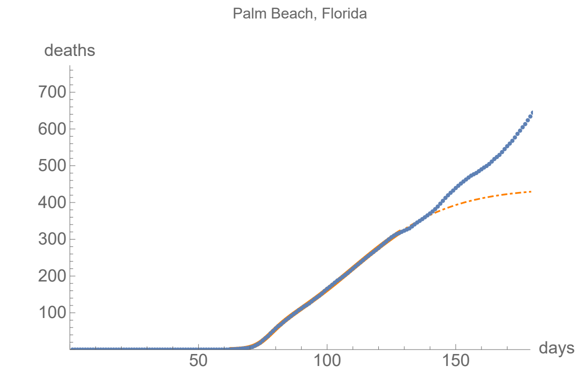


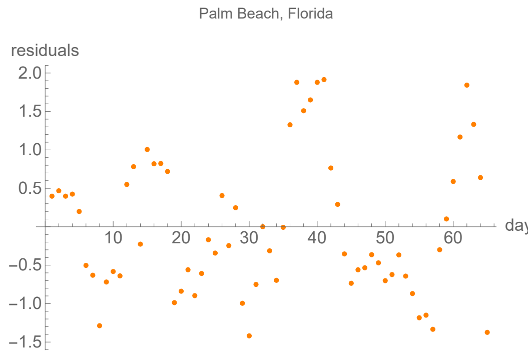


### Cook, Illinois

lag from integral overlap, using data averaged over 7 days, between 2 and 10 days, maximum overlap on day 6

lag from fits to the function I (averaged data): 24.7331 days

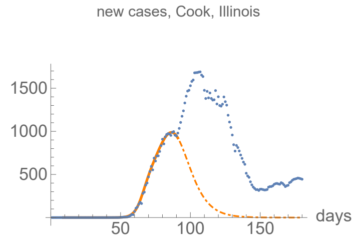


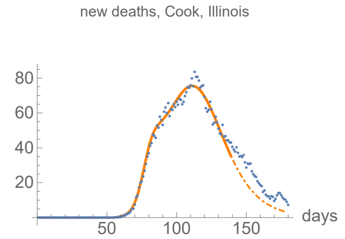


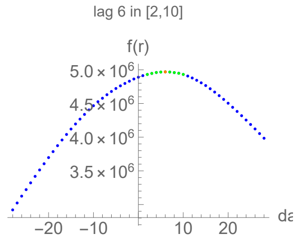


#### Parameters of fits to averaged cases data.

The best fit ends on Tue 21 Apr 2020.

| α | β | k | τ | σ |
|---|---|---|---|---|
| 0.364±0.00482 | 0.118±0.00122 | 32800.±311. | 77.9±0.156 | 57.4±0.139 |

| badness | | α | β | k | τ | σ | quality |
|---|---|---|---|---|---|---|---|
| 0.22% | error | 1.3% | 1.% | 0.95% | 0.2% | 0.24% | 3.97% |

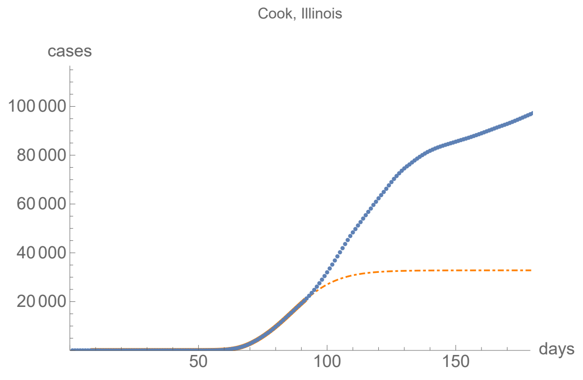


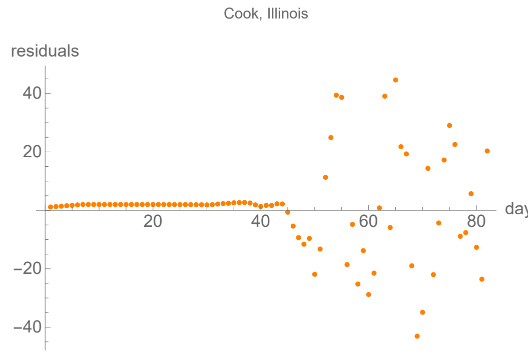


#### Parameters of fits to averaged death data.

The best fit ends on Mon 8 Jun 2020.

| α | β | k | τ | σ |
|---|---|---|---|---|
| 0.275±0.00601 | 0.0674±0.000436 | 4460.±14.7 | 52.6±0.107 | 19.8±0.156 |

| badness | | α | β | k | τ | σ | quality |
|---|---|---|---|---|---|---|---|
| 0.54% | error | 2.2% | 0.65% | 0.33% | 0.2% | 0.79% | 4.69% |

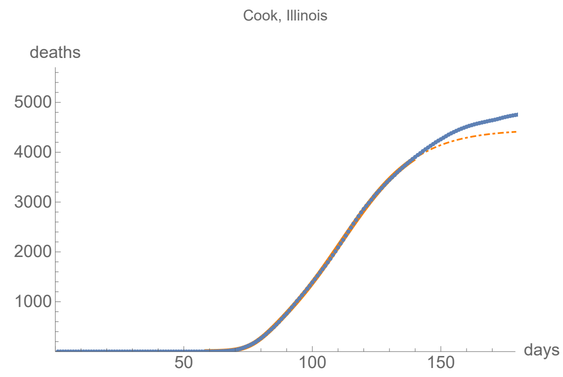


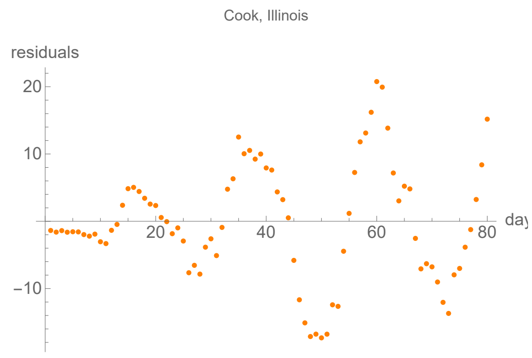


### Marion, Indiana

lag from integral overlap, using data averaged over 7 days, between 0 and 12 days, maximum overlap on day 9

lag from fits to the function I (averaged data): 16.7342 days

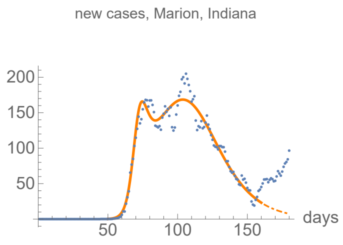


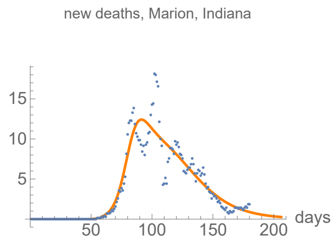


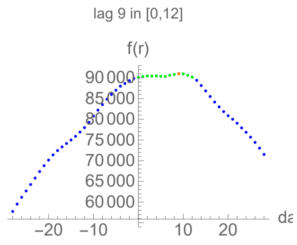


#### Parameters of fits to averaged cases data.

The best fit ends on Mon 29 Jun 2020.

| $\alpha$ | $\beta$ | k | $\tau$ | $\sigma$ |
|---|---|---|---|---|
| 0.347±0.0103 | 0.0587±0.000287 | 11500.±17.7 | 53.8±0.0722 | 21.5±0.141 |

| badness | | $\alpha$ | $\beta$ | k | $\tau$ | $\sigma$ | quality |
|---|---|---|---|---|---|---|---|
| 1.1% | error | 3.% | 0.49% | 0.15% | 0.13% | 0.66% | 5.45% |

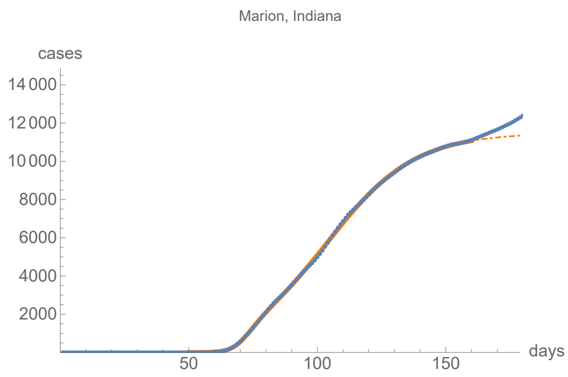


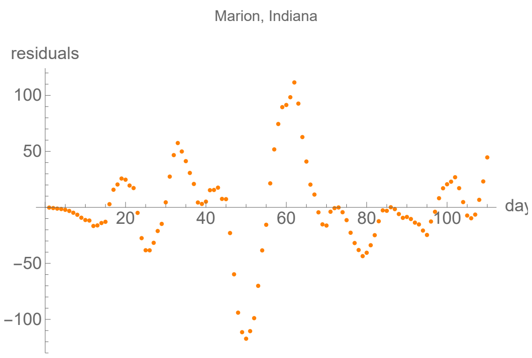


#### Parameters of fits to averaged death data.

The best fit ends on Fri 14 Aug 2020.

| $\alpha$ | $\beta$ | k | $\tau$ | $\sigma$ |
|---|---|---|---|---|
| 0.198±0.00795 | 0.0488±0.000938 | 724.±1.69 | 46.6±0.347 | 24.3±0.479 |

| badness | | $\alpha$ | $\beta$ | k | $\tau$ | $\sigma$ | quality |
|---|---|---|---|---|---|---|---|
| 2.6% | error | 4.% | 1.9% | 0.23% | 0.74% | 2.% | 11.5% |

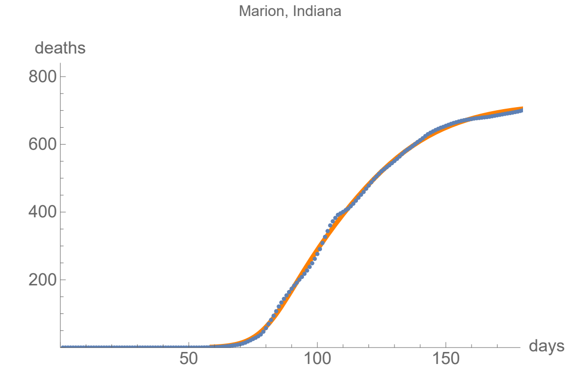


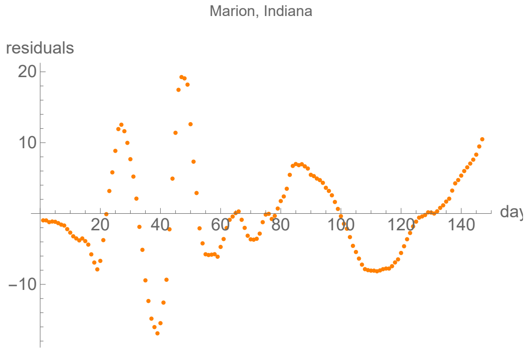


### Orleans, Louisiana

lag from integral overlap, using data averaged over 7 days, between 2 and 11 days, maximum overlap on day 7

lag from fits to the function I (averaged data): 6.7595 days

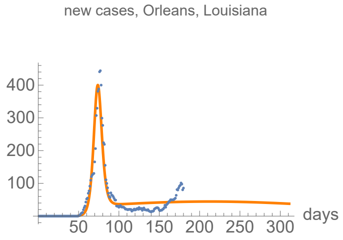


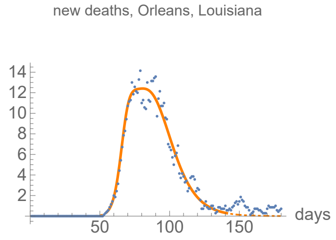


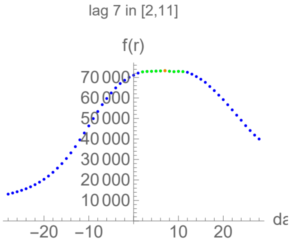


#### Parameters of fits to averaged cases data.

The best fit ends on Fri 27 Nov 2020.

| α | β | k | τ | σ |
|---|---|---|---|---|
| 0.309±0.0238 | 0.00834±0.000299 | 21300.±758. | 163.±9.07 | 23.6±0.304 |

| badness | | α | β | k | τ | σ | quality |
|---|---|---|---|---|---|---|---|
| 4.5% | error | 7.7% | 3.6% | 3.6% | 5.6% | 1.3% | 26.3% |

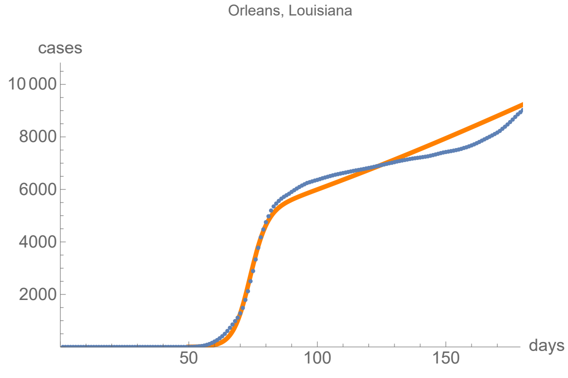


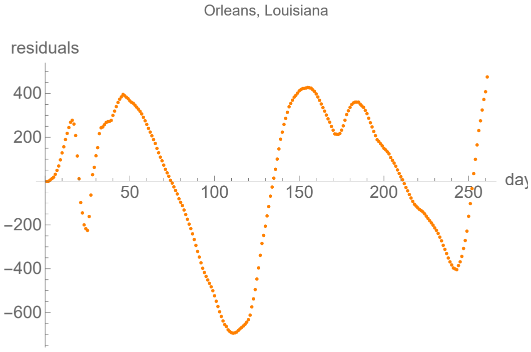


#### Parameters of fits to averaged death data.

The best fit ends on Wed 10 Jun 2020.

| α | β | k | τ | σ |
|---|---|---|---|---|
| 0.349±0.011 | 0.0912±0.000797 | 513.±0.627 | 30.3±0.0763 | 12.3±0.188 |

| badness | | α | β | k | τ | σ | quality |
|---|---|---|---|---|---|---|---|
| 1.1% | error | 3.1% | 0.87% | 0.12% | 0.25% | 1.5% | 7.01% |

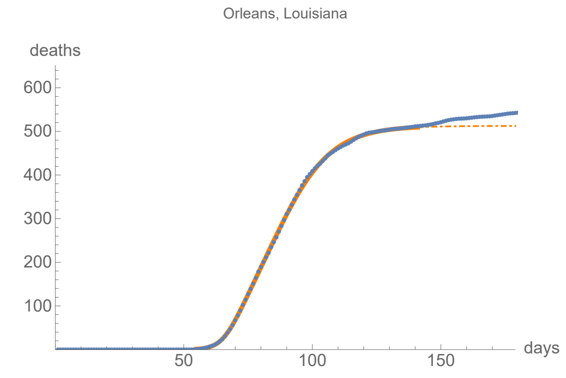


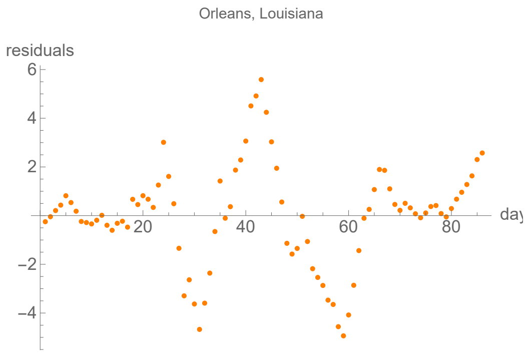


### Baltimore, Maryland

lag from integral overlap, using data averaged over 7 days, between 1 and 19 days, maximum overlap on day 14

lag from fits to the function I (averaged data): 29.6195 days

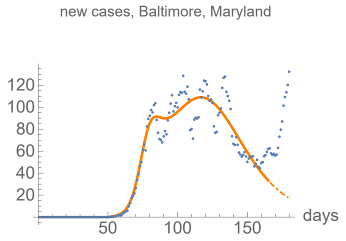


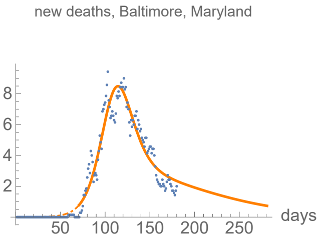


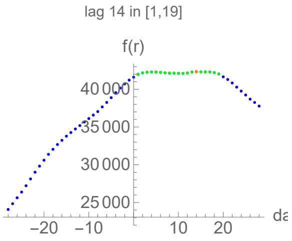


#### Parameters of fits to averaged cases data.

The best fit ends on Sat 4 Jul 2020.

| $\alpha$ | $\beta$ | k | $\tau$ | $\sigma$ |
|---|---|---|---|---|
| 0.245±0.00963 | 0.05±0.000473 | 8690.±34.4 | 64.1±0.184 | 24.2±0.286 |

| badness | | $\alpha$ | $\beta$ | k | $\tau$ | $\sigma$ | quality |
|---|---|---|---|---|---|---|---|
| 1.3% | error | 3.9% | 0.95% | 0.4% | 0.29% | 1.2% | 8.06% |

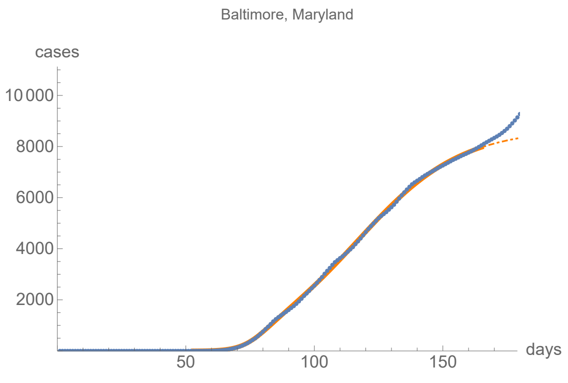


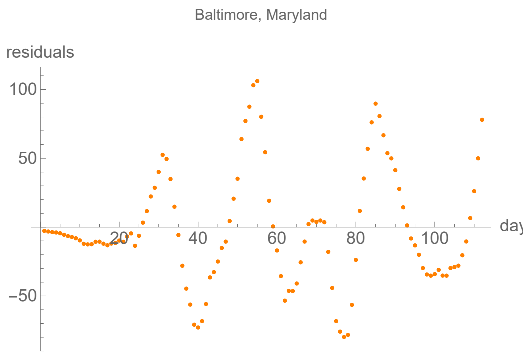


#### Parameters of fits to averaged death data.

The best fit ends on Wed 28 Oct 2020.

| $\alpha$ | $\beta$ | k | $\tau$ | $\sigma$ |
|---|---|---|---|---|
| 0.0993±0.00145 | 0.0159±0.00184 | 705.±4.11 | 38.5±0.343 | 47.7±0.692 |

| badness | | $\alpha$ | $\beta$ | k | $\tau$ | $\sigma$ | quality |
|---|---|---|---|---|---|---|---|
| 1.5% | error | 1.5% | 12.% | 0.58% | 0.89% | 1.5% | 7.26% |

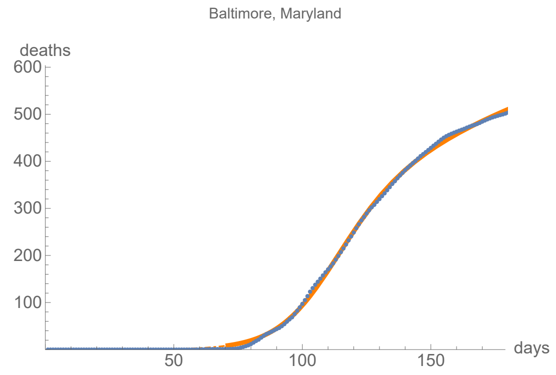


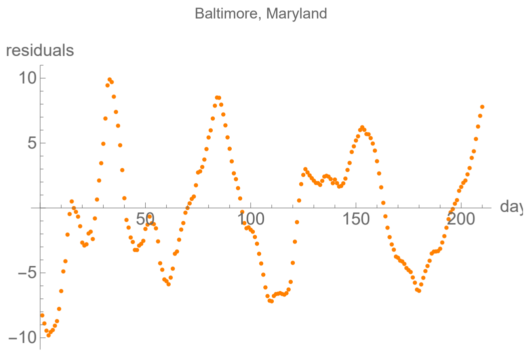


### Montgomery, Maryland

lag from integral overlap, using data averaged over 7 days, between -7 and 0 days, maximum overlap on day -4

lag from fits to the function I (averaged data): -15.057 days

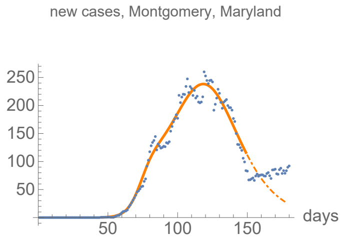


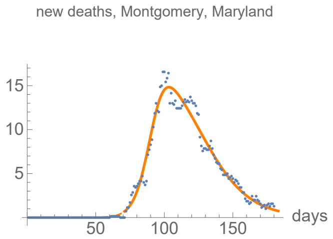


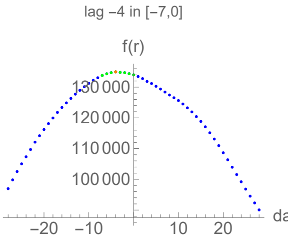


#### Parameters of fits to averaged cases data.

The best fit ends on Thu 18 Jun 2020.

| α | β | k | τ | σ |
|---|---|---|---|---|
| 0.217±0.00739 | 0.0592±0.00049 | 16000.±70.4 | 72.7±0.162 | 29.6±0.319 |

| badness | | α | β | k | τ | σ | quality |
|---|---|---|---|---|---|---|---|
| 0.73% | error | 3.4% | 0.83% | 0.44% | 0.22% | 1.1% | 6.71% |

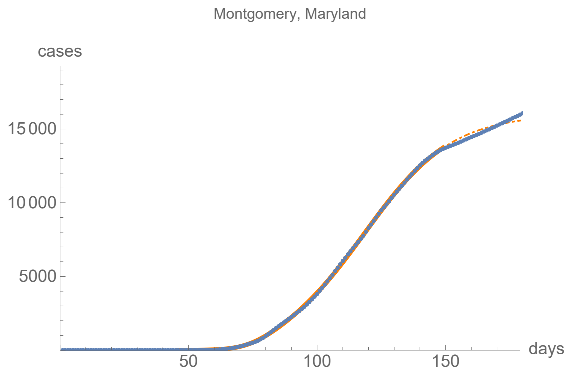


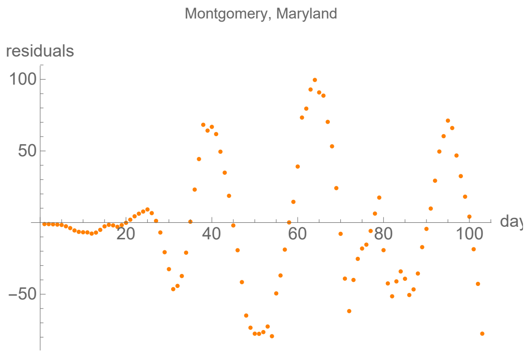


#### Parameters of fits to averaged death data.

The best fit ends on Wed 22 Jul 2020.

| α | β | k | τ | σ |
|---|---|---|---|---|
| 0.19±0.00384 | 0.0558±0.000825 | 779.±1.39 | 39.8±0.268 | 21.6±0.373 |

| badness | | α | β | k | τ | σ | quality |
|---|---|---|---|---|---|---|---|
| 0.98% | error | 2.% | 1.5% | 0.18% | 0.67% | 1.7% | 7.07% |

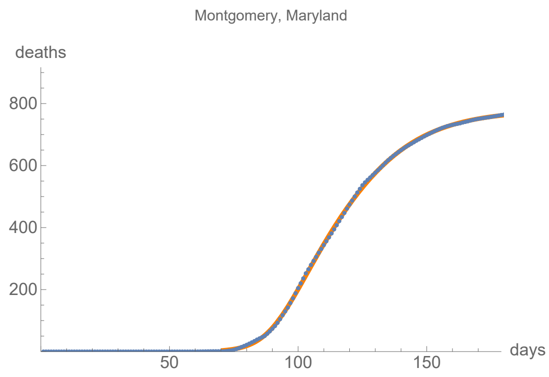


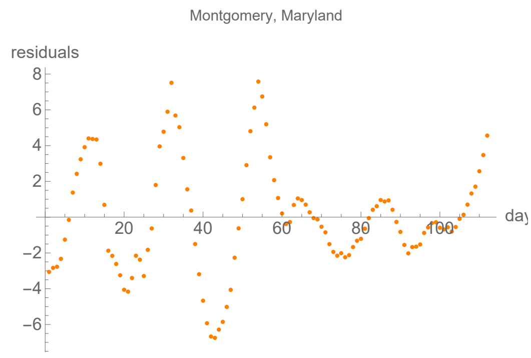


### Prince George's, Maryland

lag from integral overlap, using data averaged over 7 days, between -4 and 4 days, maximum overlap on day -1

lag from fits to the function I (averaged data): -10.4462 days

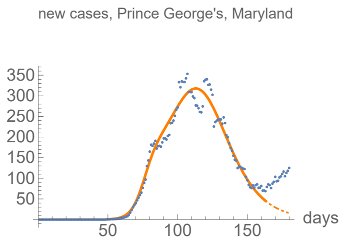


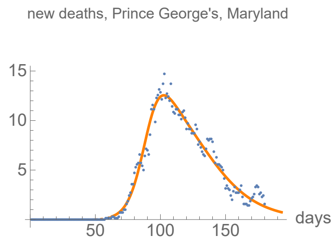


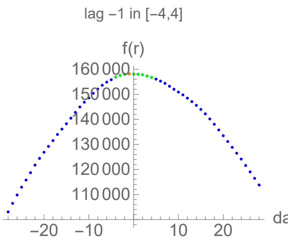


#### Parameters of fits to averaged cases data.

The best fit ends on Thu 2 Jul 2020.

| α | β | k | τ | σ |
|---|---|---|---|---|
| 0.23±0.00836 | 0.0658±0.000471 | 19100.±36.5 | 63.7±0.0725 | 27.7±0.327 |

| badness | | α | β | k | τ | σ | quality |
|---|---|---|---|---|---|---|---|
| 0.9% | error | 3.6% | 0.72% | 0.19% | 0.11% | 1.2% | 6.74% |

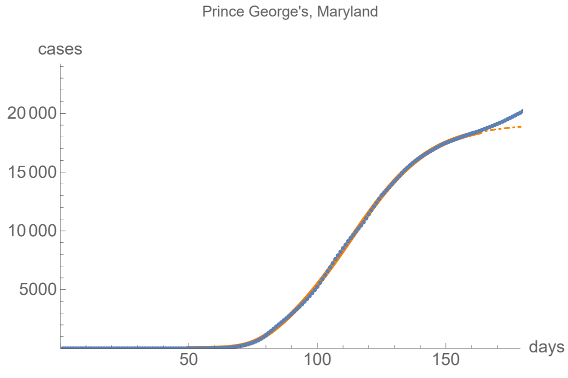


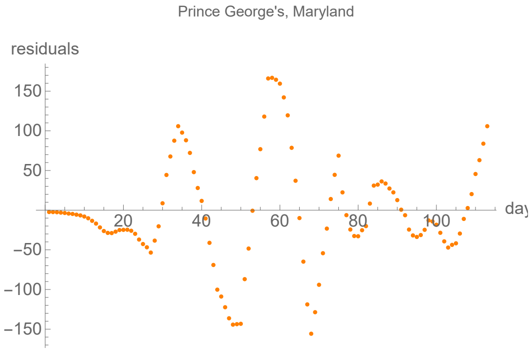


#### Parameters of fits to averaged death data.

The best fit ends on Sat 1 Aug 2020.

| α | β | k | τ | σ |
|---|---|---|---|---|
| 0.167±0.00339 | 0.047±0.000759 | 745.±1.59 | 48.8±0.314 | 29.±0.411 |

| badness | | α | β | k | τ | σ | quality |
|---|---|---|---|---|---|---|---|
| 1.% | error | 2.% | 1.6% | 0.21% | 0.64% | 1.4% | 6.95% |

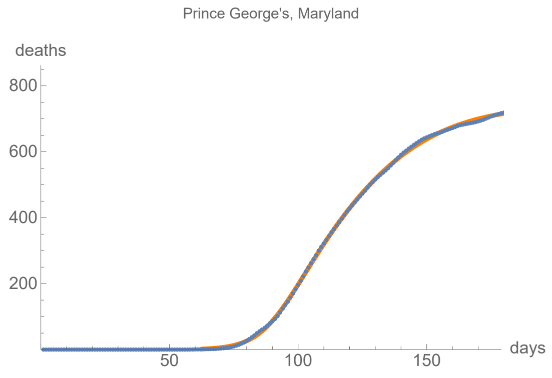


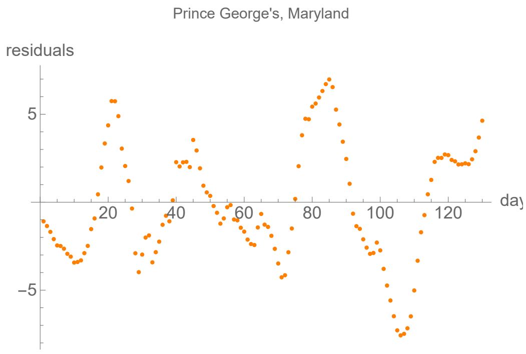


### Bristol, Massachusetts

lag from integral overlap, using data averaged over 7 days, between 9 and 19 days, maximum overlap on day 14

lag from fits to the function I (averaged data): −0.459238 days

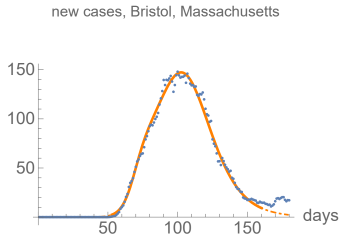


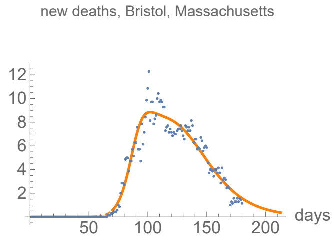


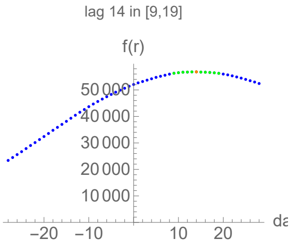


#### Parameters of fits to averaged cases data.

The best fit ends on Mon 29 Jun 2020.

| α | β | k | τ | σ |
|---|---|---|---|---|
| 0.24±0.0052 | 0.0722±0.000278 | 8040.±6.09 | 50.2±0.0372 | 17.2±0.19 |

| badness | | α | β | k | τ | σ | quality |
|---|---|---|---|---|---|---|---|
| 0.51% | error | 2.2% | 0.39% | 0.076% | 0.074% | 1.1% | 4.32% |

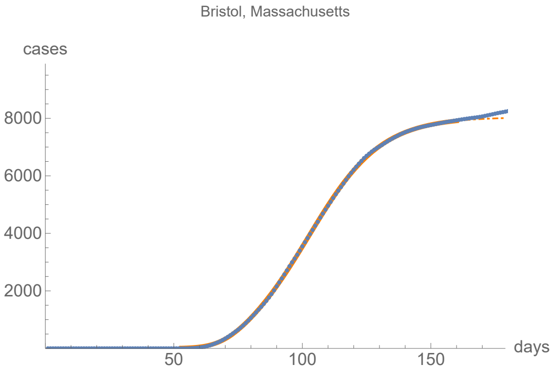


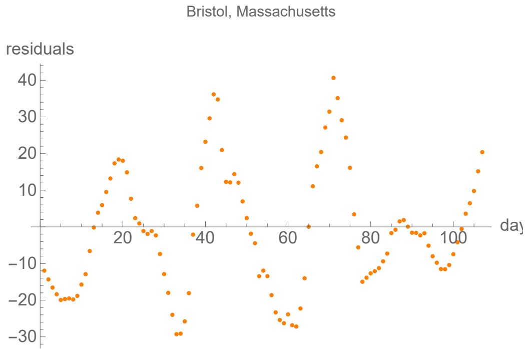


#### Parameters of fits to averaged death data.

The best fit ends on Fri 21 Aug 2020.

| α | β | k | τ | σ |
|---|---|---|---|---|
| 0.188±0.00494 | 0.0477±0.000444 | 643.±0.918 | 50.1±0.143 | 19.6±0.303 |

| badness | | α | β | k | τ | σ | quality |
|---|---|---|---|---|---|---|---|
| 1.1% | error | 2.6% | 0.93% | 0.14% | 0.29% | 1.5% | 6.65% |

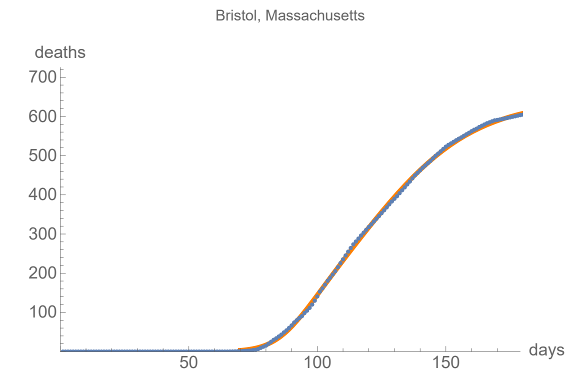


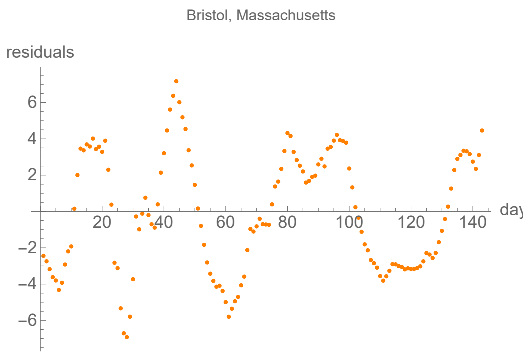


### Essex, Massachusetts

```
lag from integral overlap, using data averaged over 7 days, between 9 and 15 days, maximum overlap on day 11

lag from fits to the function I (averaged data): 11.5304 days
```

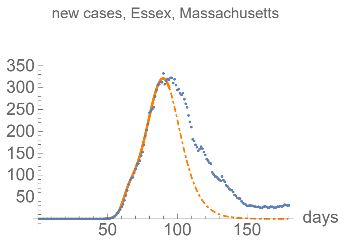


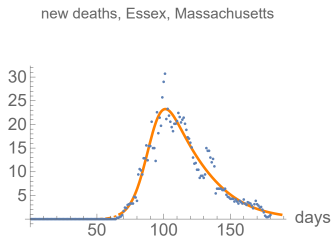


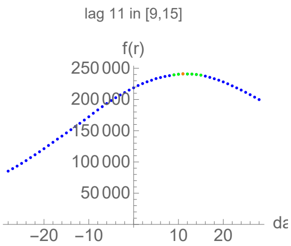


#### Parameters of fits to averaged cases data.

The best fit ends on Fri 24 Apr 2020.

| α | β | k | τ | σ |
|---|---|---|---|---|
| 0.442±0.0094 | 0.119±0.000556 | 10800.±65.5 | 40.8±0.105 | 13.3±0.0945 |

| badness | | α | β | k | τ | σ | quality |
|---|---|---|---|---|---|---|---|
| 0.25% | error | 2.1% | 0.47% | 0.61% | 0.26% | 0.71% | 4.42% |

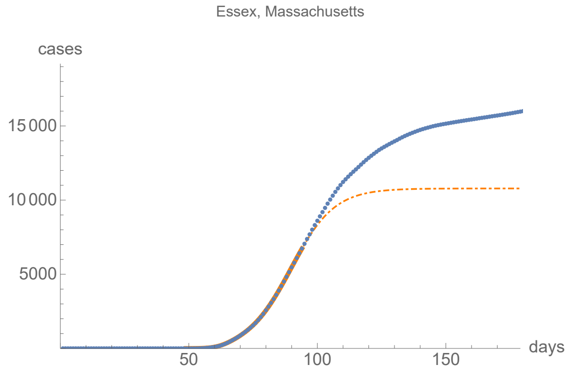


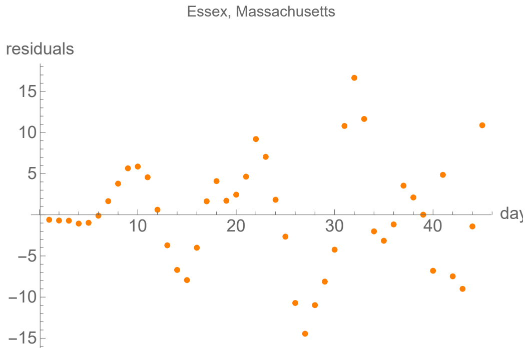


#### Parameters of fits to averaged death data.

The best fit ends on Mon 27 Jul 2020.

| α | β | k | τ | σ |
|---|---|---|---|---|
| 0.171±0.00329 | 0.0491±0.00407 | 1170.±2.57 | 26.1±0.471 | 37.7±0.541 |

| badness | | α | β | k | τ | σ | quality |
|---|---|---|---|---|---|---|---|
| 1.3% | error | 1.9% | 8.3% | 0.22% | 1.8% | 1.4% | 8.75% |

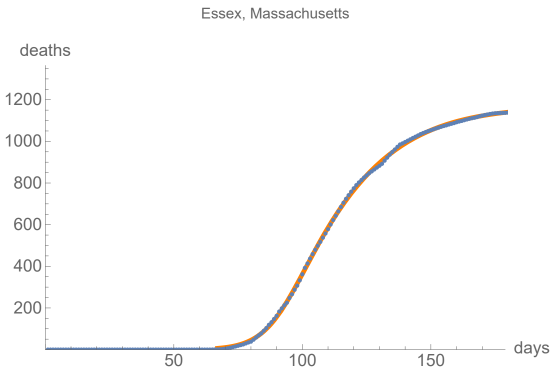


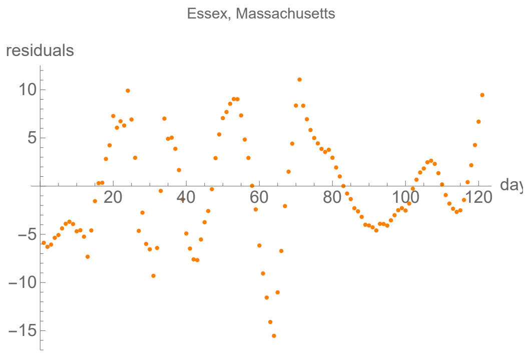


### Hampden, Massachusetts

lag from integral overlap, using data averaged over 7 days, between 6 and 12 days, maximum overlap on day 10

lag from fits to the function I (averaged data): 0.825176 days

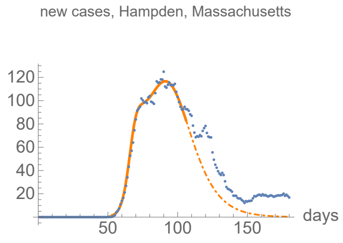


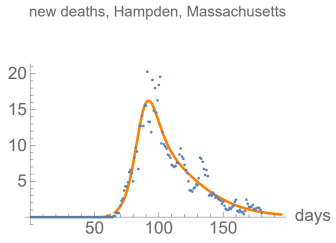


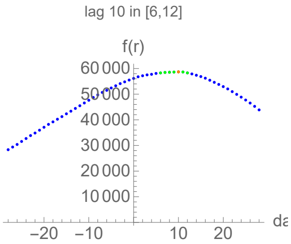


#### Parameters of fits to averaged cases data.

The best fit ends on Wed 6 May 2020.

| α | β | k | τ | σ |
|---|---|---|---|---|
| 0.355±0.00445 | 0.0825±0.000553 | 5610.±26.2 | 38.7±0.119 | 14.6±0.0771 |

| badness | | α | β | k | τ | σ | quality |
|---|---|---|---|---|---|---|---|
| 0.35% | error | 1.3% | 0.67% | 0.47% | 0.31% | 0.53% | 3.58% |

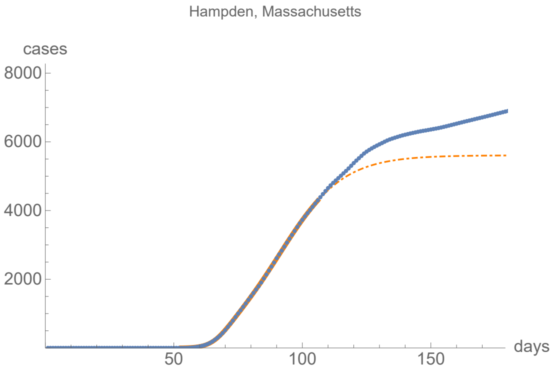


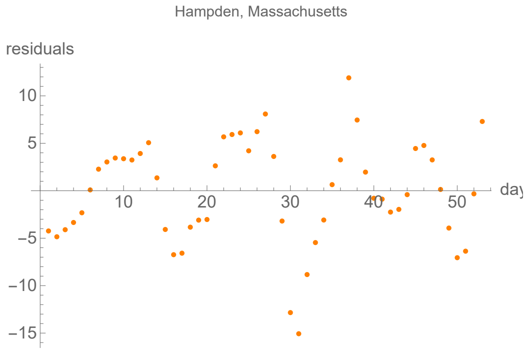


#### Parameters of fits to averaged death data.

The best fit ends on Mon 3 Aug 2020.

| α | β | k | τ | σ |
|---|---|---|---|---|
| 0.211±0.00464 | 0.0461±0.00091 | 696.±1.51 | 30.9±0.418 | 20.7±0.274 |

| badness | | α | β | k | τ | σ | quality |
|---|---|---|---|---|---|---|---|
| 1.6% | error | 2.2% | 2.% | 0.22% | 1.4% | 1.3% | 8.66% |

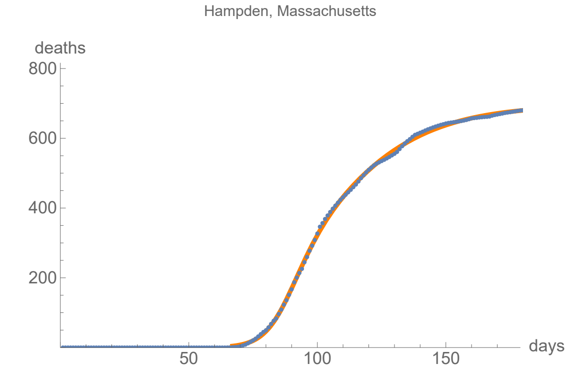


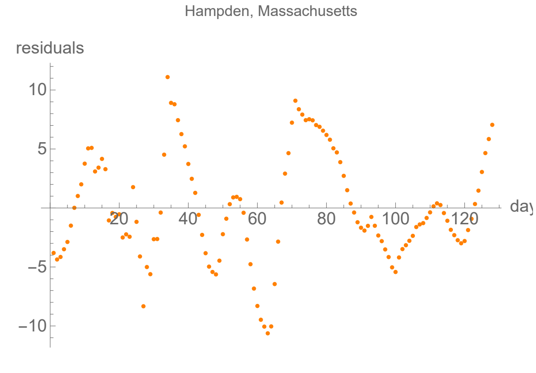


### Middlesex, Massachusetts

lag from integral overlap, using data averaged over 7 days, between 9 and 13 days, maximum overlap on day 11

lag from fits to the function I (averaged data): 10.2116 days

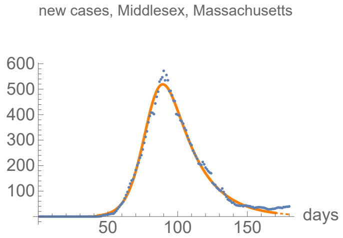


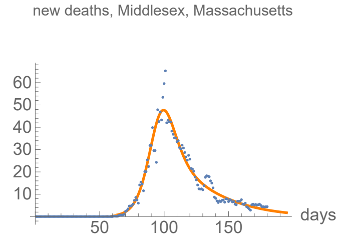


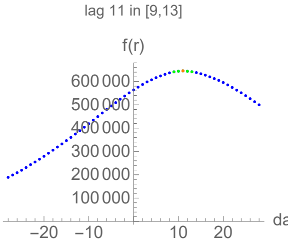


#### Parameters of fits to averaged cases data.

The best fit ends on Thu 9 Jul 2020.

| α | β | k | τ | σ |
|---|---|---|---|---|
| 0.165±0.00157 | 0.0512±0.00173 | 23000.±29.1 | 39.8±0.349 | 41.3±0.637 |

| badness | | α | β | k | τ | σ | quality |
|---|---|---|---|---|---|---|---|
| 0.55% | error | 0.95% | 3.4% | 0.13% | 0.88% | 1.5% | 4.82% |

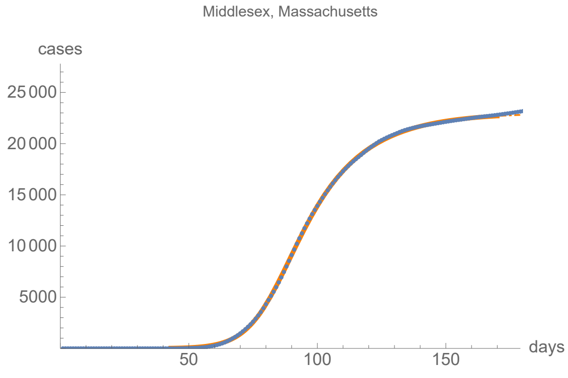


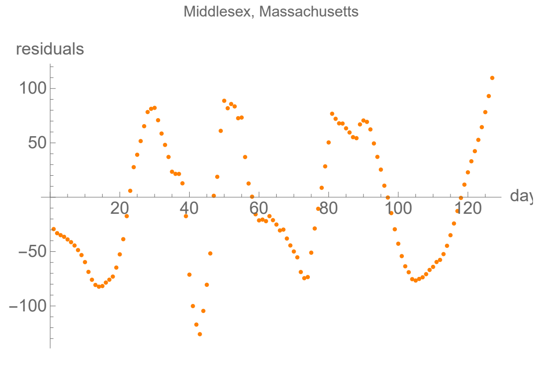


#### Parameters of fits to averaged death data.

The best fit ends on Mon 3 Aug 2020.

| α | β | k | τ | σ |
|---|---|---|---|---|
| 0.18±0.00212 | 0.0378±0.00261 | 2010.±4.62 | 32.1±0.199 | 32.1±0.551 |

| badness | | α | β | k | τ | σ | quality |
|---|---|---|---|---|---|---|---|
| 0.99% | error | 1.2% | 6.9% | 0.23% | 0.62% | 1.7% | 5.83% |

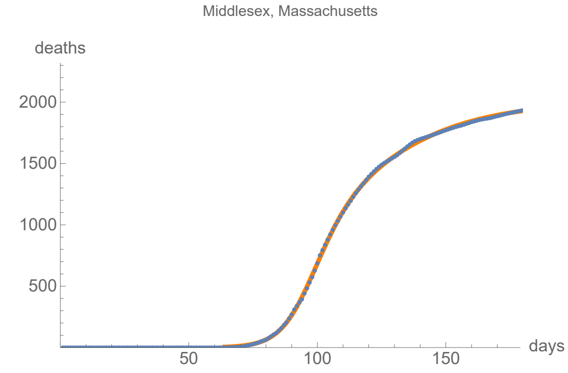


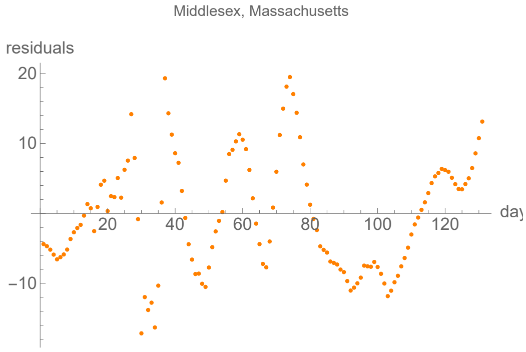


### Norfolk, Massachusetts

lag from integral overlap, using data averaged over 7 days, between 9 and 14 days, maximum overlap on day 12

lag from fits to the function I (averaged data): 9.99908 days

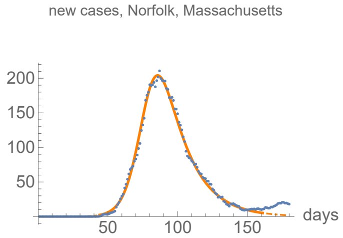


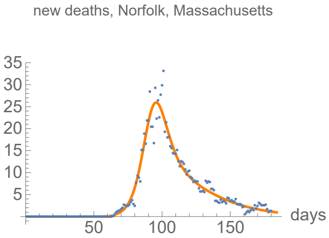


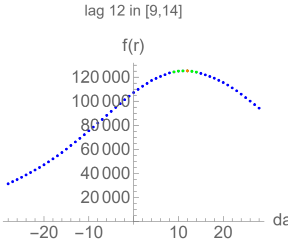


#### Parameters of fits to averaged cases data.

The best fit ends on Tue 30 Jun 2020.

| α | β | k | τ | σ |
|---|---|---|---|---|
| 0.176±0.00138 | 0.0563±0.00151 | 8460.±8.22 | 35.2±0.29 | 36.9±0.504 |

| badness | | α | β | k | τ | σ | quality |
|---|---|---|---|---|---|---|---|
| 0.43% | error | 0.78% | 2.7% | 0.097% | 0.82% | 1.4% | 4.13% |

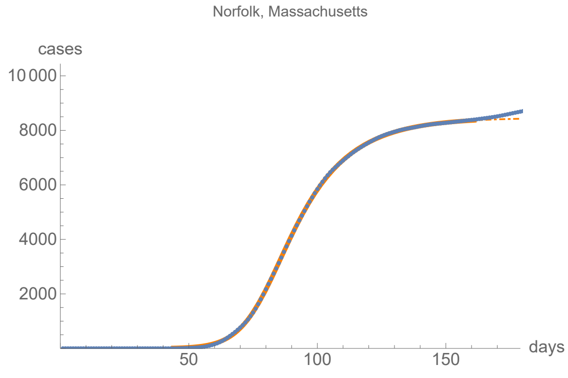


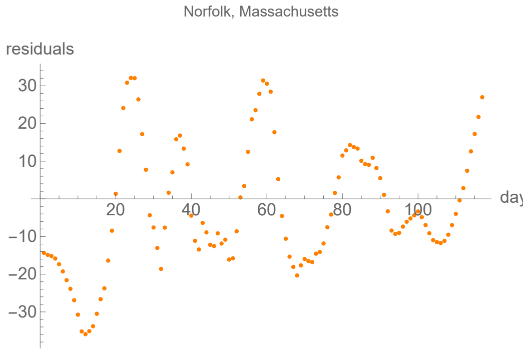


#### Parameters of fits to averaged death data.

The best fit ends on Thu 23 Jul 2020.

| α | β | k | τ | σ |
|---|---|---|---|---|
| 0.207±0.00227 | 0.0416±0.00288 | 987.±2.01 | 27.7±0.148 | 29.9±0.37 |

| badness | | α | β | k | τ | σ | quality |
|---|---|---|---|---|---|---|---|
| 1.% | error | 1.1% | 6.9% | 0.2% | 0.54% | 1.2% | 5.17% |

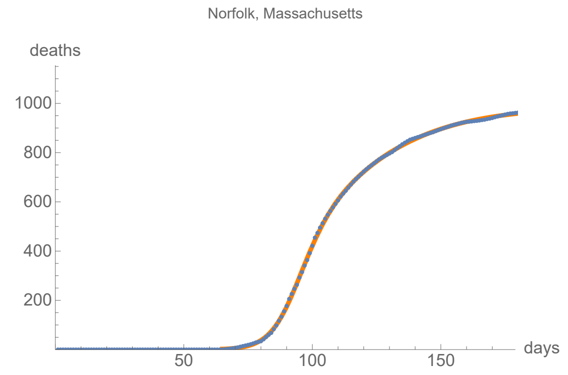


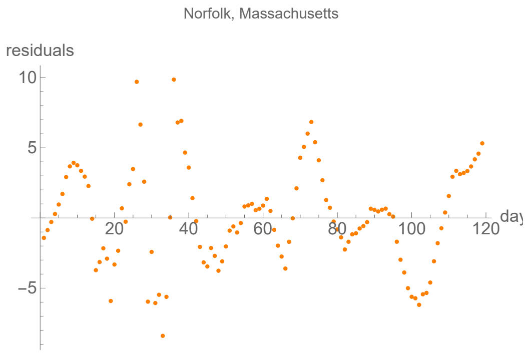


### Plymouth, Massachusetts

lag from integral overlap, using data averaged over 7 days, between 8 and 12 days, maximum overlap on day 10

lag from fits to the function I (averaged data): 8.7179 days

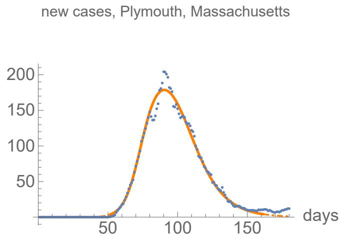


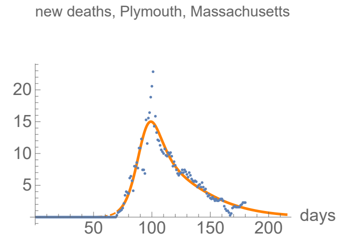


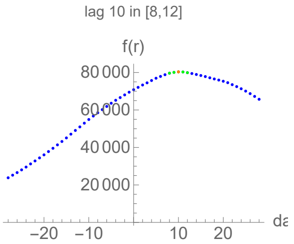


#### Parameters of fits to averaged cases data.

The best fit ends on Tue 30 Jun 2020.

| α | β | k | τ | σ |
|---|---|---|---|---|
| 0.207±0.00429 | 0.0735±0.000759 | 8320.±7.56 | 41.5±0.21 | 21.4±0.394 |

| badness | | α | β | k | τ | σ | quality |
|---|---|---|---|---|---|---|---|
| 0.85% | error | 2.1% | 1.% | 0.091% | 0.51% | 1.8% | 6.39% |

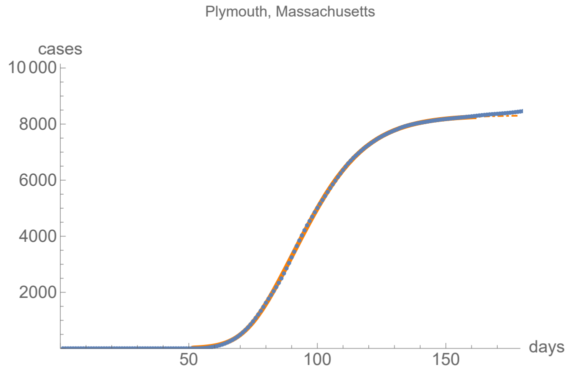


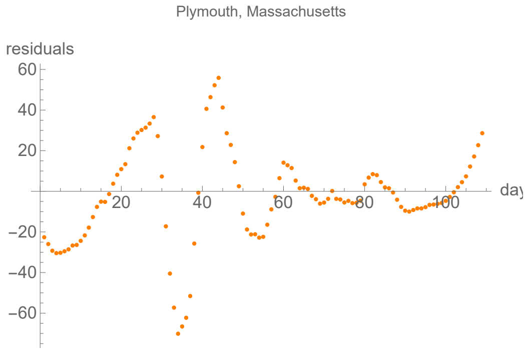


#### Parameters of fits to averaged death data.

The best fit ends on Sun 23 Aug 2020.

| α | β | k | τ | σ |
|---|---|---|---|---|
| 0.178±0.00356 | 0.0382±0.000795 | 732.±1.72 | 33.±0.526 | 23.5±0.303 |

| badness | | α | β | k | τ | σ | quality |
|---|---|---|---|---|---|---|---|
| 1.5% | error | 2.% | 2.1% | 0.23% | 1.6% | 1.3% | 8.71% |

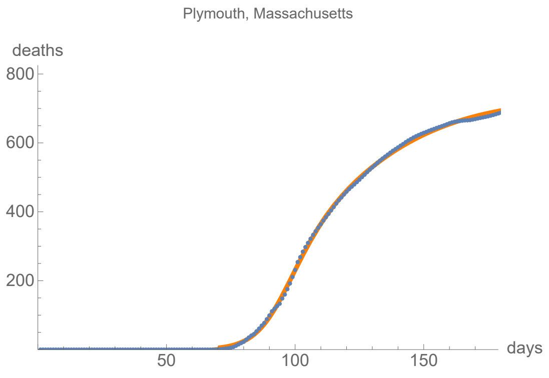


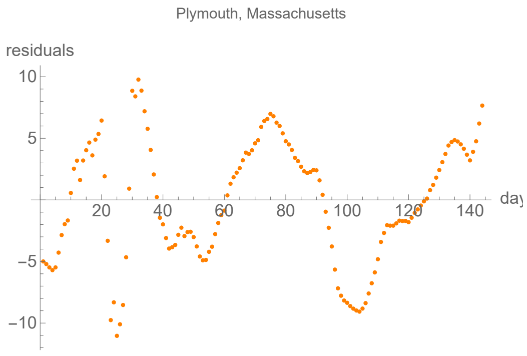


### Suffolk, Massachusetts

lag from integral overlap, using data averaged over 7 days, between 8 and 13 days, maximum overlap on day 10

lag from fits to the function I (averaged data): 15.7431 days

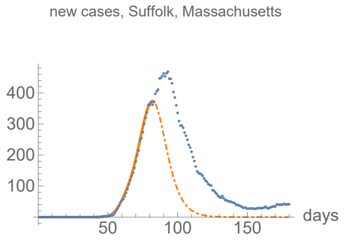


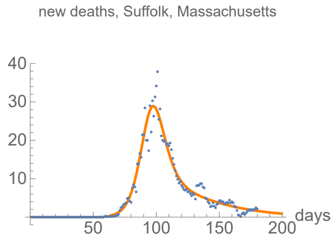


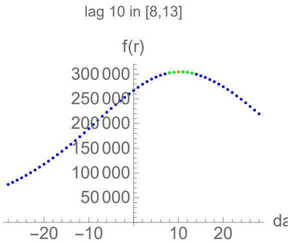


#### Parameters of fits to averaged cases data.

The best fit ends on Mon 13 Apr 2020.

| α | β | k | τ | σ |
|---|---|---|---|---|
| 0.418±0.0109 | 0.15±0.00118 | 9940.±107. | 40.±0.139 | 15.5±0.157 |

| badness | | α | β | k | τ | σ | quality |
|---|---|---|---|---|---|---|---|
| 0.14% | error | 2.6% | 0.79% | 1.1% | 0.35% | 1.% | 5.97% |

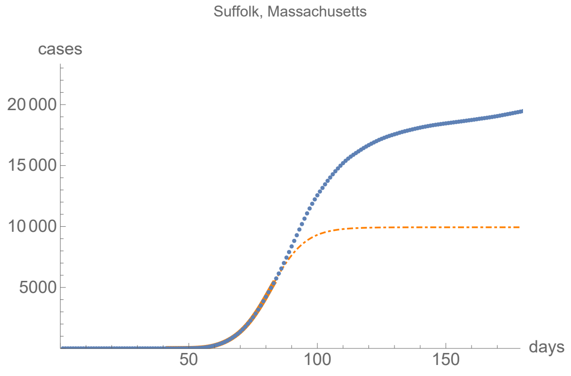


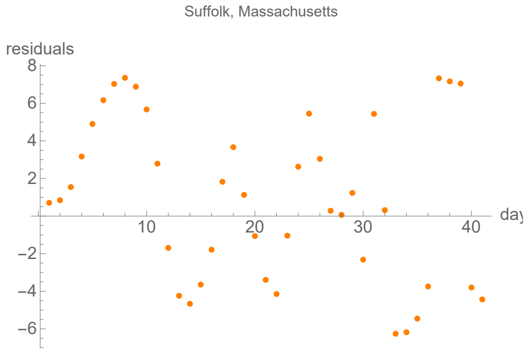


#### Parameters of fits to averaged death data.

The best fit ends on Fri 7 Aug 2020.

| α | β | k | τ | σ |
|---|---|---|---|---|
| 0.189±0.00214 | 0.0323±0.00266 | 1090.±2.96 | 31.5±0.15 | 24.±0.696 |

| badness | | α | β | k | τ | σ | quality |
|---|---|---|---|---|---|---|---|
| 1.1% | error | 1.1% | 8.2% | 0.27% | 0.48% | 2.9% | 6.91% |

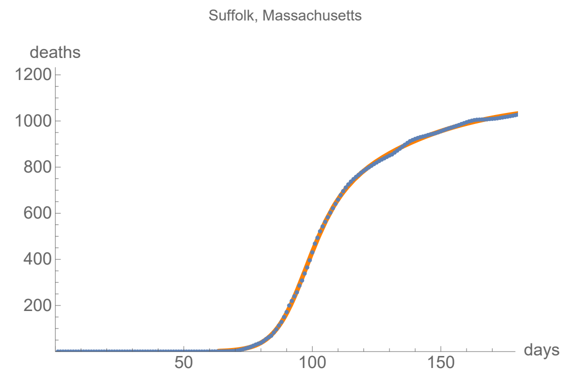


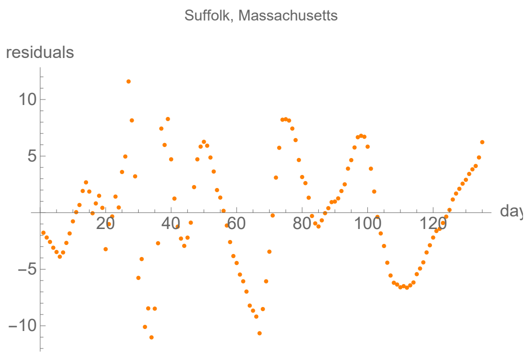


### Worcester, Massachusetts

lag from integral overlap, using data averaged over 7 days, between 9 and 16 days, maximum overlap on day 13

lag from fits to the function I (averaged data): 7.79107 days

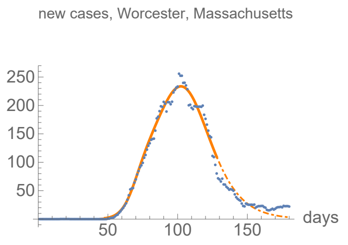


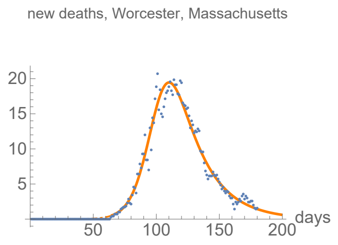


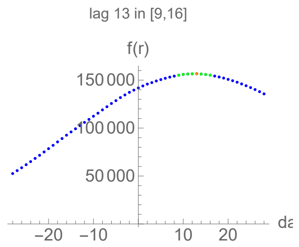


#### Parameters of fits to averaged cases data.

The best fit ends on Thu 28 May 2020.

| α | β | k | τ | σ |
|---|---|---|---|---|
| 0.225±0.00685 | 0.0735±0.000801 | 12400.±57.8 | 55.2±0.113 | 22.±0.353 |

| badness | | α | β | k | τ | σ | quality |
|---|---|---|---|---|---|---|---|
| 0.53% | error | 3.1% | 1.1% | 0.46% | 0.2% | 1.6% | 6.95% |

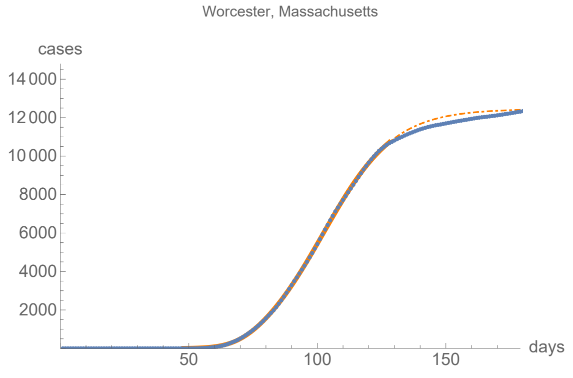


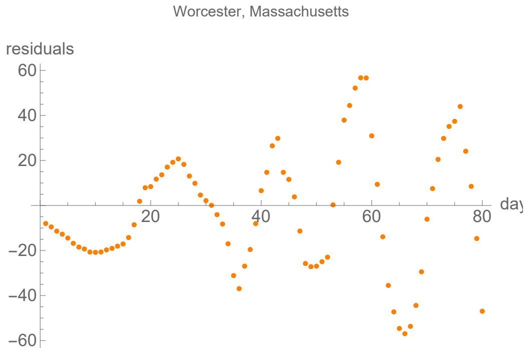


#### Parameters of fits to averaged death data.

The best fit ends on Fri 7 Aug 2020.

| α | β | k | τ | σ |
|---|---|---|---|---|
| 0.14±0.00216 | 0.0439±0.00231 | 999.±2.36 | 37.7±0.714 | 37.8±1.38 |

| badness | | α | β | k | τ | σ | quality |
|---|---|---|---|---|---|---|---|
| 0.94% | error | 1.5% | 5.3% | 0.24% | 1.9% | 3.7% | 9.44% |

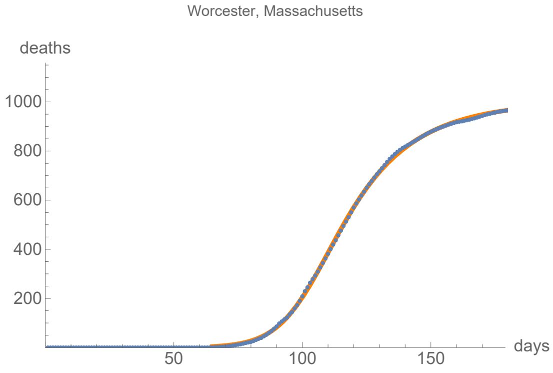


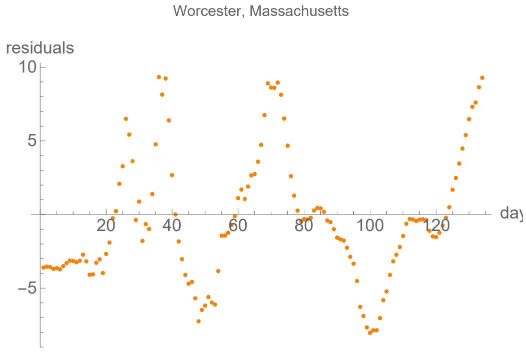


### Macomb, Michigan

lag from integral overlap, using data averaged over 7 days, between 6 and 9 days, maximum overlap on day 8

lag from fits to the function I (averaged data): 9.2049 days

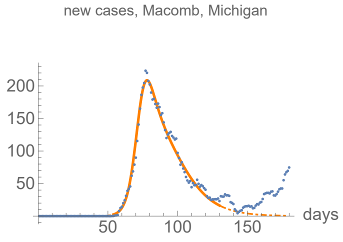


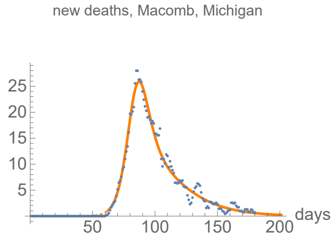


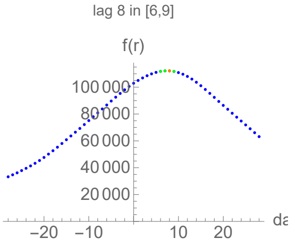


#### Parameters of fits to averaged cases data.

The best fit ends on Sat 30 May 2020.

| α | β | k | τ | σ |
|---|---|---|---|---|
| 0.295±0.00539 | 0.0726±0.0013 | 6890.±19.6 | 29.8±0.174 | 19.8±0.192 |

| badness | | α | β | k | τ | σ | quality |
|---|---|---|---|---|---|---|---|
| 0.76% | error | 1.8% | 1.8% | 0.28% | 0.58% | 0.97% | 6.22% |

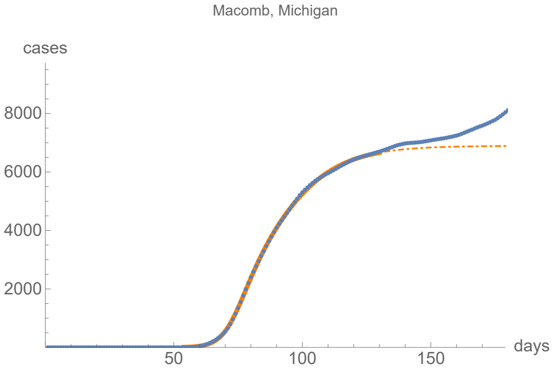


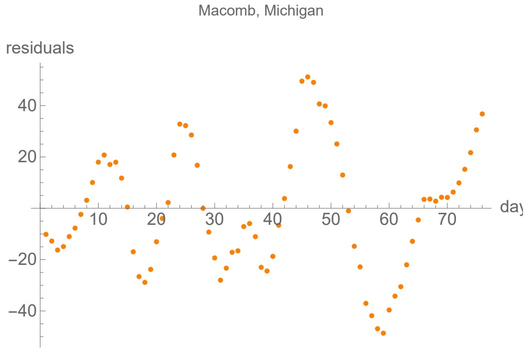


#### Parameters of fits to averaged death data.

The best fit ends on Sun 9 Aug 2020.

| α | β | k | τ | σ |
|---|---|---|---|---|
| 0.217±0.00243 | 0.0458±0.00316 | 942.±0.937 | 21.1±0.141 | 23.6±0.355 |

| badness | | α | β | k | τ | σ | quality |
|---|---|---|---|---|---|---|---|
| 0.85% | error | 1.1% | 6.9% | 0.1% | 0.67% | 1.5% | 5.44% |

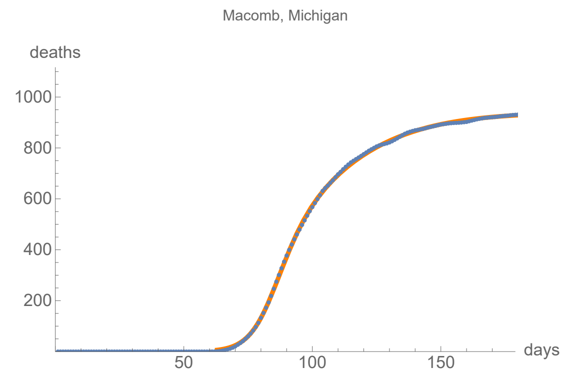


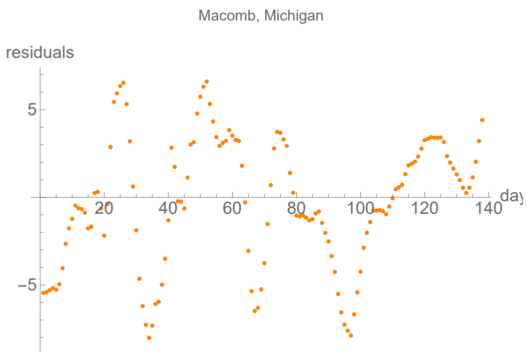


### Oakland, Michigan

lag from integral overlap, using data averaged over 7 days, between 3 and 9 days, maximum overlap on day 6

lag from fits to the function I (averaged data): 3.53659 days

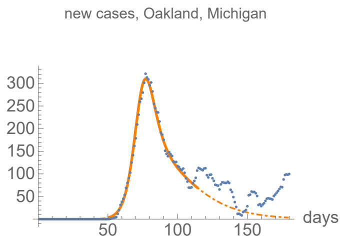


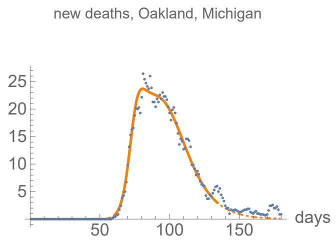


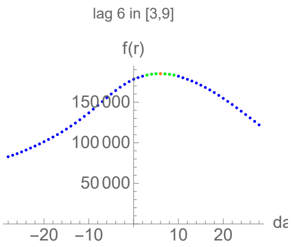


#### Parameters of fits to averaged cases data.

The best fit ends on Thu 14 May 2020.

| α | β | k | τ | σ |
|---|---|---|---|---|
| 0.253±0.0032 | 0.0508±0.00348 | 10500.±127. | 23.3±0.168 | 29.3±0.24 |

| badness | | α | β | k | τ | σ | quality |
|---|---|---|---|---|---|---|---|
| 0.53% | error | 1.3% | 6.9% | 1.2% | 0.72% | 0.82% | 5.38% |

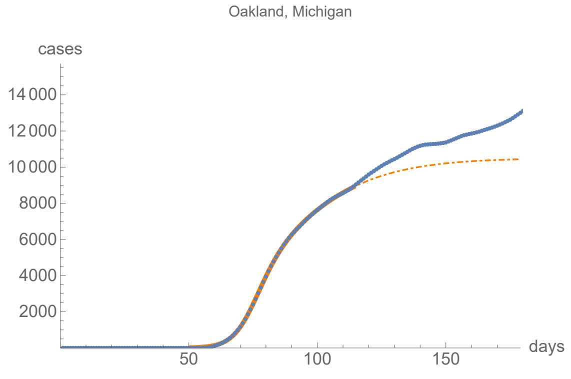


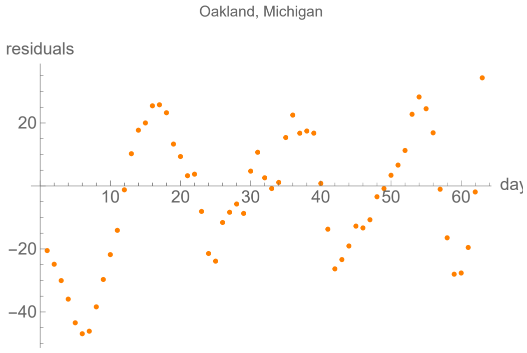


#### Parameters of fits to averaged death data.

The best fit ends on Wed 3 Jun 2020.

| α | β | k | τ | σ |
|---|---|---|---|---|
| 0.329±0.00601 | 0.0795±0.000571 | 1060.±1.77 | 32.2±0.0504 | 13.1±0.118 |

| badness | | α | β | k | τ | σ | quality |
|---|---|---|---|---|---|---|---|
| 0.61% | error | 1.8% | 0.72% | 0.17% | 0.16% | 0.9% | 4.39% |

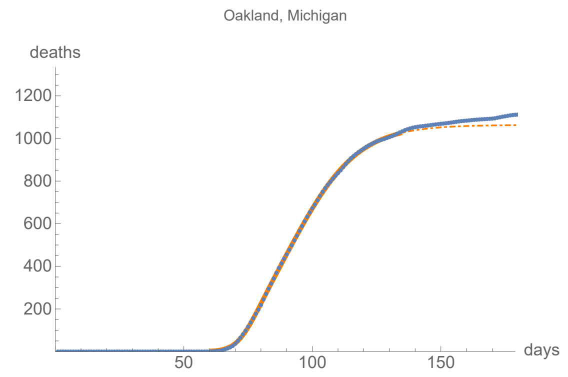


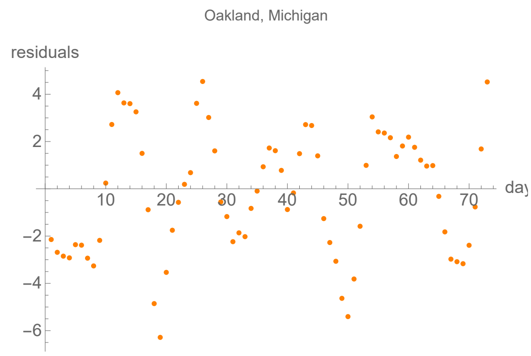


### Wayne, Michigan

lag from integral overlap, using data averaged over 7 days, between 7 and 10 days, maximum overlap on day 9

lag from fits to the function I (averaged data): 15.0953 days

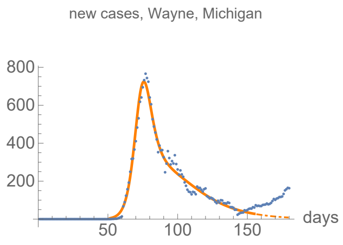


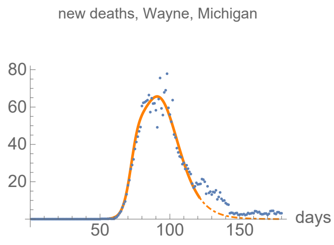


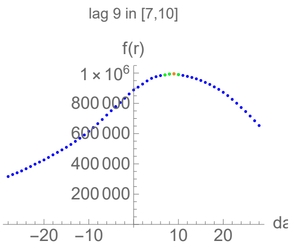


#### Parameters of fits to averaged cases data.

The best fit ends on Wed 24 Jun 2020.

| α | β | k | τ | σ |
|---|---|---|---|---|
| 0.296±0.00392 | 0.05±0.000601 | 22600.±43.3 | 30.±0.156 | 21.8±0.0933 |

| badness | | α | β | k | τ | σ | quality |
|---|---|---|---|---|---|---|---|
| 0.94% | error | 1.3% | 1.2% | 0.19% | 0.52% | 0.43% | 4.6% |

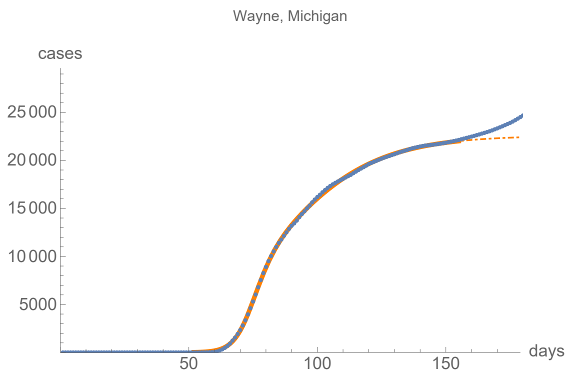


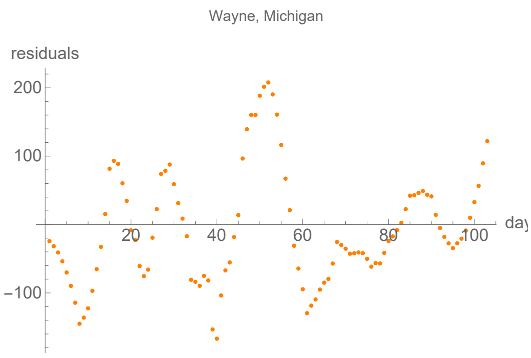


#### Parameters of fits to averaged death data.

The best fit ends on Thu 21 May 2020.

| α | β | k | τ | σ |
|---|---|---|---|---|
| 0.368±0.0147 | 0.103±0.00134 | 2470.±8.3 | 36.2±0.0723 | 17.5±0.244 |

| badness | | α | β | k | τ | σ | quality |
|---|---|---|---|---|---|---|---|
| 0.95% | error | 4.% | 1.3% | 0.34% | 0.2% | 1.4% | 8.18% |

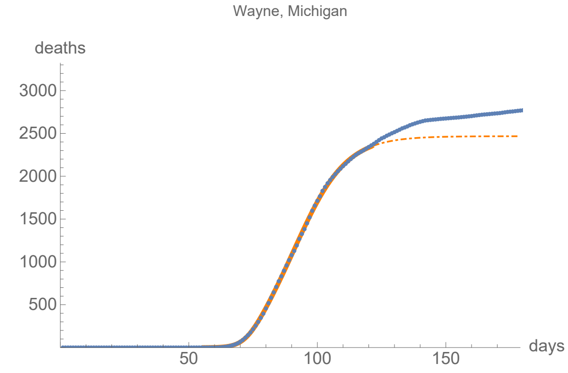


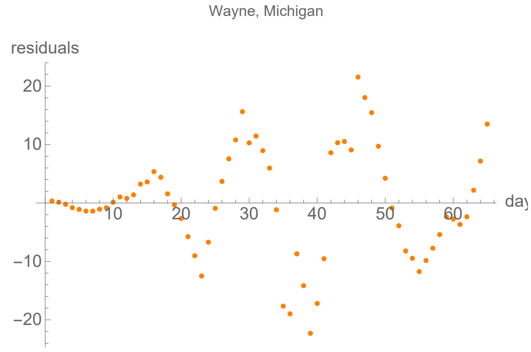


### Hennepin, Minnesota

lag from integral overlap, using data averaged over 7 days, between −13 and −8 days, maximum overlap on day −10

lag from fits to the function I (averaged data): −11.2948 days

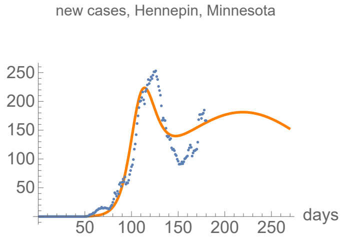


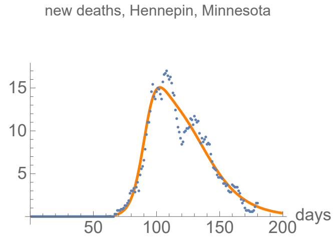


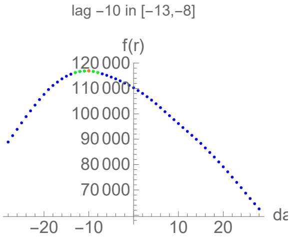


#### Parameters of fits to averaged cases data.

The best fit ends on Fri 16 Oct 2020.

| α | β | k | τ | σ |
|---|---|---|---|---|
| 0.132±0.00585 | 0.0168±0.00033 | 43100.±882. | 167.±2.61 | 56.8±0.521 |

| badness | | α | β | k | τ | σ | quality |
|---|---|---|---|---|---|---|---|
| 2.1% | error | 4.4% | 2.% | 2.% | 1.6% | 0.92% | 13.% |

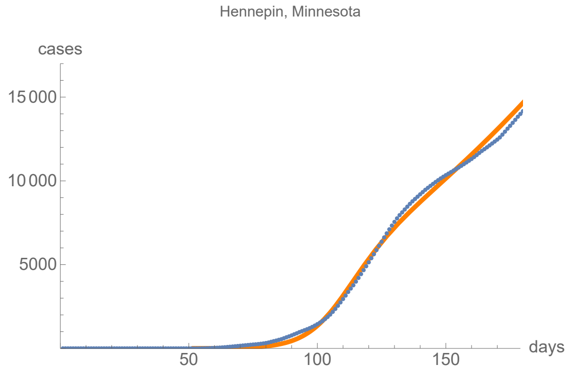


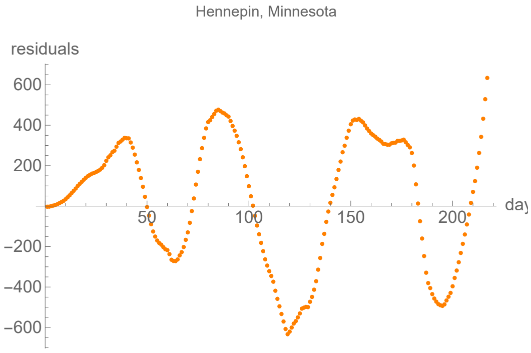


#### Parameters of fits to averaged death data.

The best fit ends on Fri 7 Aug 2020.

| α | β | k | τ | σ |
|---|---|---|---|---|
| 0.21±0.00611 | 0.0554±0.000794 | 822.±1.46 | 46.6±0.232 | 25.5±0.352 |

| badness | | α | β | k | τ | σ | quality |
|---|---|---|---|---|---|---|---|
| 1.5% | error | 2.9% | 1.4% | 0.18% | 0.5% | 1.4% | 7.85% |

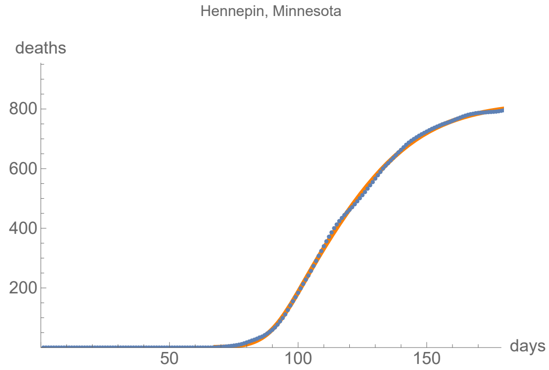


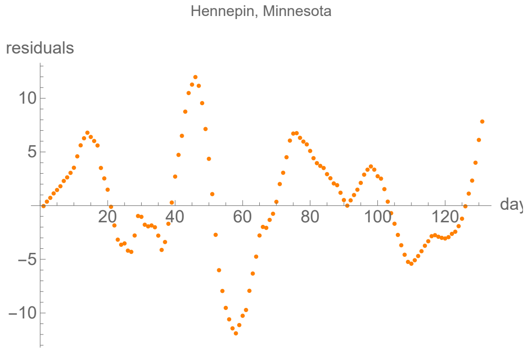


### St. Louis, Missouri

lag from integral overlap, using data averaged over 7 days, between 11 and 16 days, maximum overlap on day 14

lag from fits to the function I (averaged data): 23.5652 days

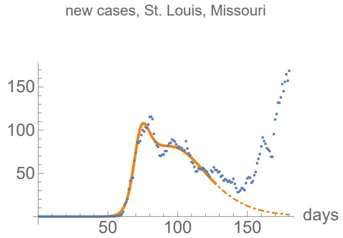


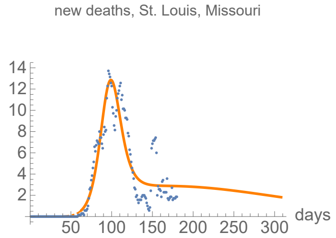


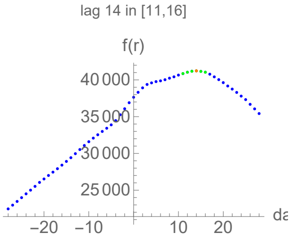


#### Parameters of fits to averaged cases data.

The best fit ends on Wed 27 May 2020.

| α | β | k | τ | σ |
|---|---|---|---|---|
| 0.32±0.00768 | 0.0587±0.000829 | 5440.±36.4 | 44.±0.232 | 20.2±0.145 |

| badness | | α | β | k | τ | σ | quality |
|---|---|---|---|---|---|---|---|
| 0.8% | error | 2.4% | 1.4% | 0.67% | 0.53% | 0.72% | 6.52% |

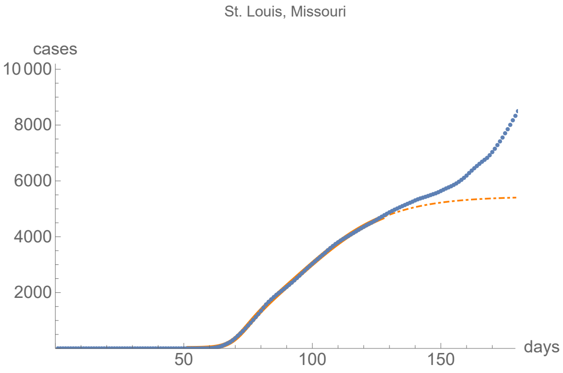


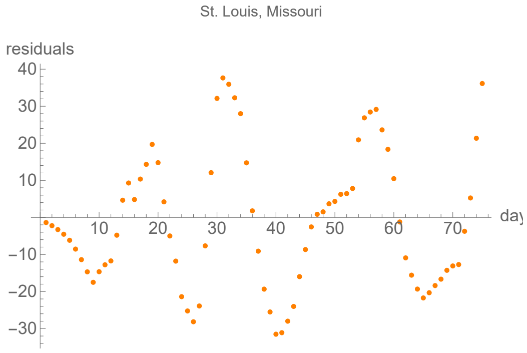


#### Parameters of fits to averaged death data.

The best fit ends on Tue 24 Nov 2020.

| α | β | k | τ | σ |
|---|---|---|---|---|
| 0.13±0.00251 | 0.01±0.00348 | 1150.±14.1 | 34.2±0.216 | 105.±2.46 |

| badness | | α | β | k | τ | σ | quality |
|---|---|---|---|---|---|---|---|
| 2.2% | error | 1.9% | 35.% | 1.2% | 0.63% | 2.4% | 10.4% |

St. Louis, Missouri

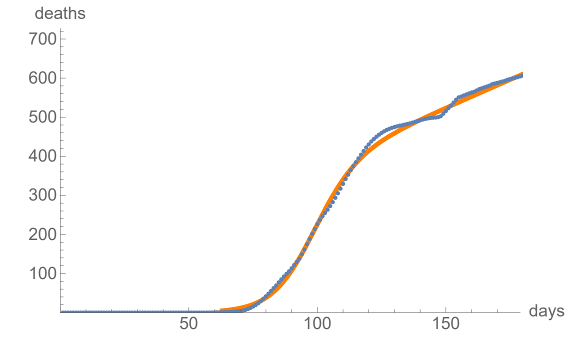


St. Louis, Missouri

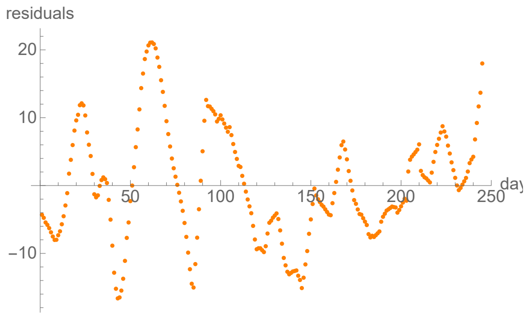


### Clark, Nevada

no clear lag in the first six months of the pandemic from integral overlap using data averaged over 7 days

lag from fits to the function I (averaged data): 7.83028 days

new cases, Clark, Nevada

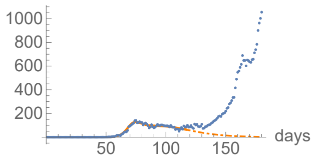


new deaths, Clark, Nevada

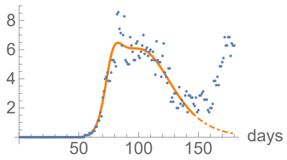


no clear lag in the first six months of the pandemic

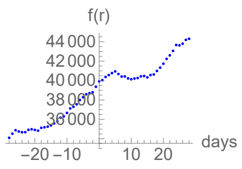


#### Parameters of fits to averaged cases data.

The best fit ends on Sun 17 May 2020.

| α | β | k | τ | σ |
|---|---|---|---|---|
| 0.307±0.0072 | 0.0573±0.00127 | 6470.±87.2 | 46.8±0.476 | 23.3±0.171 |

| badness | | α | β | k | τ | σ | quality |
|---|---|---|---|---|---|---|---|
| 0.82% | error | 2.3% | 2.2% | 1.3% | 1.% | 0.73% | 8.49% |

Clark, Nevada

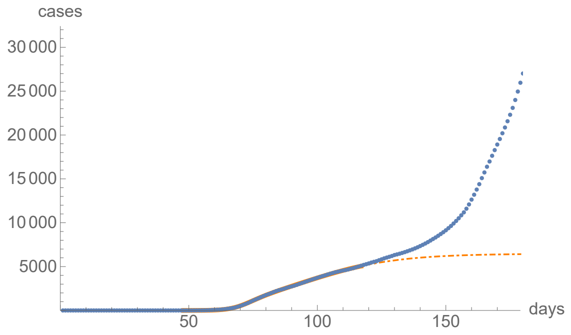


Clark, Nevada

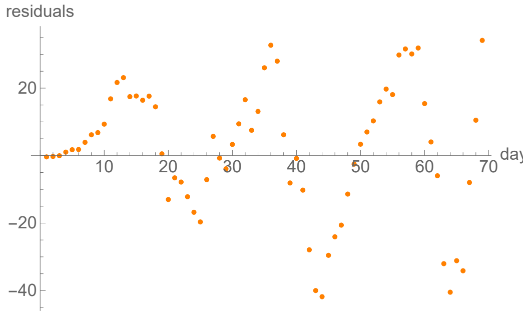


#### Parameters of fits to averaged death data.

The best fit ends on Mon 15 Jun 2020.

| α | β | k | τ | σ |
|---|---|---|---|---|
| 0.264±0.00735 | 0.0589±0.000678 | 400.±1.5 | 43.7±0.127 | 16.9±0.213 |

| badness | | α | β | k | τ | σ | quality |
|---|---|---|---|---|---|---|---|
| 1.2% | error | 2.8% | 1.2% | 0.38% | 0.29% | 1.3% | 7.05% |

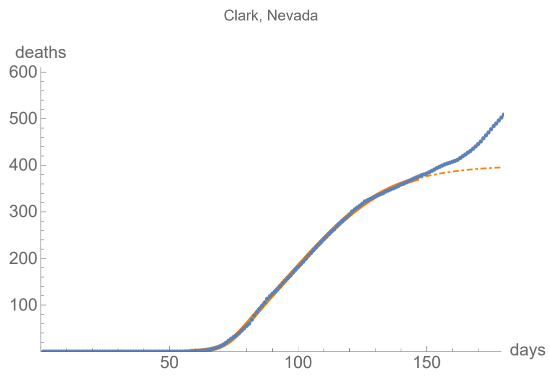


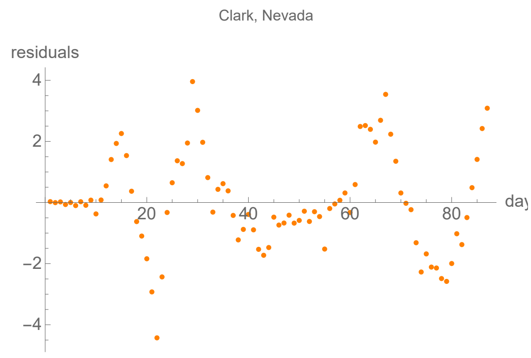


### Bergen, New Jersey

lag from integral overlap, using data averaged over 7 days, between 7 and 11 days, maximum overlap on day 9

lag from fits to the function I (averaged data): 5.729 days

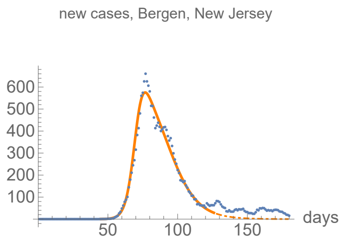


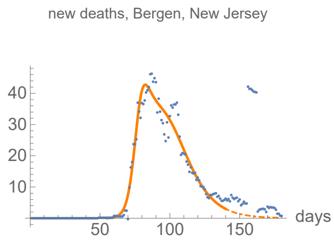


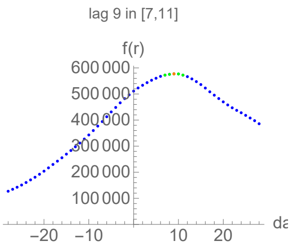


#### Parameters of fits to averaged cases data.

The best fit ends on Tue 26 May 2020.

| α | β | k | τ | σ |
|---|---|---|---|---|
| 0.333±0.00831 | 0.0916±0.00157 | 17900.±40.2 | 37.2±0.167 | 26.2±0.231 |

| badness | | α | β | k | τ | σ | quality |
|---|---|---|---|---|---|---|---|
| 0.94% | error | 2.5% | 1.7% | 0.22% | 0.45% | 0.88% | 6.7% |

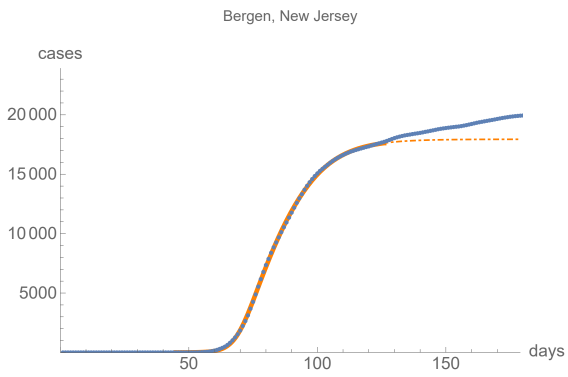


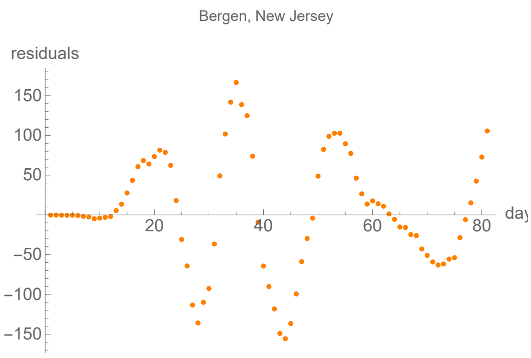


#### Parameters of fits to averaged death data.

The best fit ends on Tue 9 Jun 2020.

| α | β | k | τ | σ |
|---|---|---|---|---|
| 0.346±0.0122 | 0.0806±0.00125 | 1640.±4.72 | 38.6±0.118 | 22.7±0.222 |

| badness | | α | β | k | τ | σ | quality |
|---|---|---|---|---|---|---|---|
| 1.5% | error | 3.5% | 1.6% | 0.29% | 0.31% | 0.98% | 8.11% |

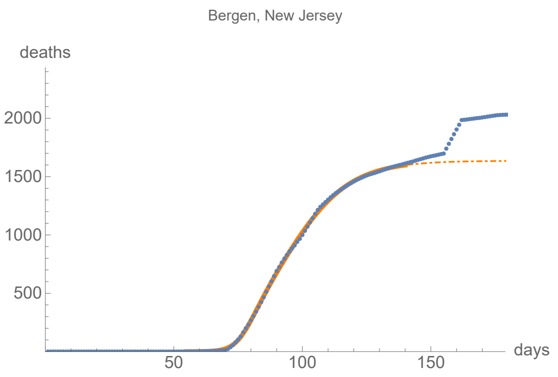


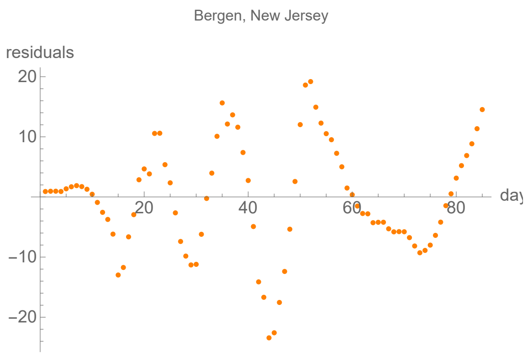


### Camden, New Jersey

lag from integral overlap, using data averaged over 7 days, between 8 and 16 days, maximum overlap on day 11

lag from fits to the function I (averaged data): 11.0318 days

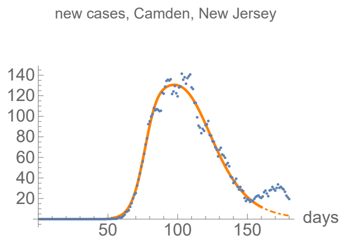


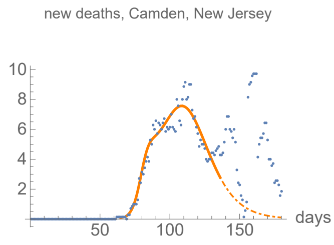


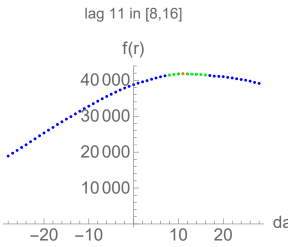


#### Parameters of fits to averaged cases data.

The best fit ends on Mon 29 Jun 2020.

| α | β | k | τ | σ |
|---|---|---|---|---|
| 0.225±0.00482 | 0.0649±0.000552 | 7390.±11.3 | 50.6±0.0883 | 25.5±0.239 |

| badness | | α | β | k | τ | σ | quality |
|---|---|---|---|---|---|---|---|
| 0.92% | error | 2.1% | 0.85% | 0.15% | 0.17% | 0.94% | 5.18% |

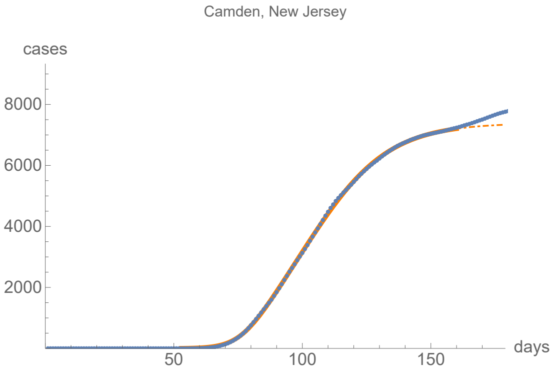


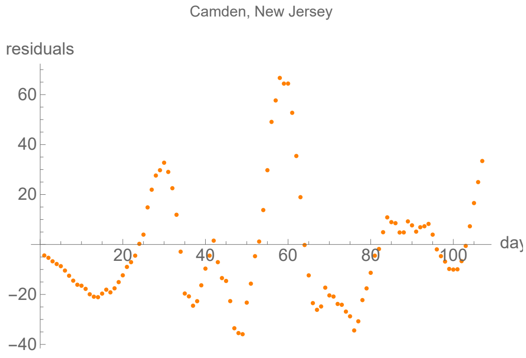


#### Parameters of fits to averaged death data.

The best fit ends on Fri 5 Jun 2020.

| α | β | k | τ | σ |
|---|---|---|---|---|
| 0.331±0.0218 | 0.0795±0.00132 | 379.±2.8 | 39.8±0.207 | 12.2±0.377 |

| badness | | α | β | k | τ | σ | quality |
|---|---|---|---|---|---|---|---|
| 1.3% | error | 6.6% | 1.7% | 0.74% | 0.52% | 3.1% | 13.9% |

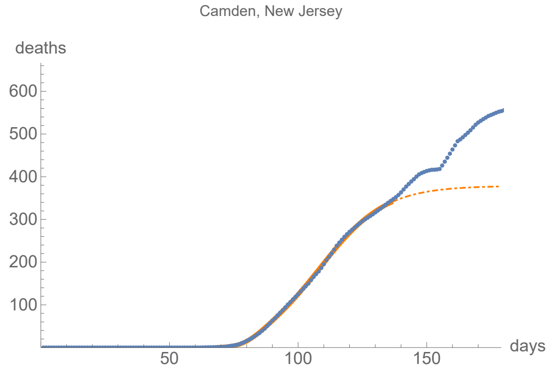


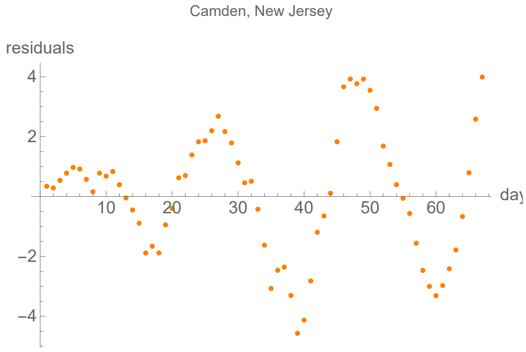


### Essex, New Jersey

lag from integral overlap, using data averaged over 7 days, between 5 and 9 days, maximum overlap on day 7

lag from fits to the function I (averaged data): 8.98589 days

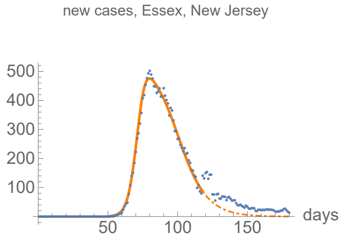


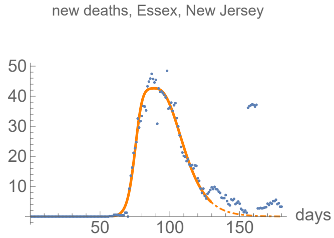


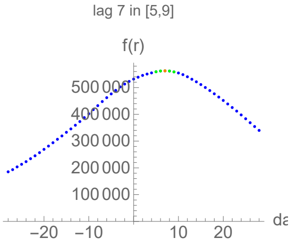


#### Parameters of fits to averaged cases data.

The best fit ends on Mon 18 May 2020.

| α | β | k | τ | σ |
|---|---|---|---|---|
| 0.321±0.00359 | 0.0884±0.00072 | 17100.±31.6 | 35.±0.0462 | 20.8±0.108 |

| badness | | α | β | k | τ | σ | quality |
|---|---|---|---|---|---|---|---|
| 0.3% | error | 1.1% | 0.82% | 0.18% | 0.13% | 0.52% | 3.07% |

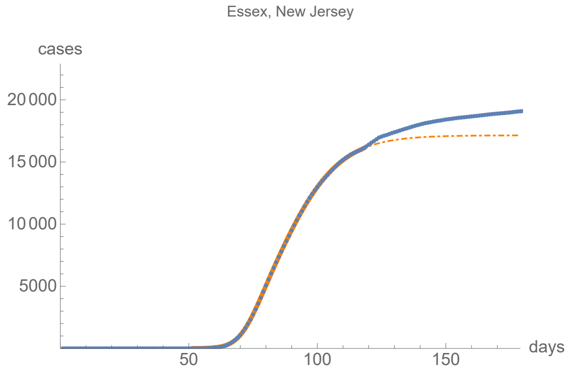


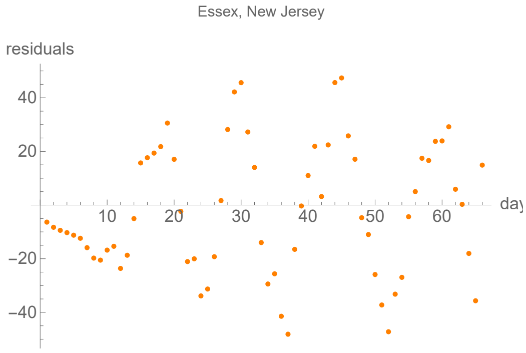


#### Parameters of fits to averaged death data.

The best fit ends on Fri 29 May 2020.

| α | β | k | τ | σ |
|---|---|---|---|---|
| 0.366±0.0108 | 0.0962±0.00107 | 1660.±3.95 | 33.9±0.0668 | 17.±0.182 |

| badness | | α | β | k | τ | σ | quality |
|---|---|---|---|---|---|---|---|
| 0.81% | error | 3.% | 1.1% | 0.24% | 0.2% | 1.1% | 6.38% |

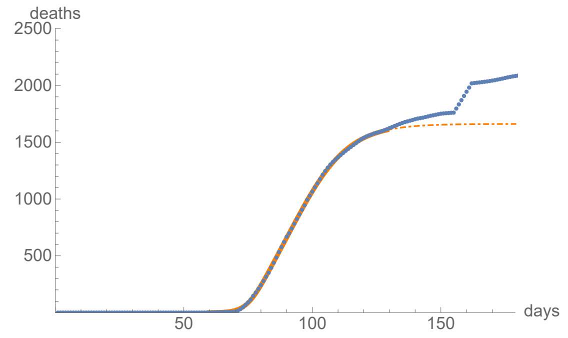


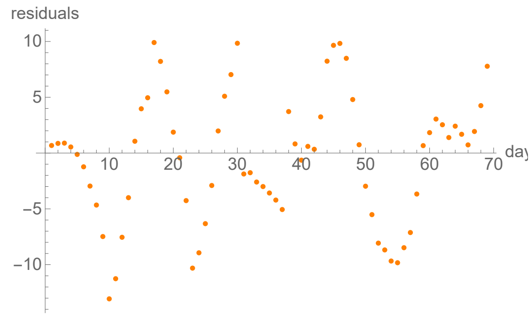


### Hudson, New Jersey

lag from integral overlap, using data averaged over 7 days, between 7 and 11 days, maximum overlap on day 9

lag from fits to the function I (averaged data): 8.35023 days

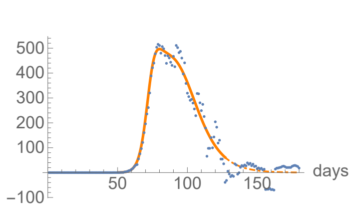


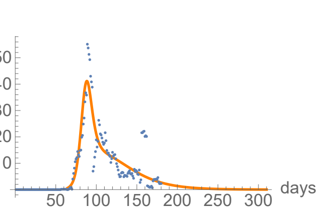


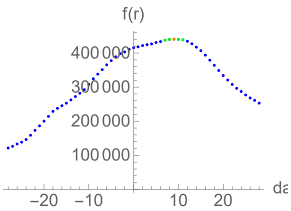


#### Parameters of fits to averaged cases data.

The best fit ends on Wed 27 May 2020.

| α | β | k | τ | σ |
|---|---|---|---|---|
| 0.35±0.00958 | 0.0884±0.00106 | 19600.±51.1 | 39.±0.0752 | 22.4±0.179 |

| badness | | α | β | k | τ | σ | quality |
|---|---|---|---|---|---|---|---|
| 0.88% | error | 2.7% | 1.2% | 0.26% | 0.19% | 0.8% | 6.07% |

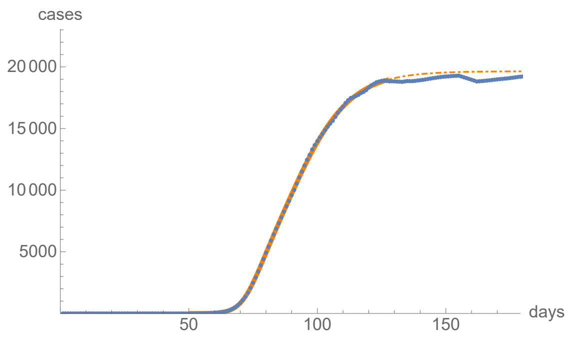


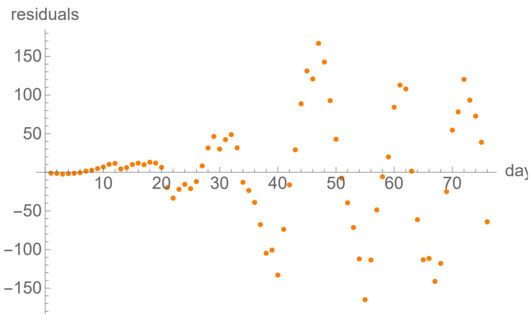


#### Parameters of fits to averaged death data.

The best fit ends on Thu 26 Nov 2020.

| α | β | k | τ | σ |
|---|---|---|---|---|
| 0.261±0.011 | 0.0365±0.000674 | 1530.±2.41 | 34.4±0.512 | 21.6±0.27 |

| badness | | α | β | k | τ | σ | quality |
|---|---|---|---|---|---|---|---|
| 3.6% | error | 4.2% | 1.8% | 0.16% | 1.5% | 1.3% | 12.6% |

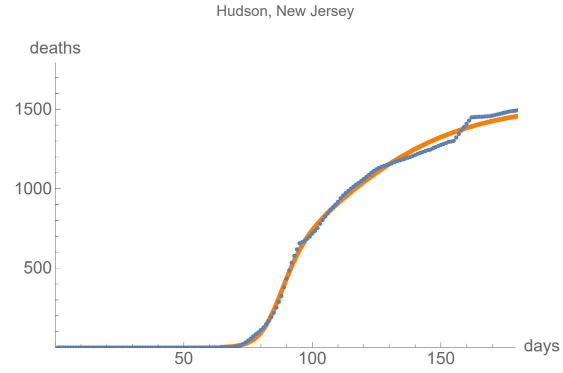


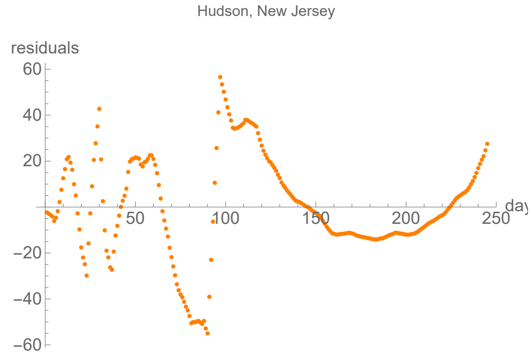


### Mercer, New Jersey

lag from integral overlap, using data averaged over 7 days, between 4 and 11 days, maximum overlap on day 8

lag from fits to the function I (averaged data): 10.9576 days

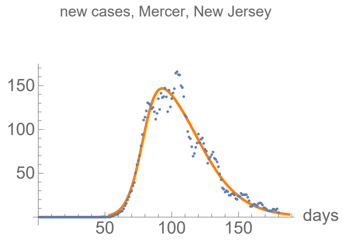


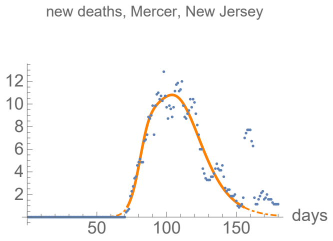


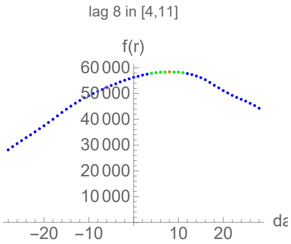


#### Parameters of fits to averaged cases data.

The best fit ends on Mon 27 Jul 2020.

| α | β | k | τ | σ |
|---|---|---|---|---|
| 0.191±0.0054 | 0.0572±0.000924 | 7880.±12.4 | 46.7±0.331 | 26.9±0.473 |

| badness | | α | β | k | τ | σ | quality |
|---|---|---|---|---|---|---|---|
| 1.4% | error | 2.8% | 1.6% | 0.16% | 0.71% | 1.8% | 8.51% |

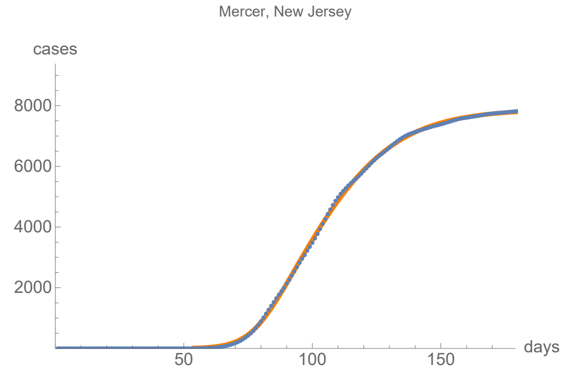


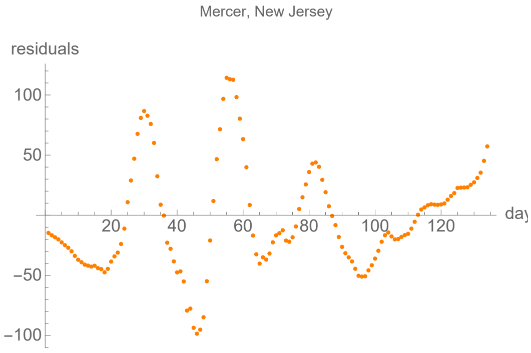


#### Parameters of fits to averaged death data.

The best fit ends on Wed 24 Jun 2020.

| α | β | k | τ | σ |
|---|---|---|---|---|
| 0.278±0.0142 | 0.0775±0.00114 | 536.±1.52 | 34.3±0.123 | 10.5±0.387 |

| badness | | α | β | k | τ | σ | quality |
|---|---|---|---|---|---|---|---|
| 1.4% | error | 5.1% | 1.5% | 0.28% | 0.36% | 3.7% | 12.3% |

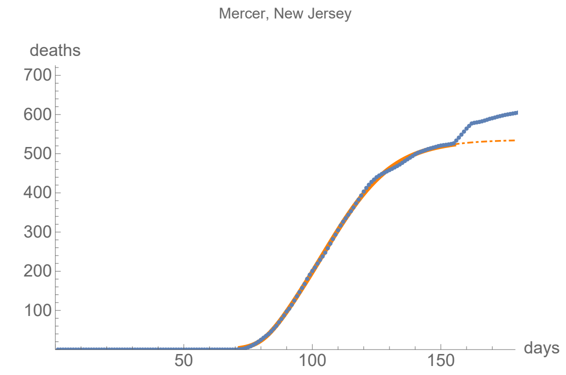


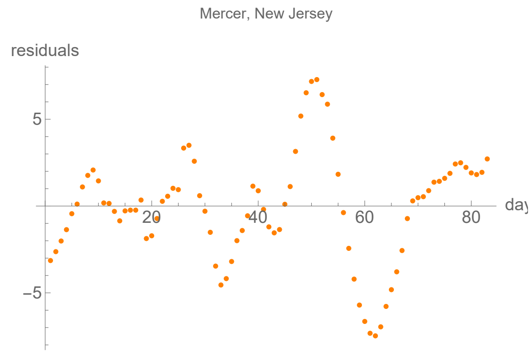


### Middlesex, New Jersey

lag from integral overlap, using data averaged over 7 days, between 6 and 11 days, maximum overlap on day 9

lag from fits to the function I (averaged data): 2.45389 days

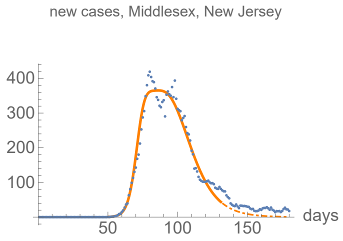


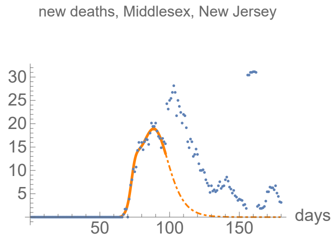


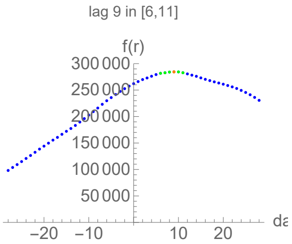


#### Parameters of fits to averaged cases data.

The best fit ends on Sun 31 May 2020.

| α | β | k | τ | σ |
|---|---|---|---|---|
| 0.334±0.0129 | 0.0859±0.0012 | 16000.±49.1 | 40.6±0.0942 | 21.6±0.253 |

| badness | | α | β | k | τ | σ | quality |
|---|---|---|---|---|---|---|---|
| 0.92% | error | 3.9% | 1.4% | 0.31% | 0.23% | 1.2% | 7.88% |

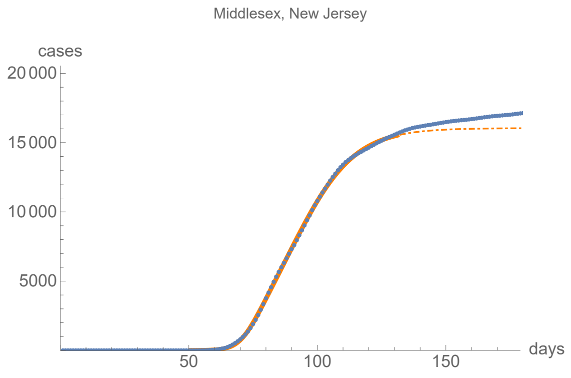


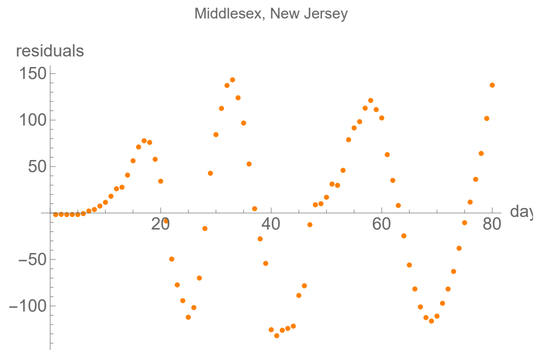


#### Parameters of fits to averaged death data.

The best fit ends on Mon 27 Apr 2020.

| α | β | k | τ | σ |
|---|---|---|---|---|
| 0.59±0.0252 | 0.141±0.00277 | 534.±7.47 | 21.8±0.211 | 6.74±0.152 |

| badness | | α | β | k | τ | σ | quality |
|---|---|---|---|---|---|---|---|
| 0.53% | error | 4.3% | 2.% | 1.4% | 0.97% | 2.3% | 11.4% |

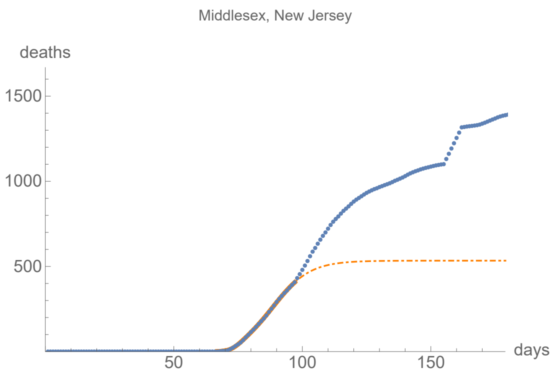


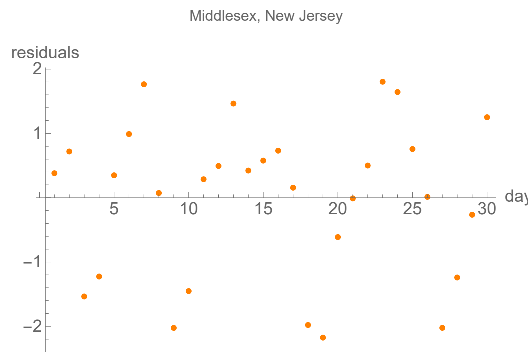


### Monmouth, New Jersey

lag from integral overlap, using data averaged over 7 days, between 9 and 12 days, maximum overlap on day 10

lag from fits to the function I (averaged data): 10.405 days

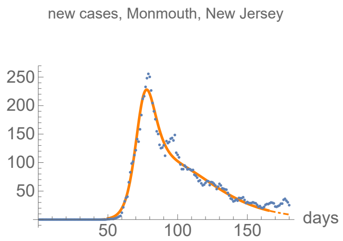


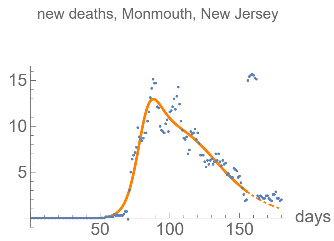


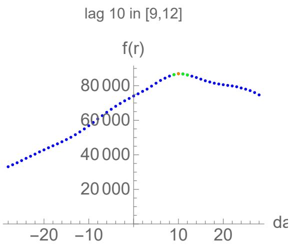


#### Parameters of fits to averaged cases data.

The best fit ends on Sat 4 Jul 2020.

| α | β | k | τ | σ |
|---|---|---|---|---|
| 0.25±0.00399 | 0.0421±0.000532 | 9520.±25.5 | 40.6±0.146 | 25.1±0.128 |

| badness | | α | β | k | τ | σ | quality |
|---|---|---|---|---|---|---|---|
| 1.3% | error | 1.6% | 1.3% | 0.27% | 0.36% | 0.51% | 5.26% |

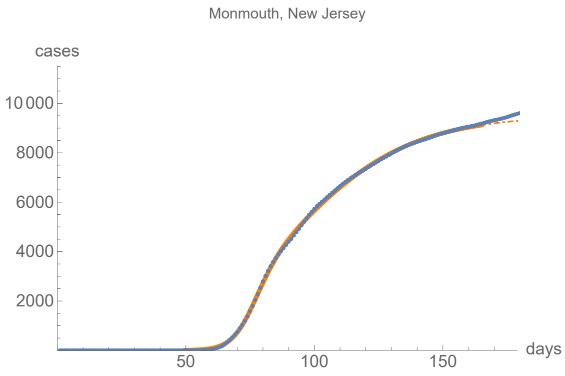


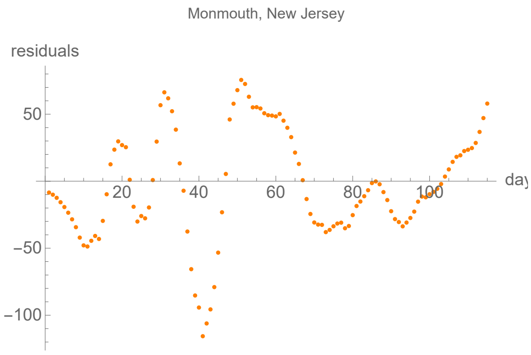


#### Parameters of fits to averaged death data.

The best fit ends on Wed 24 Jun 2020.

| α | β | k | τ | σ |
|---|---|---|---|---|
| 0.216±0.00687 | 0.0477±0.00116 | 762.±5.71 | 47.1±0.255 | 22.9±0.361 |

| badness | | α | β | k | τ | σ | quality |
|---|---|---|---|---|---|---|---|
| 1.4% | error | 3.2% | 2.4% | 0.75% | 0.54% | 1.6% | 9.87% |

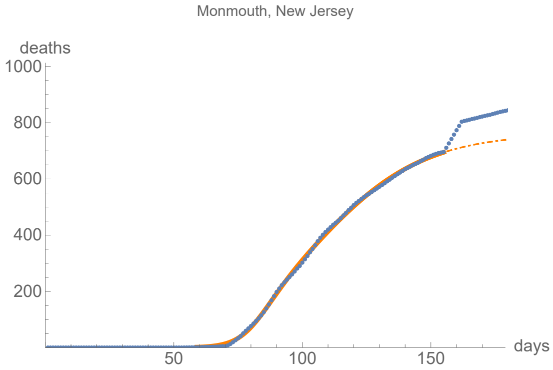


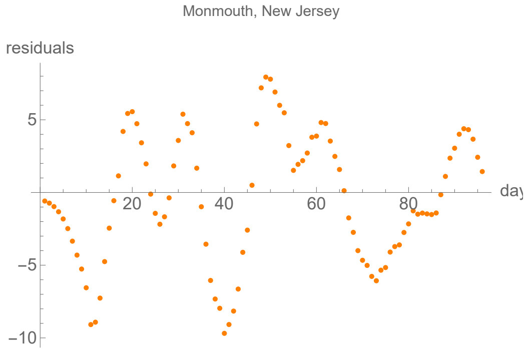


### Morris, New Jersey

lag from integral overlap, using data averaged over 7 days, between 6 and 10 days, maximum overlap on day 8

lag from fits to the function I (averaged data): 4.2849 days

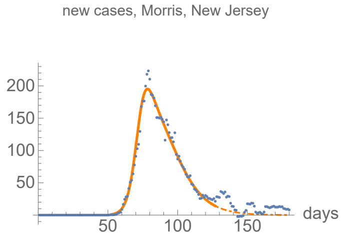


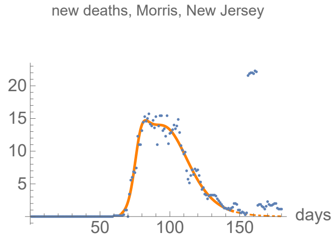


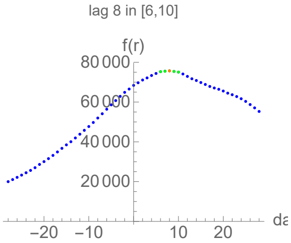


#### Parameters of fits to averaged cases data.

The best fit ends on Wed 27 May 2020.

| α | β | k | τ | σ |
|---|---|---|---|---|
| 0.313±0.00587 | 0.0834±0.00126 | 6370.±14.4 | 31.2±0.141 | 20.2±0.19 |

| badness | | α | β | k | τ | σ | quality |
|---|---|---|---|---|---|---|---|
| 0.7% | error | 1.9% | 1.5% | 0.23% | 0.45% | 0.94% | 5.71% |

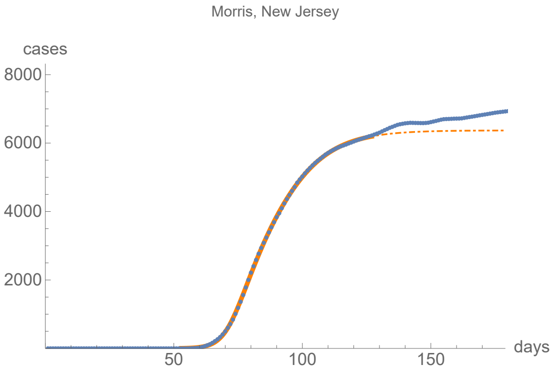


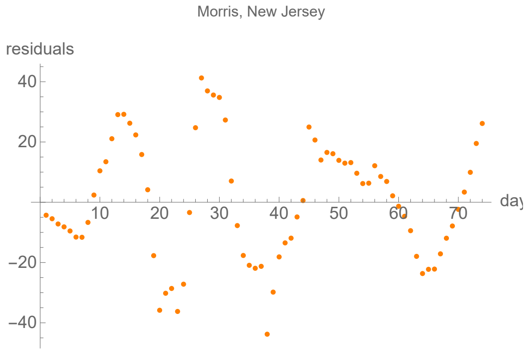


#### Parameters of fits to averaged death data.

The best fit ends on Sun 14 Jun 2020.

| α | β | k | τ | σ |
|---|---|---|---|---|
| 0.37±0.0107 | 0.0846±0.000685 | 638.±0.98 | 30.1±0.0623 | 11.4±0.149 |

| badness | | α | β | k | τ | σ | quality |
|---|---|---|---|---|---|---|---|
| 0.98% | error | 2.9% | 0.81% | 0.15% | 0.21% | 1.3% | 6.34% |

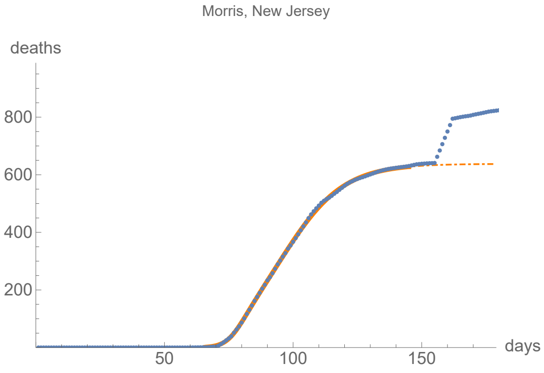


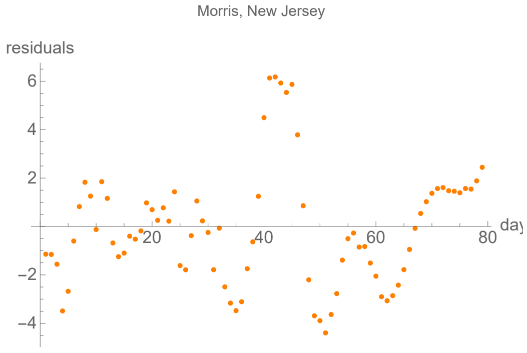


### Ocean, New Jersey

lag from integral overlap, using data averaged over 7 days, between 10 and 19 days, maximum overlap on day 14

lag from fits to the function I (averaged data): 0.709819 days

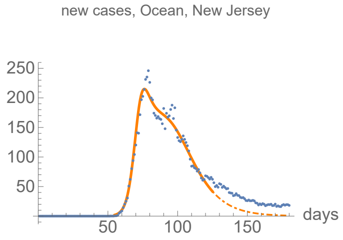


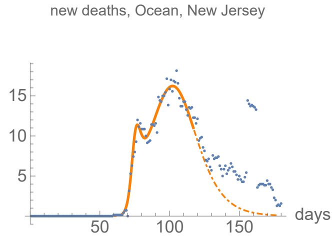


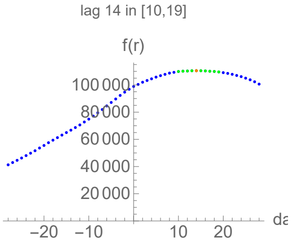


#### Parameters of fits to averaged cases data.

The best fit ends on Mon 25 May 2020.

| α | β | k | τ | σ |
|---|---|---|---|---|
| 0.345±0.00802 | 0.074±0.000925 | 8820.±32.7 | 34.8±0.0932 | 17.1±0.144 |

| badness | | α | β | k | τ | σ | quality |
|---|---|---|---|---|---|---|---|
| 0.71% | error | 2.3% | 1.2% | 0.37% | 0.27% | 0.84% | 5.77% |

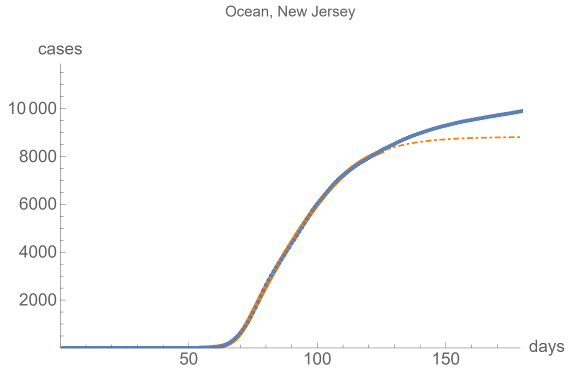


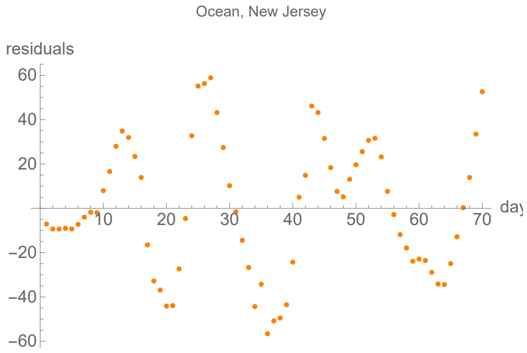


#### Parameters of fits to averaged death data.

The best fit ends on Sun 17 May 2020.

| α | β | k | τ | σ |
|---|---|---|---|---|
| 0.551±0.0169 | 0.0866±0.000512 | 750.±3.77 | 36.2±0.133 | 8.93±0.0891 |

| badness | | α | β | k | τ | σ | quality |
|---|---|---|---|---|---|---|---|
| 0.57% | error | 3.1% | 0.59% | 0.5% | 0.37% | 1.% | 6.09% |

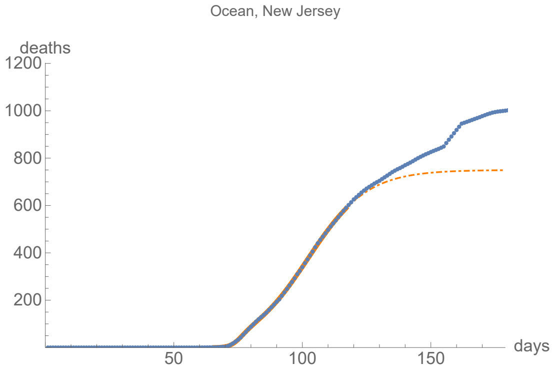


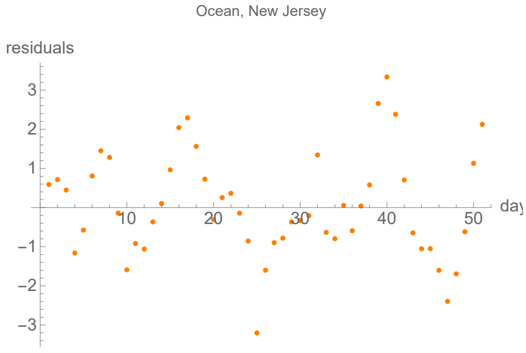


### Passaic, New Jersey

lag from integral overlap, using data averaged over 7 days, between 7 and 12 days, maximum overlap on day 8

lag from fits to the function I (averaged data): 41.9186 days

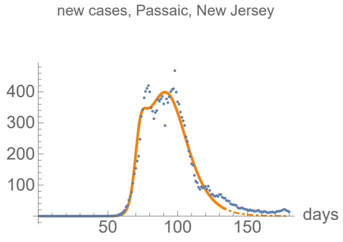


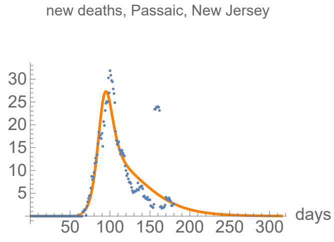


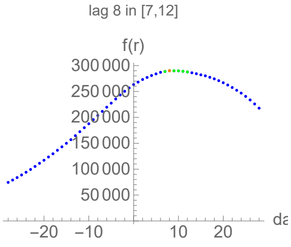


#### Parameters of fits to averaged cases data.

The best fit ends on Wed 3 Jun 2020.

| α | β | k | τ | σ |
|---|---|---|---|---|
| 0.449±0.0286 | 0.0978±0.00113 | 16200.±40.9 | 38.3±0.0843 | 18.6±0.259 |

| badness | | α | β | k | τ | σ | quality |
|---|---|---|---|---|---|---|---|
| 1.3% | error | 6.4% | 1.2% | 0.25% | 0.22% | 1.4% | 10.7% |

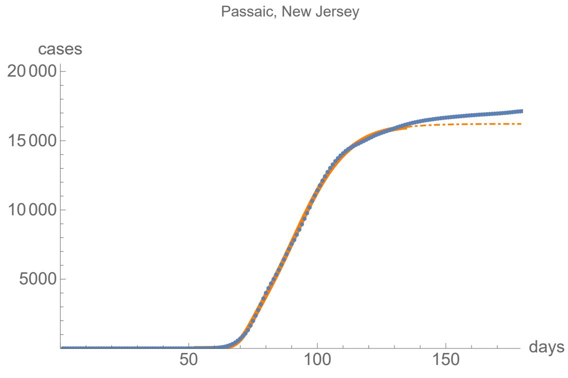


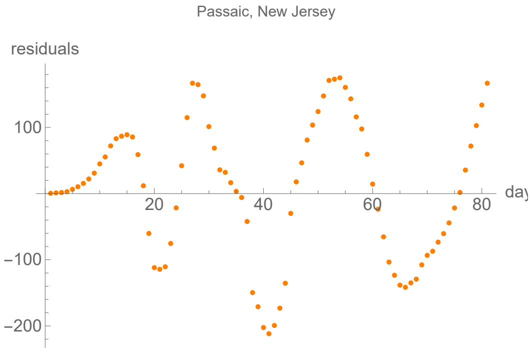


#### Parameters of fits to averaged death data.

The best fit ends on Tue 1 Dec 2020.

| α | β | k | τ | σ |
|---|---|---|---|---|
| 0.195±0.00973 | 0.0346±0.00102 | 1270.±2.83 | 37.2±0.981 | 25.7±0.505 |

| badness | | α | β | k | τ | σ | quality |
|---|---|---|---|---|---|---|---|
| 5.8% | error | 5.% | 3.% | 0.22% | 2.6% | 2.% | 18.6% |

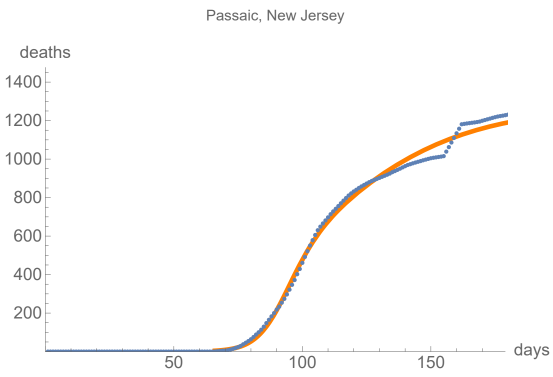


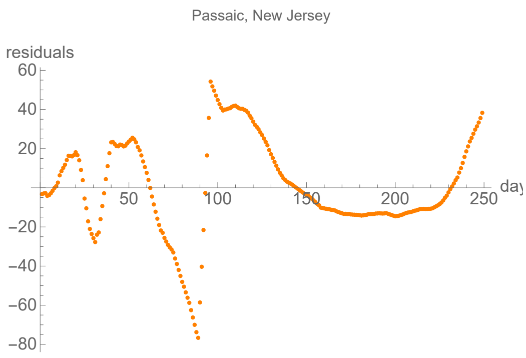


### Somerset, New Jersey

lag from integral overlap, using data averaged over 7 days, between 6 and 10 days, maximum overlap on day 8

lag from fits to the function I (averaged data): 7.75209 days

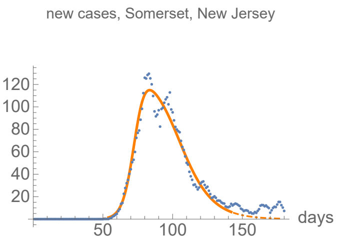


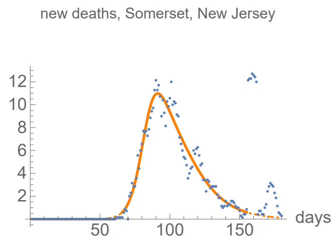


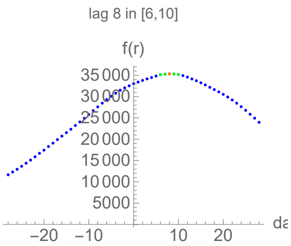


#### Parameters of fits to averaged cases data.

The best fit ends on Thu 11 Jun 2020.

| α | β | k | τ | σ |
|---|---|---|---|---|
| 0.267±0.00972 | 0.0777±0.0016 | 4720.±13.3 | 36.4±0.235 | 20.3±0.413 |

| badness | | α | β | k | τ | σ | quality |
|---|---|---|---|---|---|---|---|
| 1.2% | error | 3.6% | 2.1% | 0.28% | 0.65% | 2.% | 9.85% |

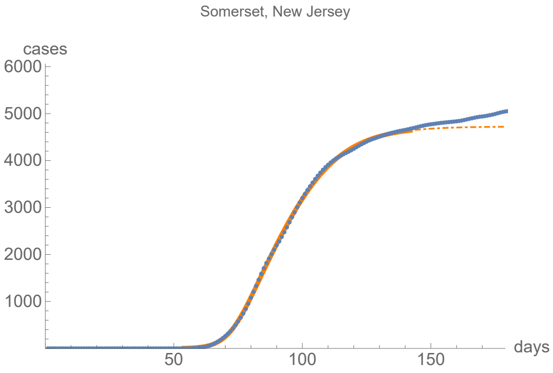


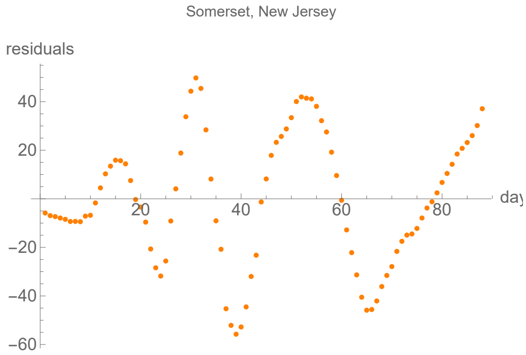


#### Parameters of fits to averaged death data.

The best fit ends on Wed 24 Jun 2020.

| α | β | k | τ | σ |
|---|---|---|---|---|
| 0.226±0.00658 | 0.0652±0.0018 | 450.±1.47 | 30.±0.441 | 18.±0.494 |

| badness | | α | β | k | τ | σ | quality |
|---|---|---|---|---|---|---|---|
| 1.5% | error | 2.9% | 2.8% | 0.33% | 1.5% | 2.8% | 11.7% |

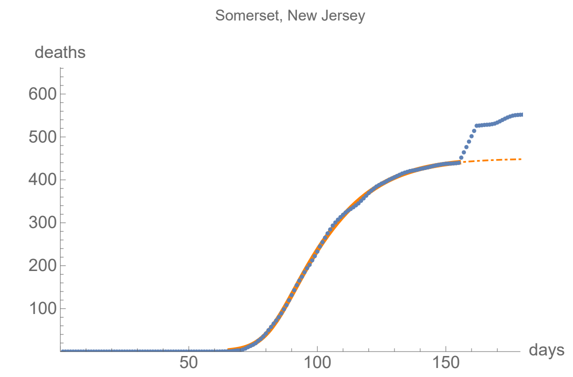


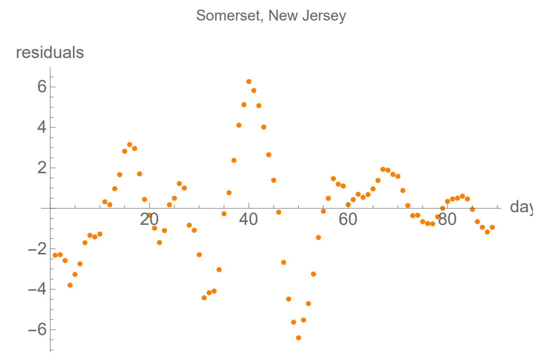


### Union, New Jersey

lag from integral overlap, using data averaged over 7 days, between 6 and 12 days, maximum overlap on day 9

lag from fits to the function I (averaged data): 10.757 days

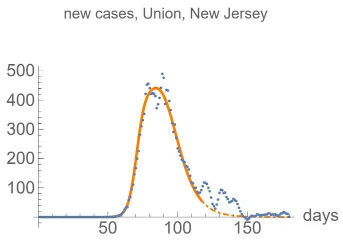


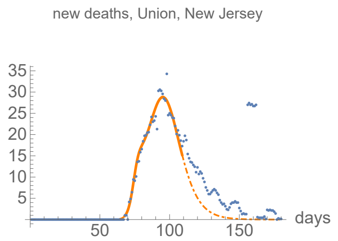


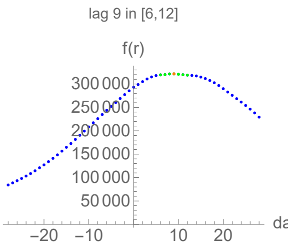


#### Parameters of fits to averaged cases data.

The best fit ends on Sun 17 May 2020.

| α | β | k | τ | σ |
|---|---|---|---|---|
| 0.37±0.0121 | 0.11±0.00149 | 14900.±40.4 | 34.2±0.0761 | 18.7±0.231 |

| badness | | α | β | k | τ | σ | quality |
|---|---|---|---|---|---|---|---|
| 0.92% | error | 3.3% | 1.4% | 0.27% | 0.22% | 1.2% | 7.27% |

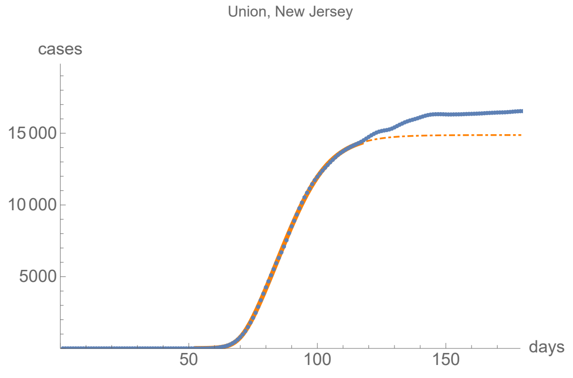


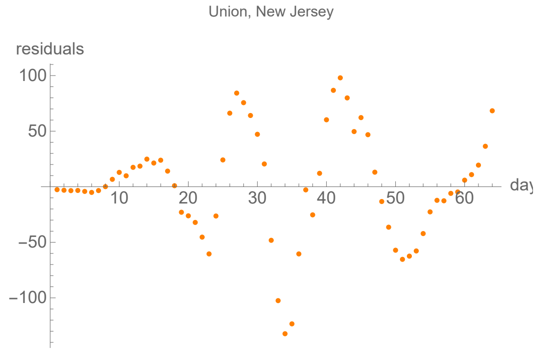


#### Parameters of fits to averaged death data.

The best fit ends on Sat 9 May 2020.

| α | β | k | τ | σ |
|---|---|---|---|---|
| 0.498±0.0209 | 0.122±0.00124 | 941.±5.57 | 27.3±0.108 | 7.35±0.159 |

| badness | | α | β | k | τ | σ | quality |
|---|---|---|---|---|---|---|---|
| 0.85% | error | 4.2% | 1.% | 0.59% | 0.4% | 2.2% | 9.21% |

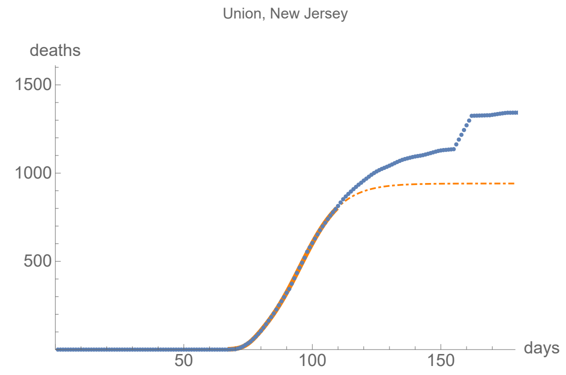


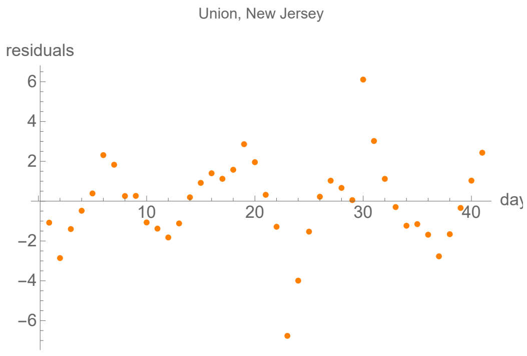


### Bronx, New York

```
lag from integral overlap, using data averaged over 7 days, between 0 and 4 days, maximum overlap on day 2

lag from fits to the function I (averaged data): -0.999217 days
```

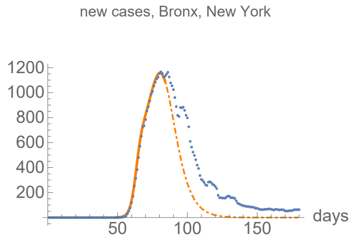


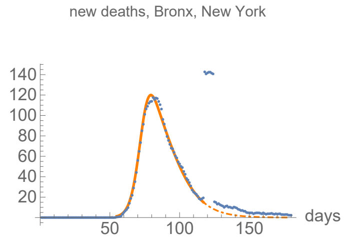


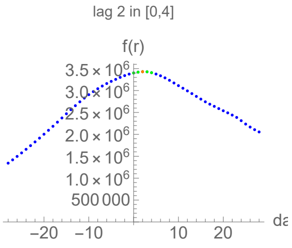


#### Parameters of fits to averaged cases data.

The best fit ends on Tue 14 Apr 2020.

| α | β | k | τ | σ |
|---|---|---|---|---|
| 0.502±0.00497 | 0.139±0.00104 | 32900.±251. | 31.7±0.111 | 15.1±0.0592 |

| badness | | α | β | k | τ | σ | quality |
|---|---|---|---|---|---|---|---|
| 0.11% | error | 0.99% | 0.75% | 0.76% | 0.35% | 0.39% | 3.35% |

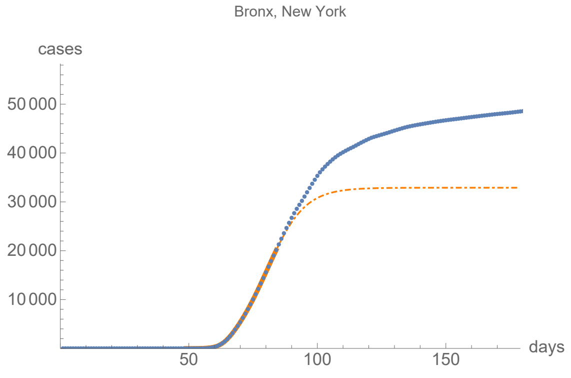


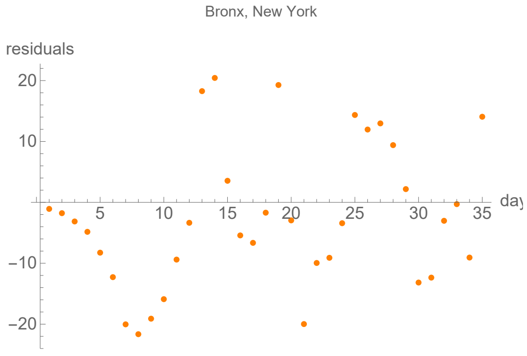


#### Parameters of fits to averaged death data.

The best fit ends on Sun 17 May 2020.

| α | β | k | τ | σ |
|---|---|---|---|---|
| 0.306±0.004 | 0.0867±0.0049 | 3650.±9.76 | 19.2±0.18 | 28.1±0.137 |

| badness | | α | β | k | τ | σ | quality |
|---|---|---|---|---|---|---|---|
| 0.43% | error | 1.3% | 5.7% | 0.27% | 0.94% | 0.49% | 4.77% |

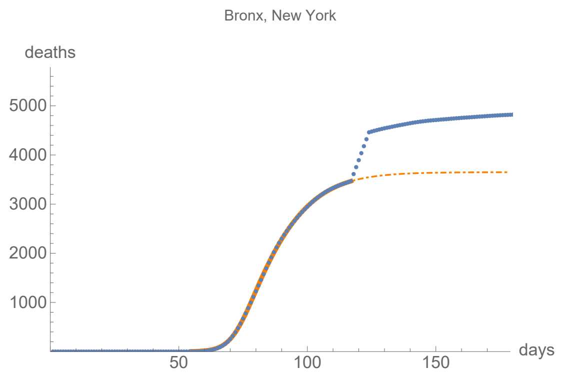


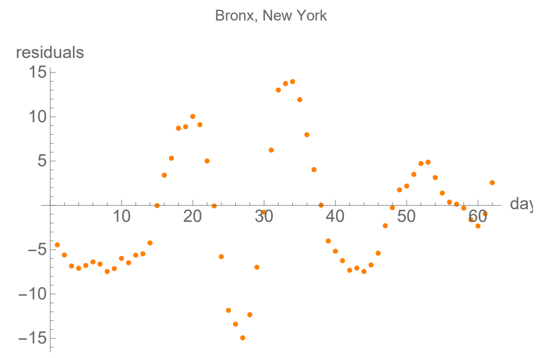


### Erie, New York

lag from integral overlap, using data averaged over 7 days, between 5 and 12 days, maximum overlap on day 10

lag from fits to the function I (averaged data): 20.463 days

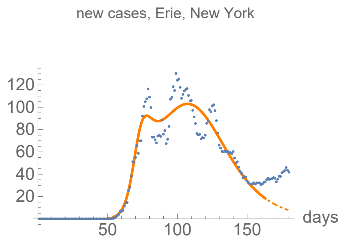


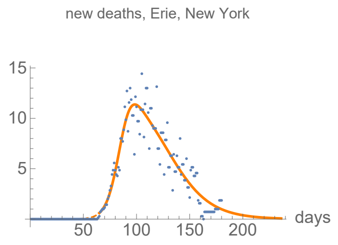


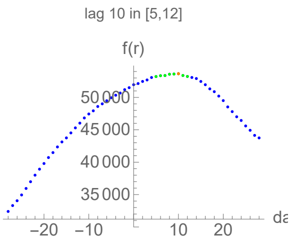


#### Parameters of fits to averaged cases data.

The best fit ends on Thu 2 Jul 2020.

| α | β | k | τ | σ |
|---|---|---|---|---|
| 0.279±0.0145 | 0.0545±0.000574 | 7550.±26.2 | 53.7±0.161 | 18.4±0.325 |

| badness | | α | β | k | τ | σ | quality |
|---|---|---|---|---|---|---|---|
| 1.6% | error | 5.2% | 1.1% | 0.35% | 0.3% | 1.8% | 10.3% |

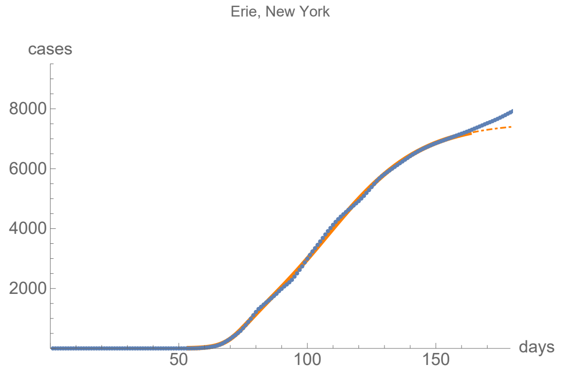


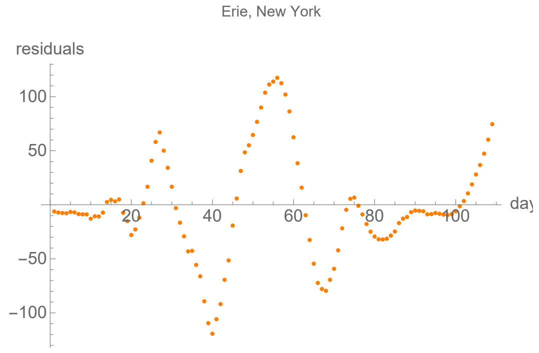


#### Parameters of fits to averaged death data.

The best fit ends on Sun 13 Sep 2020.

| α | β | k | τ | σ |
|---|---|---|---|---|
| 0.177±0.00385 | 0.0488±0.000537 | 678.±0.618 | 44.1±0.273 | 22.7±0.337 |

| badness | | α | β | k | τ | σ | quality |
|---|---|---|---|---|---|---|---|
| 1.6% | error | 2.2% | 1.1% | 0.091% | 0.62% | 1.5% | 7.04% |

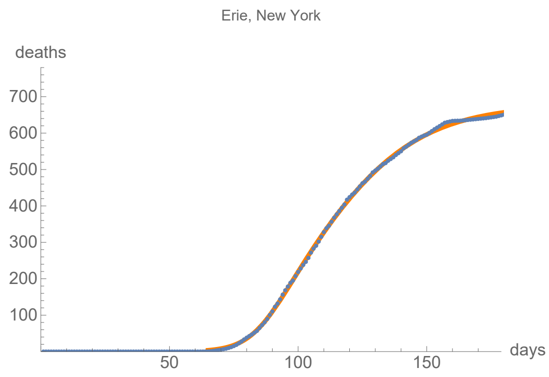


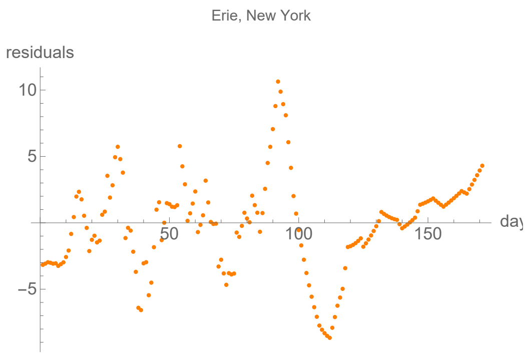


### Kings, New York

lag from integral overlap, using data averaged over 7 days, between 1 and 6 days, maximum overlap on day 3

lag from fits to the function I (averaged data): 5.82379 days

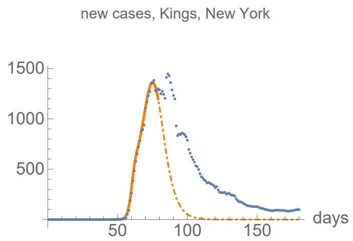


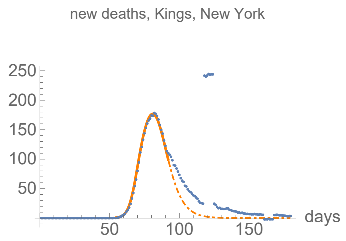


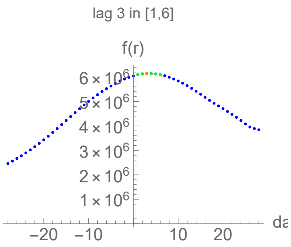


#### Parameters of fits to averaged cases data.

The best fit ends on Thu 9 Apr 2020.

| α | β | k | τ | σ |
|---|---|---|---|---|
| 0.712±0.0122 | 0.186±0.00116 | 29300.±182. | 29.1±0.0703 | 14.9±0.0502 |

| badness | | α | β | k | τ | σ | quality |
|---|---|---|---|---|---|---|---|
| 0.2% | error | 1.7% | 0.63% | 0.62% | 0.24% | 0.34% | 3.74% |

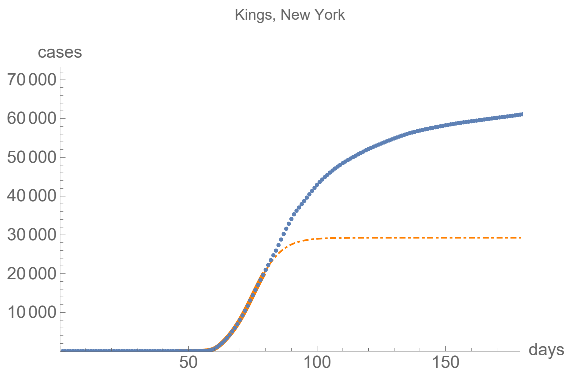


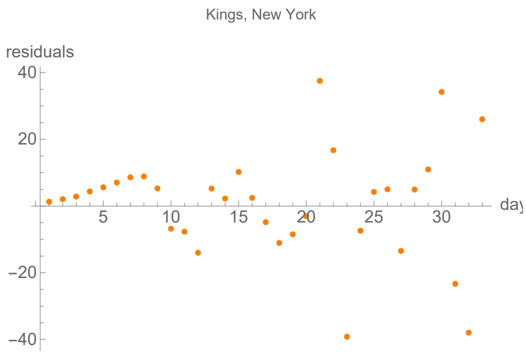


#### Parameters of fits to averaged death data.

The best fit ends on Wed 22 Apr 2020.

| α | β | k | τ | σ |
|---|---|---|---|---|
| 0.408±0.00428 | 0.148±0.00124 | 4400.±14. | 26.9±0.0285 | 13.9±0.128 |

| badness | | α | β | k | τ | σ | quality |
|---|---|---|---|---|---|---|---|
| 0.14% | error | 1.% | 0.84% | 0.32% | 0.11% | 0.92% | 3.37% |

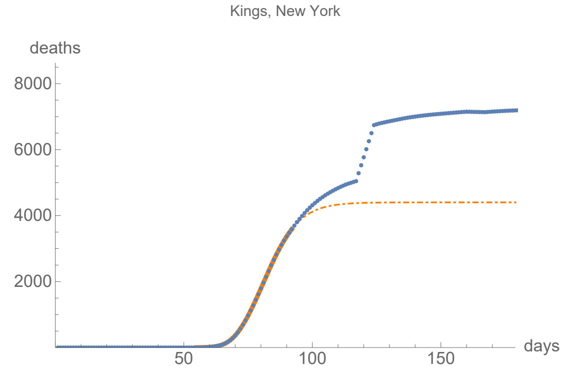


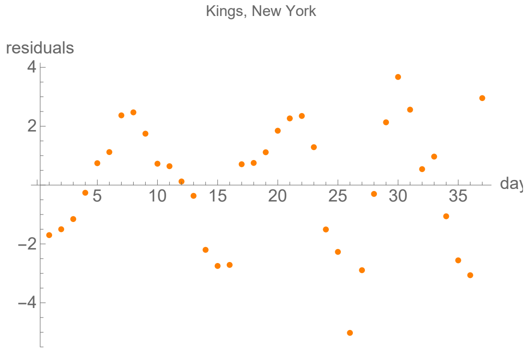


### Nassau, New York

lag from integral overlap, using data averaged over 7 days, between 5 and 8 days, maximum overlap on day 6

lag from fits to the function I (averaged data): 1.68729 days

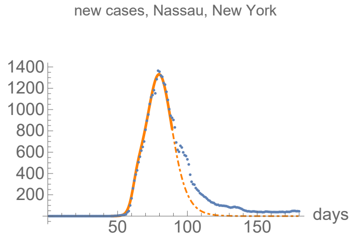


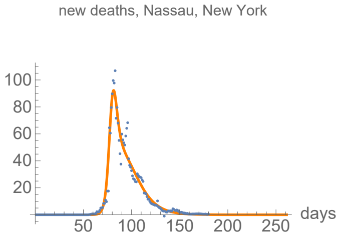


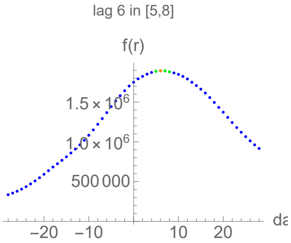


#### Parameters of fits to averaged cases data.

The best fit ends on Sun 19 Apr 2020.

| α | β | k | τ | σ |
|---|---|---|---|---|
| 0.541±0.0134 | 0.157±0.000906 | 33900.±124. | 33.8±0.0507 | 15.9±0.0943 |

| badness | | α | β | k | τ | σ | quality |
|---|---|---|---|---|---|---|---|
| 0.27% | error | 2.5% | 0.58% | 0.37% | 0.15% | 0.59% | 4.43% |

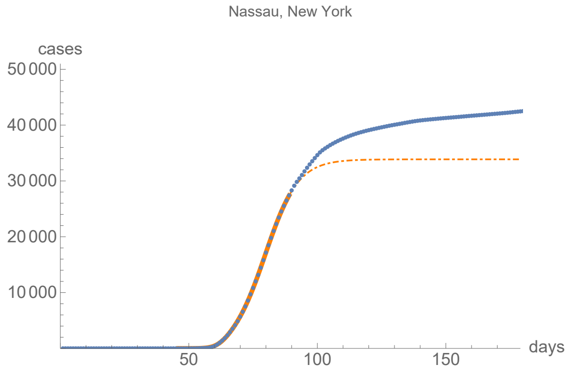


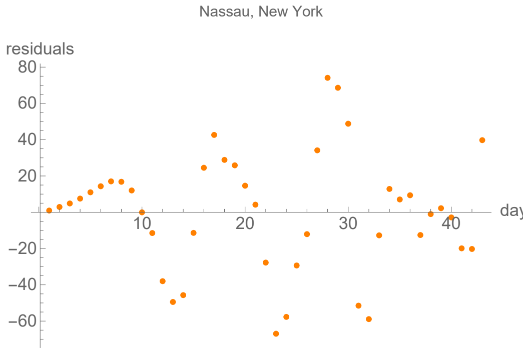


#### Parameters of fits to averaged death data.

The best fit ends on Thu 8 Oct 2020.

| α | β | k | τ | σ |
|---|---|---|---|---|
| 0.441±0.0105 | 0.0822±0.000781 | 2190.±1.11 | 30.1±0.136 | 21.6±0.103 |

| badness | | α | β | k | τ | σ | quality |
|---|---|---|---|---|---|---|---|
| 1.9% | error | 2.4% | 0.95% | 0.051% | 0.45% | 0.48% | 6.2% |

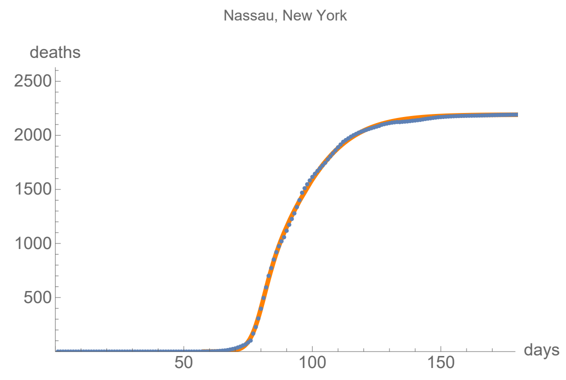


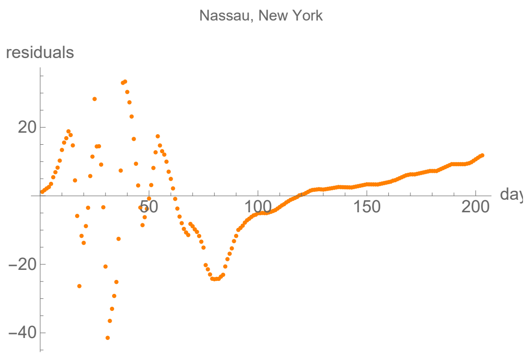


### New York, New York

lag from integral overlap, using data averaged over 7 days, between 3 and 8 days, maximum overlap on day 5

lag from fits to the function I (averaged data): 1.14256 days

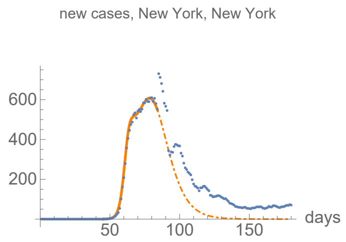


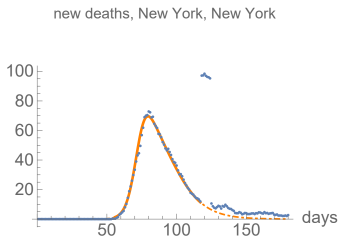


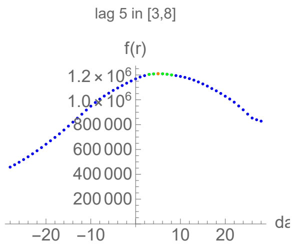


#### Parameters of fits to averaged cases data.

The best fit ends on Tue 14 Apr 2020.

| α | β | k | τ | σ |
|---|---|---|---|---|
| 0.513±0.00465 | 0.117±0.000892 | 20500.±149. | 33.7±0.127 | 17.1±0.045 |

| badness | | α | β | k | τ | σ | quality |
|---|---|---|---|---|---|---|---|
| 0.17% | error | 0.91% | 0.76% | 0.72% | 0.38% | 0.26% | 3.2% |

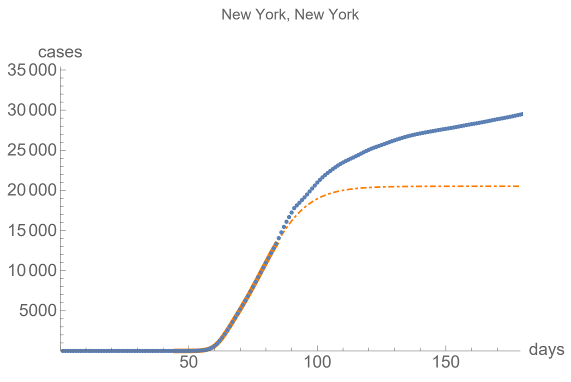


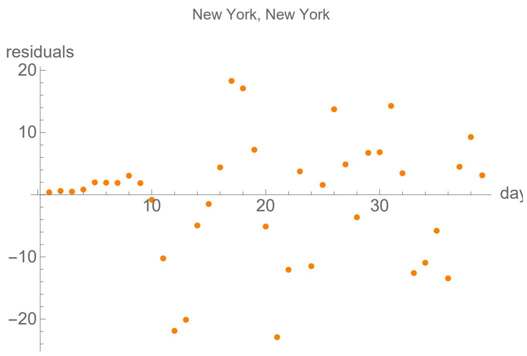


#### Parameters of fits to averaged death data.

The best fit ends on Sun 17 May 2020.

| α | β | k | τ | σ |
|---|---|---|---|---|
| 0.293±0.00373 | 0.0825±0.0046 | 2320.±6.7 | 17.5±0.175 | 28.4±0.106 |

| badness | | α | β | k | τ | σ | quality |
|---|---|---|---|---|---|---|---|
| 0.4% | error | 1.3% | 5.6% | 0.29% | 1.% | 0.37% | 4.66% |

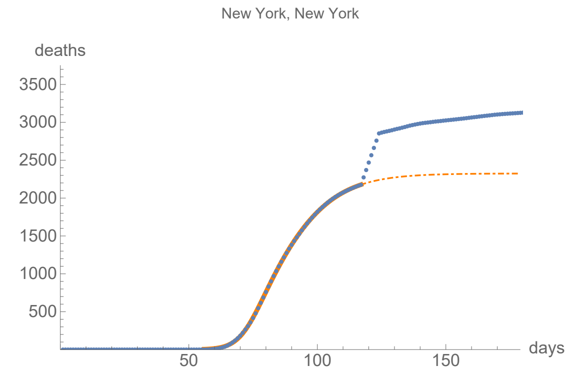


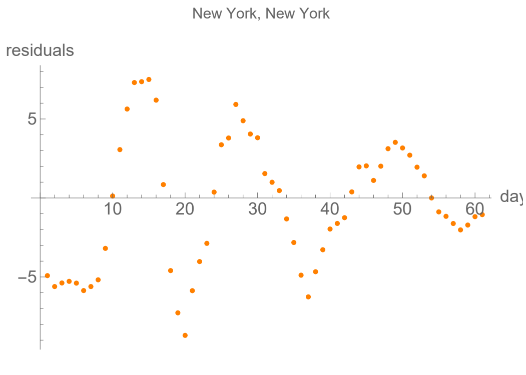


### Queens, New York

lag from integral overlap, using data averaged over 7 days, between 2 and 7 days, maximum overlap on day 5

lag from fits to the function I (averaged data): 2.65965 days

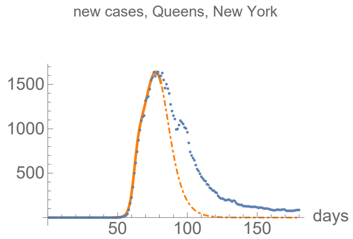


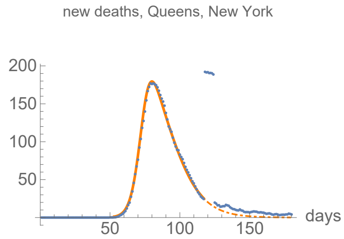


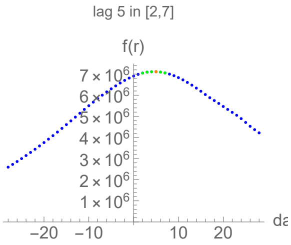


#### Parameters of fits to averaged cases data.

The best fit ends on Sat 11 Apr 2020.

| α | β | k | τ | σ |
|---|---|---|---|---|
| 0.575±0.0065 | 0.156±0.00122 | 41700.±321. | 30.6±0.1 | 15.8±0.0549 |

| badness | | α | β | k | τ | σ | quality |
|---|---|---|---|---|---|---|---|
| 0.16% | error | 1.1% | 0.78% | 0.77% | 0.33% | 0.35% | 3.52% |

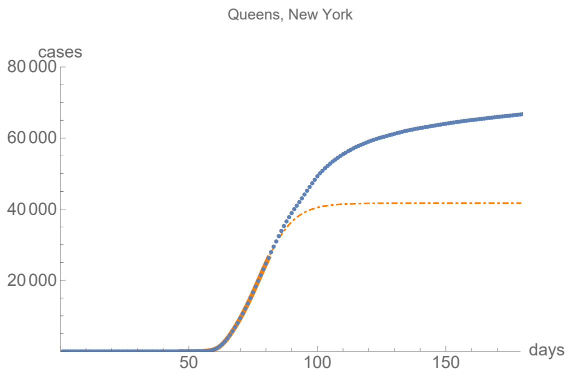


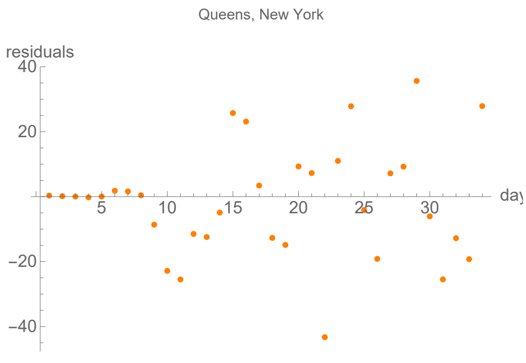


#### Parameters of fits to averaged death data.

The best fit ends on Sun 17 May 2020.

| α | β | k | τ | σ |
|---|---|---|---|---|
| 0.295±0.00252 | 0.0847±0.00305 | 5640.±10.7 | 21.4±0.126 | 30.7±0.0906 |

| badness | | α | β | k | τ | σ | quality |
|---|---|---|---|---|---|---|---|
| 0.24% | error | 0.85% | 3.6% | 0.19% | 0.59% | 0.3% | 3.02% |

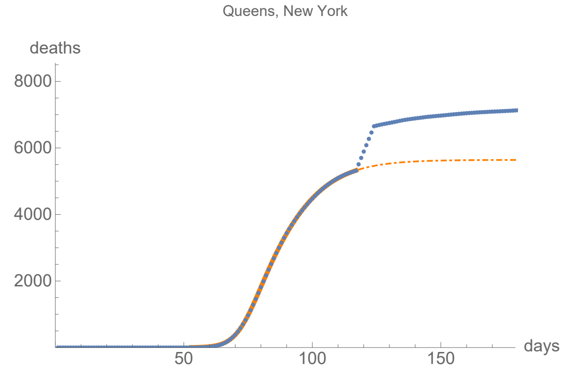


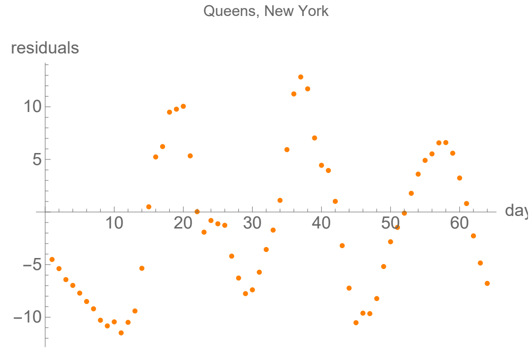


### Richmond, New York

lag from integral overlap, using data averaged over 7 days, between -1 and 4 days, maximum overlap on day 2

lag from fits to the function I (averaged data): -3.52476 days

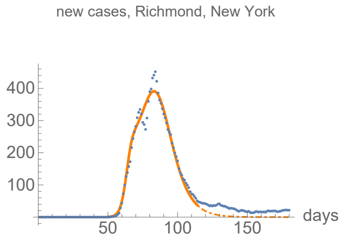


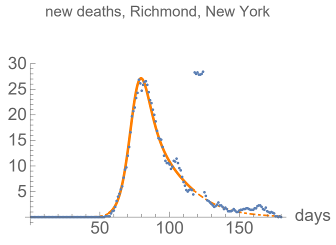


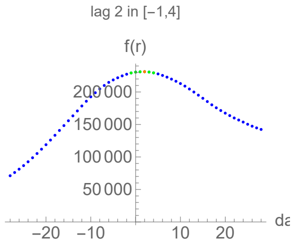


#### Parameters of fits to averaged cases data.

The best fit ends on Thu 14 May 2020.

| α | β | k | τ | σ |
|---|---|---|---|---|
| 0.471±0.0236 | 0.12±0.00104 | 13000.±27.7 | 32.3±0.0512 | 13.7±0.208 |

| badness | | α | β | k | τ | σ | quality |
|---|---|---|---|---|---|---|---|
| 0.95% | error | 5.% | 0.87% | 0.21% | 0.16% | 1.5% | 8.71% |

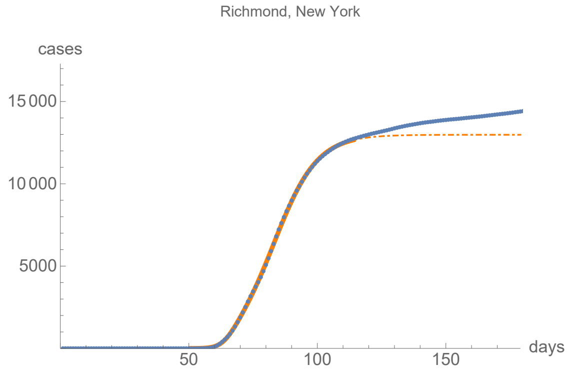


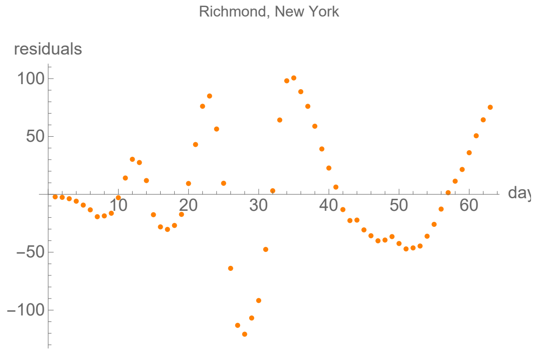


#### Parameters of fits to averaged death data.

The best fit ends on Sun 17 May 2020.

| α | β | k | τ | σ |
|---|---|---|---|---|
| 0.256±0.00445 | 0.0572±0.00489 | 890.±10.3 | 19.3±0.254 | 24.6±0.256 |

| badness | | α | β | k | τ | σ | quality |
|---|---|---|---|---|---|---|---|
| 0.69% | error | 1.7% | 8.6% | 1.2% | 1.3% | 1.% | 7.18% |

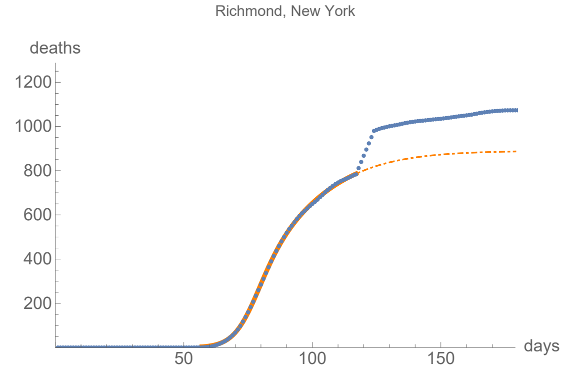


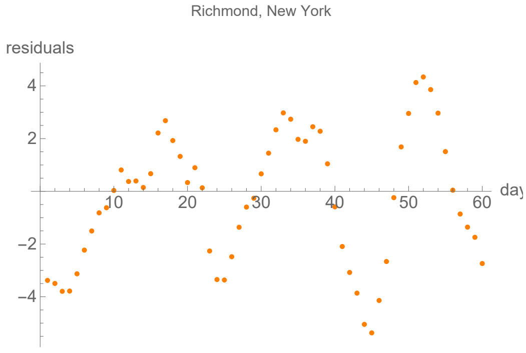


### Rockland, New York

lag from integral overlap, using data averaged over 7 days, between 7 and 10 days, maximum overlap on day 8

lag from fits to the function I (averaged data): 3.1231 days

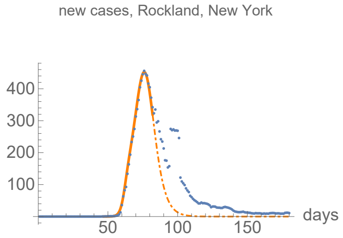


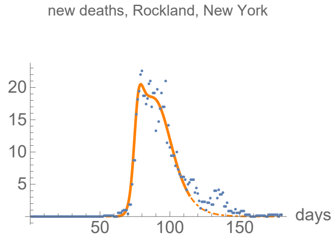


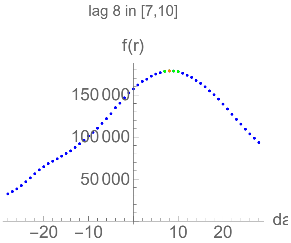


#### Parameters of fits to averaged cases data.

The best fit ends on Sun 12 Apr 2020.

| α | β | k | τ | σ |
|---|---|---|---|---|
| 0.654±0.012 | 0.205±0.00111 | 8760.±31.5 | 29.2±0.0361 | 15.4±0.0634 |

| badness | | α | β | k | τ | σ | quality |
|---|---|---|---|---|---|---|---|
| 0.17% | error | 1.8% | 0.54% | 0.36% | 0.12% | 0.41% | 3.43% |

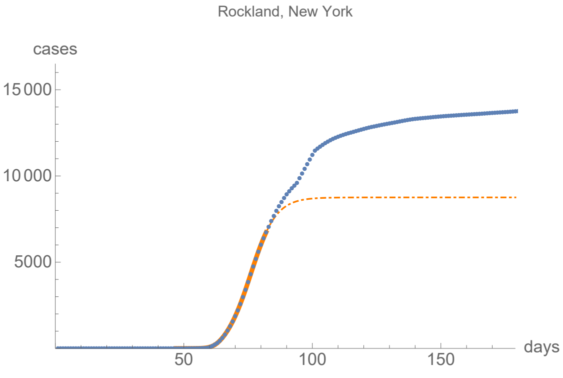


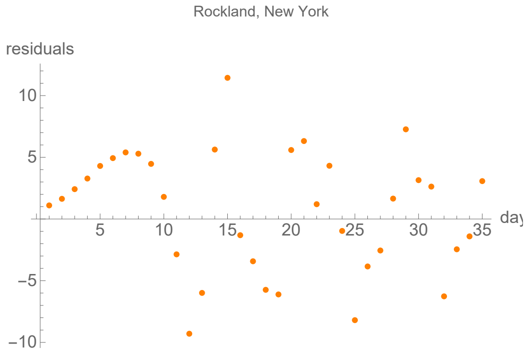


#### Parameters of fits to averaged death data.

The best fit ends on Thu 14 May 2020.

| α | β | k | τ | σ |
|---|---|---|---|---|
| 0.548±0.021 | 0.119±0.00168 | 601.±2.32 | 27.±0.0709 | 14.2±0.137 |

| badness | | α | β | k | τ | σ | quality |
|---|---|---|---|---|---|---|---|
| 1.1% | error | 3.8% | 1.4% | 0.39% | 0.26% | 0.96% | 7.96% |

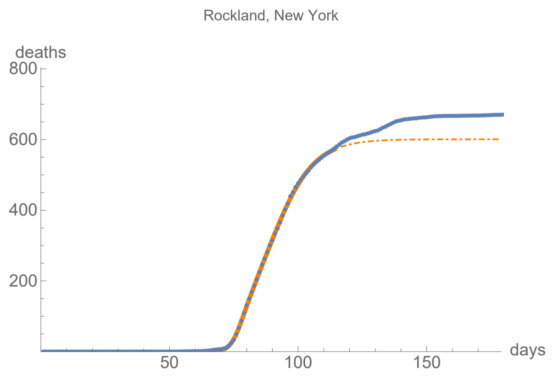


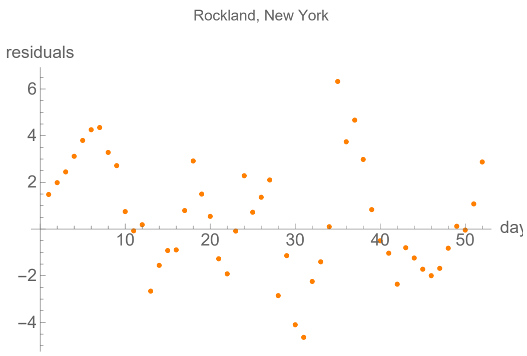


### Suffolk, New York

lag from integral overlap, using data averaged over 7 days, between 6 and 13 days, maximum overlap on day 11

lag from fits to the function I (averaged data): 0.186019 days

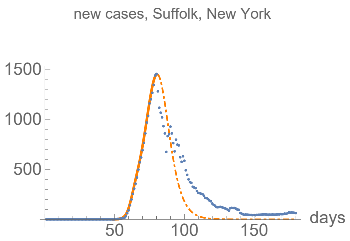


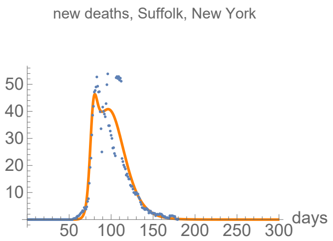


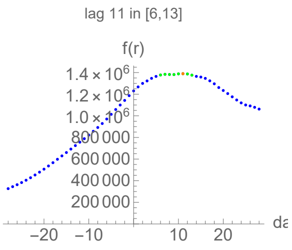


#### Parameters of fits to averaged cases data.

The best fit ends on Fri 10 Apr 2020.

| α | β | k | τ | σ |
|---|---|---|---|---|
| 0.555±0.0108 | 0.175±0.0017 | 33000.±526. | 31.5±0.173 | 13.4±0.0972 |

| badness | | α | β | k | τ | σ | quality |
|---|---|---|---|---|---|---|---|
| 0.19% | error | 1.9% | 0.97% | 1.6% | 0.55% | 0.72% | 5.96% |

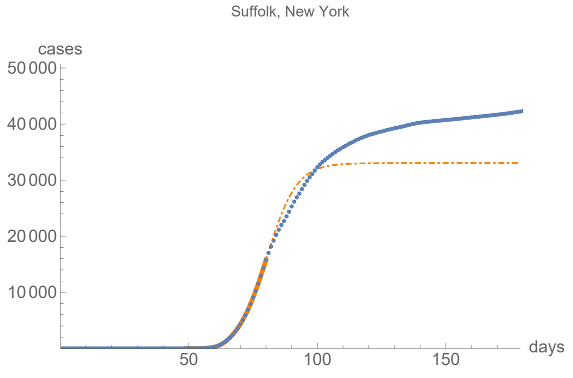


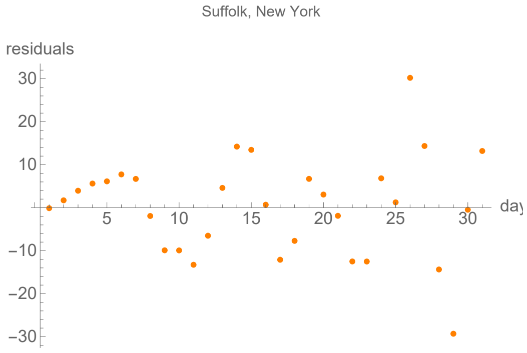


#### Parameters of fits to averaged death data.

The best fit ends on Sun 15 Nov 2020.

| α | β | k | τ | σ |
|---|---|---|---|---|
| 0.42±0.0244 | 0.0806±0.00072 | 2000.±1.33 | 41.4±0.104 | 21.6±0.241 |

| badness | | α | β | k | τ | σ | quality |
|---|---|---|---|---|---|---|---|
| 3.5% | error | 5.8% | 0.89% | 0.066% | 0.25% | 1.1% | 11.7% |

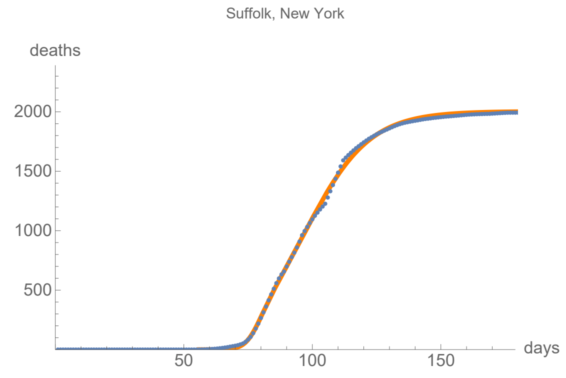


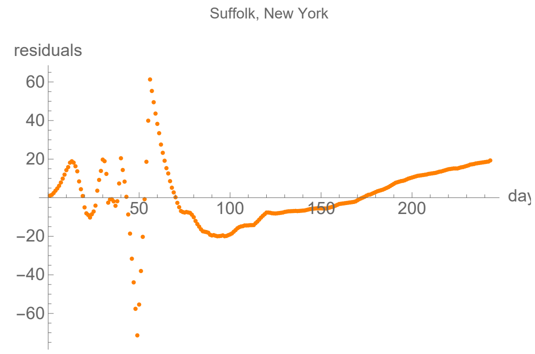


### Westchester, New York

```
lag from integral overlap, using data averaged over 7 days, between 12 and 14 days, maximum overlap on day 13

lag from fits to the function I (averaged data): 13.9184 days
```

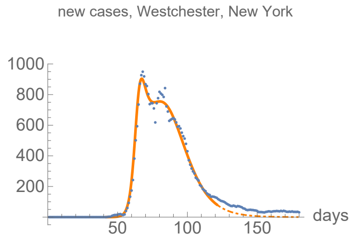


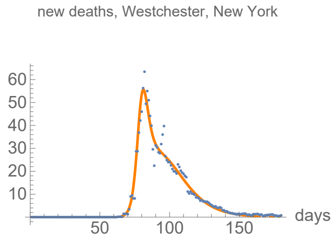


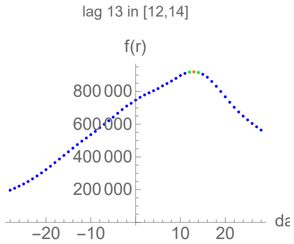


#### Parameters of fits to averaged cases data.

The best fit ends on Thu 21 May 2020.

| α | β | k | τ | σ |
|---|---|---|---|---|
| 0.472±0.00843 | 0.0899±0.000437 | 33100.±39.7 | 39.7±0.0354 | 22.7±0.0666 |

| badness | | α | β | k | τ | σ | quality |
|---|---|---|---|---|---|---|---|
| 0.5% | error | 1.8% | 0.49% | 0.12% | 0.089% | 0.29% | 3.27% |

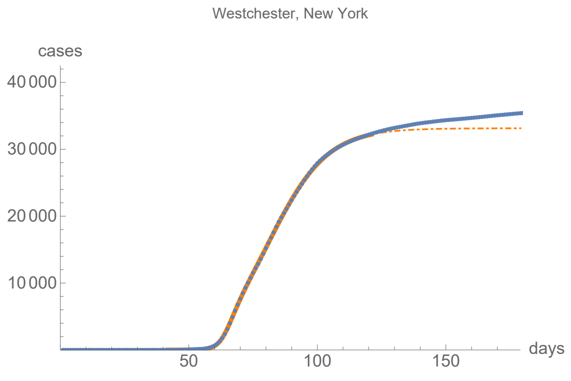


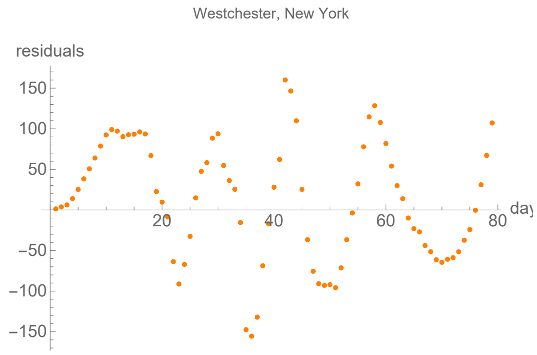


#### Parameters of fits to averaged death data.

The best fit ends on Tue 30 Jun 2020.

| α | β | k | τ | σ |
|---|---|---|---|---|
| 0.459±0.00873 | 0.0778±0.000573 | 1420.±1.33 | 24.6±0.0754 | 13.6±0.0735 |

| badness | | α | β | k | τ | σ | quality |
|---|---|---|---|---|---|---|---|
| 1.% | error | 1.9% | 0.74% | 0.094% | 0.31% | 0.54% | 4.58% |

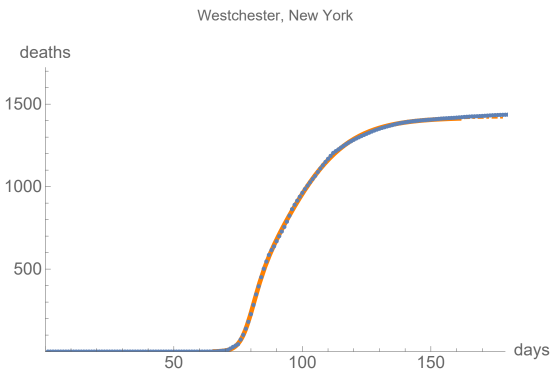


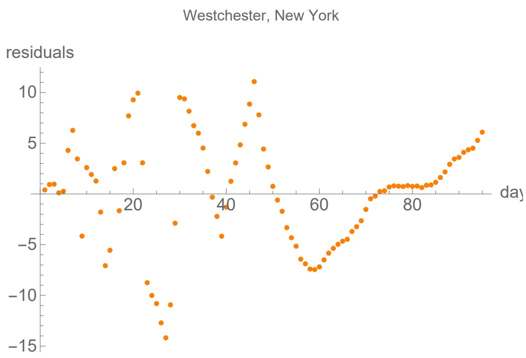


### Bucks, Pennsylvania

lag from integral overlap, using data averaged over 7 days, between 7 and 13 days, maximum overlap on day 11

lag from fits to the function I (averaged data): 4.31689 days

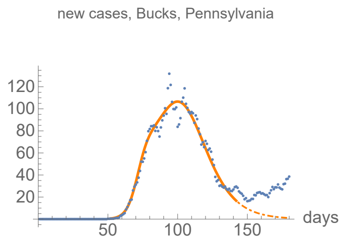


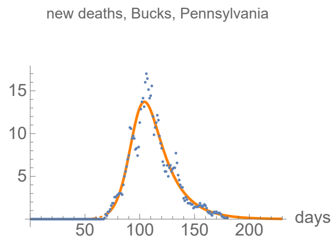


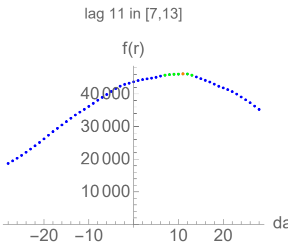


#### Parameters of fits to averaged cases data.

The best fit ends on Thu 11 Jun 2020.

| $\alpha$ | $\beta$ | k | $\tau$ | $\sigma$ |
|---|---|---|---|---|
| 0.274±0.00761 | 0.0763±0.000525 | 5490.±9.95 | 48.9±0.0564 | 21.±0.211 |

| badness | | $\alpha$ | $\beta$ | k | $\tau$ | $\sigma$ | quality |
|---|---|---|---|---|---|---|---|
| 0.83% | error | 2.8% | 0.69% | 0.18% | 0.12% | 1.% | 5.6% |

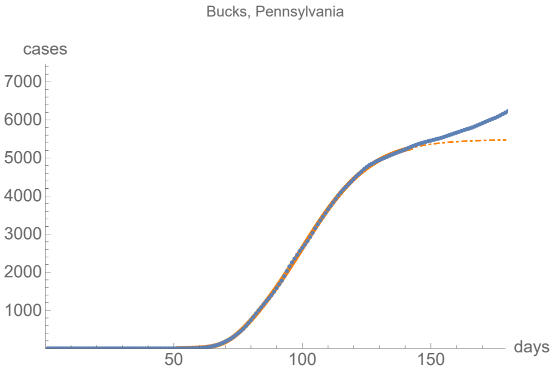


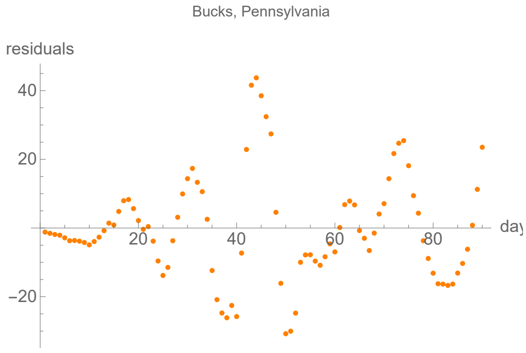


#### Parameters of fits to averaged death data.

The best fit ends on Sun 6 Sep 2020.

| $\alpha$ | $\beta$ | k | $\tau$ | $\sigma$ |
|---|---|---|---|---|
| 0.169±0.00337 | 0.0535±0.00388 | 582.±0.663 | 29.1±0.713 | 29.1±1.41 |

| badness | | $\alpha$ | $\beta$ | k | $\tau$ | $\sigma$ | quality |
|---|---|---|---|---|---|---|---|
| 1.7% | error | 2.% | 7.3% | 0.11% | 2.5% | 4.8% | 12.9% |

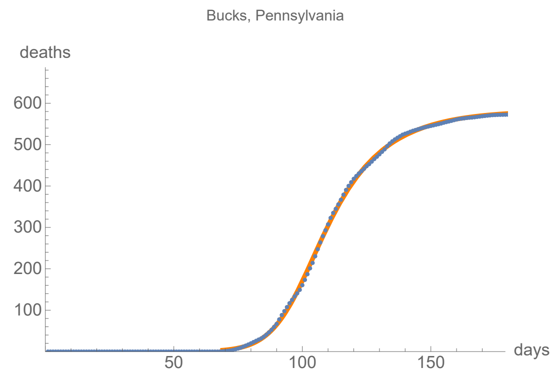


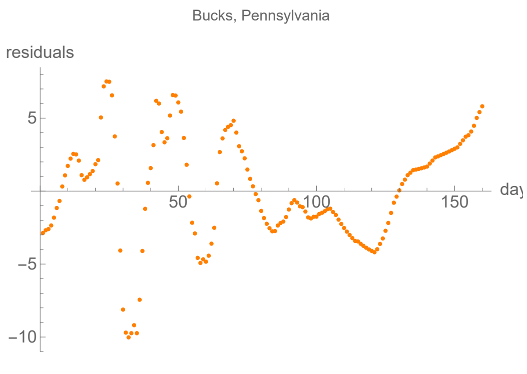


### Delaware, Pennsylvania

```
lag from integral overlap, using data averaged over 7 days, between 7 and 16 days, maximum overlap on day 12

lag from fits to the function I (averaged data): 18.386 days
```

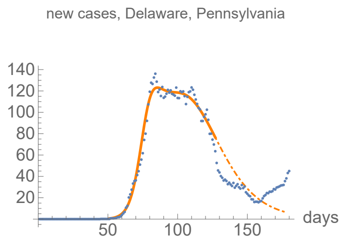


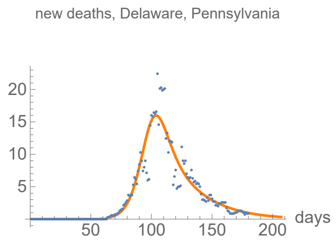


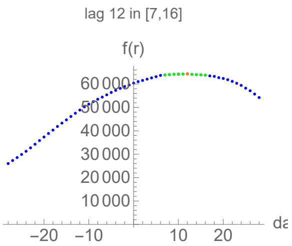


#### Parameters of fits to averaged cases data.

The best fit ends on Wed 27 May 2020.

| α | β | k | τ | σ |
|---|---|---|---|---|
| 0.247±0.00436 | 0.0578±0.00086 | 7870.±67.5 | 53.5±0.292 | 26.±0.189 |

| badness | | α | β | k | τ | σ | quality |
|---|---|---|---|---|---|---|---|
| 0.6% | error | 1.8% | 1.5% | 0.86% | 0.55% | 0.72% | 5.98% |

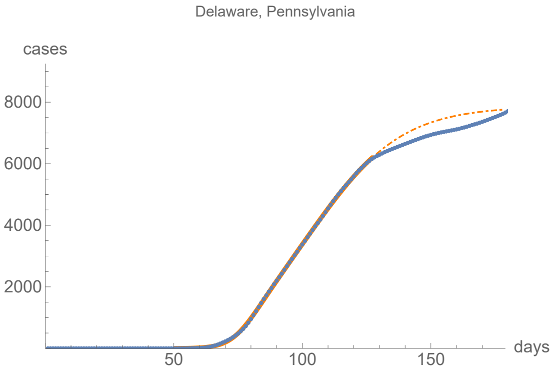


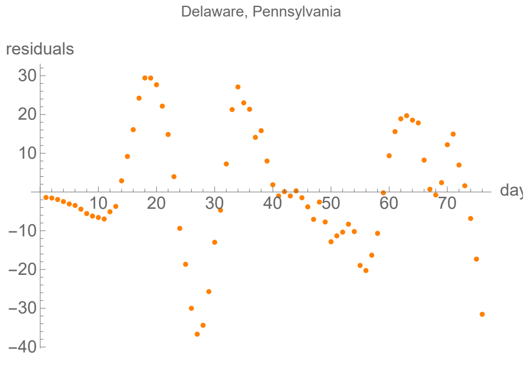


#### Parameters of fits to averaged death data.

The best fit ends on Sat 15 Aug 2020.

| α | β | k | τ | σ |
|---|---|---|---|---|
| 0.171±0.00484 | 0.0416±0.00585 | 700.±2.63 | 33.4±0.616 | 33.4±1.56 |

| badness | | α | β | k | τ | σ | quality |
|---|---|---|---|---|---|---|---|
| 2.4% | error | 2.8% | 14.% | 0.38% | 1.8% | 4.7% | 14.8% |

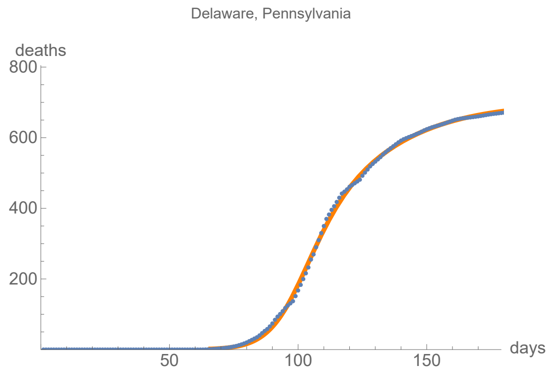


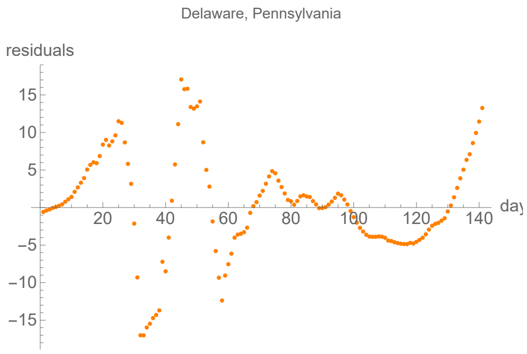


### Montgomery, Pennsylvania

lag from integral overlap, using data averaged over 7 days, between 8 and 12 days, maximum overlap on day 10

lag from fits to the function I (averaged data): 17.4783 days

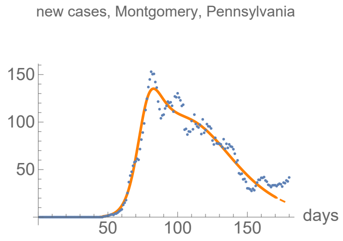


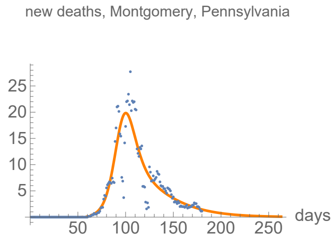


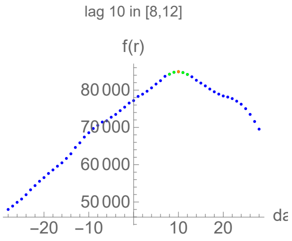


#### Parameters of fits to averaged cases data.

The best fit ends on Thu 9 Jul 2020.

| α | β | k | τ | σ |
|---|---|---|---|---|
| 0.213±0.00369 | 0.0442±0.000372 | 9100.±21.2 | 59.±0.105 | 29.5±0.166 |

| badness | | α | β | k | τ | σ | quality |
|---|---|---|---|---|---|---|---|
| 0.82% | error | 1.7% | 0.84% | 0.23% | 0.18% | 0.56% | 4.37% |

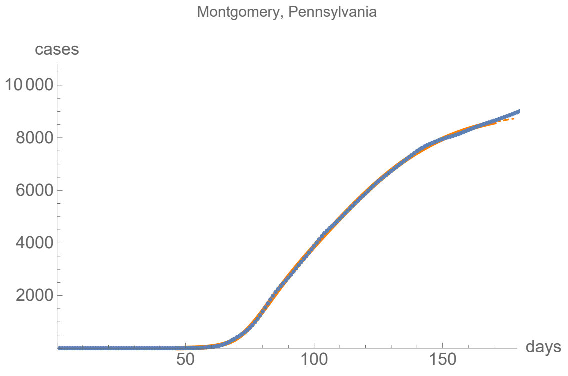


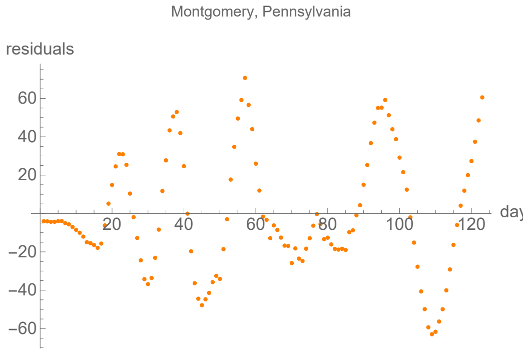


#### Parameters of fits to averaged death data.

The best fit ends on Sat 10 Oct 2020.

| α | β | k | τ | σ |
|---|---|---|---|---|
| 0.173±0.00379 | 0.0357±0.00494 | 876.±1.46 | 32.6±0.344 | 32.6±1.02 |

| badness | | α | β | k | τ | σ | quality |
|---|---|---|---|---|---|---|---|
| 3.1% | error | 2.2% | 14.% | 0.17% | 1.1% | 3.1% | 12.% |

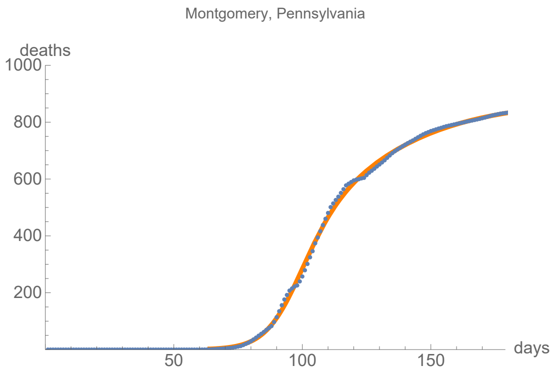


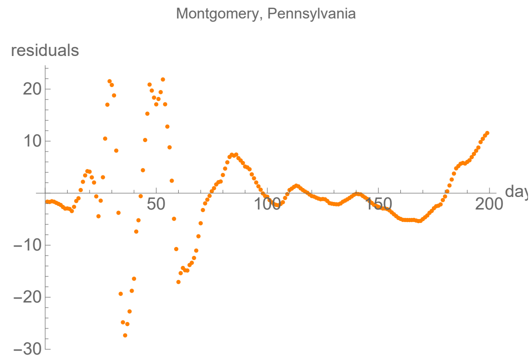


### Philadelphia, Pennsylvania

lag from integral overlap, using data averaged over 7 days, between 8 and 12 days, maximum overlap on day 10

lag from fits to the function I (averaged data): −3.30921 days

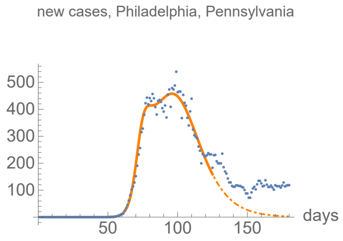


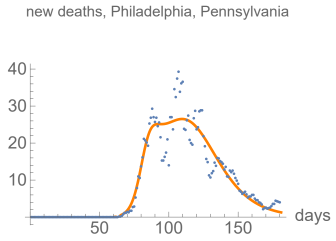


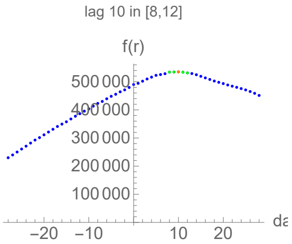


#### Parameters of fits to averaged cases data.

The best fit ends on Mon 25 May 2020.

| α | β | k | τ | σ |
|---|---|---|---|---|
| 0.35±0.00916 | 0.0788±0.0007 | 23000.±83. | 44.7±0.0972 | 21.1±0.145 |

| badness | | α | β | k | τ | σ | quality |
|---|---|---|---|---|---|---|---|
| 0.68% | error | 2.6% | 0.89% | 0.36% | 0.22% | 0.69% | 5.45% |

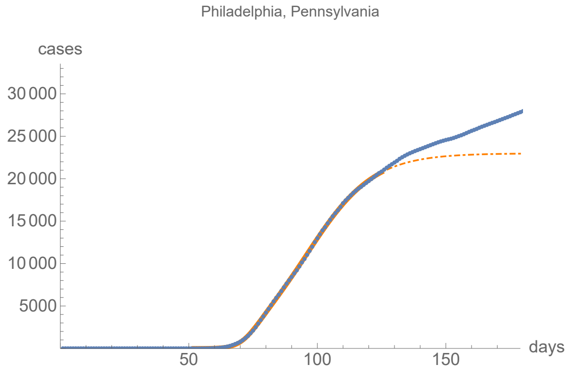


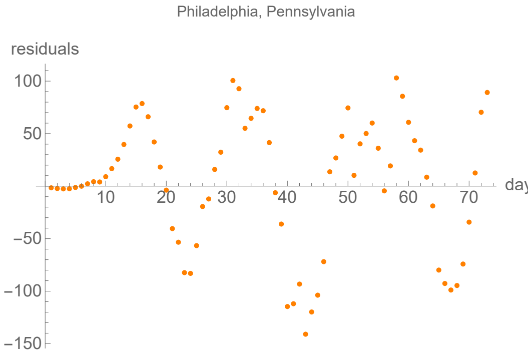


#### Parameters of fits to averaged death data.

The best fit ends on Mon 20 Jul 2020.

| α | β | k | τ | σ |
|---|---|---|---|---|
| 0.273±0.0174 | 0.0628±0.00089 | 1660.±4.51 | 45.3±0.154 | 17.±0.441 |

| badness | | α | β | k | τ | σ | quality |
|---|---|---|---|---|---|---|---|
| 2.4% | error | 6.4% | 1.4% | 0.27% | 0.34% | 2.6% | 13.4% |

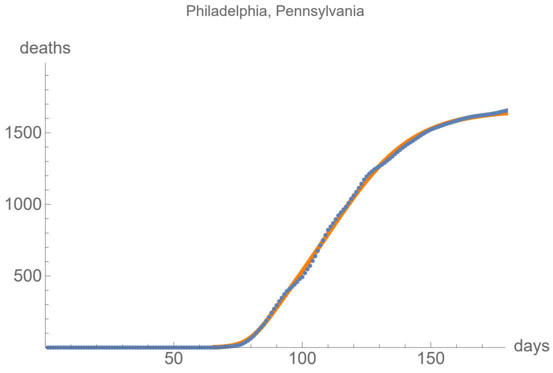


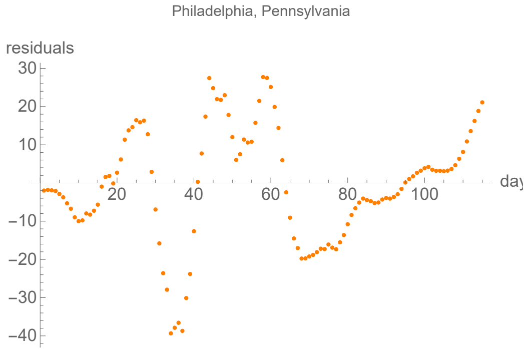


### Providence, Rhode Island

lag from integral overlap, using data averaged over 7 days, between 10 and 10 days, maximum overlap on day 10

lag from fits to the function I (averaged data): 3.85265 days

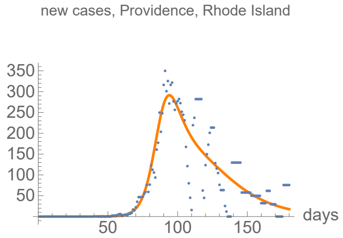


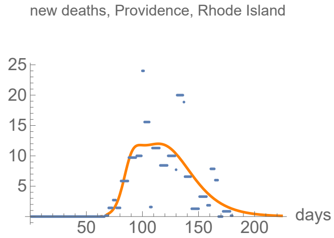


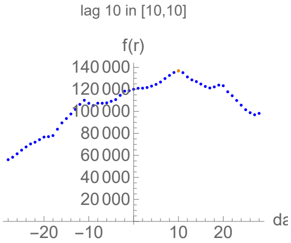


#### Parameters of fits to averaged cases data.

The best fit ends on Sun 19 Jul 2020.

| α | β | k | τ | σ |
|---|---|---|---|---|
| 0.21±0.00743 | 0.0462±0.00146 | 13700.±69.4 | 60.7±0.462 | 46.2±0.445 |

| badness | | α | β | k | τ | σ | quality |
|---|---|---|---|---|---|---|---|
| 3.4% | error | 3.5% | 3.2% | 0.51% | 0.76% | 0.96% | 12.3% |

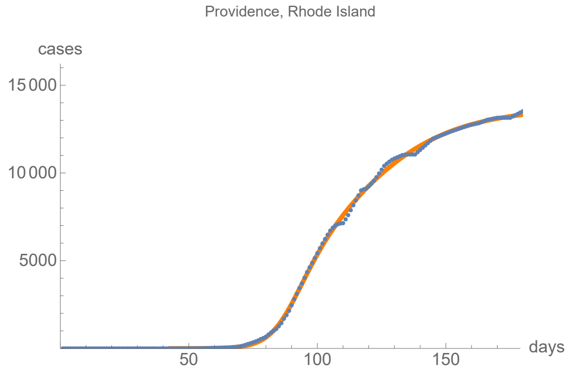


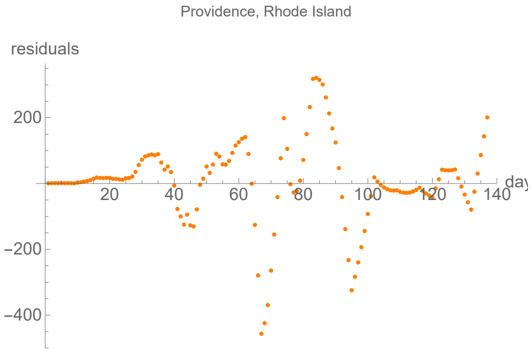


#### Parameters of fits to averaged death data.

The best fit ends on Tue 1 Sep 2020.

| α | β | k | τ | σ |
|---|---|---|---|---|
| 0.239±0.025 | 0.0565±0.00125 | 826.±2.45 | 48.3±0.315 | 18.2±0.834 |

| badness | | α | β | k | τ | σ | quality |
|---|---|---|---|---|---|---|---|
| 4.8% | error | 10.% | 2.2% | 0.3% | 0.65% | 4.6% | 23.% |

Providence, Rhode Island

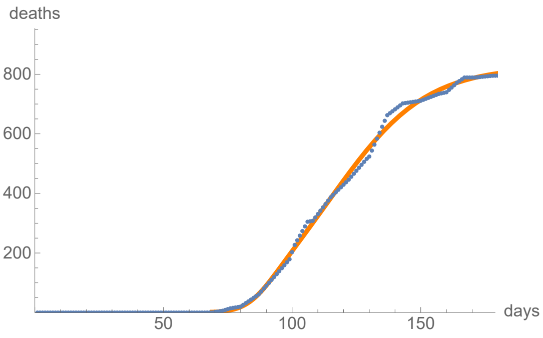


Providence, Rhode Island

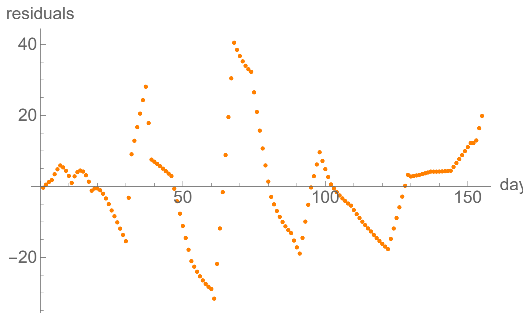


### Harris, Texas

no clear lag in the first six months of the pandemic from integral overlap using data averaged over 7 days

lag from fits to the function I (averaged data): 8.16663 days

new cases, Harris, Texas

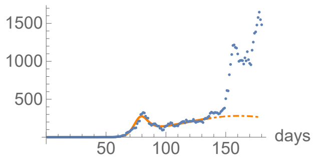


new deaths, Harris, Texas

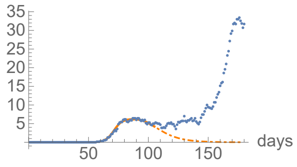


no clear lag in the first six months of the pandemic

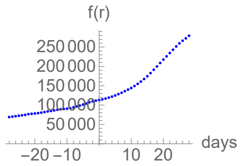


#### Parameters of fits to averaged cases data.

The best fit ends on Sat 6 Jun 2020.

| α | β | k | τ | σ |
|---|---|---|---|---|
| 0.288±0.00647 | 0.0285±0.000644 | 39500.±2920. | 116.±4.37 | 32.9±0.121 |

| badness | | α | β | k | τ | σ | quality |
|---|---|---|---|---|---|---|---|
| 1.% | error | 2.2% | 2.3% | 7.4% | 3.8% | 0.37% | 17.1% |

Harris, Texas

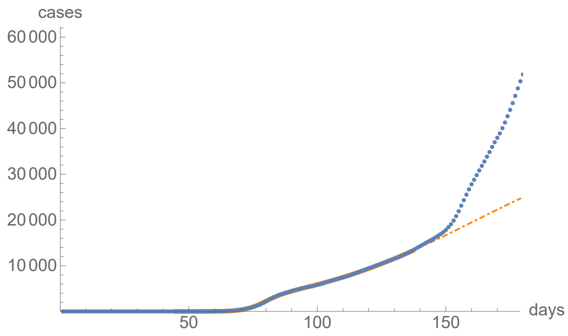


Harris, Texas

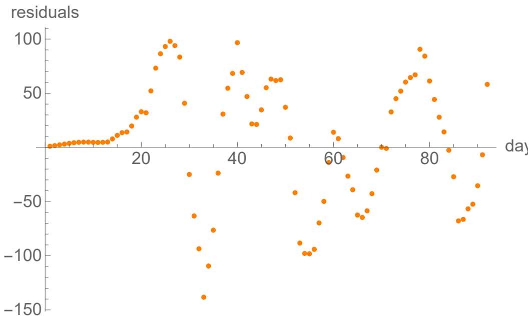


#### Parameters of fits to averaged death data.

The best fit ends on Tue 5 May 2020.

| α | β | k | τ | σ |
|---|---|---|---|---|
| 0.314±0.00716 | 0.088±0.00199 | 262.±3.31 | 30.9±0.261 | 11.9±0.259 |

| badness | | α | β | k | τ | σ | quality |
|---|---|---|---|---|---|---|---|
| 0.5% | error | 2.3% | 2.3% | 1.3% | 0.84% | 2.2% | 9.33% |

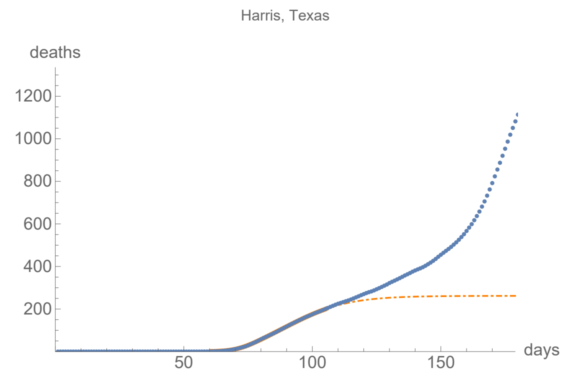


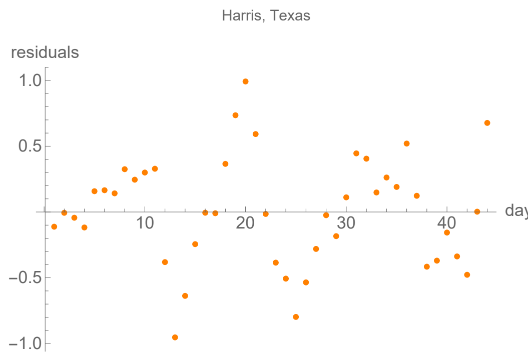


### Fairfax, Virginia

lag from integral overlap, using data averaged over 7 days, between -7 and 0 days, maximum overlap on day -2

lag from fits to the function I (averaged data): -9.76437 days

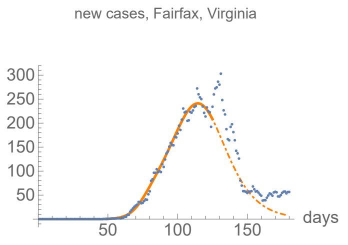


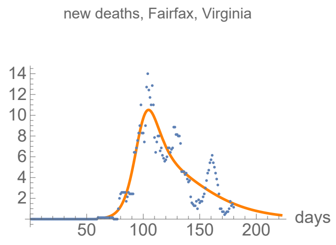


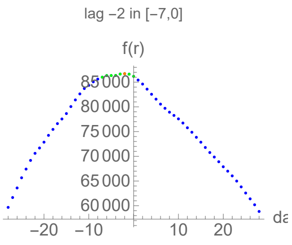


#### Parameters of fits to averaged cases data.

The best fit ends on Mon 25 May 2020.

| α | β | k | τ | σ |
|---|---|---|---|---|
| 0.213±0.00666 | 0.0745±0.000944 | 12800.±134. | 65.8±0.274 | 26.2±0.425 |

| badness | | α | β | k | τ | σ | quality |
|---|---|---|---|---|---|---|---|
| 0.46% | error | 3.1% | 1.3% | 1.% | 0.42% | 1.6% | 7.94% |

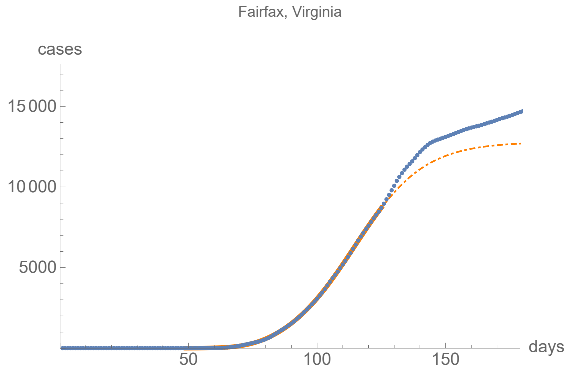


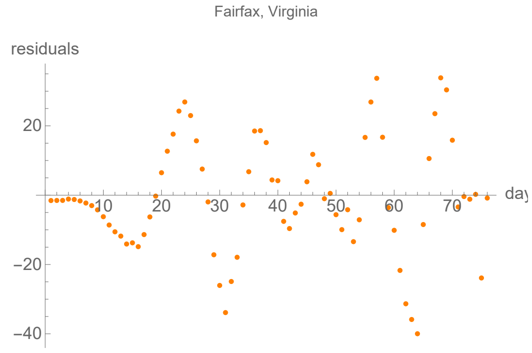


#### Parameters of fits to averaged death data.

The best fit ends on Sun 30 Aug 2020.

| α | β | k | τ | σ |
|---|---|---|---|---|
| 0.174±0.00558 | 0.036±0.00112 | 551.±2.27 | 46.8±0.7 | 34.±0.469 |

| badness | | α | β | k | τ | σ | quality |
|---|---|---|---|---|---|---|---|
| 2.5% | error | 3.2% | 3.1% | 0.41% | 1.5% | 1.4% | 12.1% |

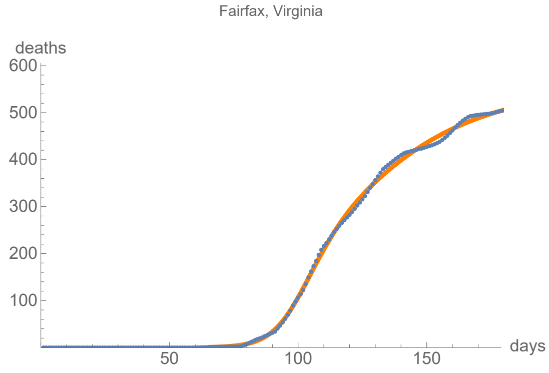


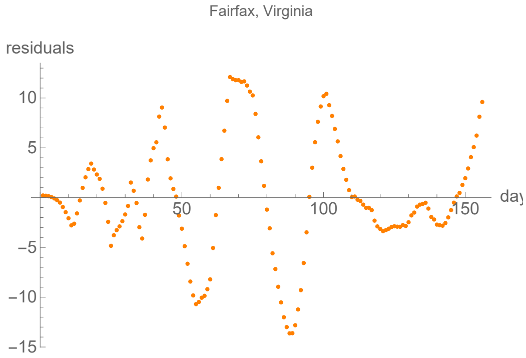


### King, Washington

lag from integral overlap, using data averaged over 7 days, between -2 and 2 days, maximum overlap on day 0

lag from fits to the function I (averaged data): 5.00642 days

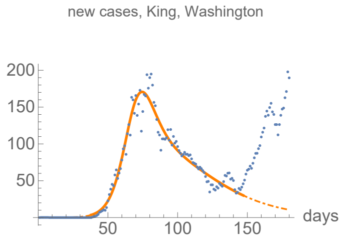


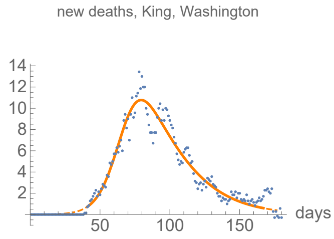


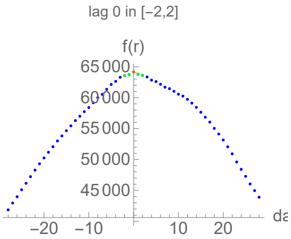


#### Parameters of fits to averaged cases data.

The best fit ends on Tue 16 Jun 2020.

| α | β | k | τ | σ |
|---|---|---|---|---|
| 0.171±0.00302 | 0.036±0.00373 | 9540.±72.7 | 31.2±0.317 | 47.±0.278 |

| badness | | α | β | k | τ | σ | quality |
|---|---|---|---|---|---|---|---|
| 0.94% | error | 1.8% | 10.% | 0.76% | 1.% | 0.59% | 6.75% |

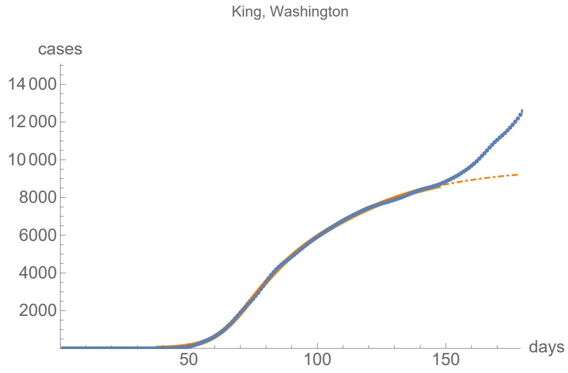


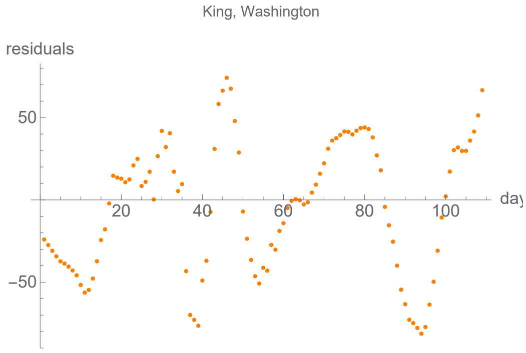


#### Parameters of fits to averaged death data.

The best fit ends on Mon 6 Jul 2020.

| α | β | k | τ | σ |
|---|---|---|---|---|
| 0.141±0.00328 | 0.0437±0.00385 | 624.±2.02 | 28.±0.871 | 39.6±0.997 |

| badness | | α | β | k | τ | σ | quality |
|---|---|---|---|---|---|---|---|
| 1.4% | error | 2.3% | 8.8% | 0.32% | 3.1% | 2.5% | 11.9% |

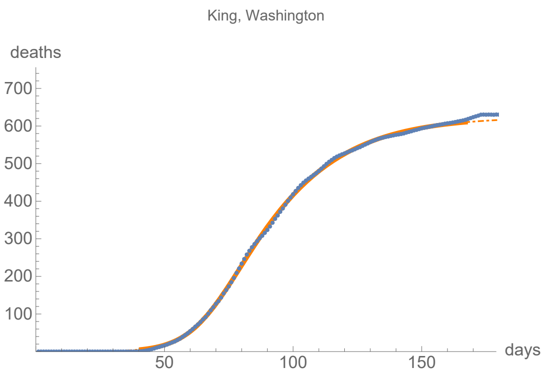
King, Washington
deaths
700
600
500
400
300
200
100
50
100
150
days

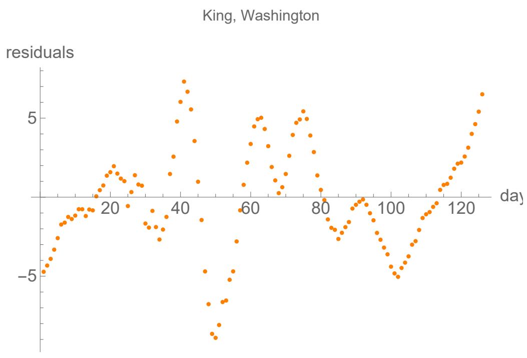
King, Washington
residuals
5
−5
20
40
60
80
100
120
day
